# Associations between disease-specific polygenic risk scores and disease-specific causes of death in the UK Biobank cohort

**DOI:** 10.64898/2025.12.21.25342770

**Authors:** Wenyu Liu, Jennifer A Collister, Lei Clifton, Thomas J Littlejohns, David J Hunter

## Abstract

1.

Disease-specific Polygenic Risk Scores (PRS) are usually evaluated against the incidence of diseases they were derived for. Individuals may be more interested in how these PRS influence their probable cause of death. Using UK Biobank data, we examined the top 10 causes of death among individuals in the highest quintile of disease-specific PRS for Alzheimer’s disease, bowel cancer, cardiovascular disease, coronary artery disease, ischaemic stroke, breast cancer, epithelial ovarian cancer, and prostate cancer. Analyses were stratified by sex, age at death, and smoking status (never, past, current). We also assessed varying PRS percentile thresholds to identify when the target disease became the leading cause of death, and evaluated the impact of each disease-specific PRS on all-cause mortality using Cox proportional hazards models. For most disease-specific PRS, individuals in the high-risk group were more likely to die from other common diseases. The leading causes of death varied according to demographic and behavioural subgroup: breast cancer in women, ischaemic heart disease in men, dementia in the oldest age groups, and lung cancer among smokers. For instance, while prostate cancer was the leading cause of death among older never-smoking men in the highest quintile of the prostate cancer PRS; in other age and smoking status categories, ischaemic heart disease or lung cancer were more common. While a high PRS is predictive of disease diagnosis, most individuals die from other common conditions, depending on their demographic and behavioural subgroups. These findings highlight the importance of contextualising PRS results in clinical settings and risk communication.

**Key messages:**

- What is already known on this topic:

Disease-specific PRS have been investigated for their ability to predict incidence, not death, from the specific target disease.

- What this study adds:

We evaluated PRS for the most common diseases against death from the target disease, as well as other common causes of death.

- How this study might affect research, practice or policy:

Providing the probabilities of death from each target disease, and from other diseases, to the probability of PRS-specific incidence may help contextualise communication of risks associated with high disease-specific PRS.

## 1. Introduction

Polygenic Risk Scores (PRS) are available for dozens of diseases and phenotypes and are beginning to be used clinically as a tool for individual disease risk stratification ^1–4^. Most analyses of PRS focus on their ability to predict incident disease, typically reported as relative risks comparing individuals at the extremes with the middle of the disease-specific PRS distribution. However, this approach blurs the distinction between common and less common diseases, and may overlook absolute disease burden and mortality risk across diseases. These limitations complicate efforts to offer individuals meaningful, personalised risk information across multiple PRS.

Studies examining the communication of PRS across multiple conditions provide further insight into implementation challenges. Connolly et al. ^5^ reported the return of multi-condition PRS results in an ethnically mixed population in the US. They found that participant comprehension, engagement, and equitable use of risk information required scalable, culturally tailored education tools. Lewis et al. ^6^ emphasised the ethical risk of misinterpreting differential PRS performance across genetic ancestry categories as social structure categories and highlighted the need for carefully considered grouping, labelling, and language use in reporting. Moreover, a recent systematic review ^7^ highlighted major challenges in PRS-based risk communication and emphasised the importance of plain language, transparent assumptions, and multimodal visual aids to reduce misinterpretation and enhance understanding.

Although PRS are beginning to enter clinical practice, a gap remains in understanding how disease-specific genetic risk scores relate to actual mortality outcomes, particularly across demographic or behavioural subgroups. Indeed, individuals receiving PRS-based results may assume that the genetically predicted disease is their most likely cause of death. However, diseases differ substantially in their environmental risk factors, their frequency and their mortality consequences. Reducing premature mortality is a major goal of public health ^8^. Yet, evidence on the relationship between disease-specific PRS and cause of death remains limited.

Our primary objective is to investigate how PRS for several major causes of death relate to both death from the target disease and other leading causes of mortality. Specifically, we assess whether individuals at high genetic risk for a particular condition are more likely to die from that target disease or from other common causes, and how this varies by age, sex, and smoking status—factors known to strongly influence mortality. By shifting focus from disease incidence to mortality, this study aims to provide additional practically relevant risk information for the utility of PRS in clinical and public health decision-making.

## 2. Methods

### 2.1 Study population

The study population comprised participants from the UK Biobank (UKB), a prospective cohort study which recruited about half a million individuals aged between 40 and 69 years from England, Scotland and Wales in 2006-2010 ^9^. At recruitment, participants attended a baseline assessment centre, where they provided sociodemographic, lifestyle, and medical history information via a touchscreen questionnaire and a nurse-led interview, and underwent physical examinations. Electronic informed consent was obtained from all participants at the assessment centre. We included participants with complete data on smoking status and selected disease-specific PRS, as described in the Exposure section below. Individuals who had withdrawn from the study were excluded.

### 2.2 Outcome

Mortality data were obtained through linkage to national death registries. The primary outcome was all-cause mortality, with the primary causes of death coded according to the International Classification of Diseases, Tenth Revision (ICD-10), as recorded in UKB data-field 40001. ICD-10 codes were grouped based on Appendix 1 of the UK Biobank Death Summary Report (28 March 2025) ^10^, which categorises common causes of death within the UKB cohort. Linked death data were available from April 2006 onwards. The administrative censoring dates were 31 May 2024 for England and Wales, and 31 December 2023 for Scotland. Follow-up time was measured from baseline until the date of death, the earliest administrative censoring date applicable among the three countries, or the date of loss to follow-up, whichever occurred first.

### 2.3 Exposure

We considered the Standard PRS provided by Genomics PLC and available in the UKB under category 301 ^11^. These scores were derived using external GWAS data only, in contrast to the Enhanced PRS set which was validated using a combination of external and internal UKB data, making it unsuitable to include the whole UKB cohort in analysis due to potential overfitting. We used the most recent PRS datasets that were updated to Version 2 in May 2024.

The Standard PRS we considered were for Alzheimer’s disease (AD), bowel cancer (CRC), cardiovascular disease (CVD), coronary artery disease (CAD), ischaemic stroke (ISS), breast cancer (BC), epithelial ovarian cancer (EOC) and prostate cancer (PC), for which BC and EOC were for females and PC was for males only. For each PRS, the high-risk group was defined as those in the top quintile of PRS risk (unless stated otherwise).

### 2.4 Statistical analysis

We first examined the top 10 causes of deaths and their proportions among overall deaths, comparing the full cohort (denoted as “All” in the figures) to the high-risk group for each disease-specific PRS, stratified by sex, age at death and smoking status (derived from the touchscreen questionnaire where participants self-reported smoking habits). More specifically, participants who died during follow-up were classified into nine subgroups based on age at death (≤59, 60–69, ≥70) and smoking status (never, previous, current), for females (**Figure 1**) and males (**Figure 2**), respectively.

**Figure 1.**
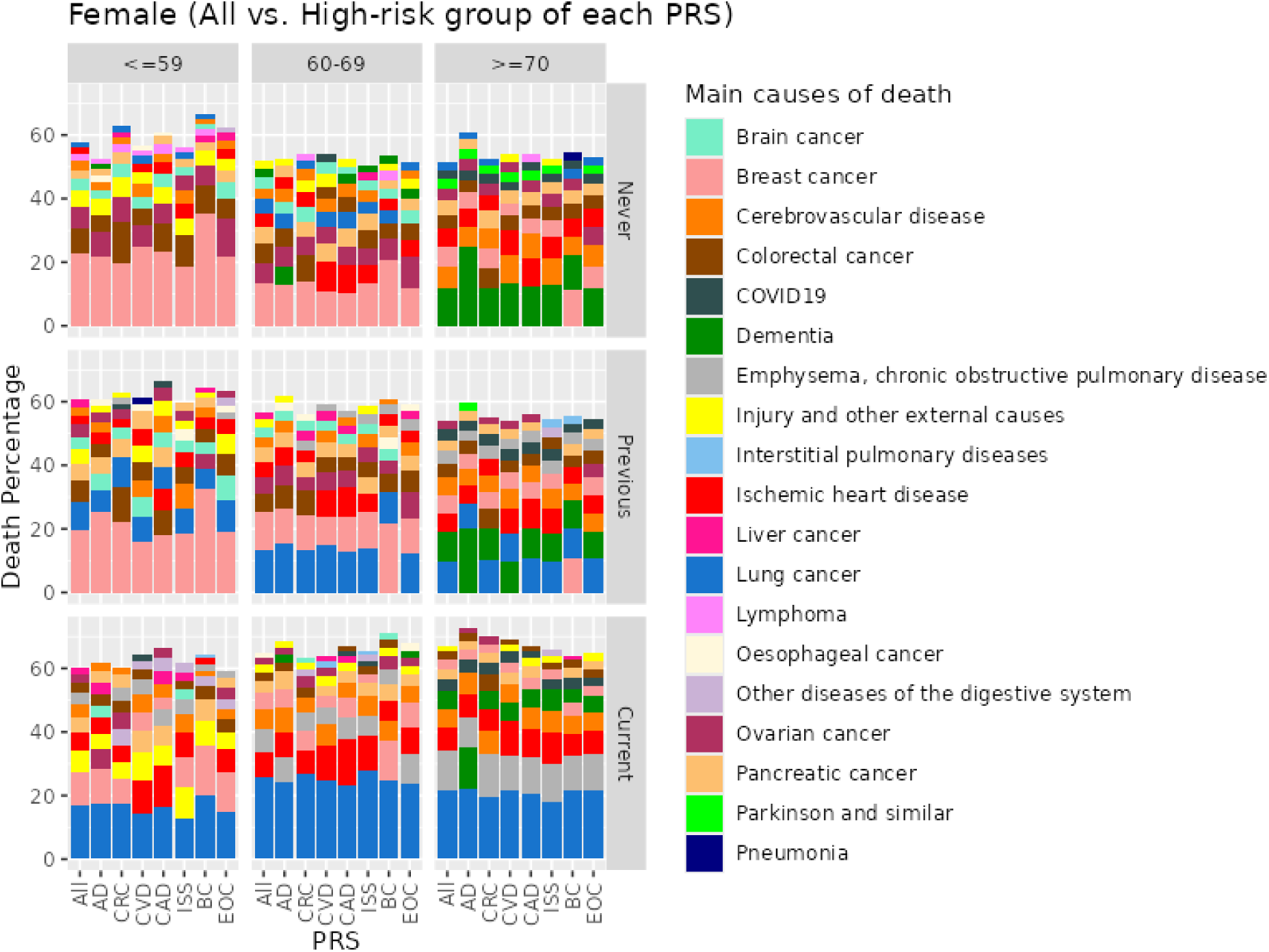
Top 10 causes of death for the high-risk group of the Standard PRS for UKB female participants stratified by age of death group and smoking status. Alzheimer’s disease (AD); bowel cancer (CRC); cardiovascular disease (CVD); coronary artery disease (CAD); ischaemic stroke (ISS); breast cancer (BC); epithelial ovarian cancer (EOC). Age of death groups: <=59, 60-69, >=70; smoking status groups: Never, Previous, Current.

**Figure 2.**
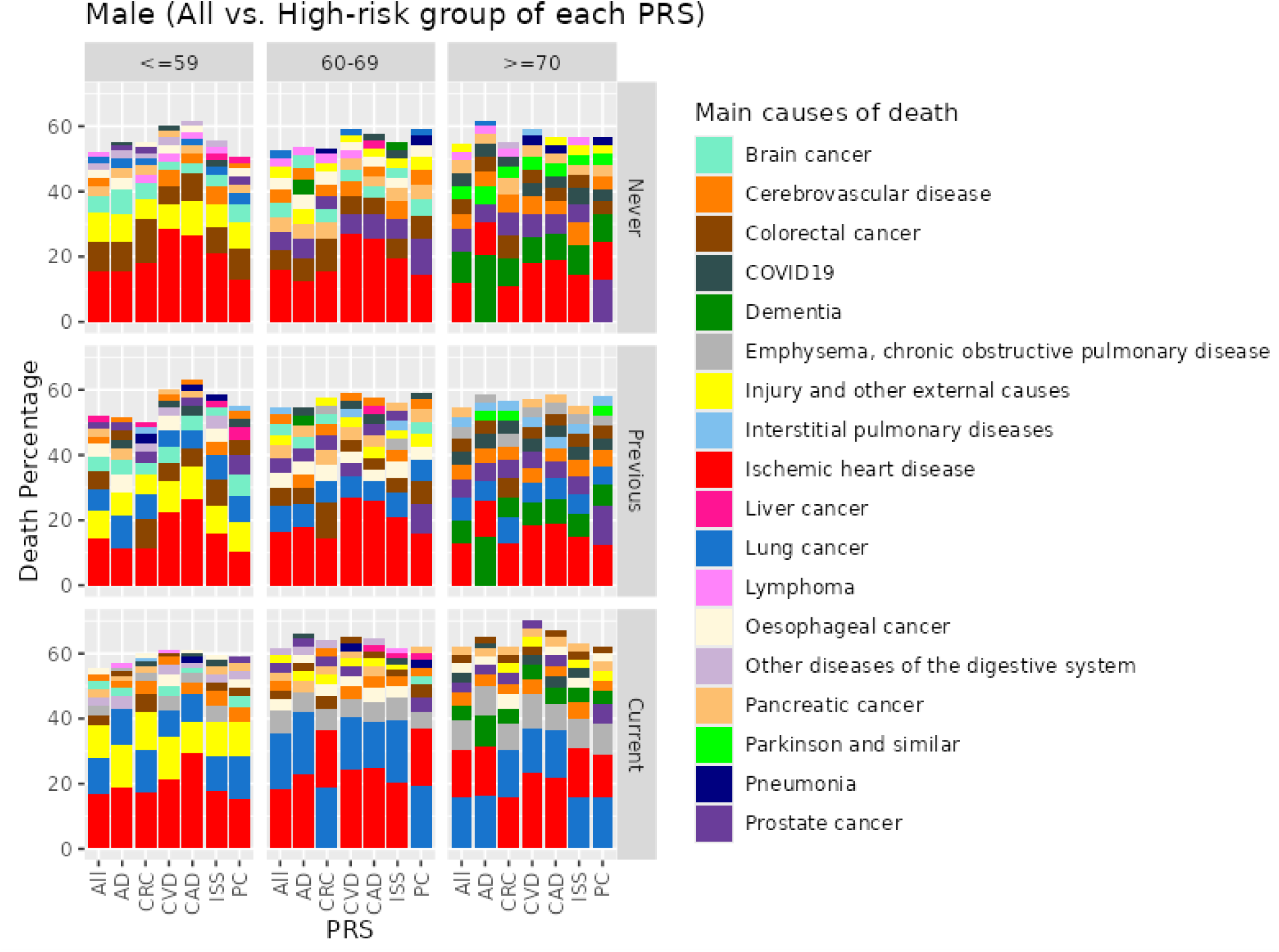
Top 10 causes of death for the high-risk group of the Standard PRS for UKB male participants stratified by age of death group and smoking status. Alzheimer’s disease (AD); bowel cancer (CRC); cardiovascular disease (CVD); coronary artery disease (CAD); ischaemic stroke (ISS); prostate cancer (PC). Age of death groups: <=59, 60-69, >=70; smoking status groups: Never, Previous, Current.

We categorised the PRS into quintiles, with the top 20% considered high-risk, i.e., their predicted likelihood of developing the disease was in the top 20% of polygenic risk. We also explored a wide range of PRS percentile cut points (1^st^ to 99^th^) to identify at what threshold (if any) the target disease became the leading cause of death within demographic and behavioural subgroups (**Table 1**).

**Table 1.** Percentile cut point at which the target disease for the PRS first became the leading cause of death.

| Females; Never smokers |  |  |  |  |  |  |  |
| --- | --- | --- | --- | --- | --- | --- | --- |
| Death age | AD<sup>1</sup> | CRC<sup>2</sup> | CVD<sup>3</sup> | PRS<br>CAD<sup>4</sup> | ISS<sup>5</sup> | BC<sup>6</sup> | EOC<sup>7</sup> |
| <=59 | NA | 98 | NA | NA | NA | 1 | NA |
| 60-69 | NA | NA | 90 | 95 | NA | 1 | 90 |
| 70+ | 1 | NA | NA | NA | NA | 80 | NA |
| Females; Previous smokers |  |  |  |  |  |  |  |
| Death age | AD | CRC | CVD | PRS<br>CAD | ISS | BC | EOC |
| <=59 | NA | NA | NA | 90 | NA | 1 | NA |
| 60-69 | NA | 98 | 99 | 90 | NA | 15 | 98 |
| 70+ | 10 | NA | 85 | 90 | 99 | 60 | NA |
| Females; Current smokers |  |  |  |  |  |  |  |
| Death age | AD | CRC | CVD | PRS<br>CAD | ISS | BC | EOC |
| <=59 | NA | 98 | 80 | 90 | NA | 80 | NA |
| 60-69 | NA | NA | NA | 99 | NA | 97 | NA |
| 70+ | 99 | NA | NA | NA | NA | NA | NA |
| Males; Never smokers |  |  |  |  |  |  |  |
| Death age | AD | CRC | CVD | PRS<br>CAD | ISS | PC<sup>8</sup> |  |
| <=59 | NA | 90 | 1 | 1 | 1 | 99 |  |
| 60-69 | 99 | 95 | 1 | 1 | 1 | 99 |  |
| 70+ | 30 | NA | 1 | 1 | 1 | 55 |  |
| Males; Previous smokers |  |  |  |  |  |  |  |
| Death age | AD | CRC | CVD | PRS<br>CAD | ISS | PC |  |
| <=59 | NA | NA | 1 | 1 | 1 | NA |  |
| 60-69 | NA | NA | 1 | 1 | 1 | 97 |  |
| 70+ | 65 | NA | 1 | 1 | 1 | 85 |  |
| Males; Current smokers |  |  |  |  |  |  |  |
| Death age | AD | CRC | CVD | PRS<br>CAD | ISS | PC |  |
| <=59 | NA | 95 | 1 | 1 | 1 | NA |  |
| 60-69 | NA | NA | 1 | 1 | 1 | 99 |  |
| 70+ | NA | NA | 25 | 20 | 50 | NA |  |

<sup>1</sup>: Alzheimer's disease (AD), <sup>2</sup>: bowel cancer (CRC), <sup>3</sup>: cardiovascular disease (CVD), <sup>4</sup>: coronary artery disease (CAD), <sup>5</sup>: ischaemic stroke (ISS), <sup>6</sup>: breast cancer (BC), <sup>7</sup>: epithelial ovarian cancer (EOC), <sup>8</sup>: prostate cancer (PC).
NA means that the target disease for the PRS never became the major cause of death.
For each cell in the table (by age at death, sex, smoking status, and PRS), the corresponding plot showing the percentile cut point (if any) at which the target disease became the leading cause of death is shown in the Supplemental Material.

To investigate the influence of each disease-specific PRS on all-cause mortality, we fitted Cox proportional hazard models adjusted for age, sex (for PRS that are not sex-specific), and smoking status using the whole study population described above. PRS were evaluated (i) as quintiles (reference: third quintile, i.e., the middle risk group) (**Table 2**) and (ii) per 1 standard deviation (SD) increase (**Figure 3**). The proportional hazards assumption was assessed using scaled Schoenfeld residuals.

**Table 2.** Hazard ratio of all-cause mortality by PRS risk group, adjusted for smoking status, baseline age as well as sex if the PRS is not sex-specific.

| PRS |  |  |  | PRS |  |  |  |
| --- | --- | --- | --- | --- | --- | --- | --- |
| Quintile | HR | 95% CI | p-value | Quintile | HR | 95% CI | p-value |
| AD |  |  |  | ISS |  |  |  |
| Q5 | 1.18 | 1.15, 1.22 | <0.001 | Q5 | 1.15 | 1.12, 1.18 | <0.001 |
| Q4 | 1.04 | 1.01, 1.07 | 0.007 | Q4 | 1.05 | 1.02, 1.08 | <0.001 |
| Q3 | — | — | — | Q3 | — | — | — |
| Q2 | 1.01 | 0.98, 1.03 | 0.66 | Q2 | 0.95 | 0.93, 0.98 | 0.001 |
| Q1 | 0.99 | 0.96, 1.01 | 0.29 | Q1 | 0.90 | 0.87, 0.92 | <0.001 |
| CRC |  |  |  | BC<sup>1</sup> |  |  |  |
| Q5 | 1.08 | 1.05, 1.10 | <0.001 | Q5 | 1.15 | 1.10, 1.19 | <0.001 |
| Q4 | 1.01 | 0.98, 1.03 | 0.69 | Q4 | 1.06 | 1.01, 1.10 | 0.01 |
| Q3 | — | — | — | Q3 | — | — | — |
| Q2 | 0.98 | 0.96, 1.01 | 0.24 | Q2 | 0.95 | 0.91, 1.00 | 0.04 |
| Q1 | 0.94 | 0.92, 0.97 | <0.001 | Q1 | 0.98 | 0.94, 1.02 | 0.29 |
| CVD |  |  |  | EOC<sup>1</sup> |  |  |  |
| Q5 | 1.12 | 1.09, 1.15 | <0.001 | Q5 | 1.04 | 1.00, 1.09 | 0.06 |
| Q4 | 1.03 | 1.01, 1.06 | 0.02 | Q4 | 0.99 | 0.95, 1.03 | 0.60 |
| Q3 | — | — | — | Q3 | — | — | — |
| Q2 | 0.97 | 0.94, 0.99 | 0.02 | Q2 | 0.98 | 0.93, 1.02 | 0.25 |
| Q1 | 0.91 | 0.88, 0.93 | <0.001 | Q1 | 0.96 | 0.92, 1.00 | 0.07 |
| CAD |  |  |  | PC<sup>2</sup> |  |  |  |
| Q5 | 1.09 | 1.07, 1.12 | <0.001 | Q5 | 1.07 | 1.03, 1.11 | <0.001 |
| Q4 | 1.03 | 1.01, 1.06 | 0.02 | Q4 | 1.03 | 1.00, 1.07 | 0.08 |
| Q3 | — | — | — | Q3 | — | — | — |
| Q2 | 0.96 | 0.93, 0.98 | 0.001 | Q2 | 1.01 | 0.97, 1.04 | 0.72 |
| Q1 | 0.90 | 0.88, 0.93 | <0.001 | Q1 | 0.99 | 0.96, 1.03 | 0.63 |

<sup>1</sup>: Females only; <sup>2</sup>: Males only. HR = Hazard Ratio; CI = Confidence Interval; Alzheimer’s disease (AD); bowel cancer (CRC); cardiovascular disease (CVD); coronary artery disease (CAD); ischaemic stroke (ISS); breast cancer (BC); epithelial ovarian cancer (EOC); prostate cancer (PC).

**Figure 3.**
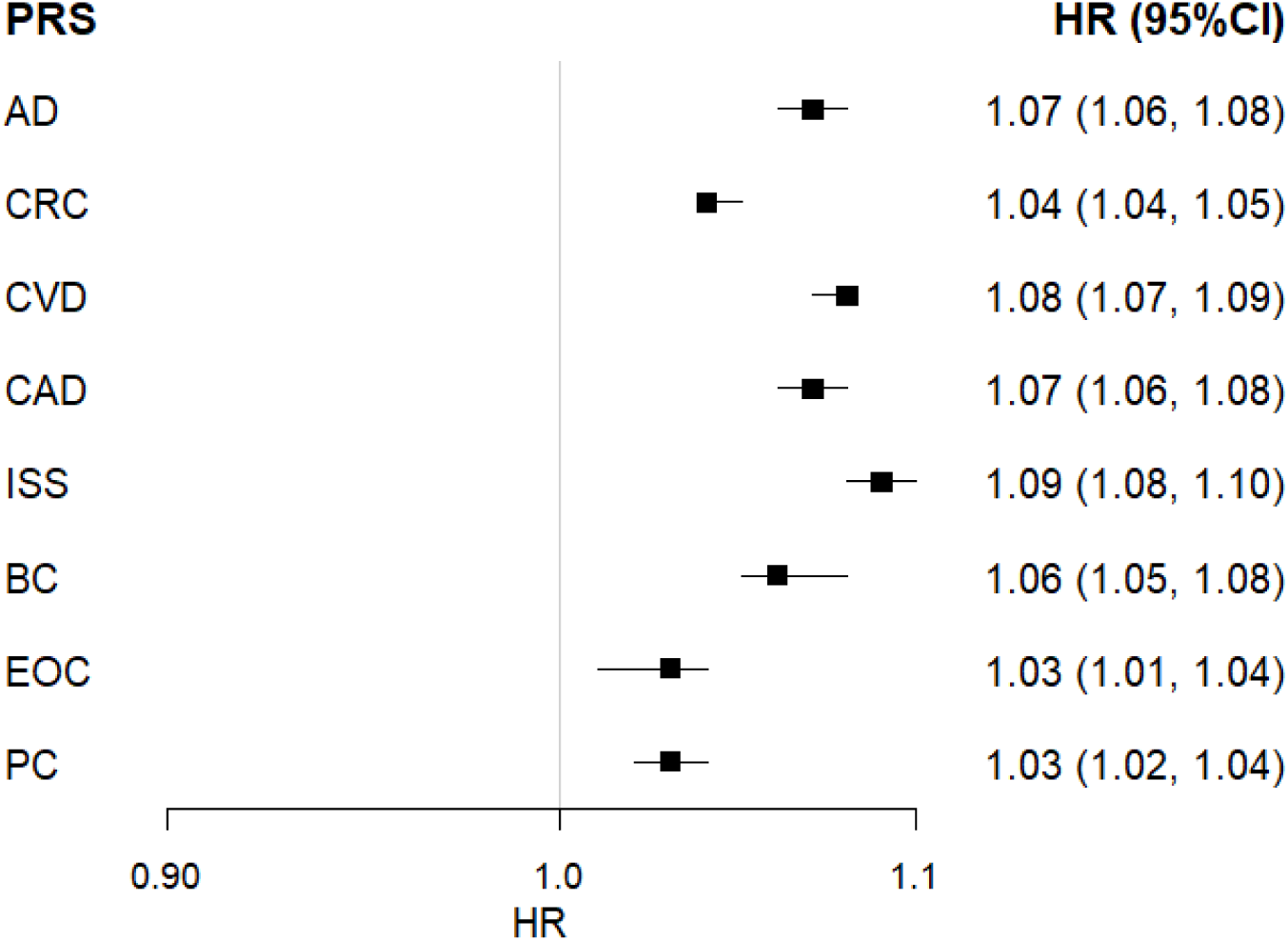
Hazard ratio of all-cause mortality per 1 SD increase of the disease-specific PRS, adjusted for smoking status, age at baseline as well as sex if the PRS is not sex-specific.

We used data available in UKB Data Release v19.1. All analyses were performed using RStudio version 4.4.0 on the cloud-based UK Biobank Research Analysis Platform (UKB-RAP).

## 3. Results

Among the 261,020 female participants, 8.1% died during follow-up. Within the high-risk group for each disease-specific PRS, the proportion who died from any cause was 9.2% for the AD PRS, 8.5% for the CRC PRS, 8.7% for the CVD PRS, 8.7% for the CAD PRS, 9.0% for the ISS PRS, 8.9% for the BC PRS, and 8.4% for the EOC PRS. For males (N=220,019), 13.6% died during follow-up, and the corresponding mortality proportions in the high-risk group for each PRS were 15.1% for AD, 14.9% for CRC, 15.1% for CVD, 14.7% for CAD, 15.5% for ISS, and 14.2% for PC.

**Figures 1 and 2** display the top 10 causes of death by sex, age at death, smoking status, and PRS group, that accounted for approximately 60% of all deaths in both sexes. The percentage of deaths in the figures was calculated based on their corresponding subgroup. For instance, for all female never smokers who died <=59 years of age, 22.8% died from breast cancer. Whereas, among individuals who were at high genetic risk (i.e., in the top 20% of PRS for BC), 35.5% died from breast cancer.

**Figure 1** shows that breast cancer was the major cause of death for young females who never or previously smoked. For women who died after 70, dementia became the main cause of death, except in the high-risk BC PRS group where breast cancer was still the leading cause of death. For all current smokers, regardless of PRS, the leading cause of death was lung cancer. This was also true for some previous smokers who died at 60 or older, except those with high polygenic risk of breast cancer and dementia.

**Figure 2** shows that Ischemic heart disease was the major cause of death for males across various PRS and most demographic/behavioural subgroups. Dementia emerged as the second leading cause of death for never and former smokers who died at age 70 or above. Lung cancer was the most common cause of death among older current smokers in several high-risk categories of PRS for other diseases, however, the number of males who died from lung cancer only marginally exceeded the number who died from ischemic heart disease.

**Table 1** presents the disease-specific PRS percentile cut points (if any) at which the target disease became the leading cause of death by subgroup. For example, dementia became the most frequent cause of death among female never-smokers aged ≥70 even at the 1st percentile of AD PRS, suggesting its dominance regardless of polygenic risk of developing Alzheimer’s disease. More specifically, the target disease, dementia, was the leading cause of death for people in the top 99% of the AD risk scores. This association was less strong for female previous smokers older than 70, still, the leading cause of death was dementia from as low as the 10^th^ percentile of the AD PRS. The same findings applied for males although the association was not as pronounced as for females. Similarly, the table reconfirmed that the leading cause of death was breast cancer for women, especially for younger female never/previous smokers, and ischemic heart disease accounted for a high proportion of the causes of death for men across subgroups, where the percentile cut point was 1 (except for older current smokers) for the PRS associated with Ischemic heart disease (i.e., CVD, CAD and ISS).

On the other hand, for male never smokers who died after age 70, the PC PRS percentile cut point of 55 and above demonstrated that prostate cancer was the major cause of death among individuals in the top 45% of the PC PRS, despite ischemic heart disease being the major cause of death for men overall. Note that for some disease-specific PRS, the target disease never became the leading cause of death even at the 99^th^ percentile cut point (denoted as NA in **Table 1**) because other causes of death were more common, such as lung cancer for current smokers.

In the overall survival analysis, the median follow-up time is 15.2 years. **Table 2** summarises hazard ratios (HRs) of all-cause mortality adjusted for age, sex (if applicable) and cigarette smoking by PRS quintiles. Compared to the middle-risk group (the 3^rd^ quintile), individuals in the top quintile of ISS PRS had a 15% higher risk of death, while those in the lowest quintile had a 10% lower risk. Similar strength and direction of the associations were observed from other PRS, although the 95% CI of some adjusted HRs contained 1.

HRs displayed in **Table 2** are adjusted for key covariates. Overall, the risk of all-cause mortality was higher among older individuals, males, and those who smoked cigarettes. Compared to never-smokers, the adjusted HR was 1.38 (95% CI: 1.35–1.41) for former smokers, and 2.86 (95% CI: 2.78–2.94) for current smokers. Each additional year of age at baseline was associated with a 11% increased risk of all-cause mortality. Males had a higher risk of all-cause mortality than females (HR=1.58, 95% CI: 1.55–1.61). In sex-stratified models risk associated with current smoking at baseline remained high: HR 2.94 (95% CI: 2.82–3.07) for females, and 2.83 (95% CI: 2.73–2.93) for males.

As shown in Figure 3, a 1 SD increase in each PRS was associated with a modest increase in all-cause mortality. The adjusted HR for other covariates were similar to Table 2. We observed no evidence of violation of the proportional hazard assumption for all Cox models. (Figure 3 here)

## 4. Discussion

Most presentations of risk associated with PRS focus on relative or absolute risk over time of the target disease. People might naturally assume that if they are at high risk of a specific disease that disease is likely to cause their death. However, the absolute risk of a low or average polygenic risk of a common cause of death may be higher than a higher polygenic risk of a less common cause of death. Presenting information about disease-specific PRS in the context of all causes of death may help people focus on the causes of preventable mortality, rather than only a specific disease for which they may be at higher risk of diagnosis, but still at low absolute risk of death. PRS are increasingly proposed as tools for stratifying disease risk, however, our findings demonstrate that a high polygenic predisposition to a particular disease does not necessarily mean it will be the most likely cause of death. This finding is also consistent with Yang et. al ^12^, who compared the genetic architectures of disease susceptibility and disease-specific mortality across nine common diseases in seven biobanks, and concluded that genetic variants or PRS with strong effects on disease susceptibility had little to no association with disease-specific mortality after diagnosis. This mismatch between genetic risk of a disease and actual cause of death underscores the importance of contextualising PRS within broader epidemiological and behavioural frameworks.

In the UKB cohort, the leading causes of death were more strongly influenced by age, sex, and smoking status than by polygenic predisposition to developing a specific disease alone. For instance, ischaemic heart disease was the dominant cause of death among men, while dementia and breast cancer were leading causes among older women and younger non-smokers, respectively. Lung cancer remained the primary cause of death for smokers regardless of their genetic risk for other conditions. For many PRS, the target disease did not emerge as the most common cause of death even at extreme PRS thresholds (e.g. the 99th percentile). Most people with a high disease-specific PRS were still more likely to die from unrelated but more prevalent diseases.

Our findings also align with previous studies highlighting the complexity of interpreting and communicating polygenic risk in clinical or public health settings. Misinterpretation of high PRS as deterministic can lead to unnecessary anxiety or misplaced health priorities ^2^. For individuals and clinicians, understanding the relative importance of a PRS within the broader landscape of mortality risk may influence decision-making about preventive actions. A high genetic risk for prostate cancer might warrant increased screening, but in many older men, cardiovascular disease remained a far more likely cause of death—even at lower cardiovascular PRS levels.

Limitations of the study include the known evidence of a “healthy volunteer” bias in the UK Biobank population ^9^, in which smoking prevalence and age-specific mortality was somewhat lower than in the UK general population. However, stratifying by smoking status should have removed of this bias with respect to smoking-specific mortality. Additionally, the UKB cohort consists predominantly of individuals of European ancestry. Previous studies ^1, 13^ have highlighted the potential concern of exacerbating ethnic inequities as PRS often exhibits reduced predictive performance in non-European individuals (who are typically underrepresented in the genome-wide association studies used to derive these scores). Our study has several strengths, including the use of a large, well-characterised cohort, stratification by key behavioural and demographic factors, and the use of validated PRS derived from external data. In summary, presenting disease-specific PRS in the context of risk of death from that disease and other diseases, may help focus risk communication on diseases that are more common causes of death, and exposures such as smoking that cause most preventable deaths.

## Data Availability

This research has been conducted using the UK Biobank (https://www.ukbiobank.ac.uk) Resource under Application Number 33952. All results presented in this manuscript, including the code used to generate them, will be returned to UK Biobank within 6 months of publication at which point they are made available for researchers to request (subject to UK Biobank approval). It is not feasible to share the data from us but from UKBB directly upon application approval. This is due to the UKBB restrictions that no individual–level data can be downloaded from the UK Biobank Research Analysis Platform (UKB–RAP) to a local machine. The cloud-based platform UKB–RAP is where a registered researcher can access the UKBB data and conduct their analyses.

## 4. Declarations

### 4.1 Ethics approval and consent to participate

The UK Biobank study was approved by the North West Multi-centre Research Ethics Committee (REC reference: 11/NW/03820). All participants provided written informed consent prior to enrolment, and the study was conducted in accordance with the principles of the Declaration of Helsinki. The analyses reported here use existing data from the UK Biobank cohort; the authors had no role in participant recruitment. Patients were not involved in the interpretation or writing of these results. Findings from UK Biobank are routinely shared with participants through the study website and social media channels.

### 4.2 Data and code availability

This research has been conducted using the UK Biobank (https://www.ukbiobank.ac.uk) Resource under Application Number 33952. All results presented in this manuscript, including the code used to generate them, will be returned to UK Biobank within 6 months of publication at which point they are made available for researchers to request (subject to UK Biobank approval). It is not feasible to share the data from us but from UKBB directly upon application approval. This is due to the UKBB restrictions that no individual-level data can be downloaded from the UK Biobank Research Analysis Platform (UKB-RAP) to a local machine. The cloud-based platform UKB-RAP is where a registered researcher can access the UKBB data and conduct their analyses.

### 4.3 Author contributions

**DJH**: Funding acquisition, Conceptualization, Methodology, Supervision, Writing – review & editing. **WL**: Data curation, Software, Methodology, Formal analysis, Visualization, Writing – original draft. **JAC**: Software, Methodology, Writing – review. **LC**: Methodology, Writing – review. **TJL**: Methodology, Writing – review.

### 4.4 Transparency statement

The lead author affirms that this manuscript is an honest, accurate, and transparent account of the study being reported; that no important aspects of the study have been omitted; and that any discrepancies from the study as planned have been explained.

## 4.5 Acknowledgements

We thank the participants and staff of the UK Biobank for enabling us to conduct this research. This study has been conducted using the UK Biobank Resource under Application Number 33952. The work was supported by the Wellcome Trust, Medical Research Council, Department of Health, Scottish government, and Northwest Regional Development Agency. It has also received funding from the Welsh Assembly government and British Heart Foundation. The analyses here were funded by the Cancer Research UK [grant number C16077/A29186], and supported by the Nuffield Department of Population Health, Oxford University.

