## Supplemental Material for "Associations between disease-specific polygenic risk scores and disease-specific causes of death in the UK Biobank cohort"

### Examining the percentile cut point for which the disease-specific PRS becomes the leading cause of death

#### 1 Alzheimer's disease (AD) PRS for UKB females

##### 1.1 Female never smokers who died at age <=59

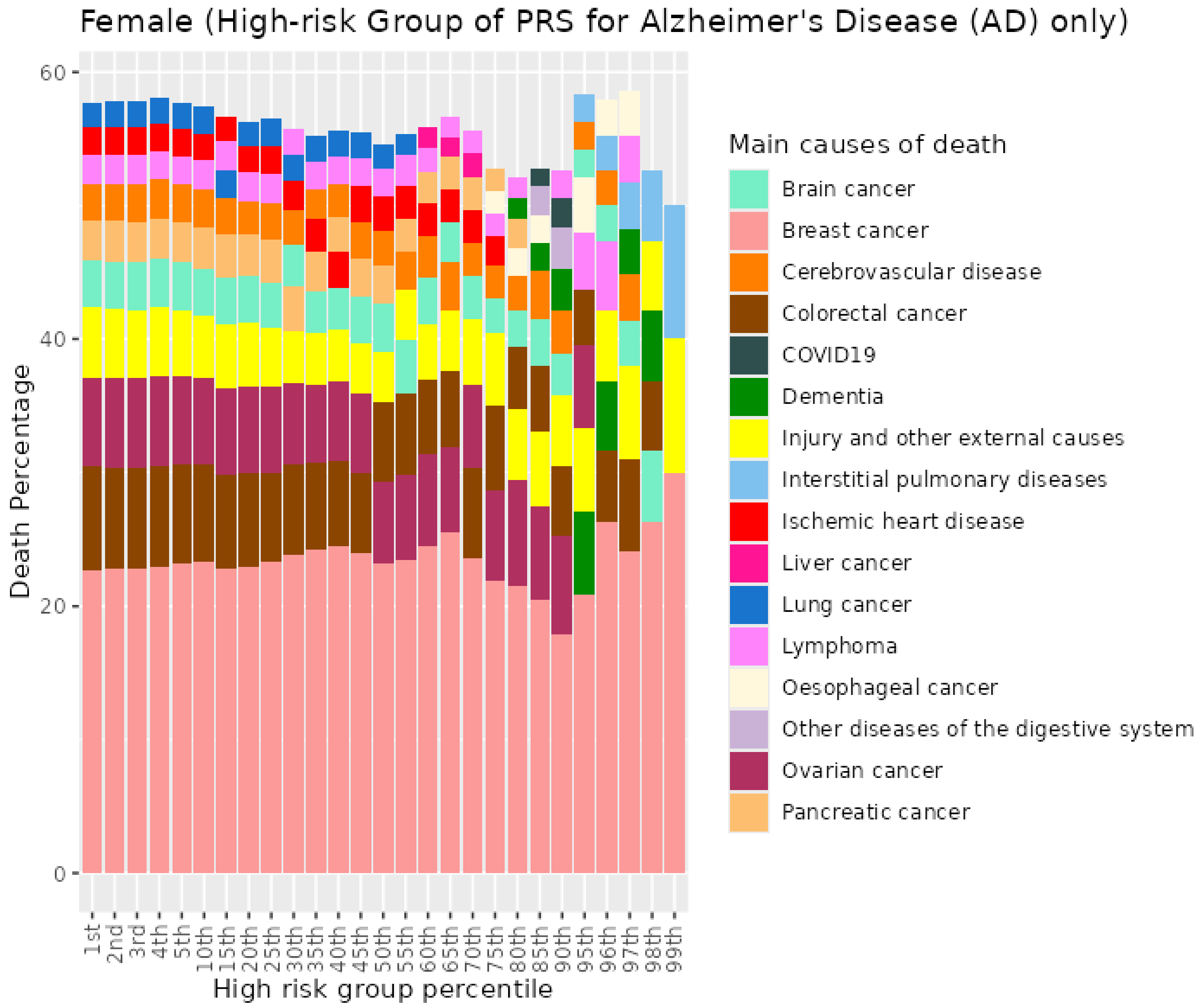


*Figure S1. Percentiles from 1 to 99 for Alzheimer's disease PRS among female never smokers who died at age <=59*

#### 1.2 Female never smokers who died at age 60-69

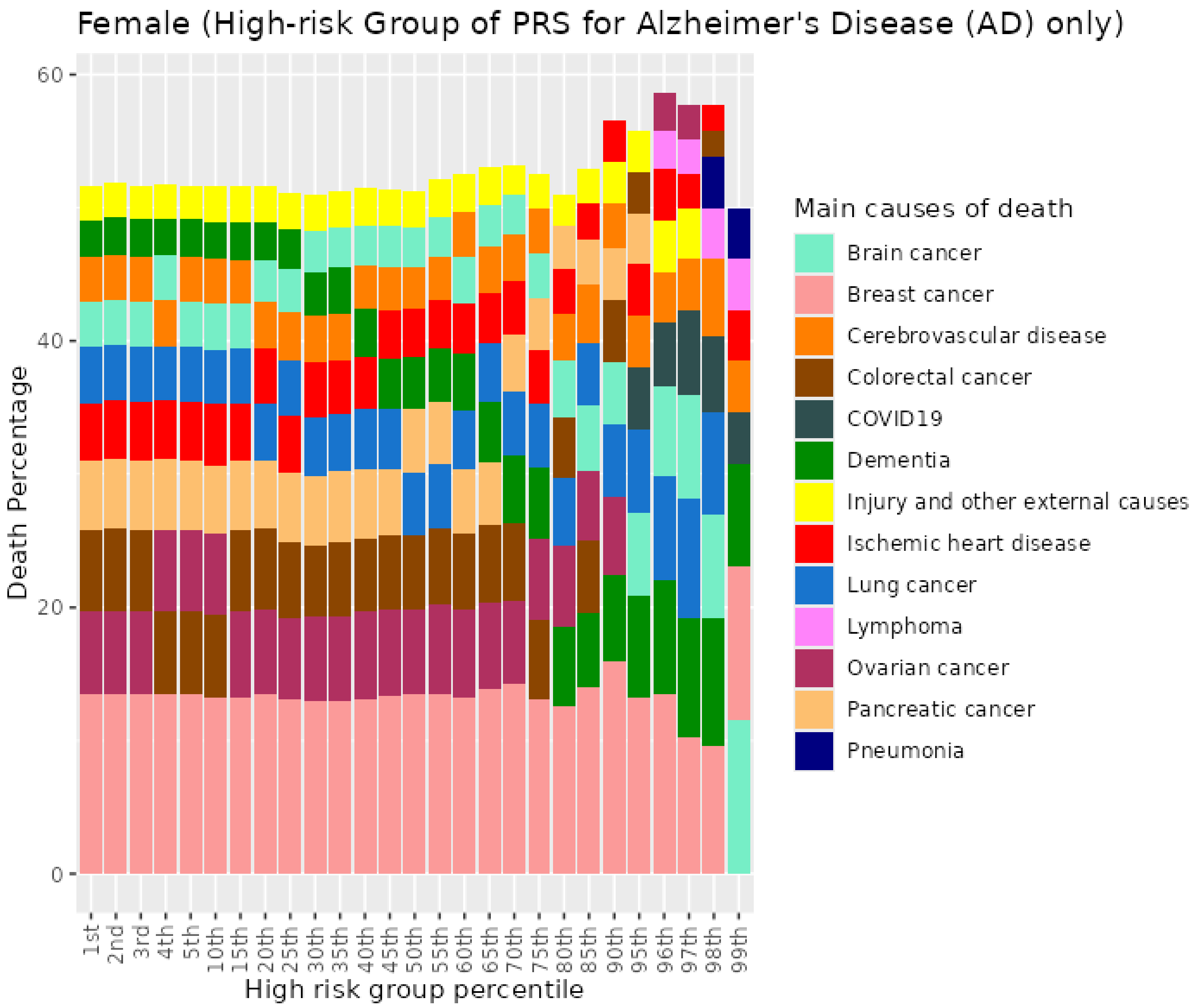


*Figure S2. Percentiles from 1 to 99 for Alzheimer's disease PRS among female never smokers who died at age 60-69*

##### 1.3 Female never smokers who died at age 70+

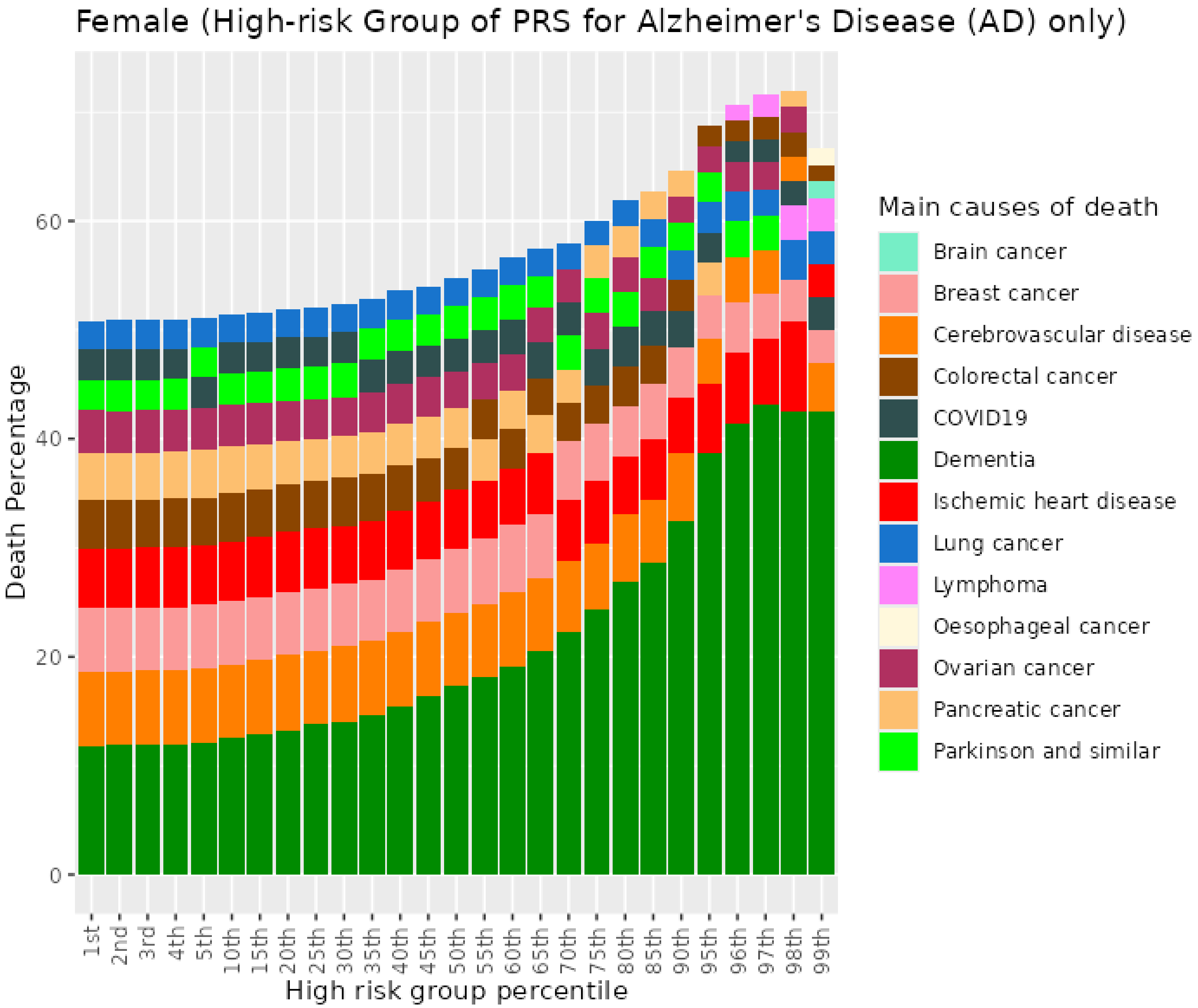


*Figure S3. Percentiles from 1 to 99 for Alzheimer's disease PRS among female never smokers who died at age 70+*

##### 1.4 Female previous smokers who died at age <=59

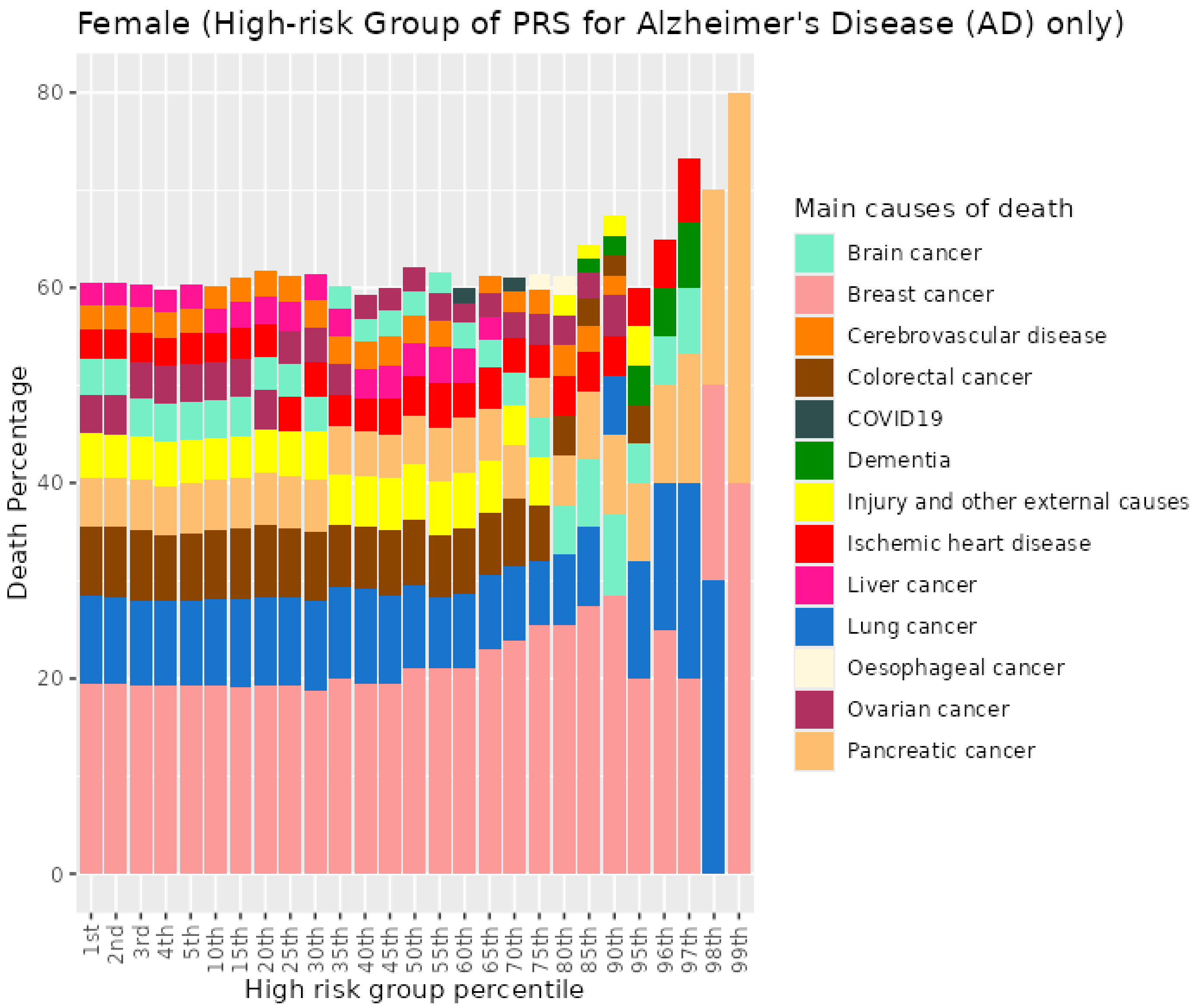


*Figure S4. Percentiles from 1 to 99 for Alzheimer's disease PRS among female previous smokers who died at age <=59*

#### 1.5 Female previous smokers who died at age 60-69

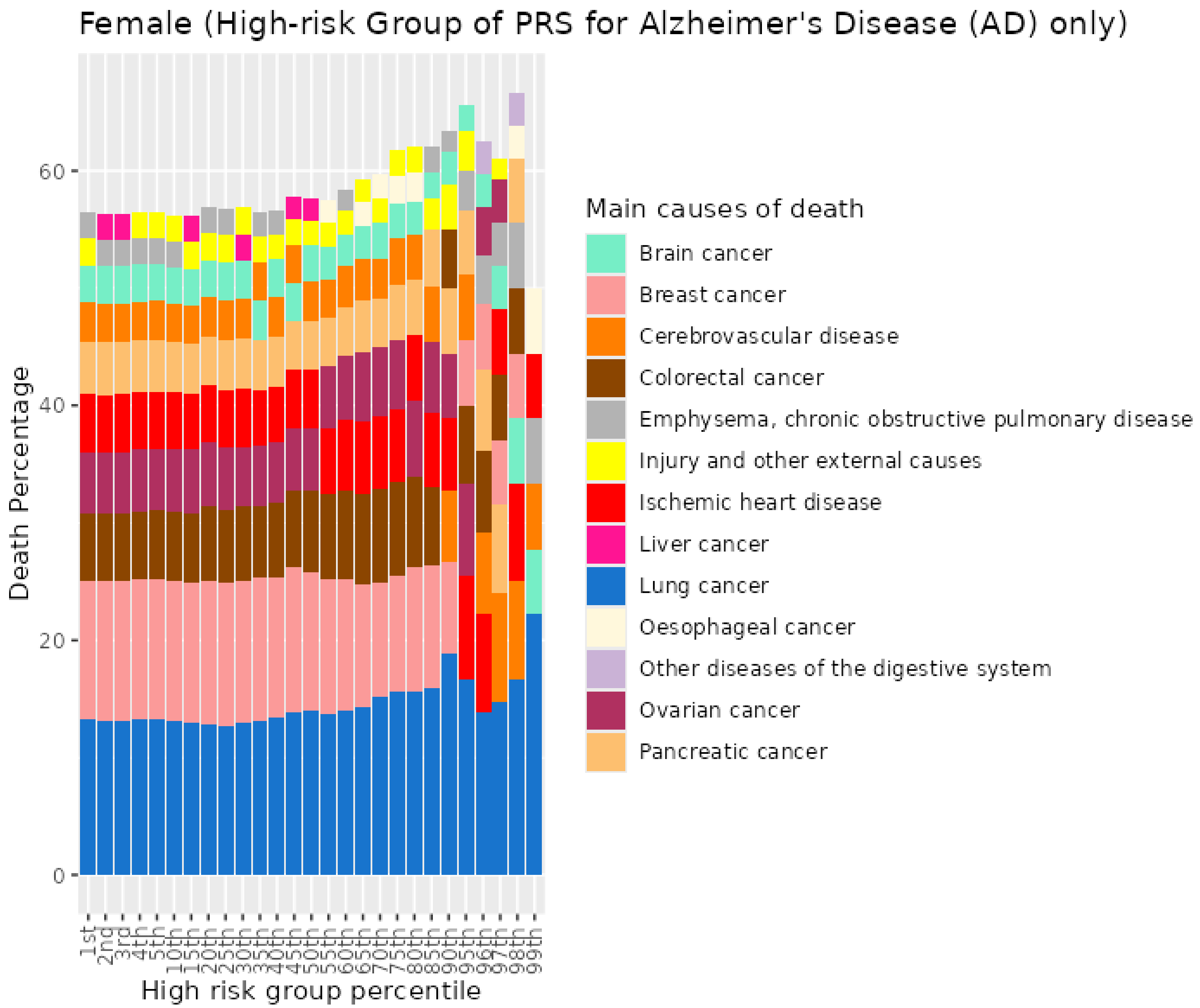


*Figure S5. Percentiles from 1 to 99 for Alzheimer's disease PRS among female previous smokers who died at age 60-69*

#### 1.6 Female previous smokers who died at age 70+

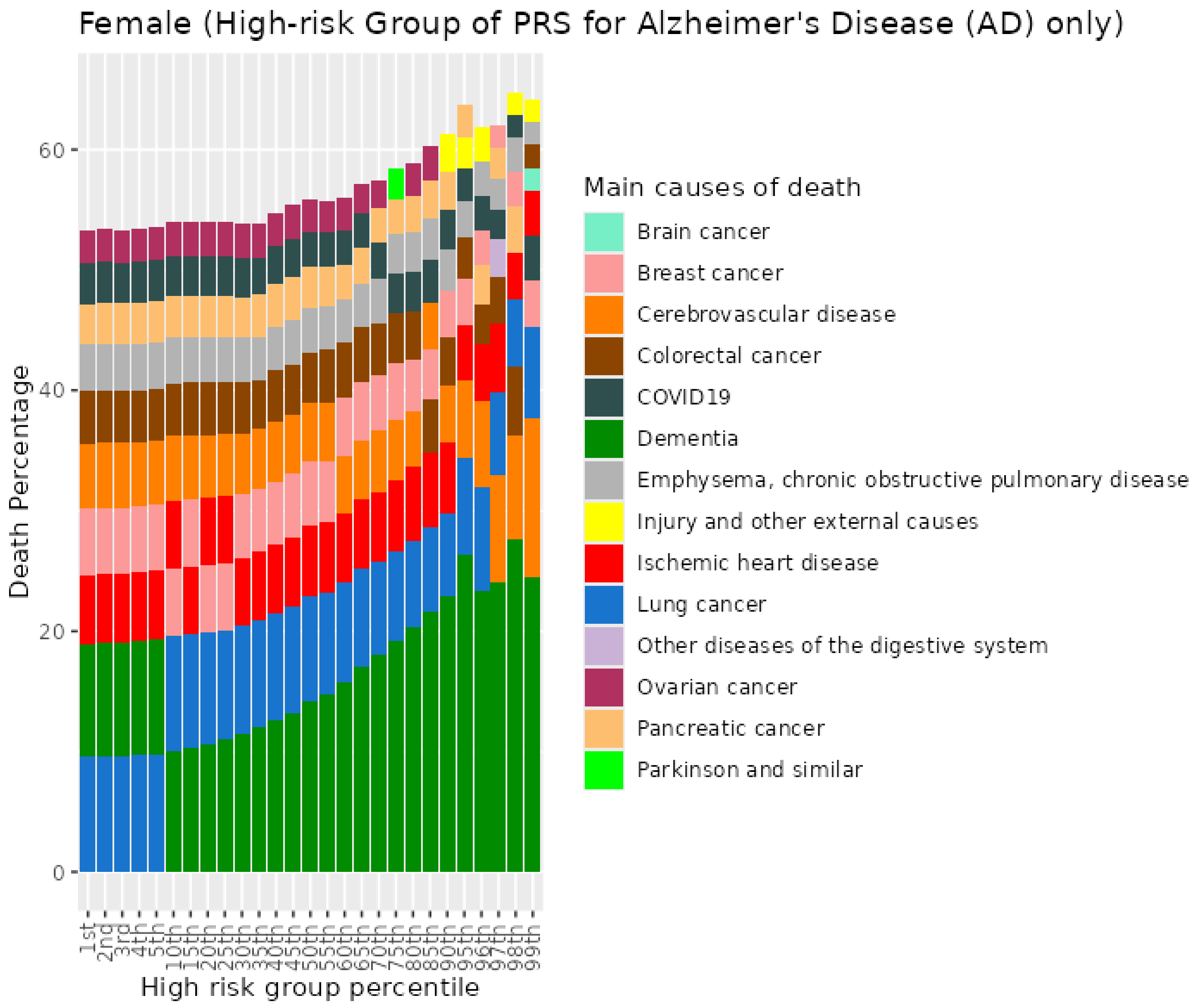


*Figure S6. Percentiles from 1 to 99 for Alzheimer's disease PRS among female previous smokers who died at age 70+*

##### 1.7 Female current smokers who died at age <=59

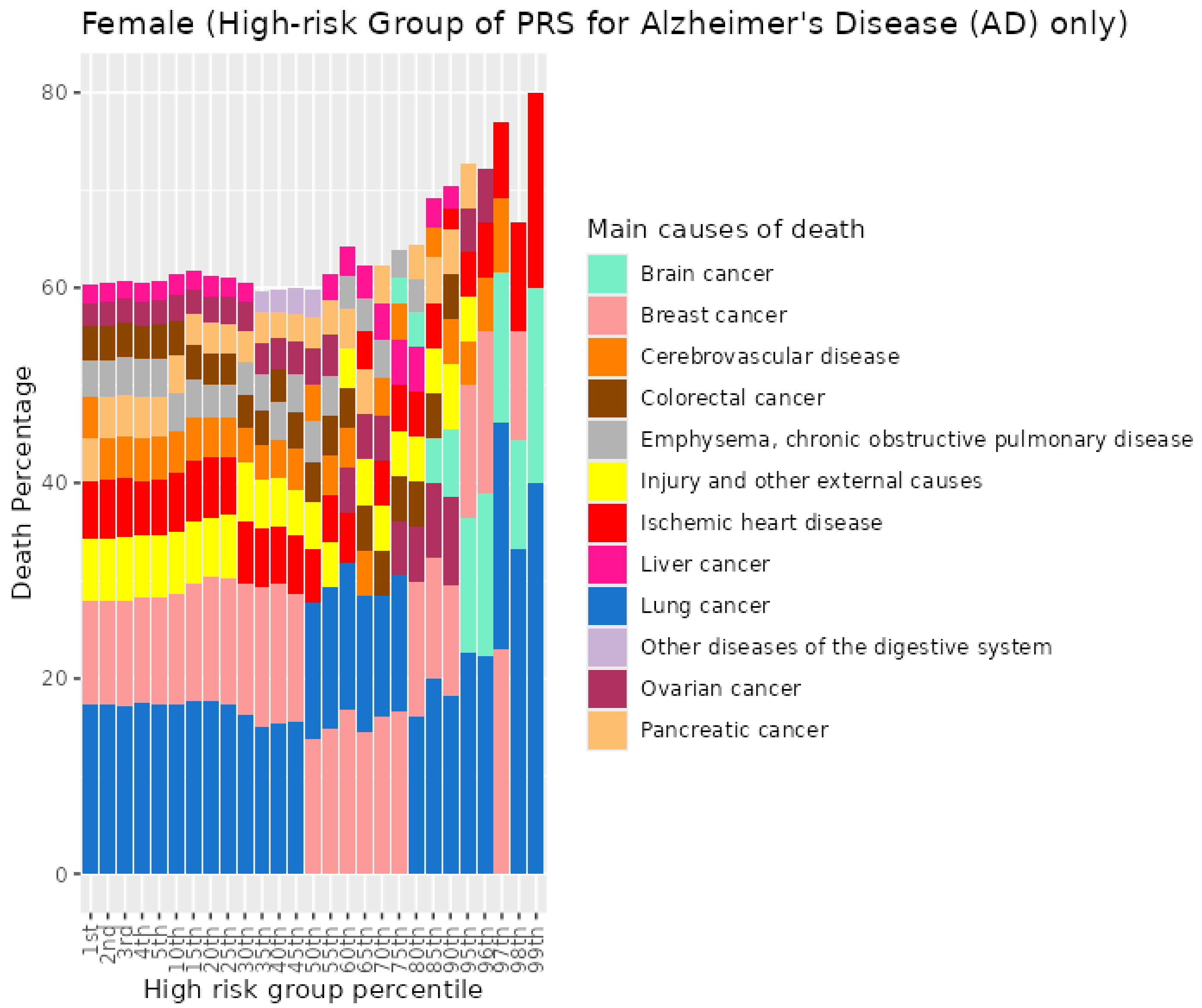


*Figure S7. Percentiles from 1 to 99 for Alzheimer's disease PRS among female current smokers who died at age <=59*

##### 1.8 Female current smokers who died at age 60-69

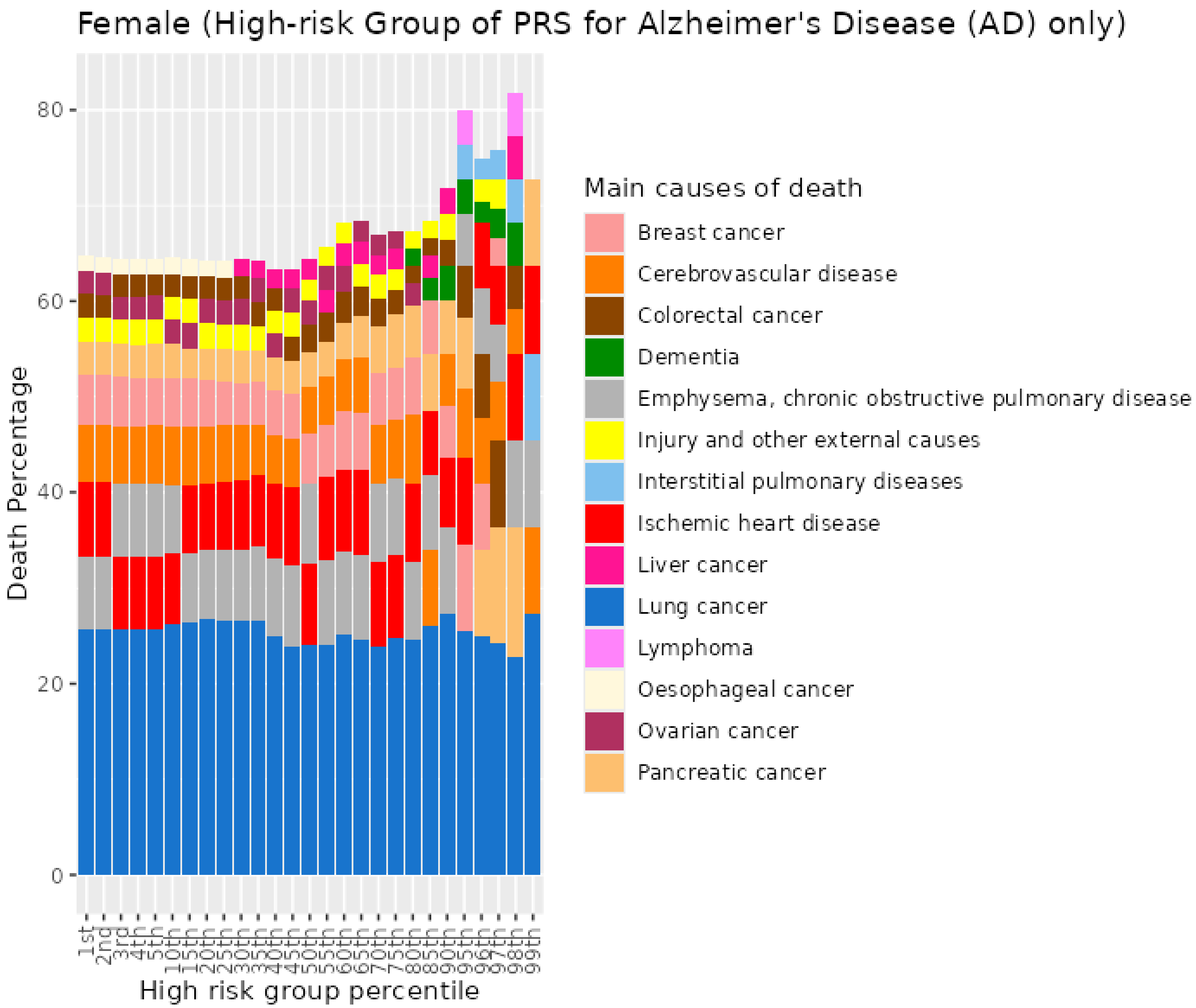


*Figure S8. Percentiles from 1 to 99 for Alzheimer's disease PRS among female current smokers who died at age 60-69*

##### 1.9 Female current smokers who died at age 70+

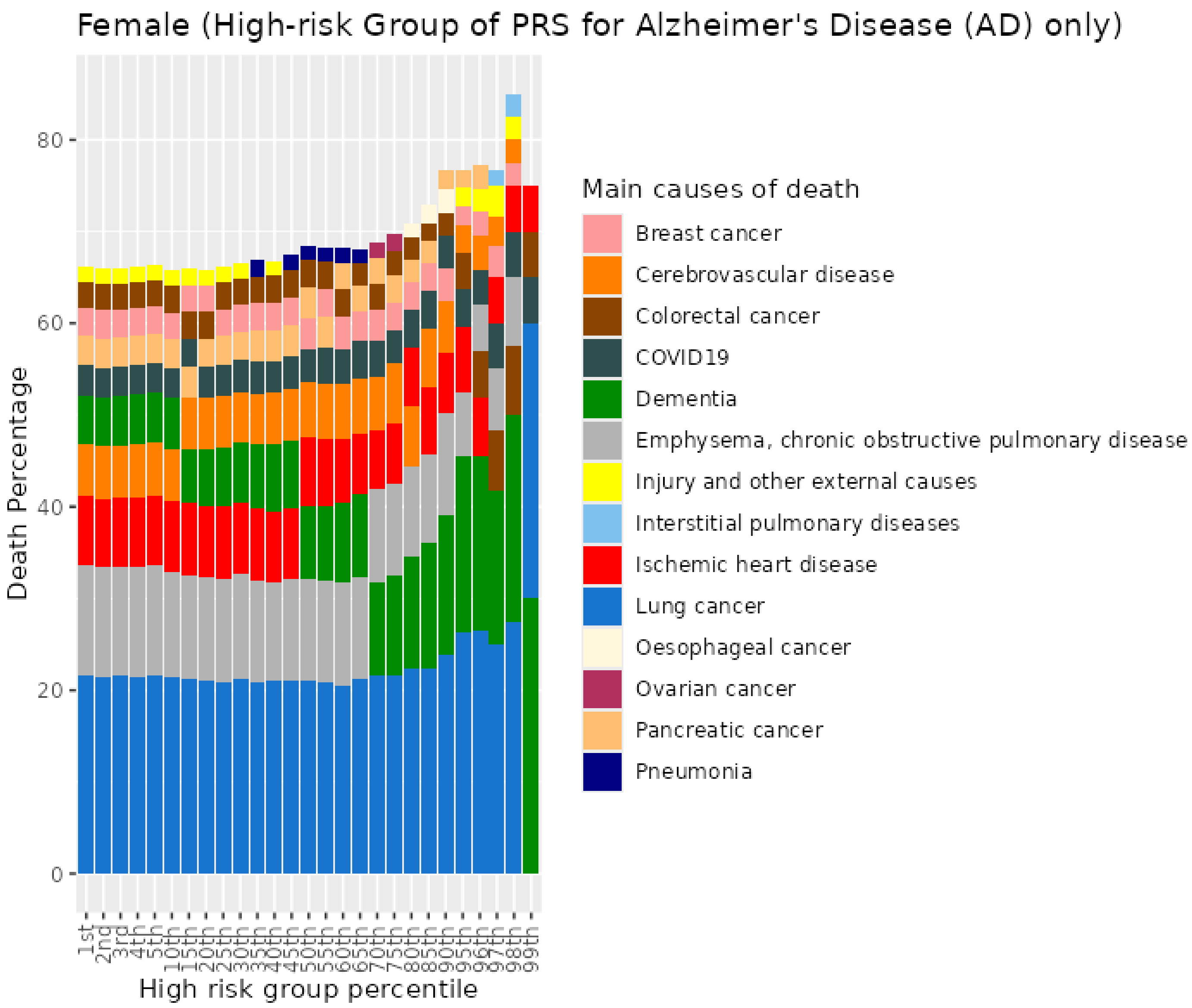


*Figure S9. Percentiles from 1 to 99 for Alzheimer's disease PRS among female current smokers who died at age 70+*

#### 2 Bowel Cancer (CRC) PRS for UKB females

##### 2.1 Female never smokers who died at age <=59

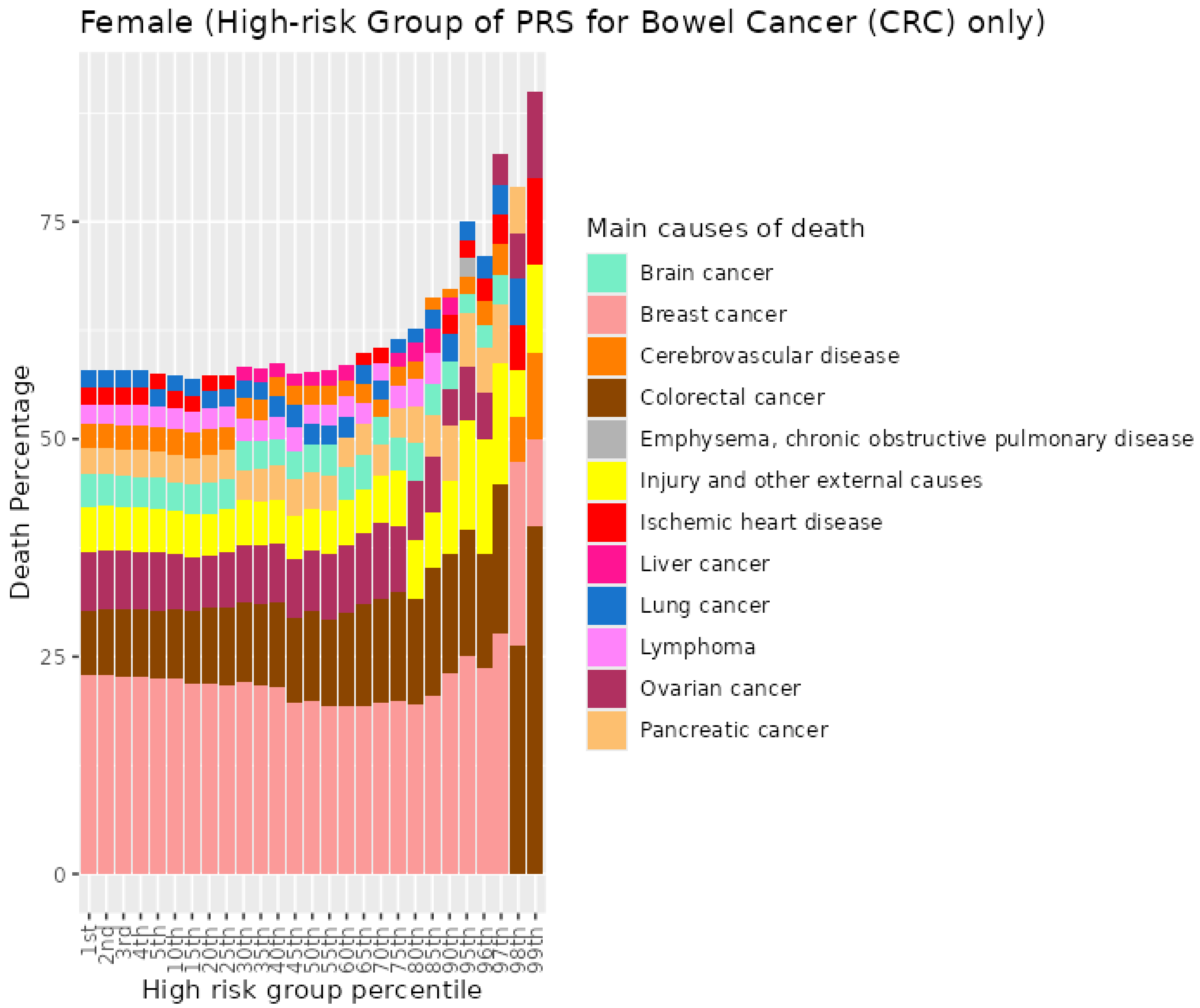


*Figure S10. Percentiles from 1 to 99 for bowel cancer PRS among females never smokers who died at age <=59*

##### 2.2 Female never smokers who died at age 60-69

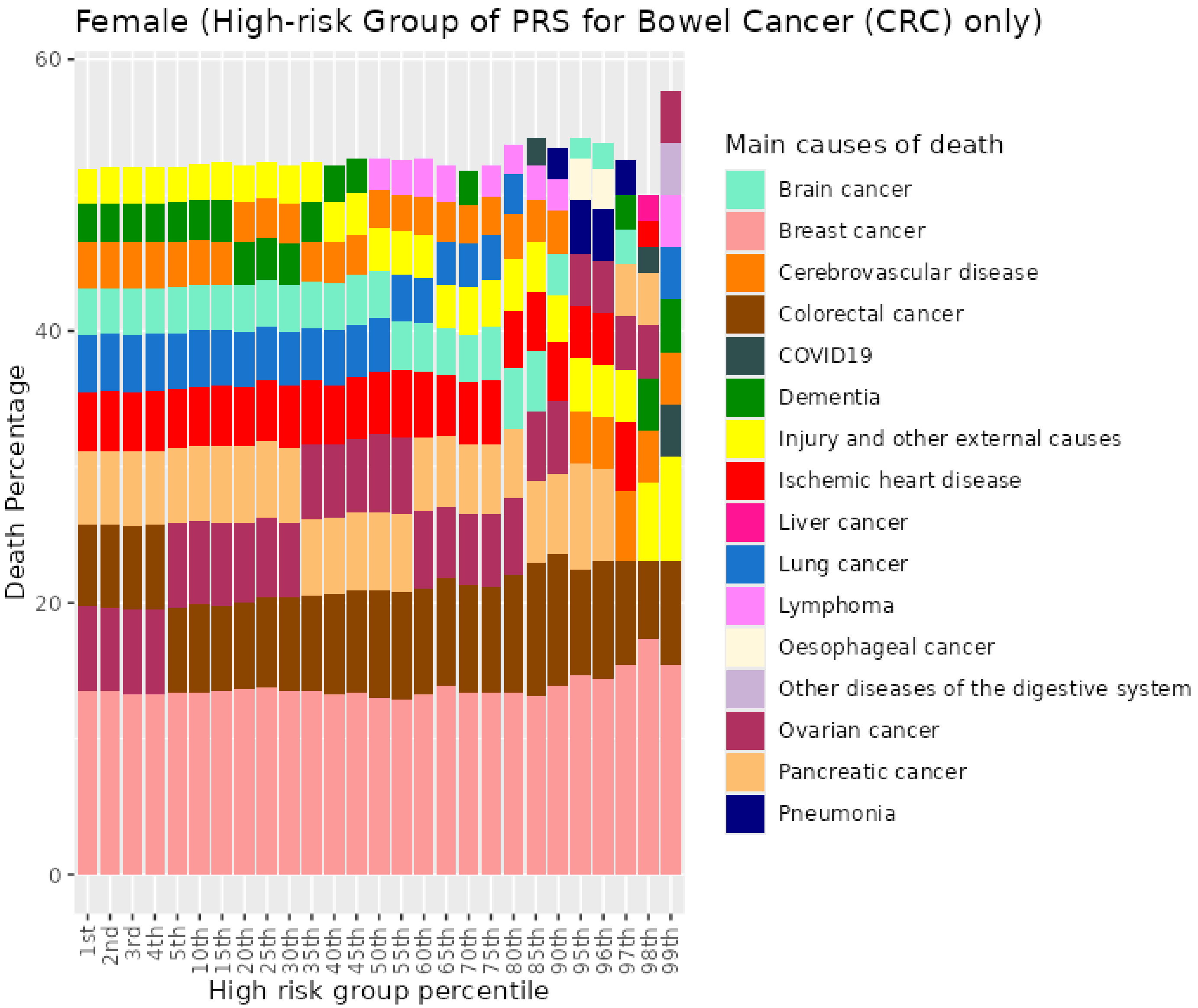


*Figure S11. Percentiles from 1 to 99 for bowel cancer PRS among female never smokers who died at age 60-69*

##### 2.3 Female never smokers who died at age 70+

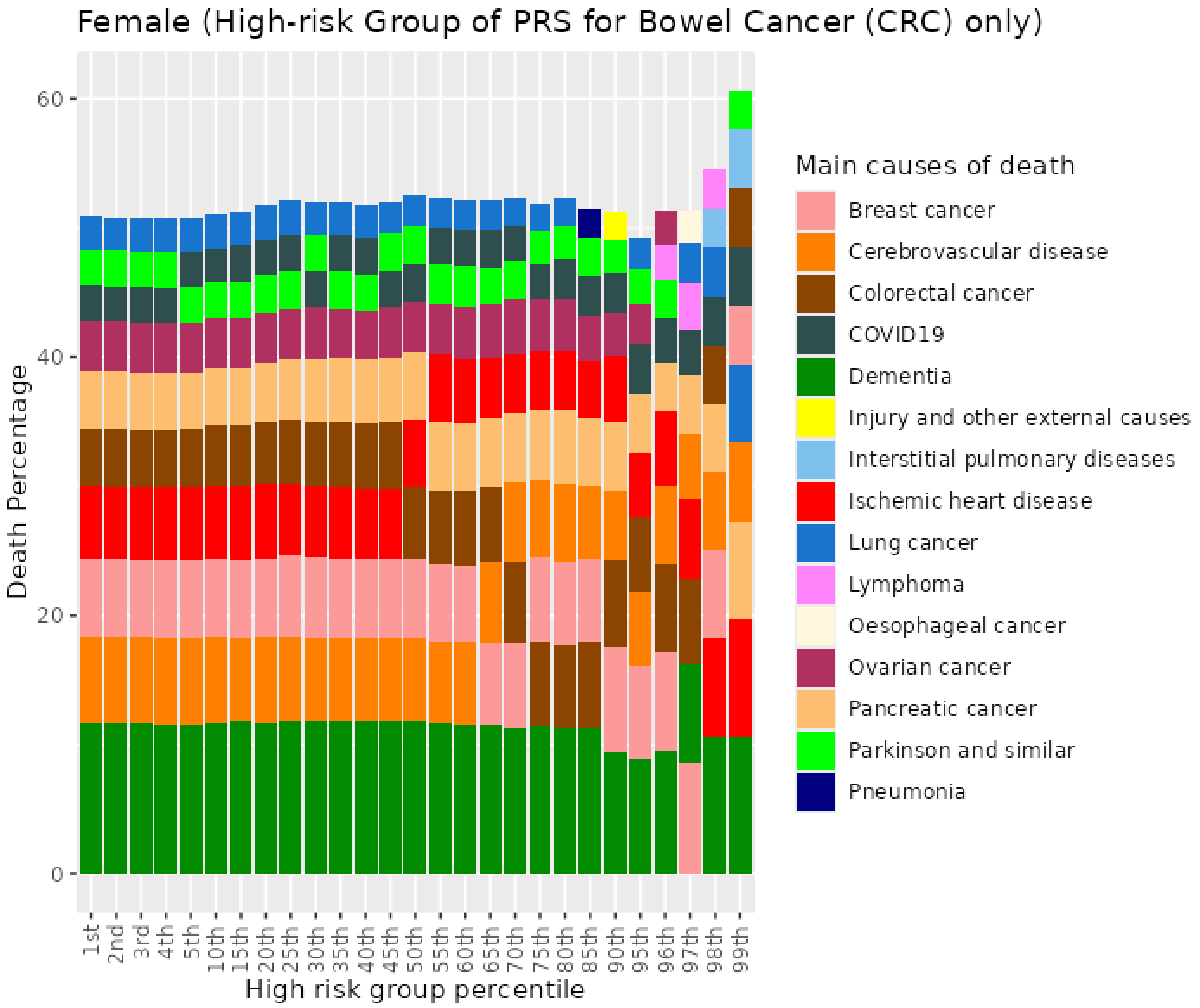


*Figure S12. Percentiles from 1 to 99 for bowel cancer PRS among female never smokers who died at age 70+*

#### 2.4 Female previous smokers who died at age <=59

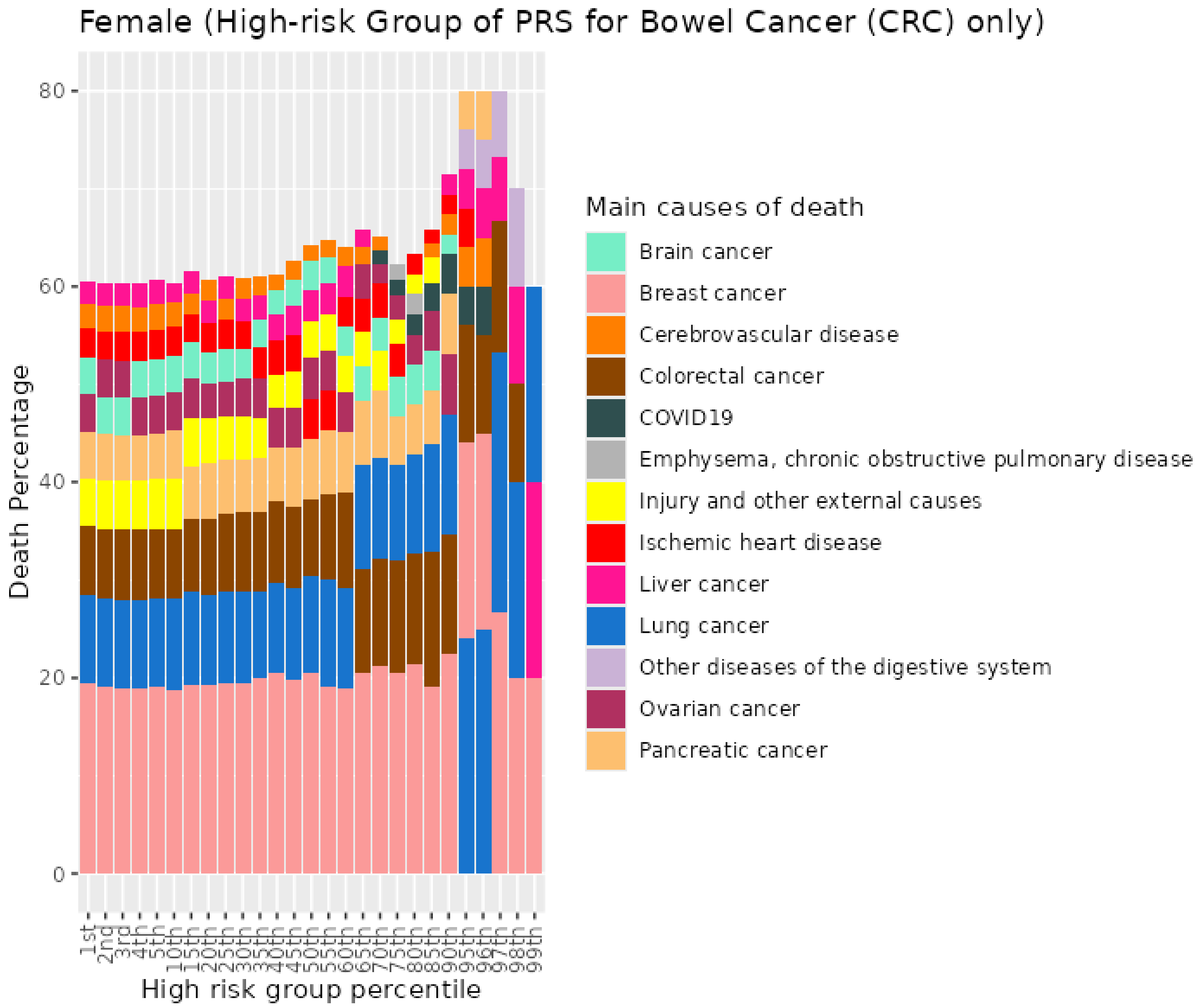


*Figure S13. Percentiles from 1 to 99 for bowel cancer PRS among female previous smokers who died at age <=59*

#### 2.5 Female previous smokers who died at age 60-69

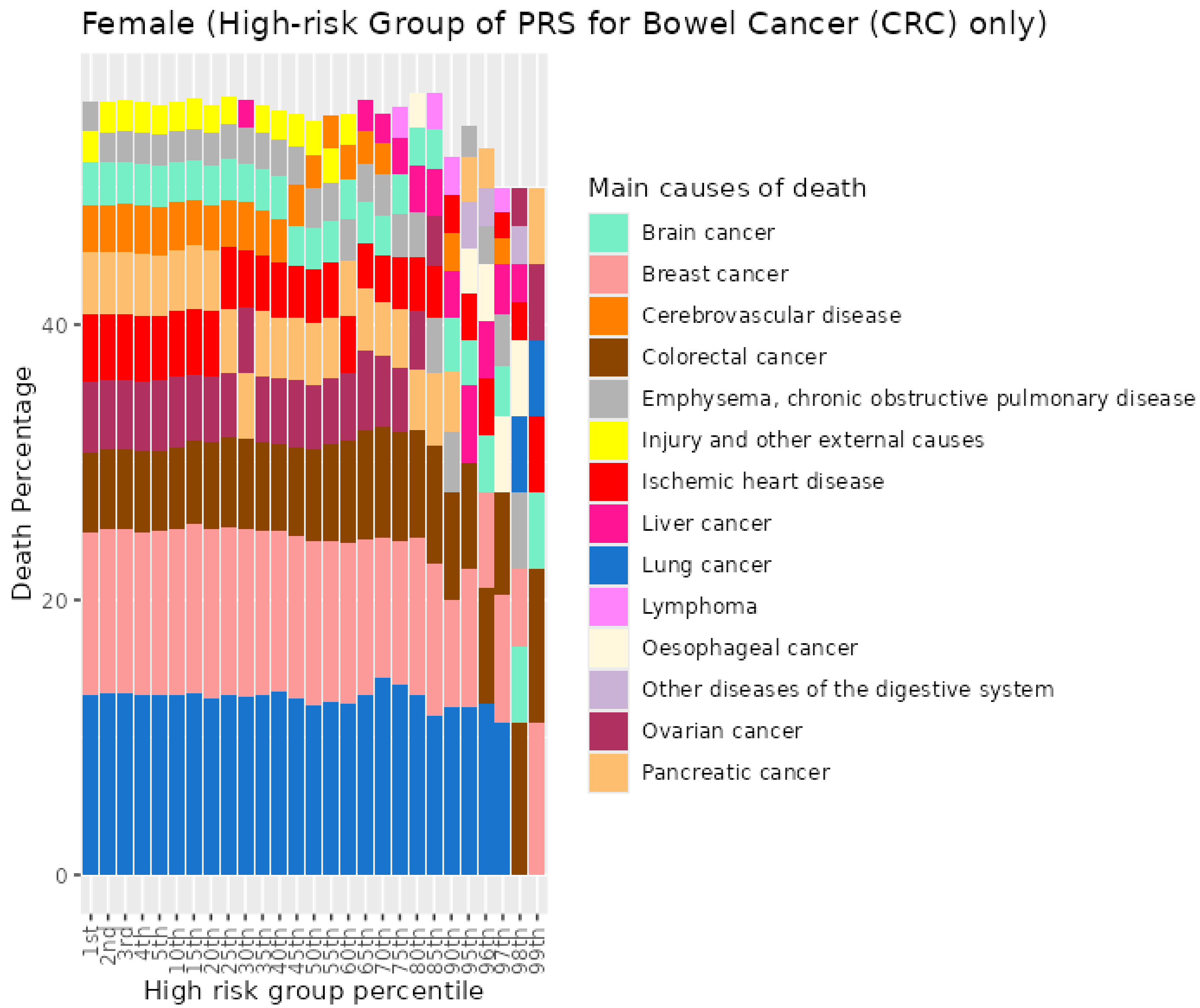


*Figure S14. Percentiles from 1 to 99 for bowel cancer PRS among female previous smokers who died at age 60-69*

#### 2.6 Female previous smokers who died at age 70+

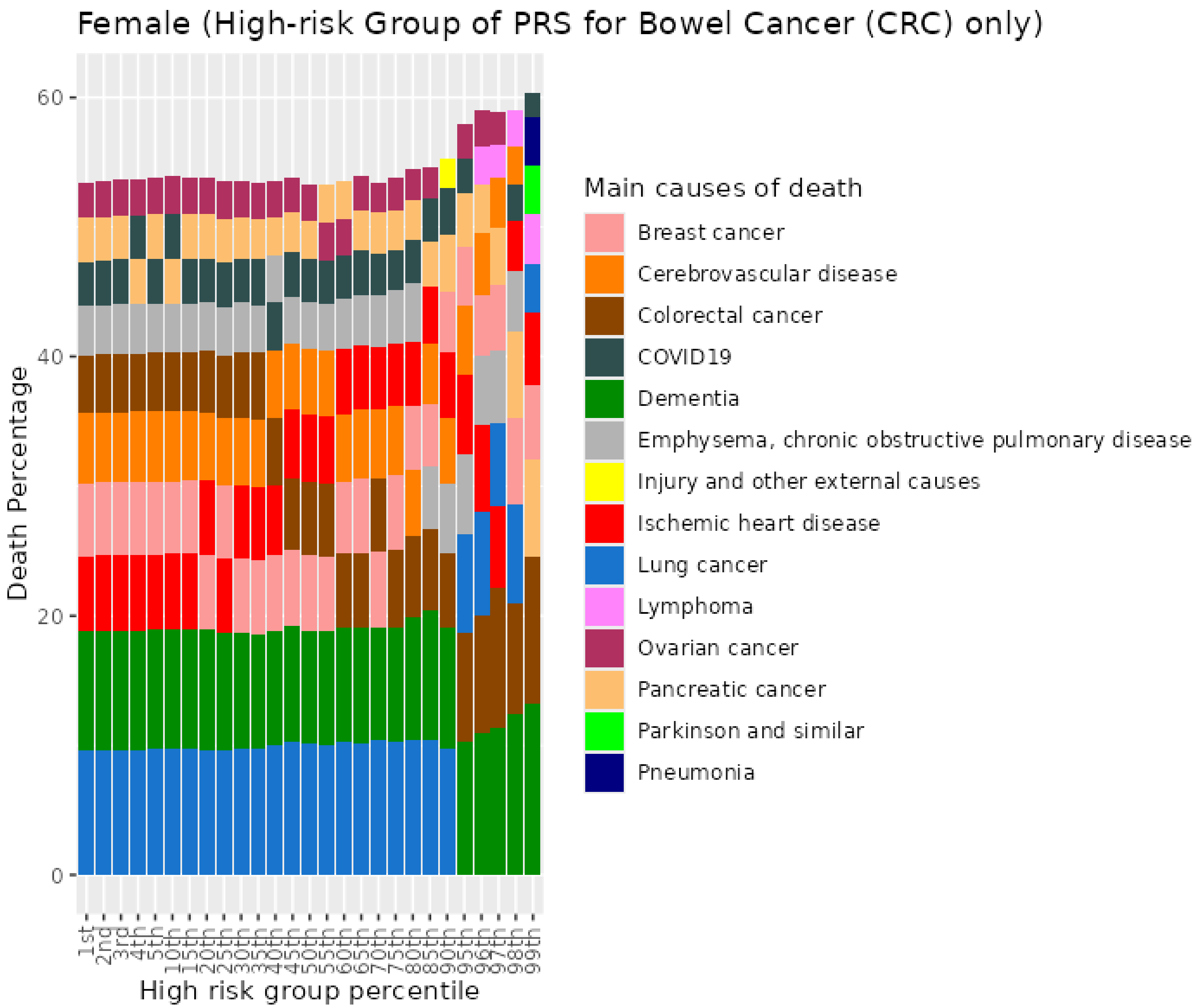


*Figure S15. Percentiles from 1 to 99 for bowel cancer PRS among female previous smokers who died at age 70+*

#### 2.7 Female current smokers who died at age <=59

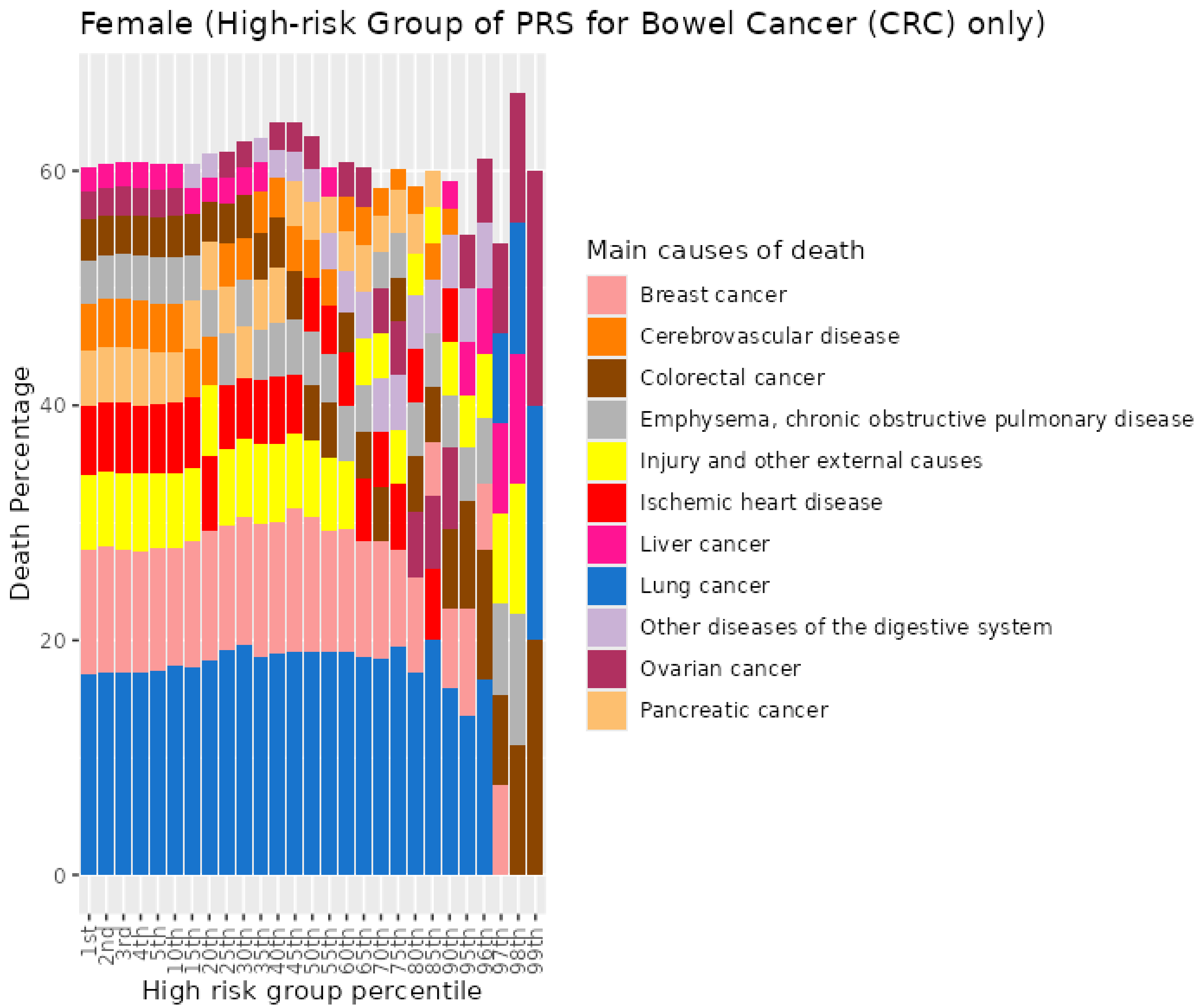


*Figure S16. Percentiles from 1 to 99 for bowel cancer PRS among female current smokers who died at age <=59*

#### 2.8 Female current smokers who died at age 60-69

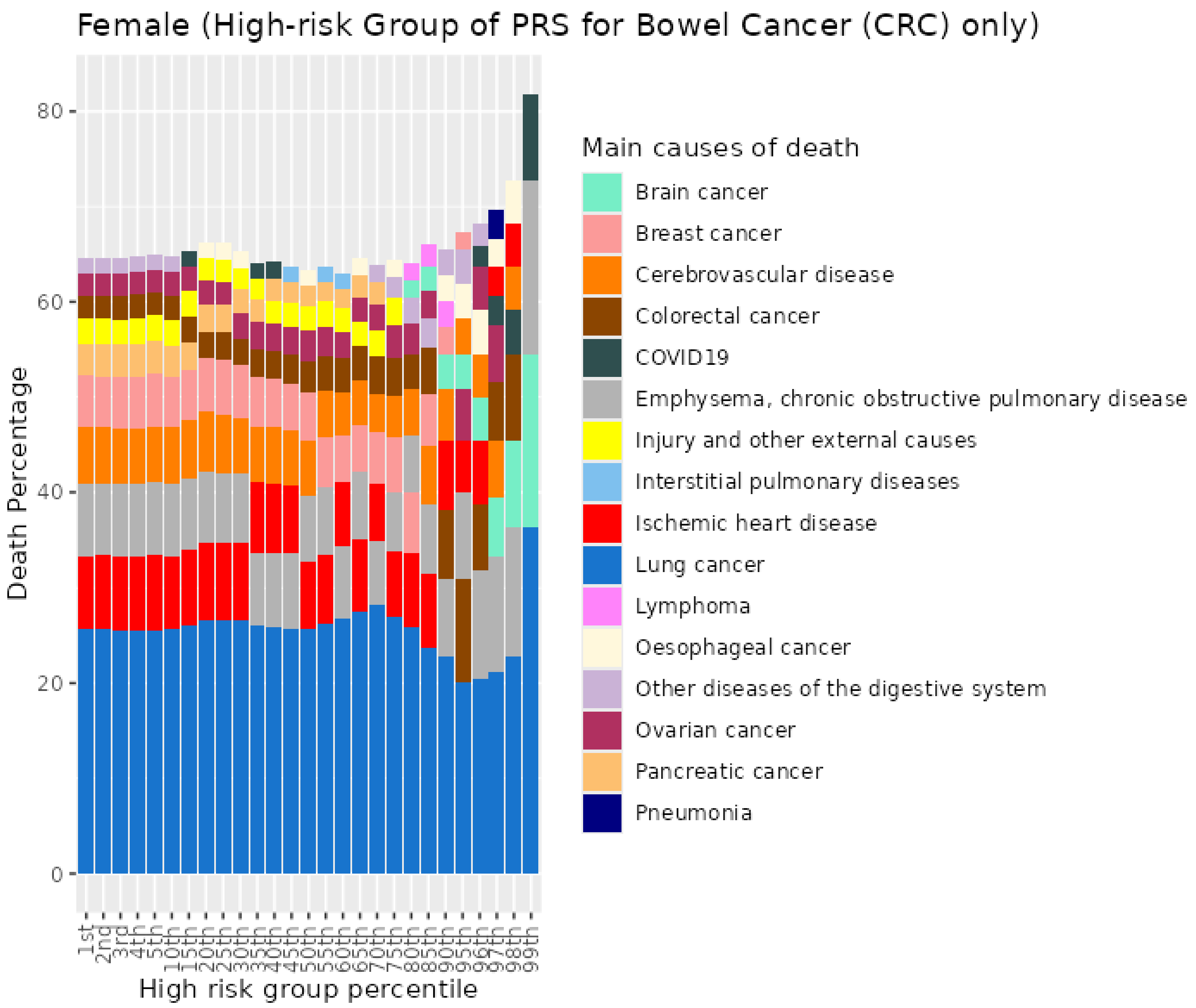


*Figure S17. Percentiles from 1 to 99 for bowel cancer PRS among female current smokers who died at age 60-69*

#### 2.9 Female current smokers who died at age 70+

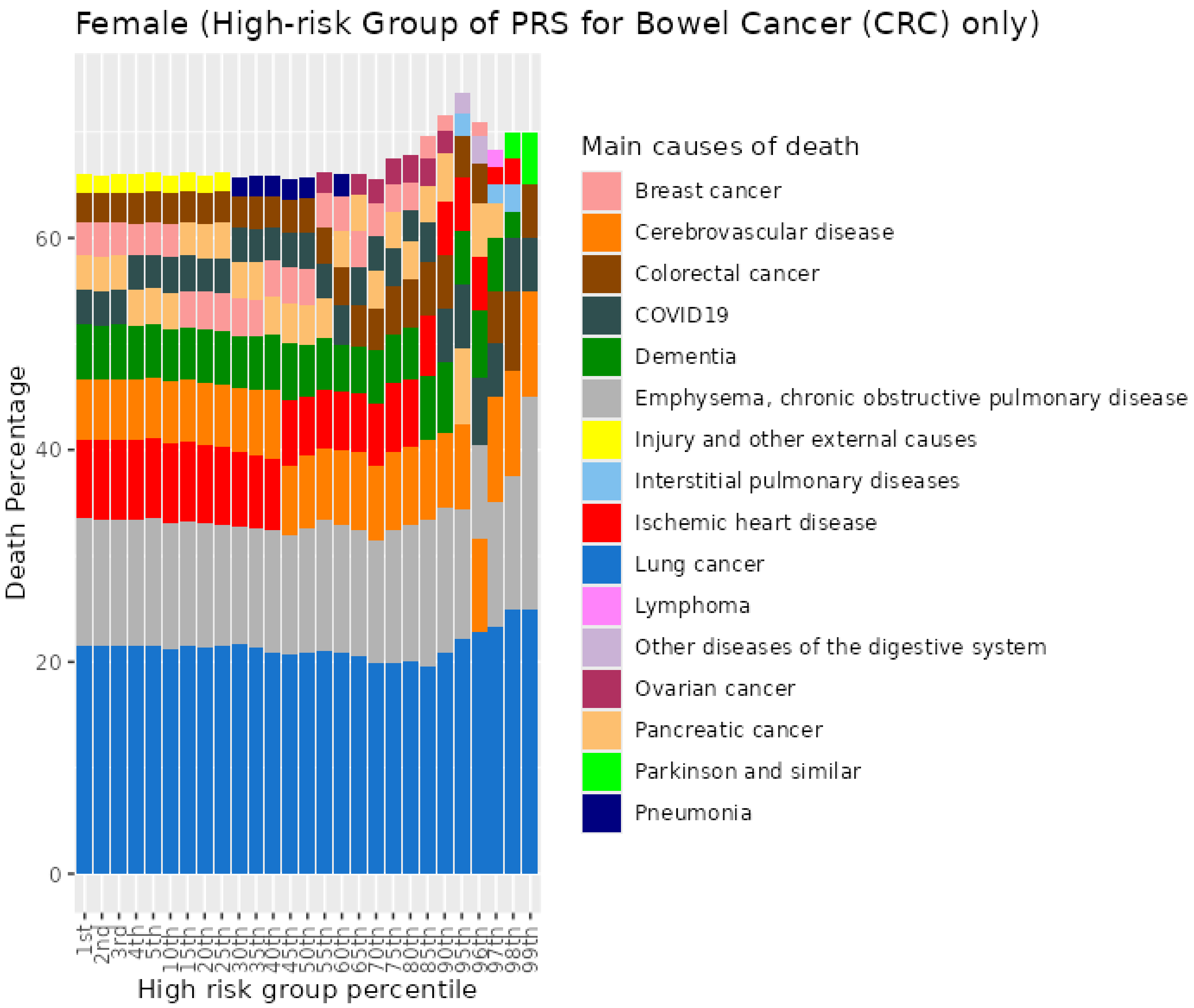


*Figure S18. Percentiles from 1 to 99 for bowel cancer PRS among female current smokers who died at age 70+*

### 3 Cardiovascular disease (CVD) PRS for UKB females

#### 3.1 Female never smokers who died at age <=59

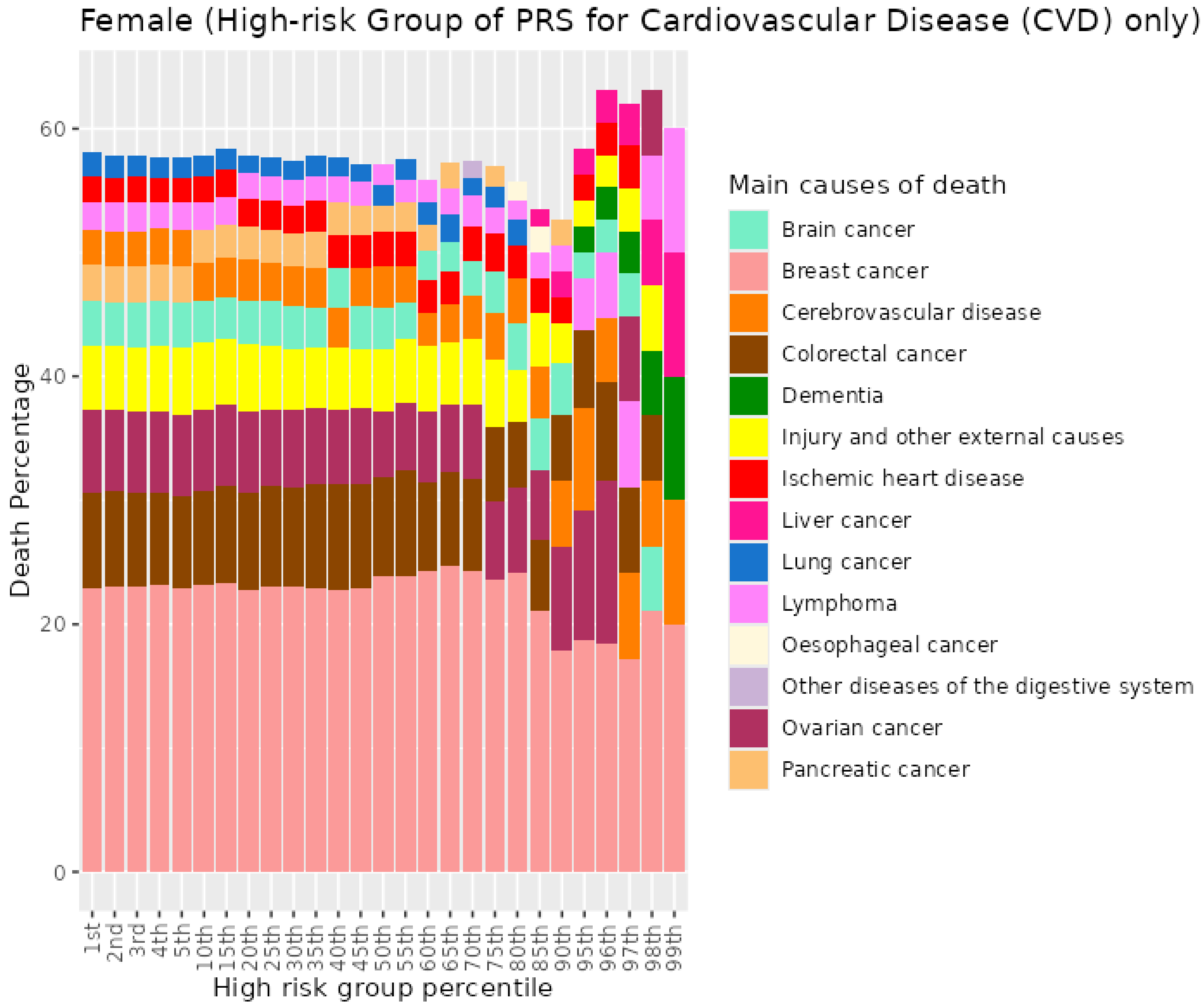


*Figure S19. Percentiles from 1 to 99 for cardiovascular disease PRS among female never smokers who died at age <=59*

##### 3.2 Female never smokers who died at age 60-69

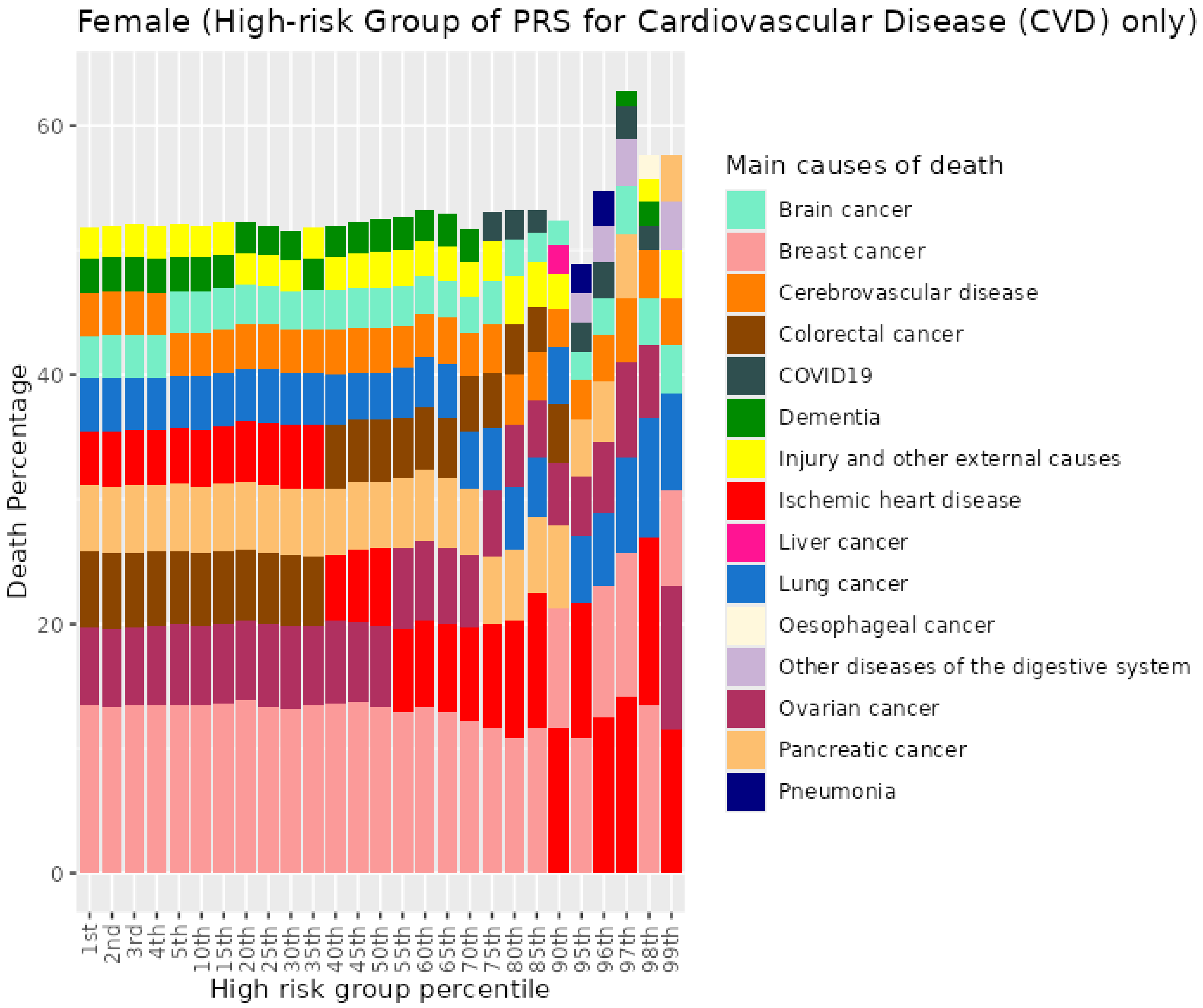


*Figure S20. Percentiles from 1 to 99 for cardiovascular disease PRS among female never smokers who died at age 60-69*

##### 3.3 Female never smokers who died at age 70+

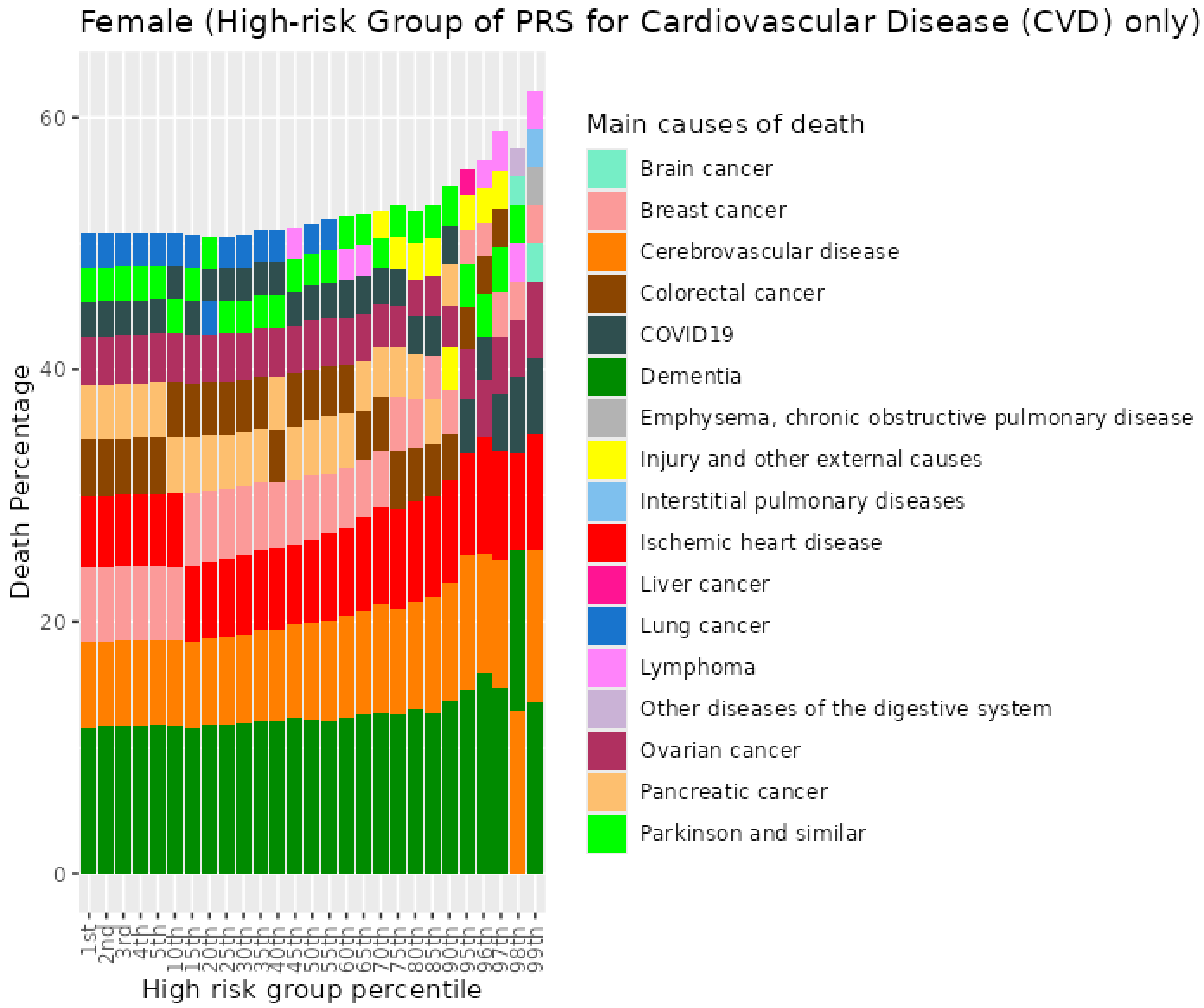


*Figure S21. Percentiles from 1 to 99 for cardiovascular disease PRS among female never smokers who died at age 70+*

##### 3.4 Female previous smokers who died at age <=59

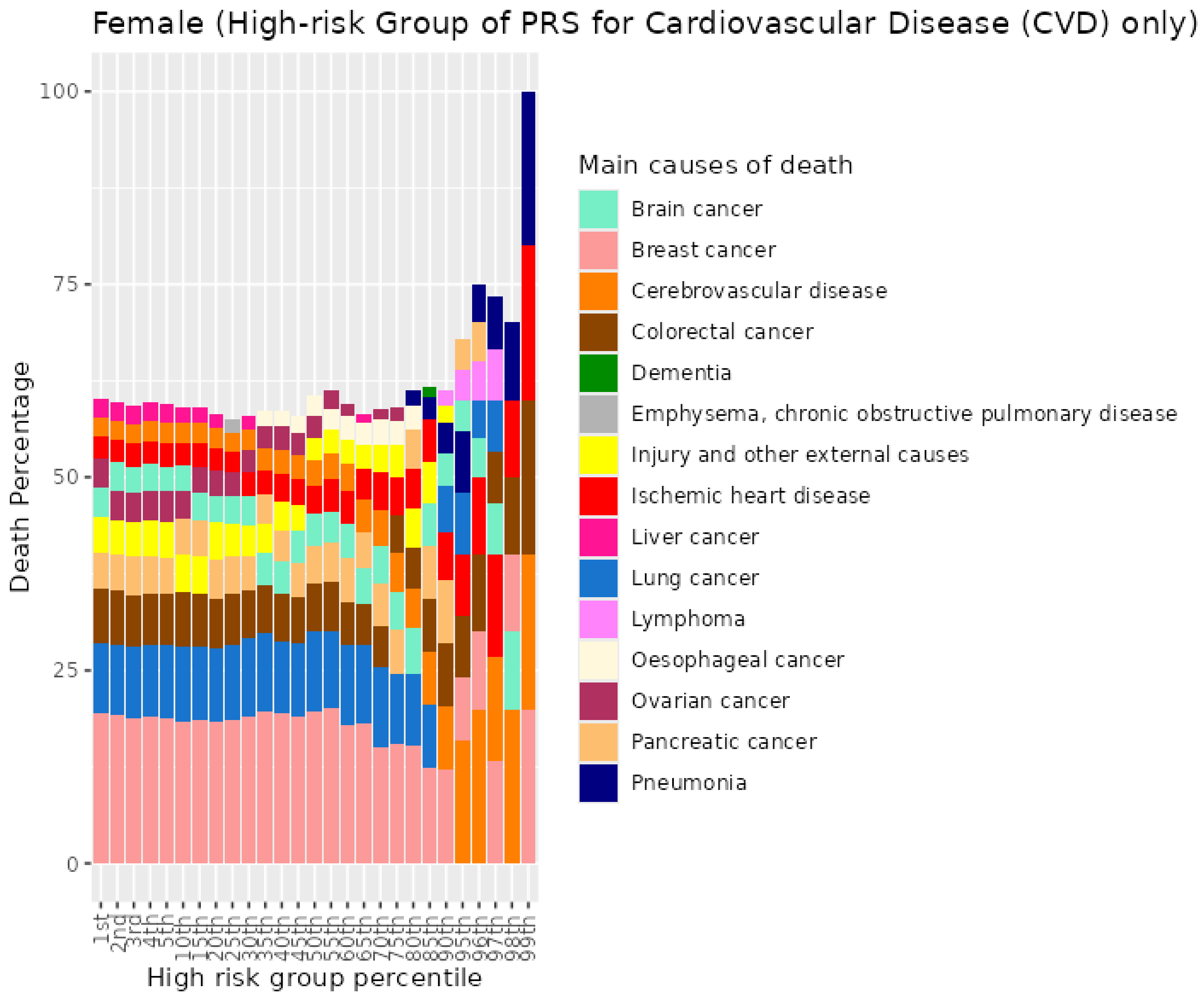


*Figure S22. Percentiles from 1 to 99 for cardiovascular disease PRS among female previous smokers who died at age <=59*

##### 3.5 Female previous smokers who died at age 60-69

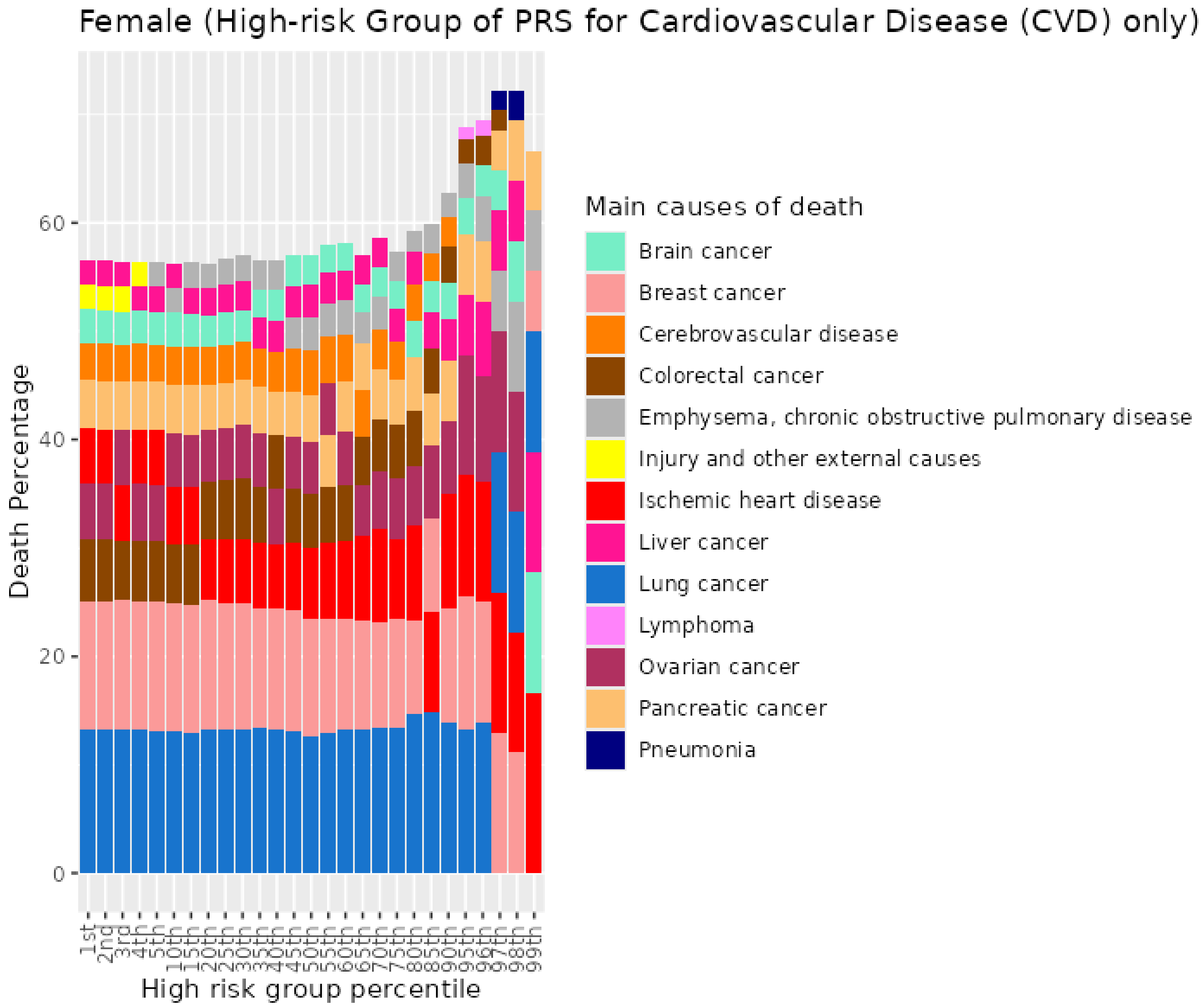


*Figure S23. Percentiles from 1 to 99 for cardiovascular disease PRS among female previous smokers who died at age 60-69*

##### 3.6 Female previous smokers who died at age 70+

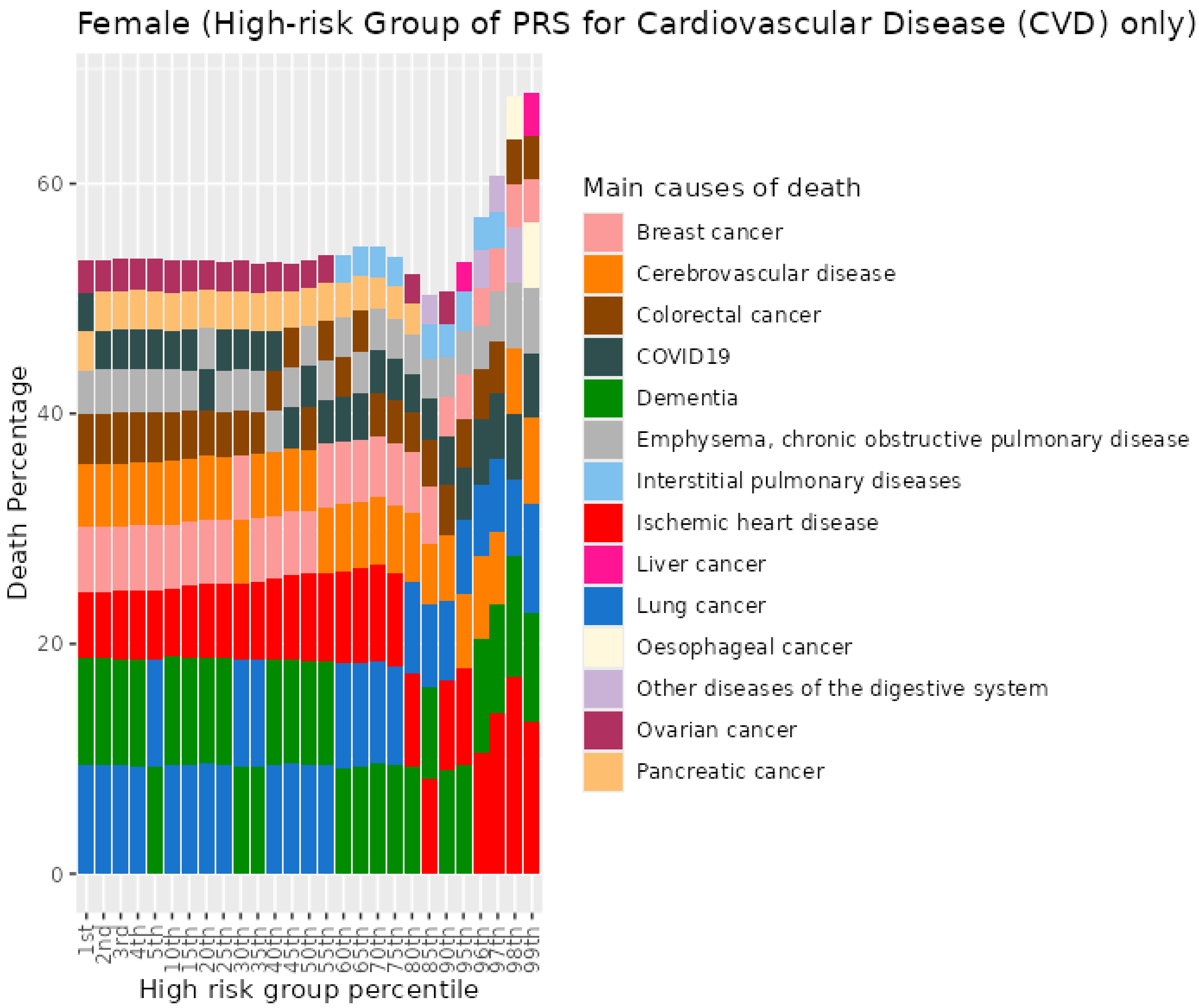


*Figure S24. Percentiles from 1 to 99 for cardiovascular disease PRS among female previous smokers who died at age 70+*

##### 3.7 Female current smokers who died at age <=59

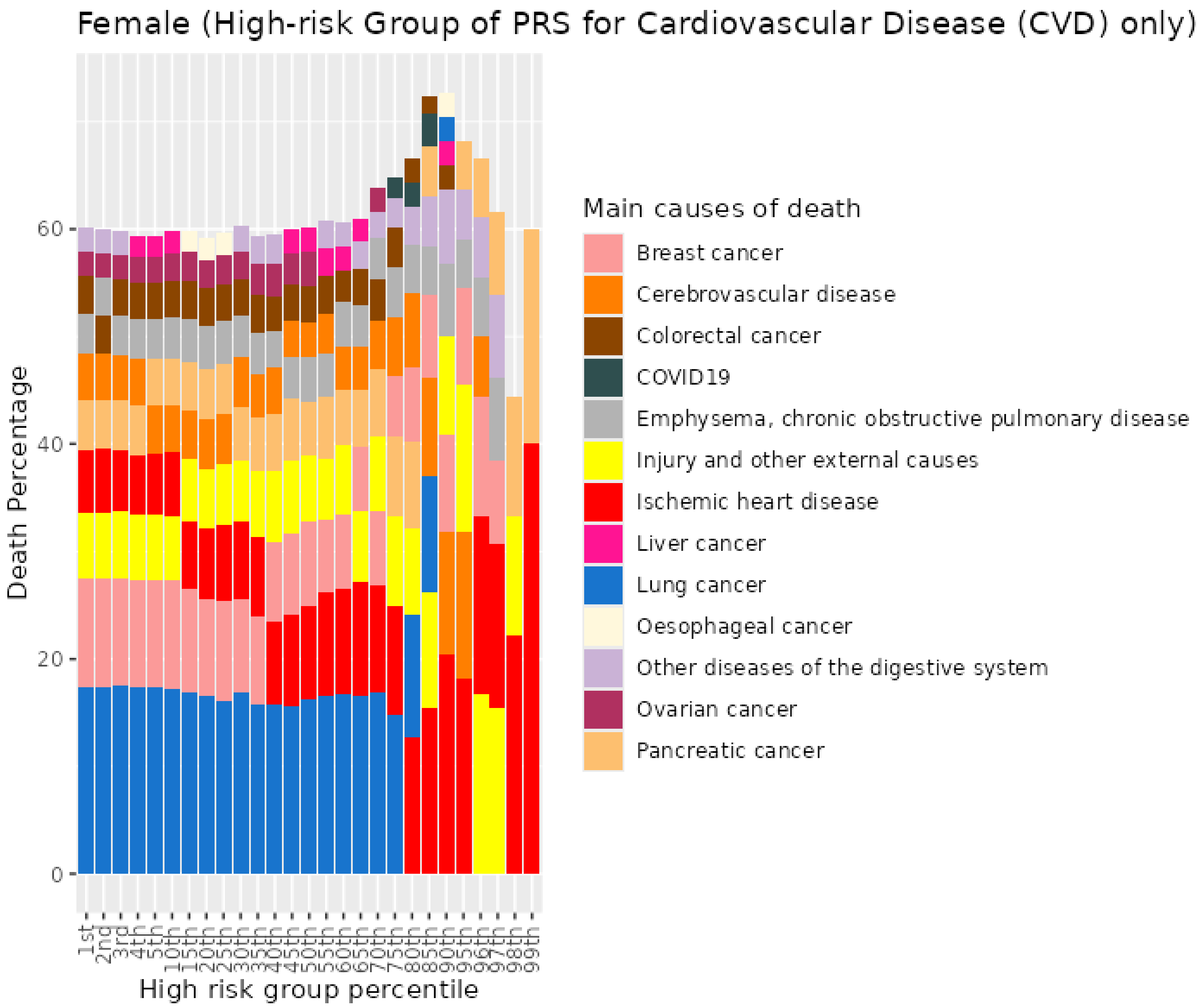


*Figure S25. Percentiles from 1 to 99 for cardiovascular disease PRS among female current smokers who died at age <=59*

##### 3.8 Female current smokers who died at age 60-69

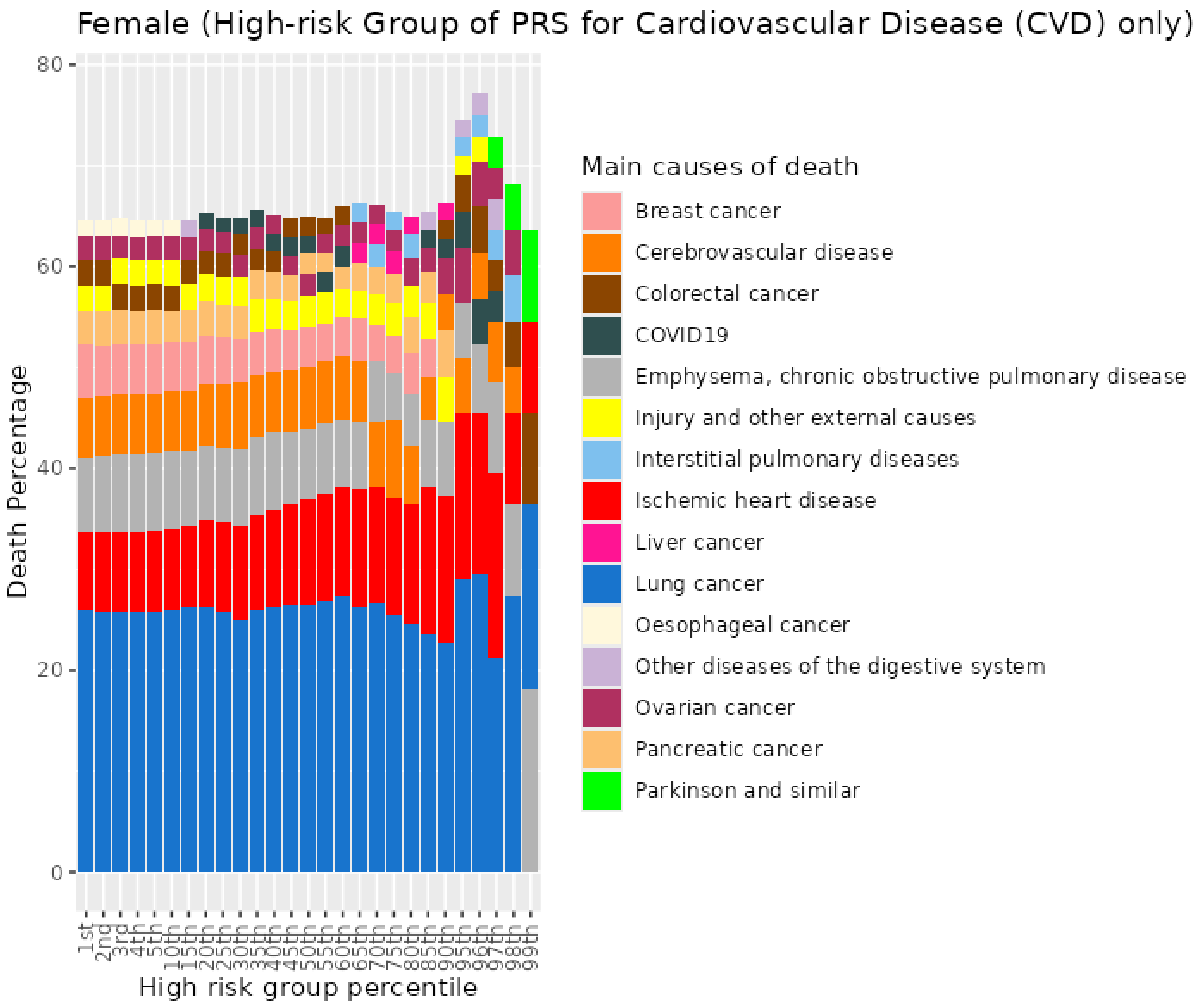


*Figure S26. Percentiles from 1 to 99 for cardiovascular disease PRS among female current smokers who died at age 60-69*

##### 3.9 Female current smokers who died at age 70+

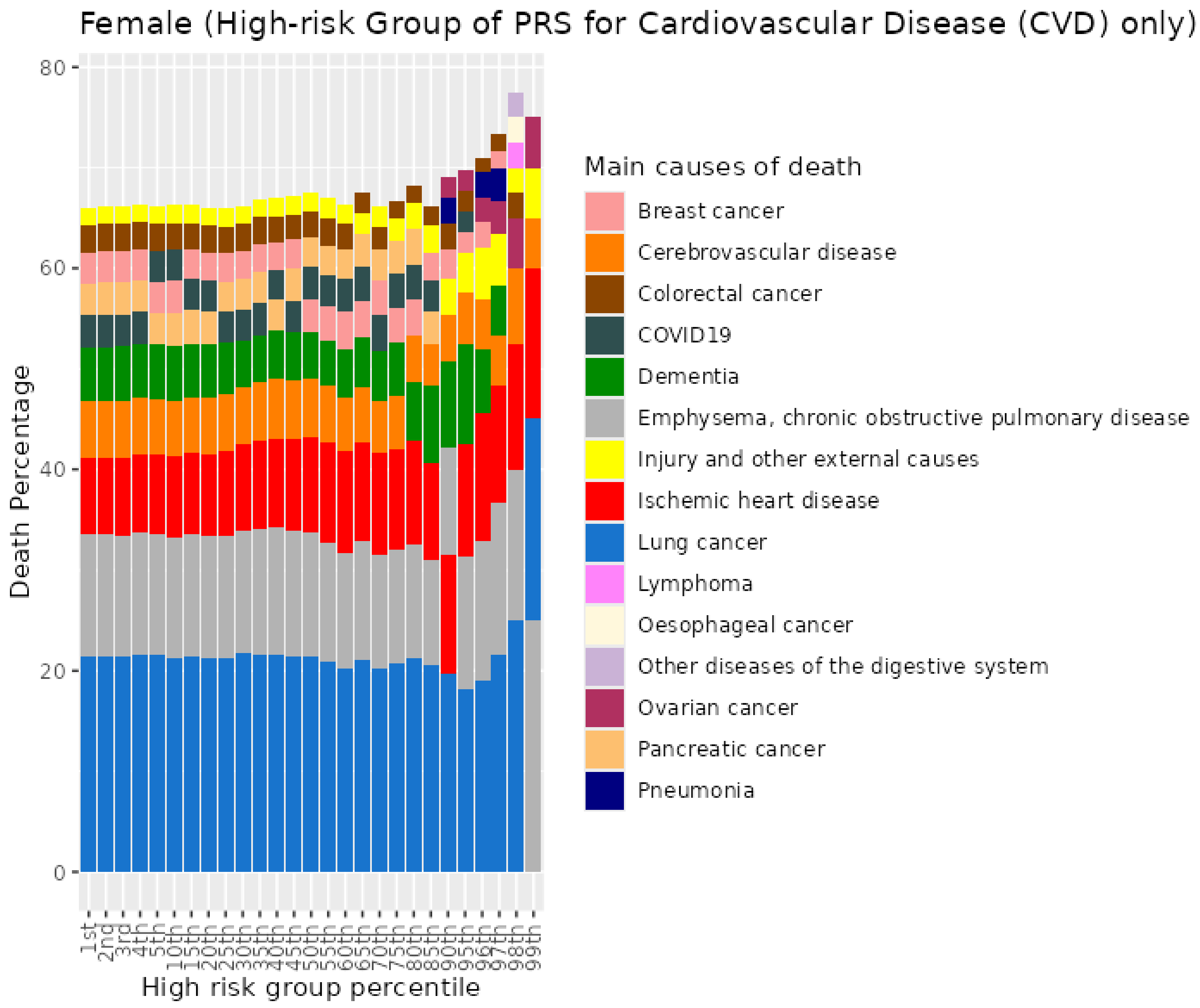


*Figure S27. Percentiles from 1 to 99 for cardiovascular disease PRS among female current smokers who died at age 70+*

### 4 Coronary artery disease (CAD) PRS for UKB females

#### 4.1 Female never smokers who died at age <=59

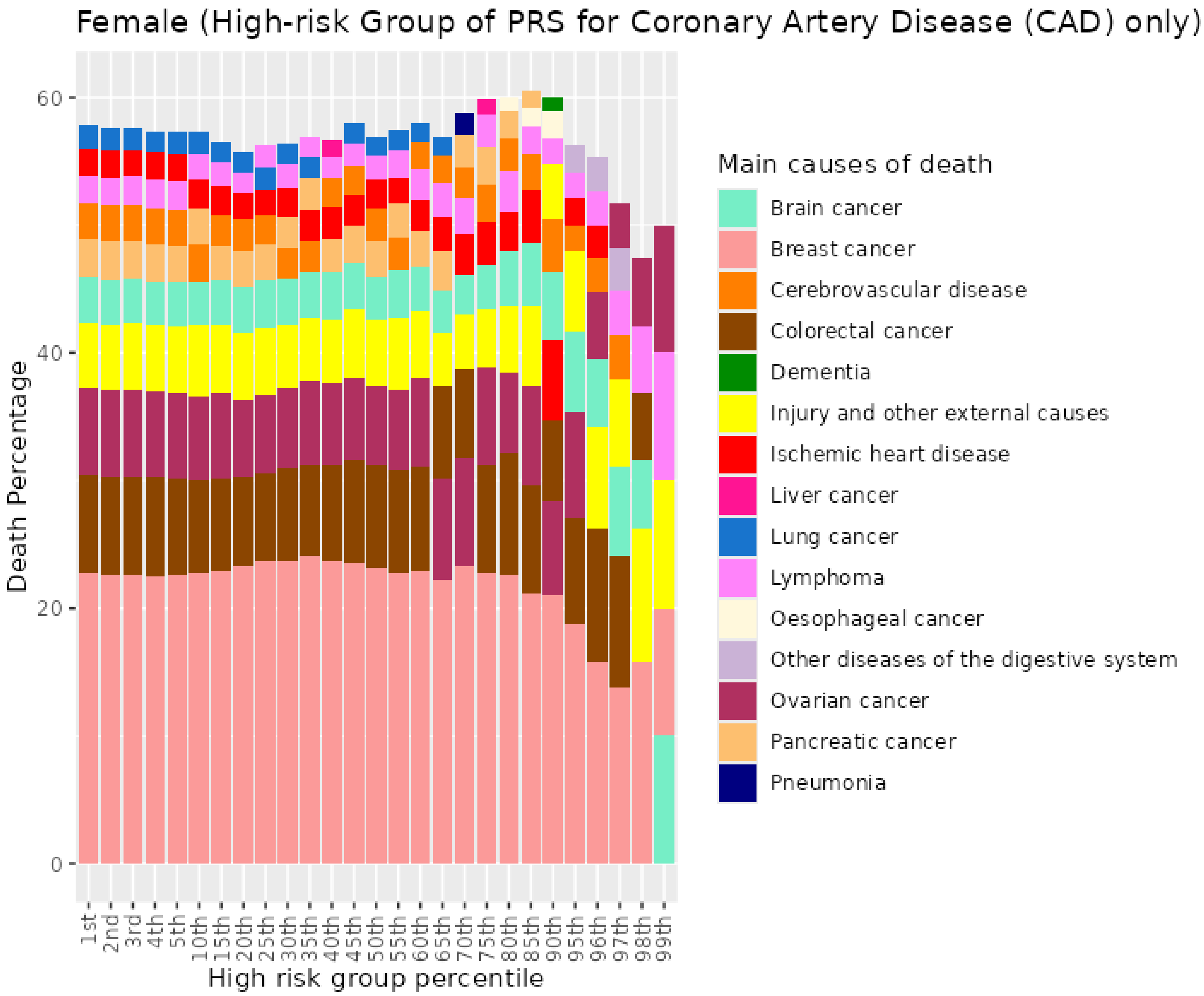


*Figure S28. Percentiles from 1 to 99 for coronary artery disease PRS among female never smokers who died at age <=59*

##### 4.2 Female never smokers who died at age 60-69

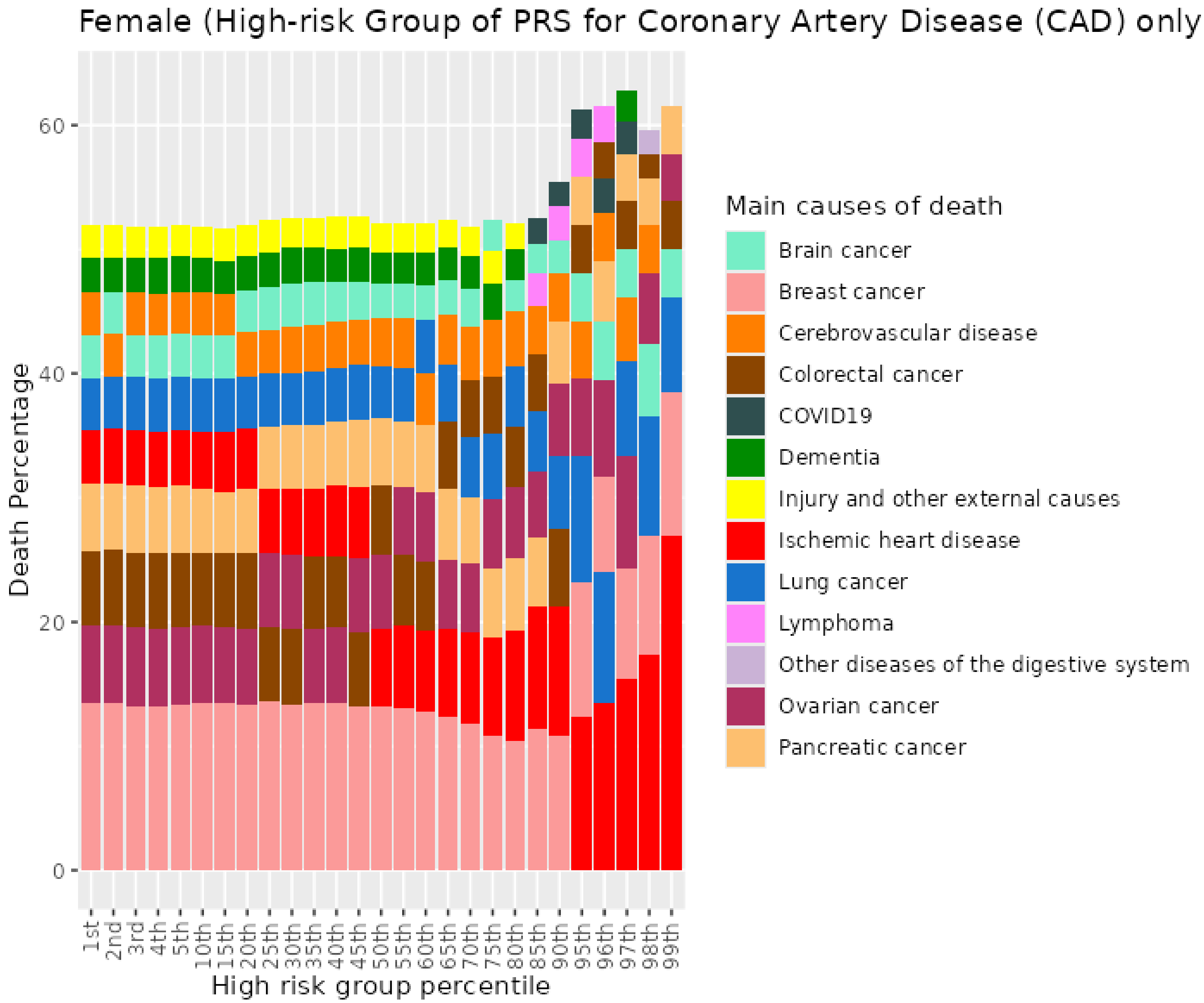


*Figure S29. Percentiles from 1 to 99 for coronary artery disease PRS among female never smokers who died at age 60-69*

##### 4.3 Female never smokers who died at age 70+

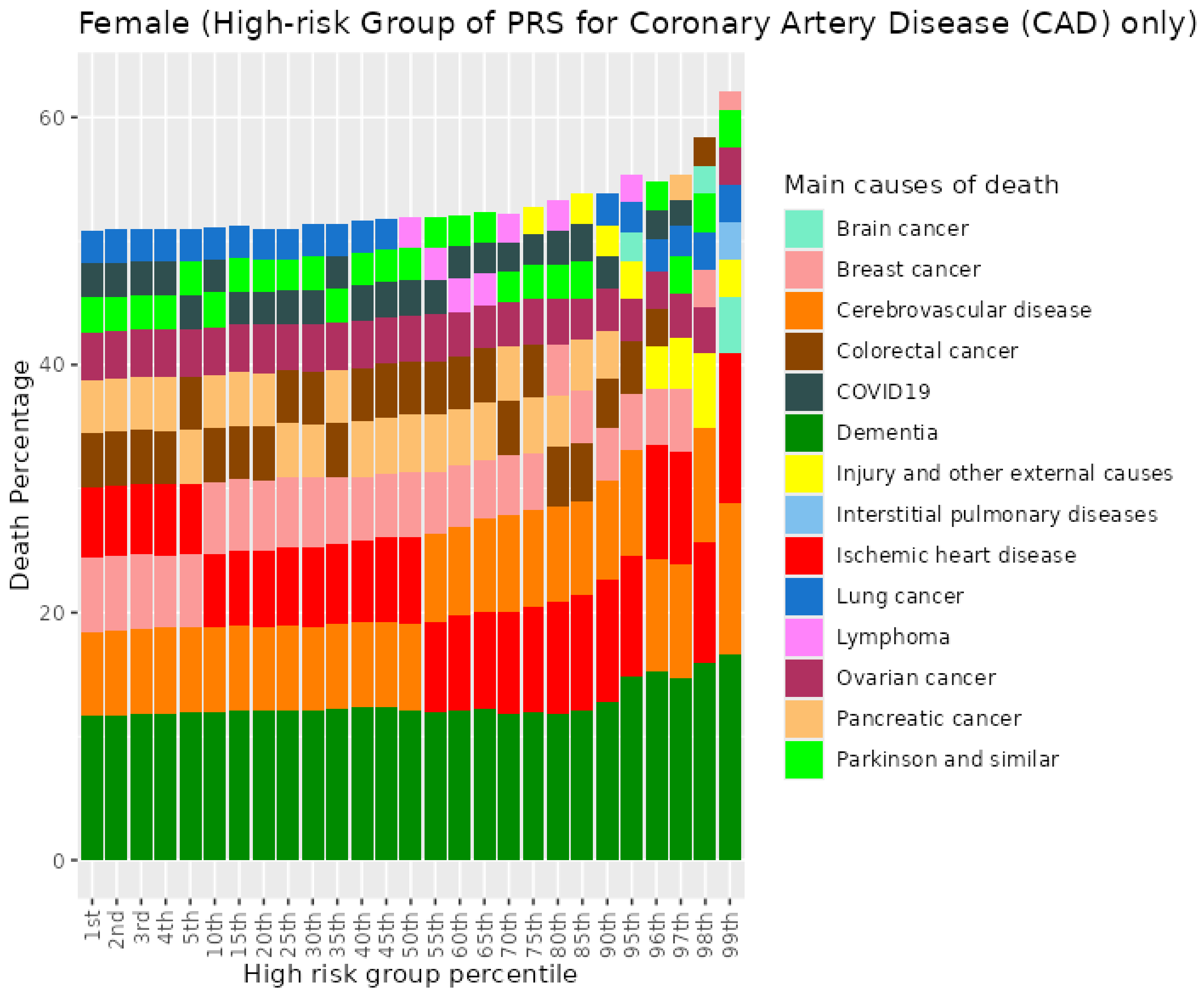


*Figure S30. Percentiles from 1 to 99 for coronary artery disease PRS among female never smokers who died at age 70+*

##### 4.4 Female previous smokers who died at age <=59

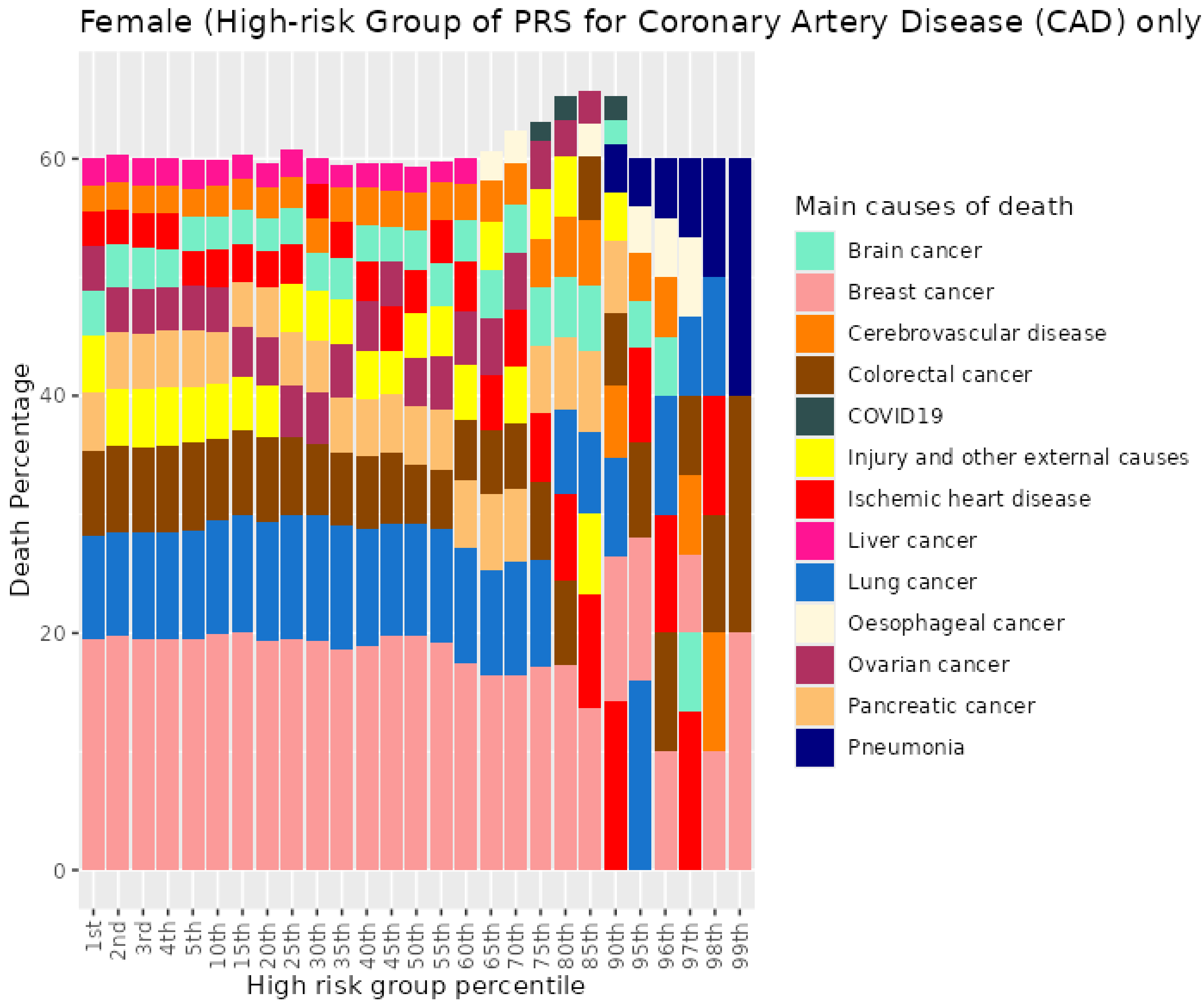


*Figure S31. Percentiles from 1 to 99 for coronary artery disease PRS among female previous smokers who died at age <=59*

##### 4.5 Female previous smokers who died at age 60-69

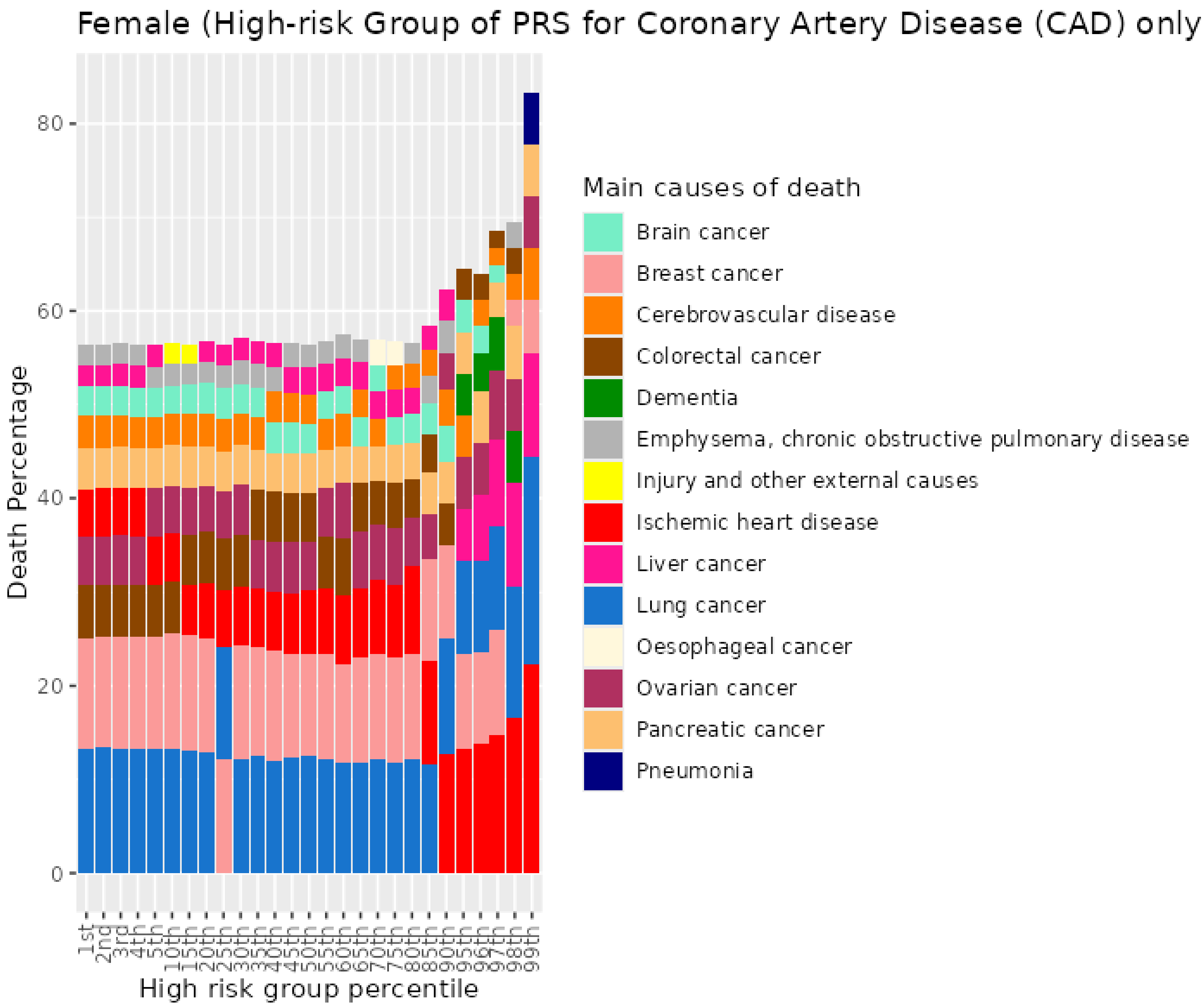


*Figure S32. Percentiles from 1 to 99 for coronary artery disease PRS among female previous smokers who died at age 60-69*

##### 4.6 Female previous smokers who died at age 70+

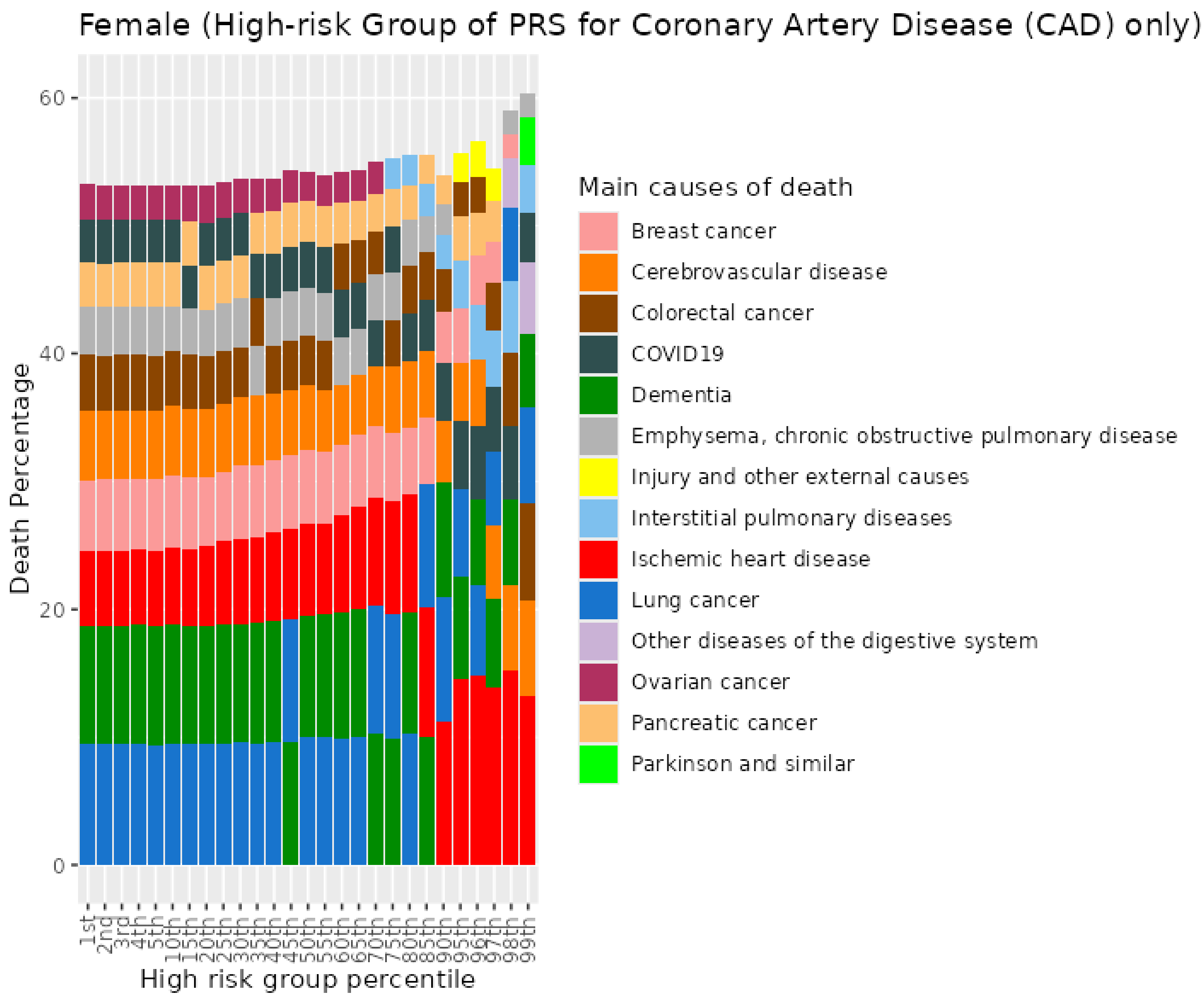


*Figure S33. Percentiles from 1 to 99 for coronary artery disease PRS among female previous smokers who died at age 70+*

##### 4.7 Female current smokers who died at age <=59

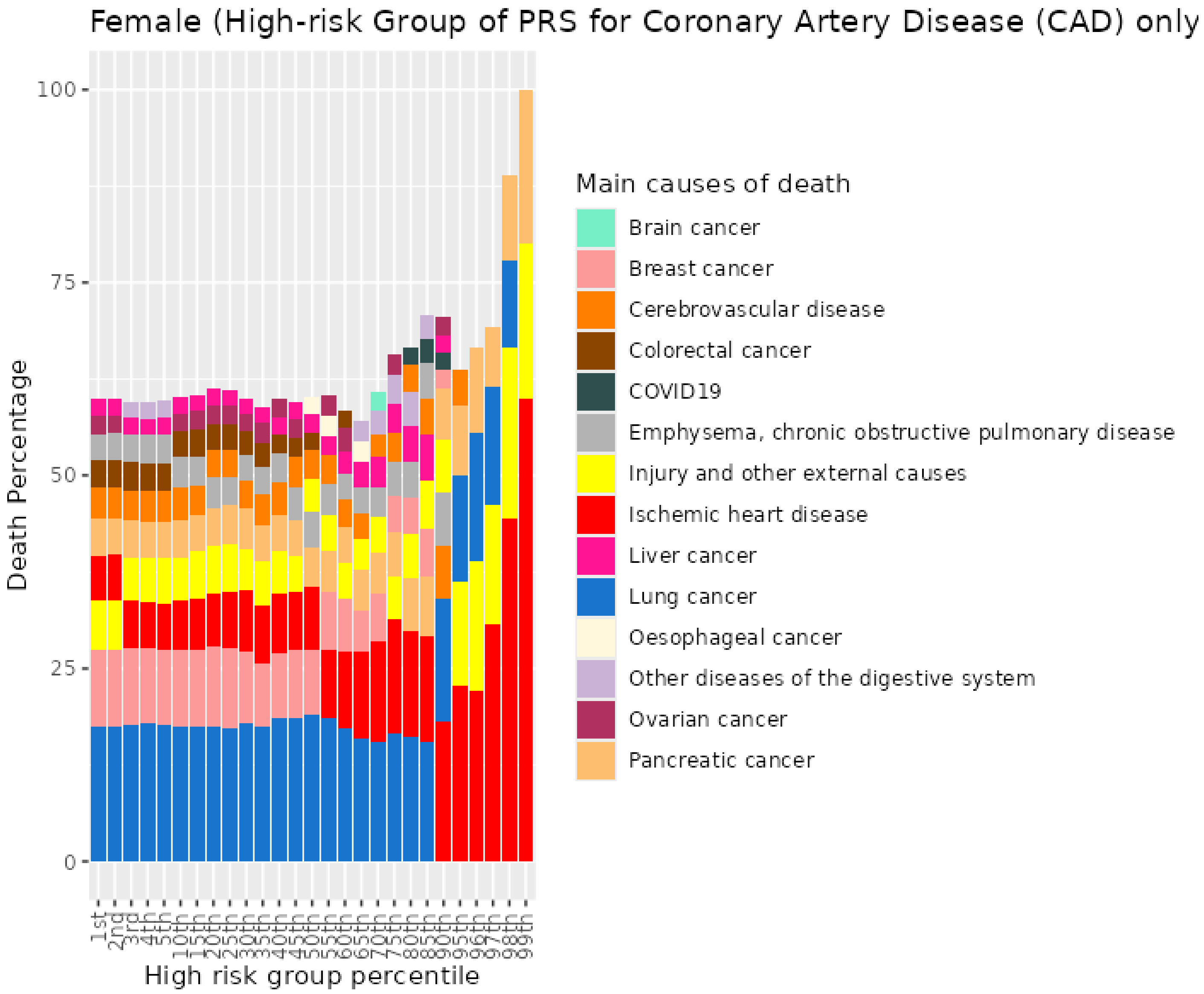


*Figure S34. Percentiles from 1 to 99 for coronary artery disease PRS among female current smokers who died at age <=59*

#### 4.8 Female current smokers who died at age 60-69

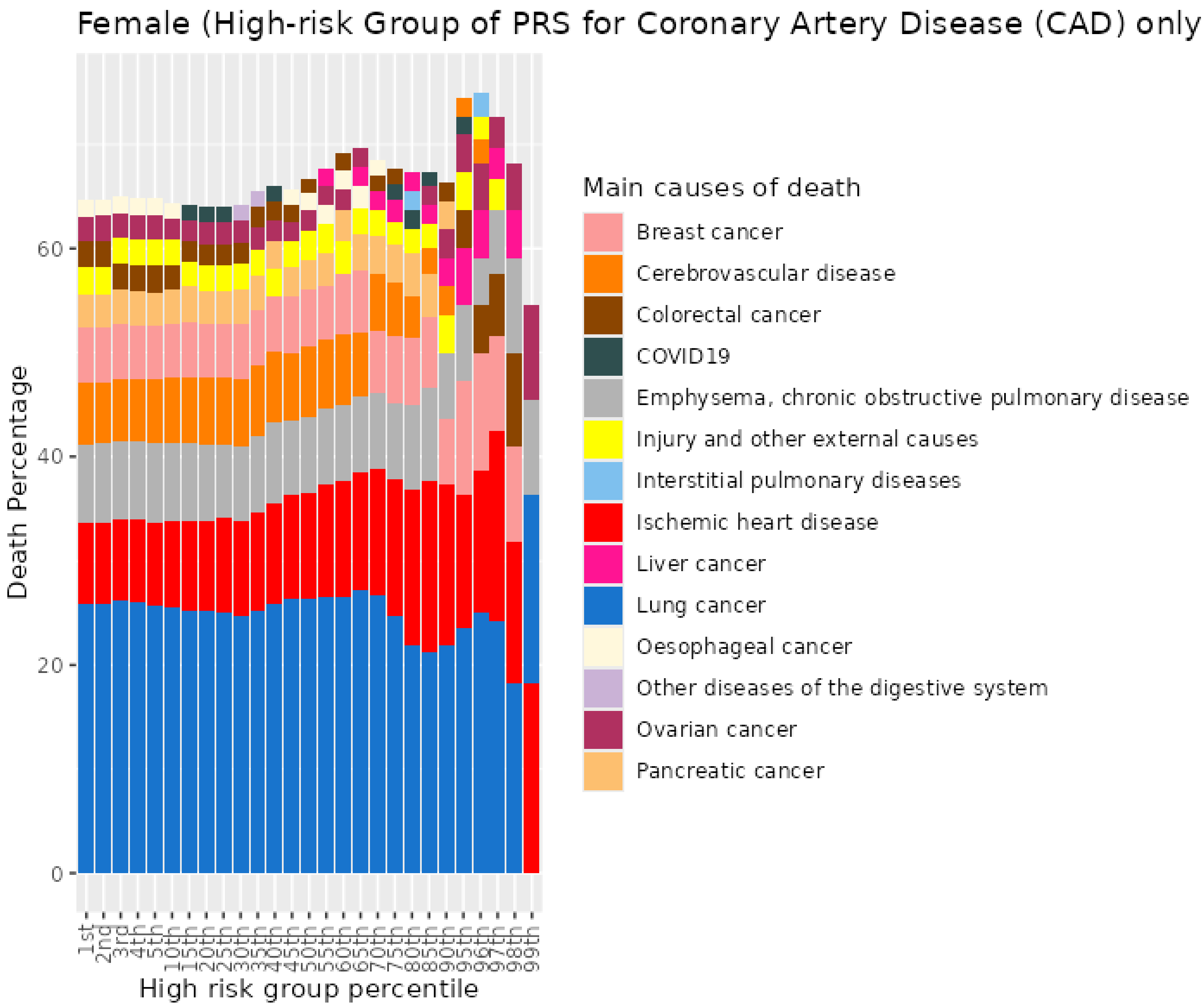


*Figure S35. Percentiles from 1 to 99 for coronary artery disease PRS among female current smokers who died at age 60-69*

##### 4.9 Female current smokers who died at age 70+

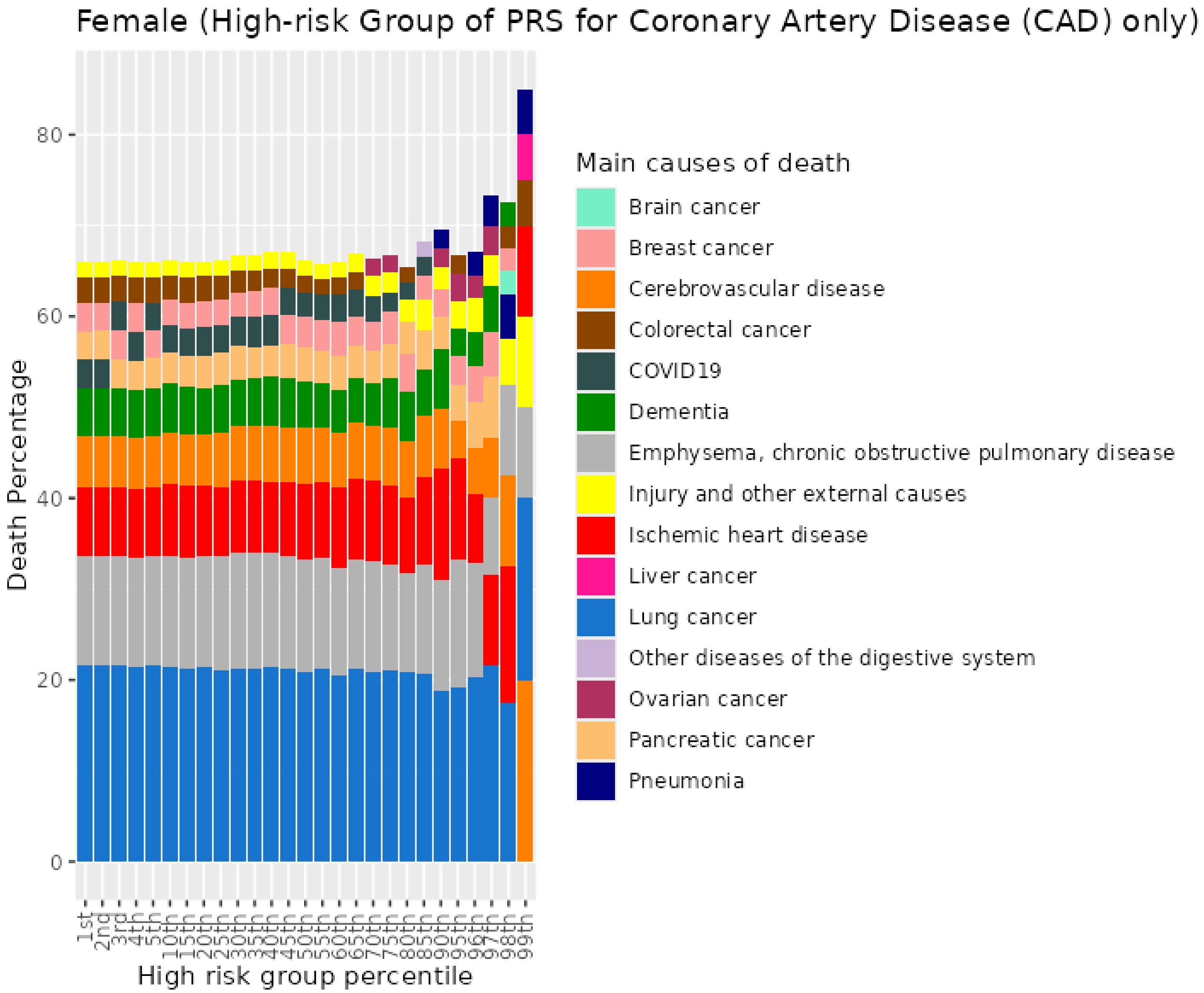


*Figure S36. Percentiles from 1 to 99 for coronary artery disease among female current smokers who died at age 70+*

### 5 Ischaemic stroke (ISS) PRS for UKB females

#### 5.1 Female never smokers who died at age <=59

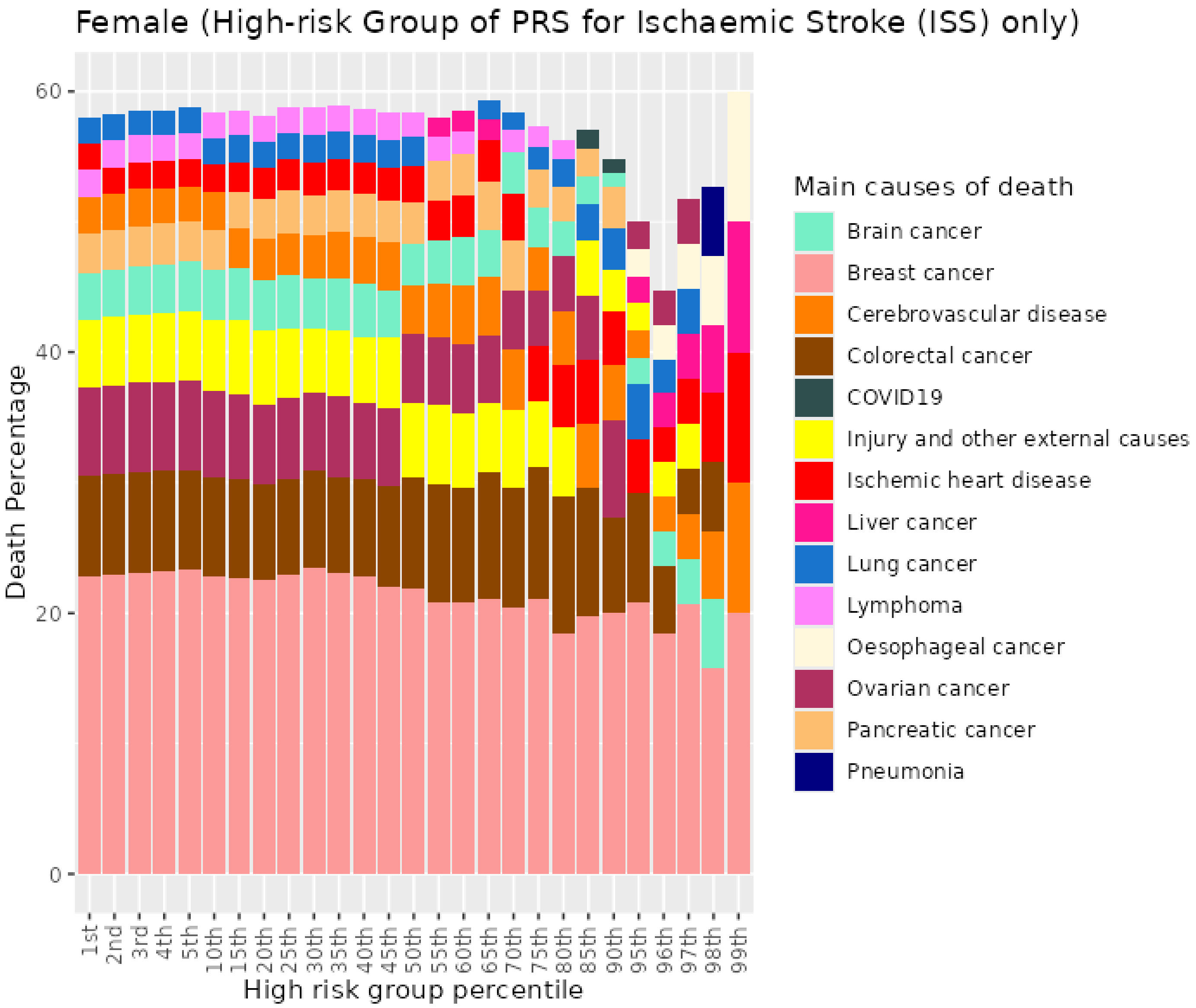


*Figure S37. Percentiles from 1 to 99 for ischaemic stroke PRS among female previous never who died at age <=59*

#### 5.2 Female never smokers who died at age 60-69

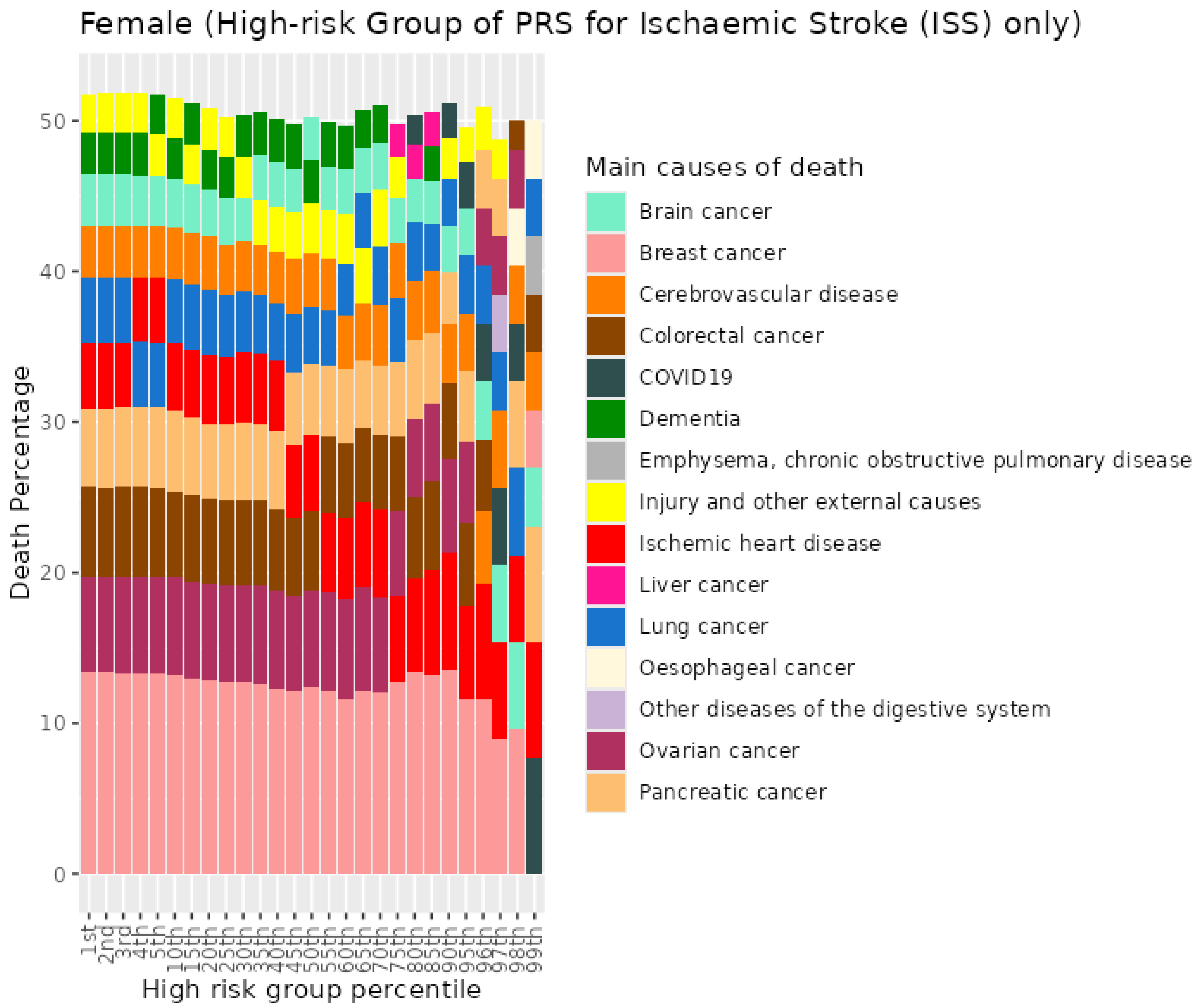


*Figure S38. Percentiles from 1 to 99 for ischaemic stroke PRS among female previous never who died at age 60-69*

#### 5.3 Female never smokers who died at age 70+

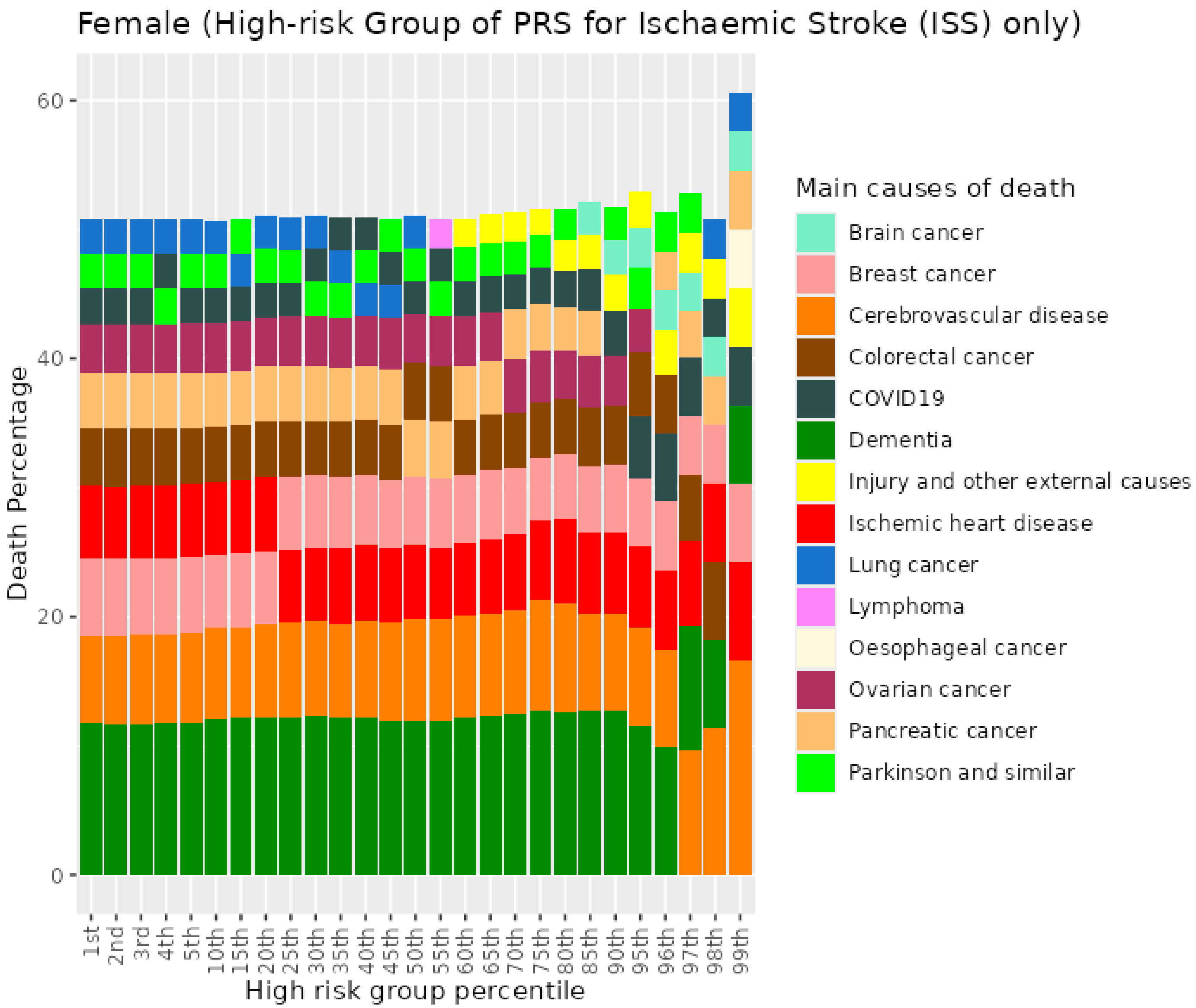


Figure S39. Percentiles from 1 to 99 for ischaemic stroke PRS among female previous never who died at age 70+

#### 5.4 Female previous smokers who died at age <=59

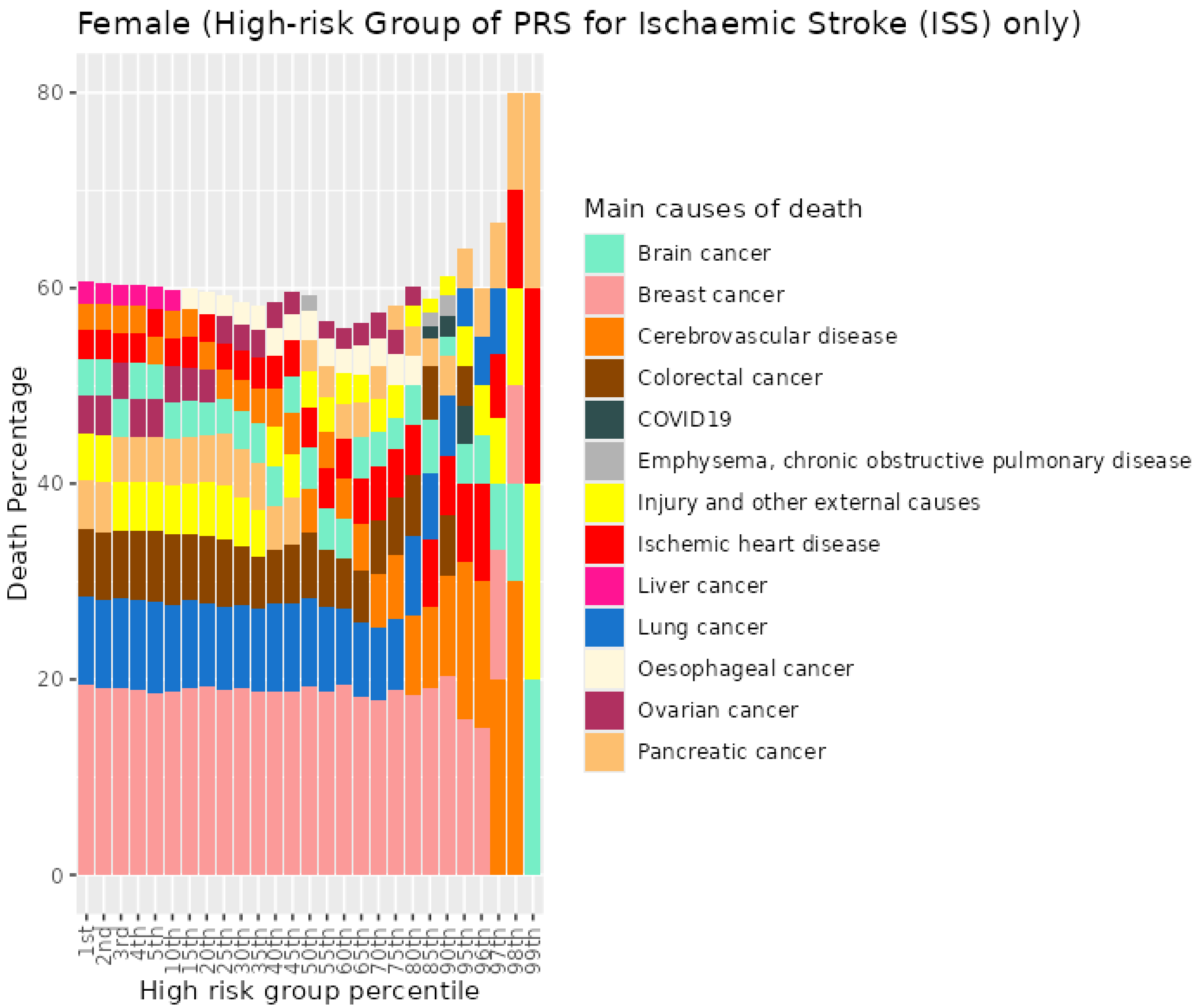


*Figure S40. Percentiles from 1 to 99 for ischaemic stroke PRS among female previous smokers who died at age <=59*

#### 5.5 Female previous smokers who died at age 60-69

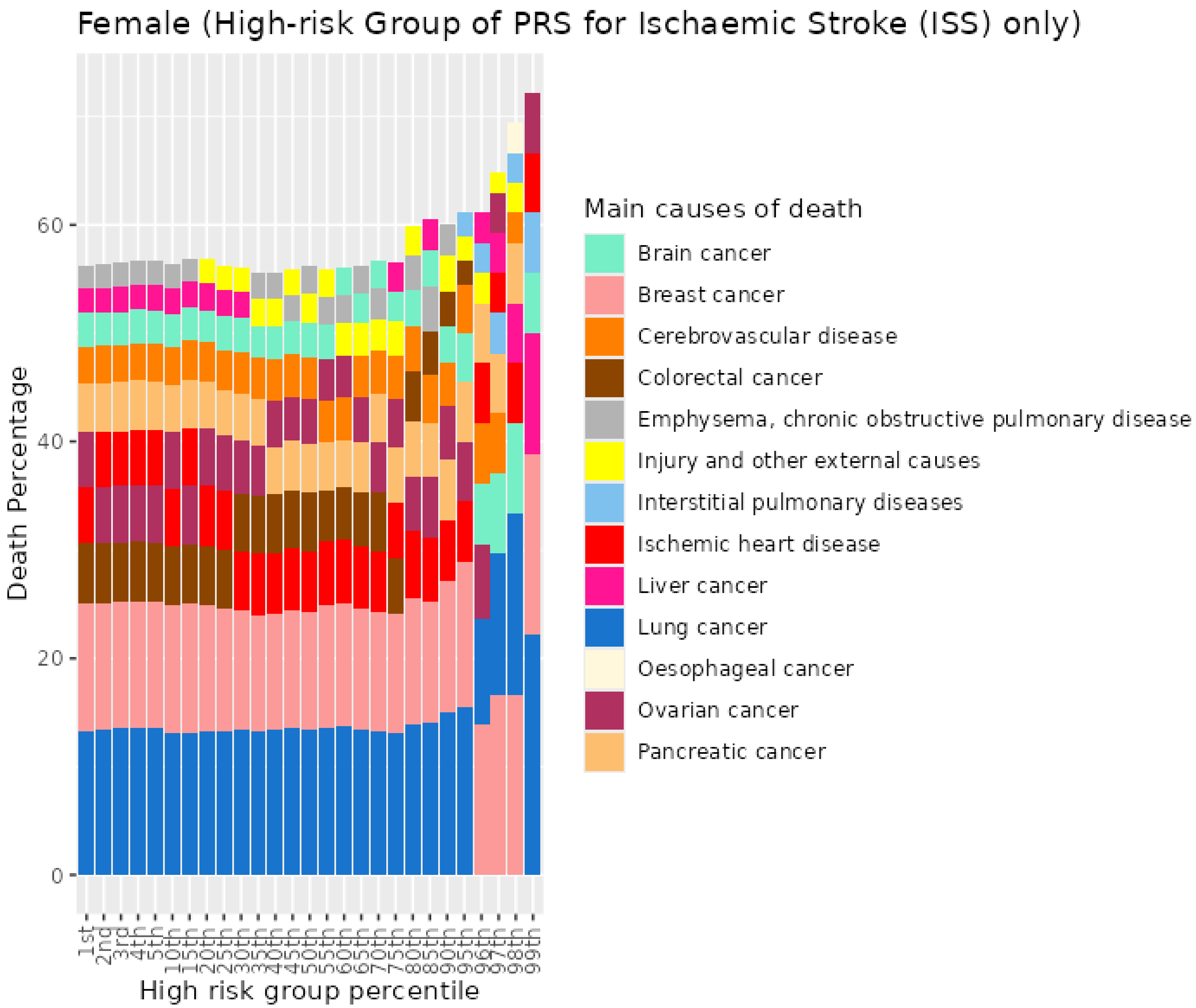


*Figure S41. Percentiles from 1 to 99 for ischaemic stroke PRS among female previous smokers who died at age 60-69*

#### 5.6 Female previous smokers who died at age 70+

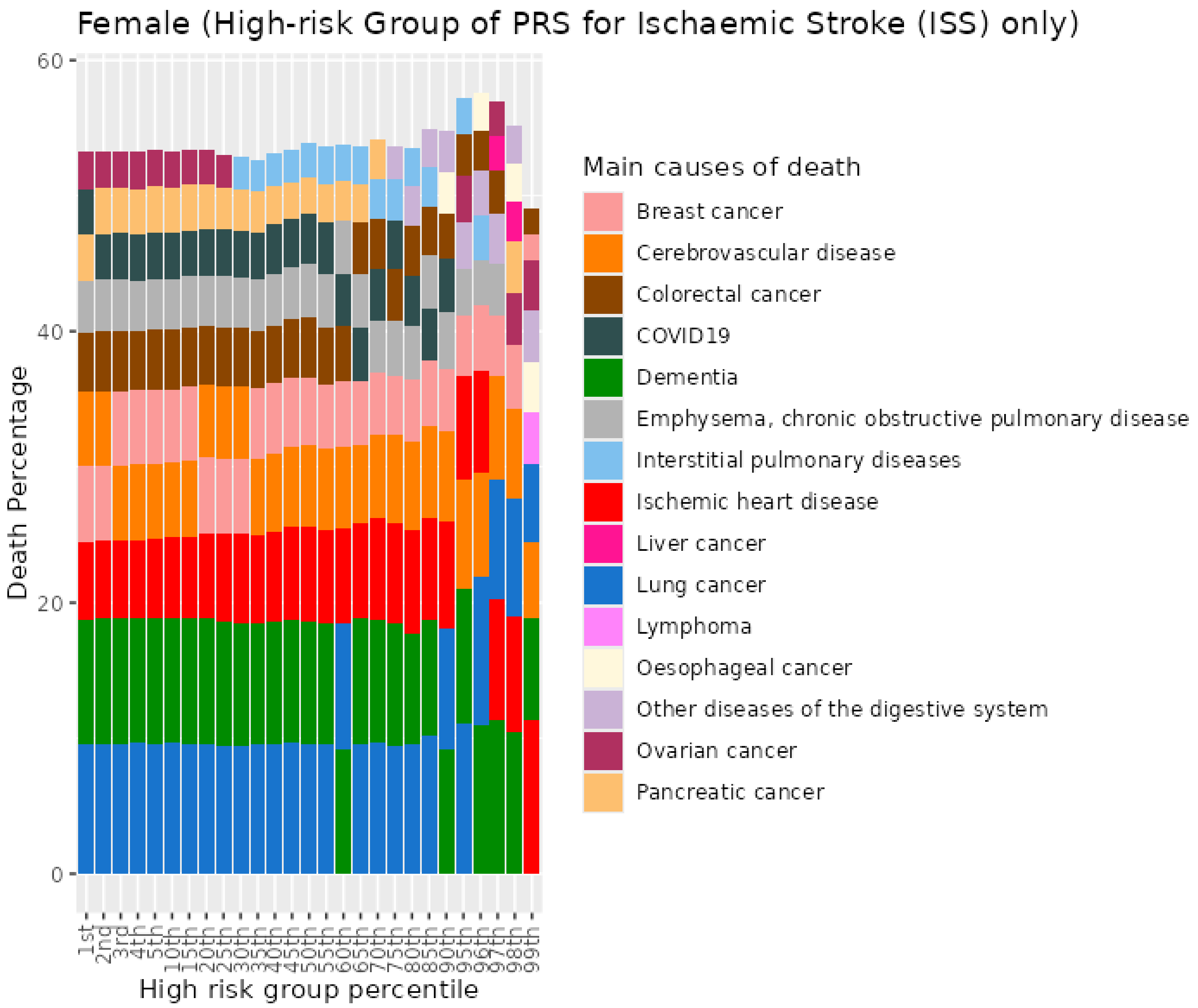


Figure S42. Percentiles from 1 to 99 for ischaemic stroke PRS among female previous smokers who died at age 70+

#### 5.7 Female current smokers who died at age <=59

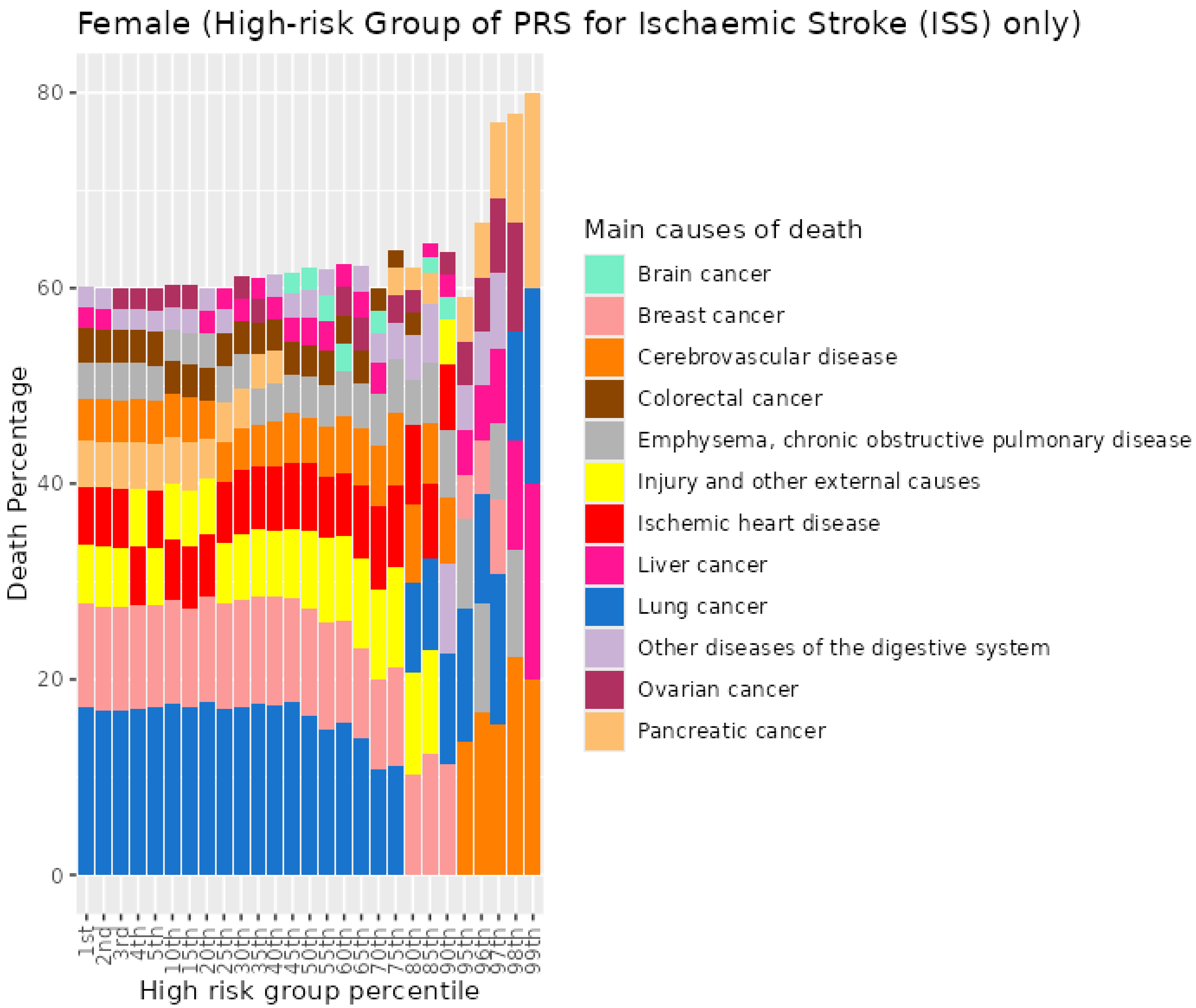


*Figure S43. Percentiles from 1 to 99 for ischaemic stroke PRS among female current smokers who died at age <=59*

##### 5.8 Female current smokers who died at age 60-69

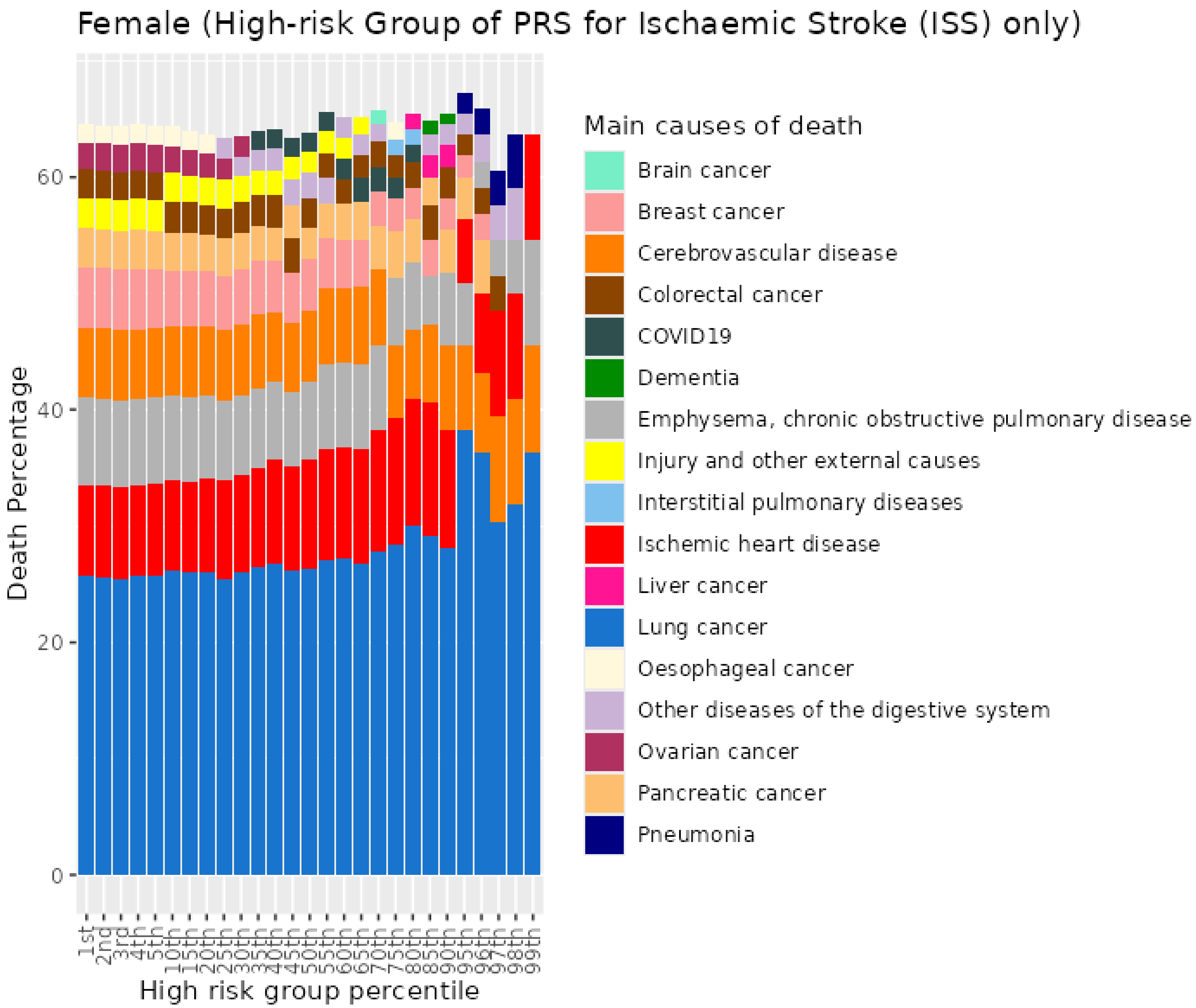


*Figure S44. Percentiles from 1 to 99 for ischaemic stroke PRS among female current smokers who died at age 60-69*

##### 5.9 Female current smokers who died at age 70+

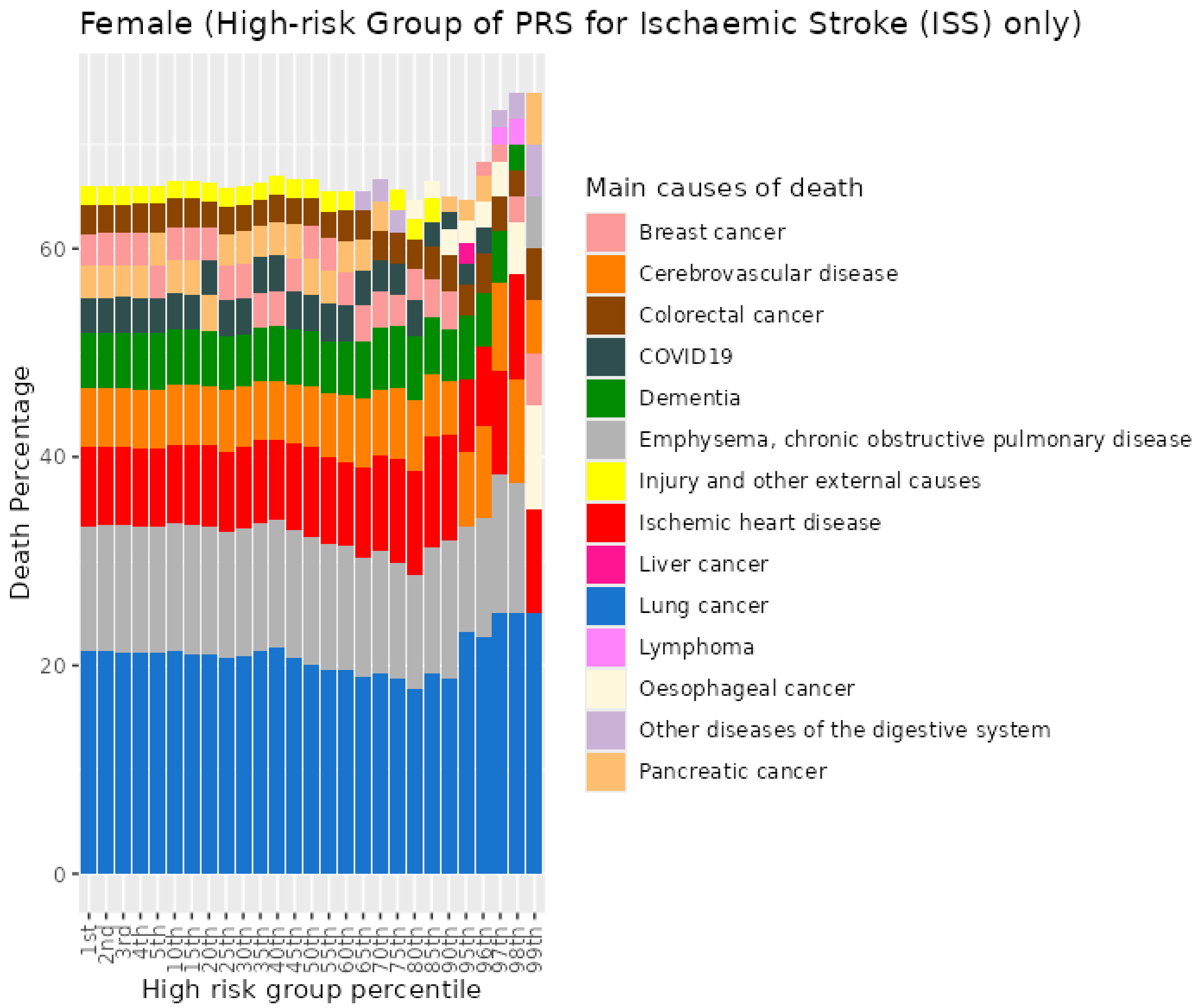


Figure S45. Percentiles from 1 to 99 for ischaemic stroke PRS among female current smokers who died at age 70+

### 6 Breast cancer (BC) PRS for UKB females

#### 6.1 Female never smokers who died at age <=59

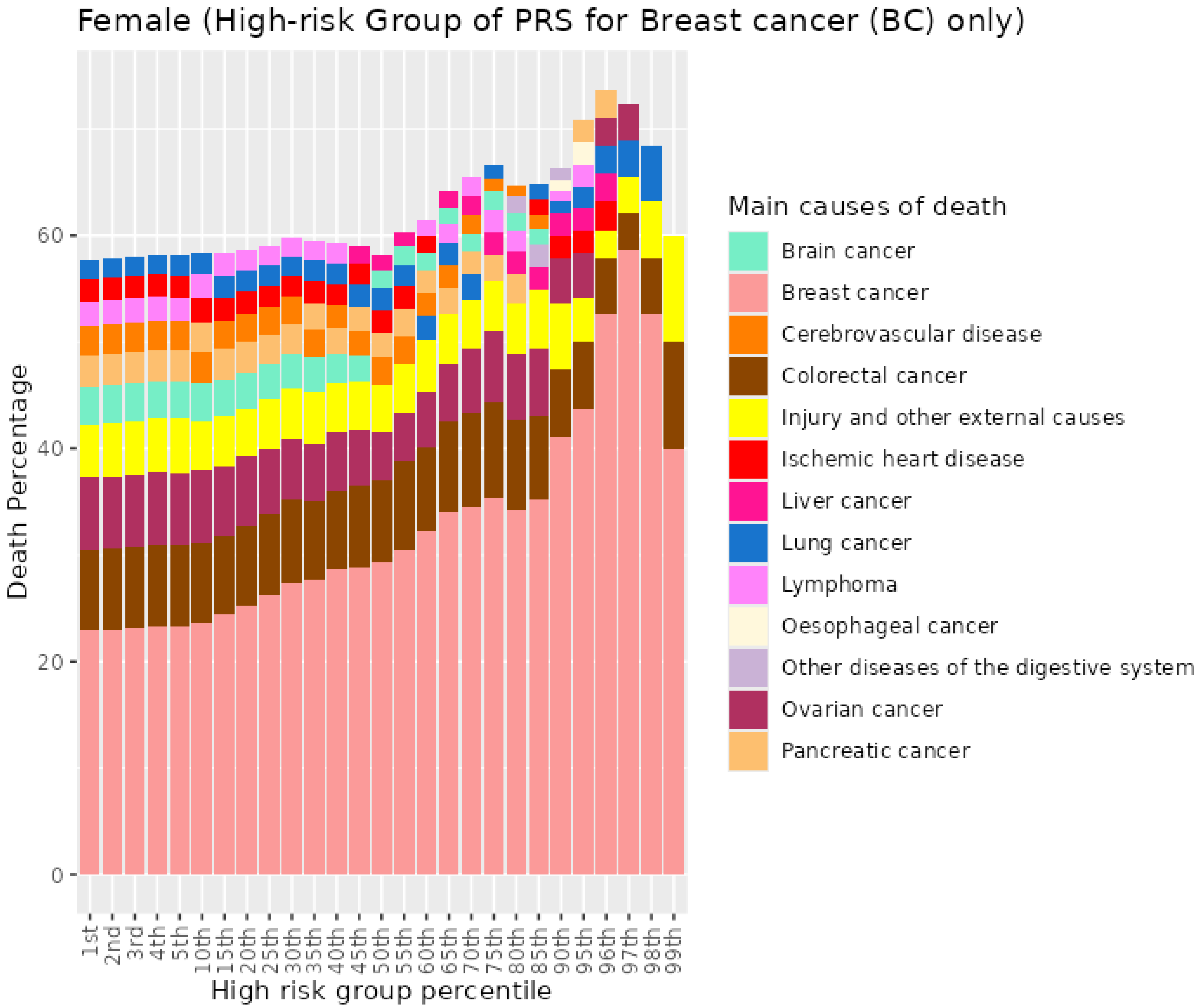


Figure S46. Percentiles from 1 to 99 for breast cancer PRS among female previous never who died at age <=59

##### 6.2 Female never smokers who died at age 60-69

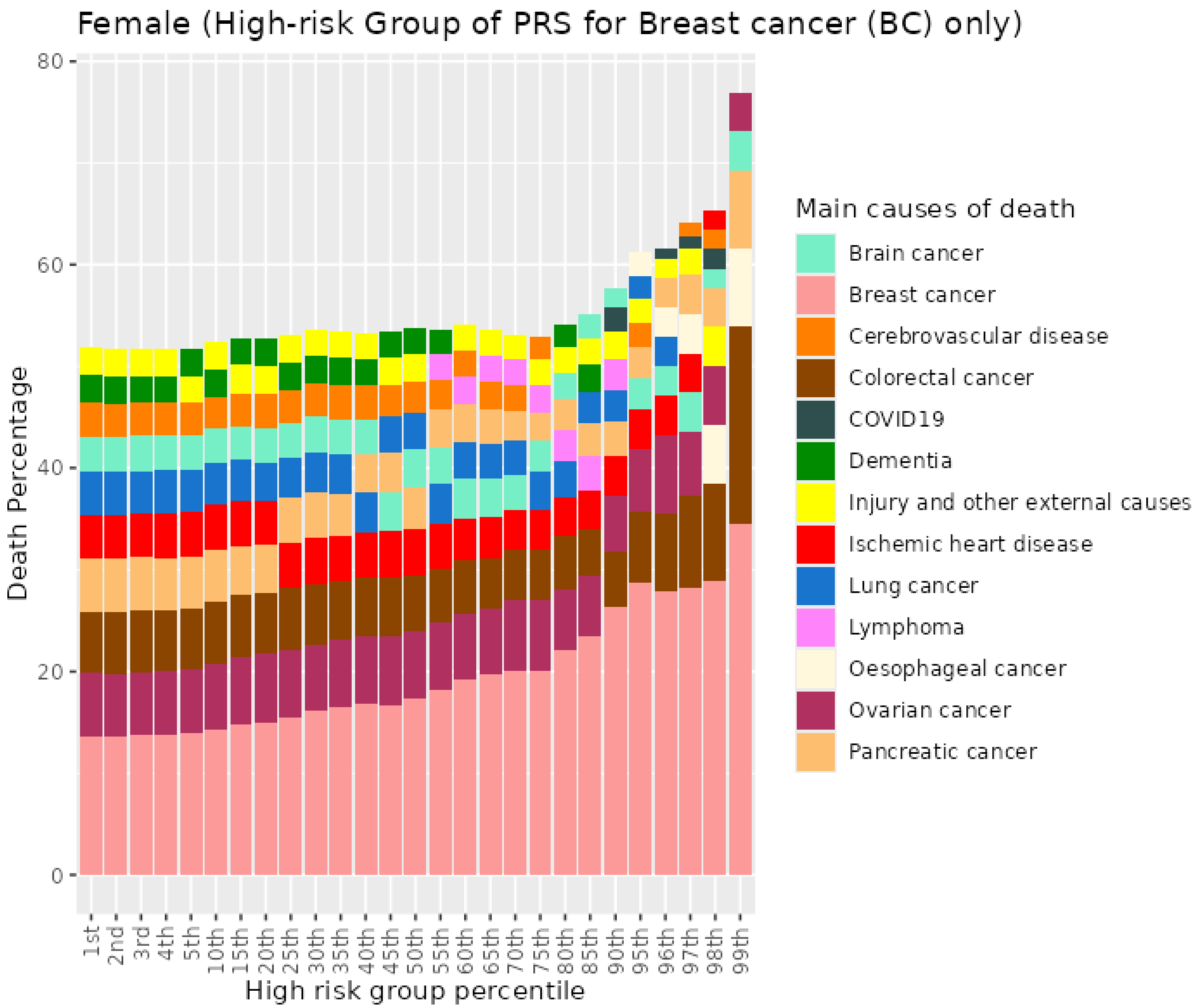


Figure S47. Percentiles from 1 to 99 for breast cancer PRS among female previous never who died at age 60-69

##### 6.3 Female never smokers who died at age 70+

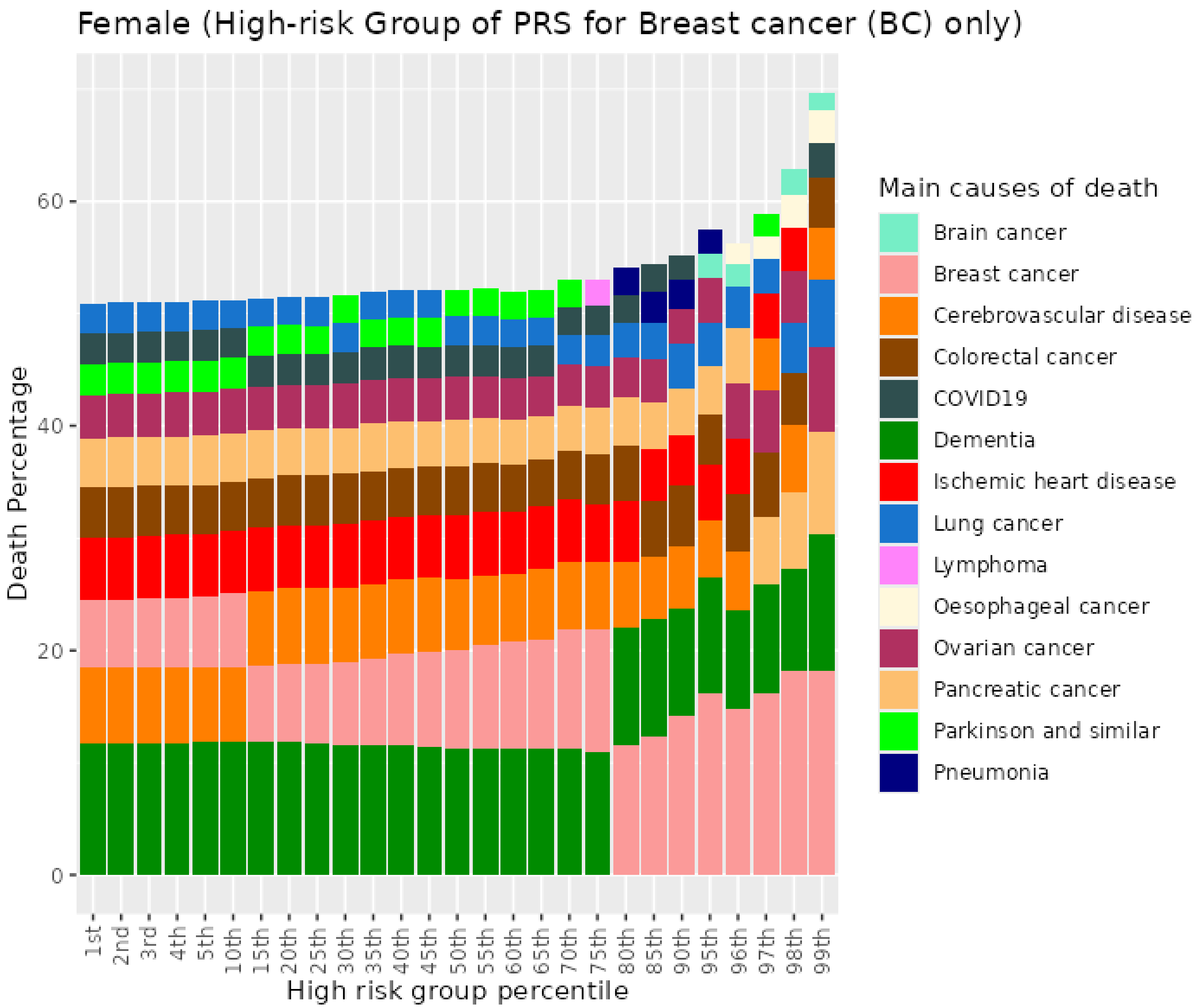


Figure S48. Percentiles from 1 to 99 for breast cancer PRS among female previous never who died at age 70+

#### 6.4 Female previous smokers who died at age <=59

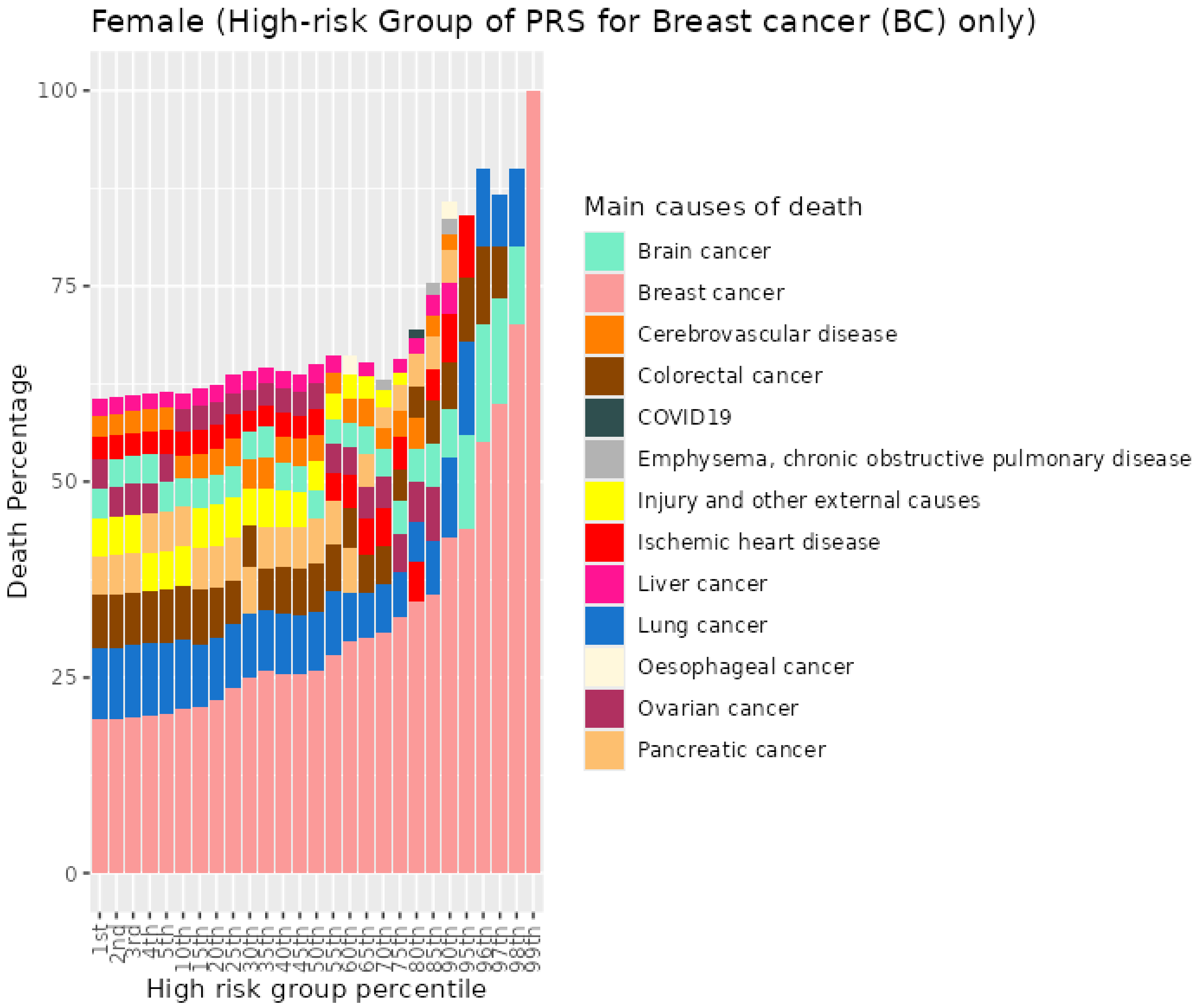


Figure S49. Percentiles from 1 to 99 for breast cancer PRS among female previous smokers who died at age <=59

##### 6.5 Female previous smokers who died at age 60-69

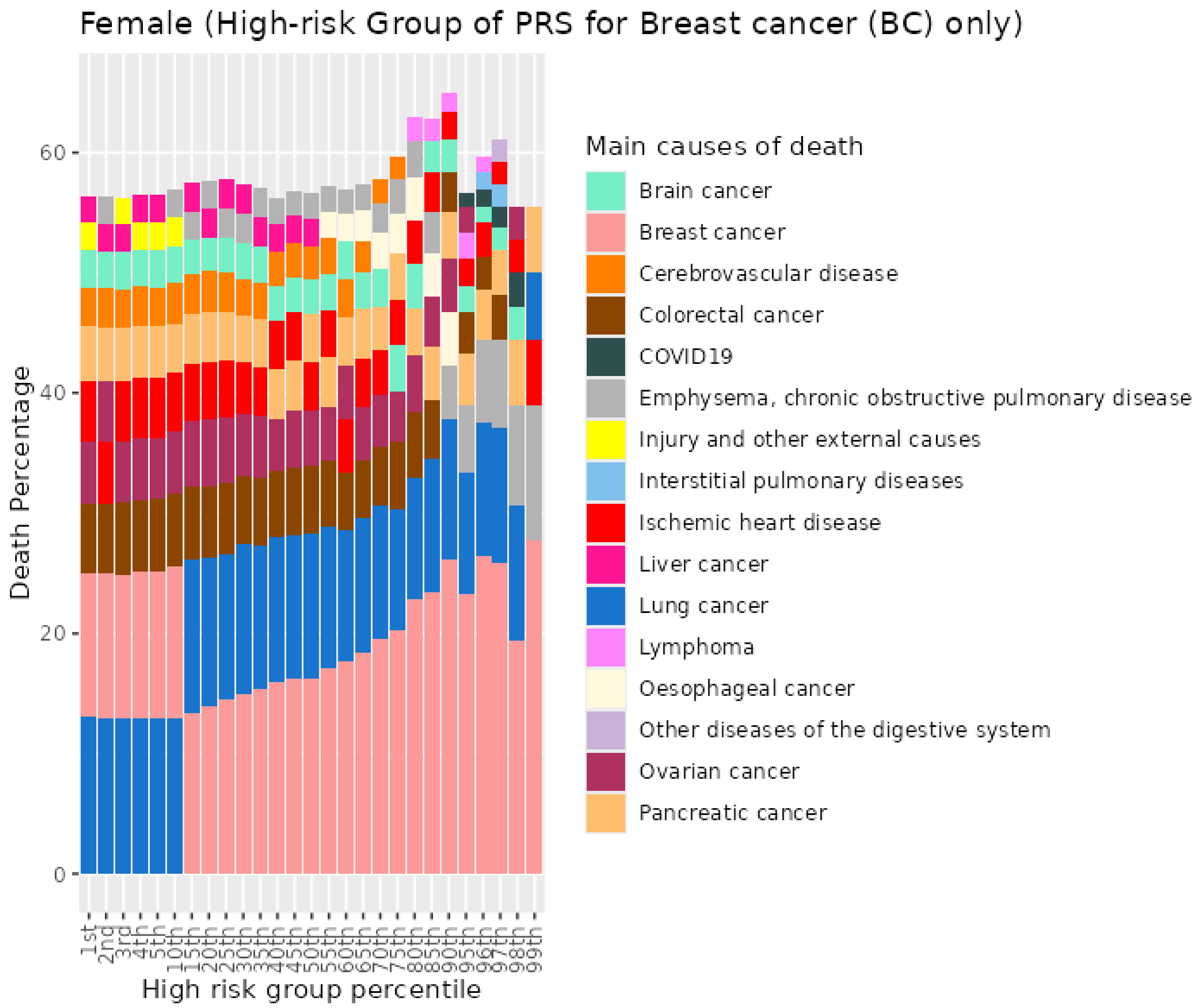


Figure S50. Percentiles from 1 to 99 for breast cancer PRS among female previous smokers who died at age 60-69

##### 6.6 Female previous smokers who died at age 70+

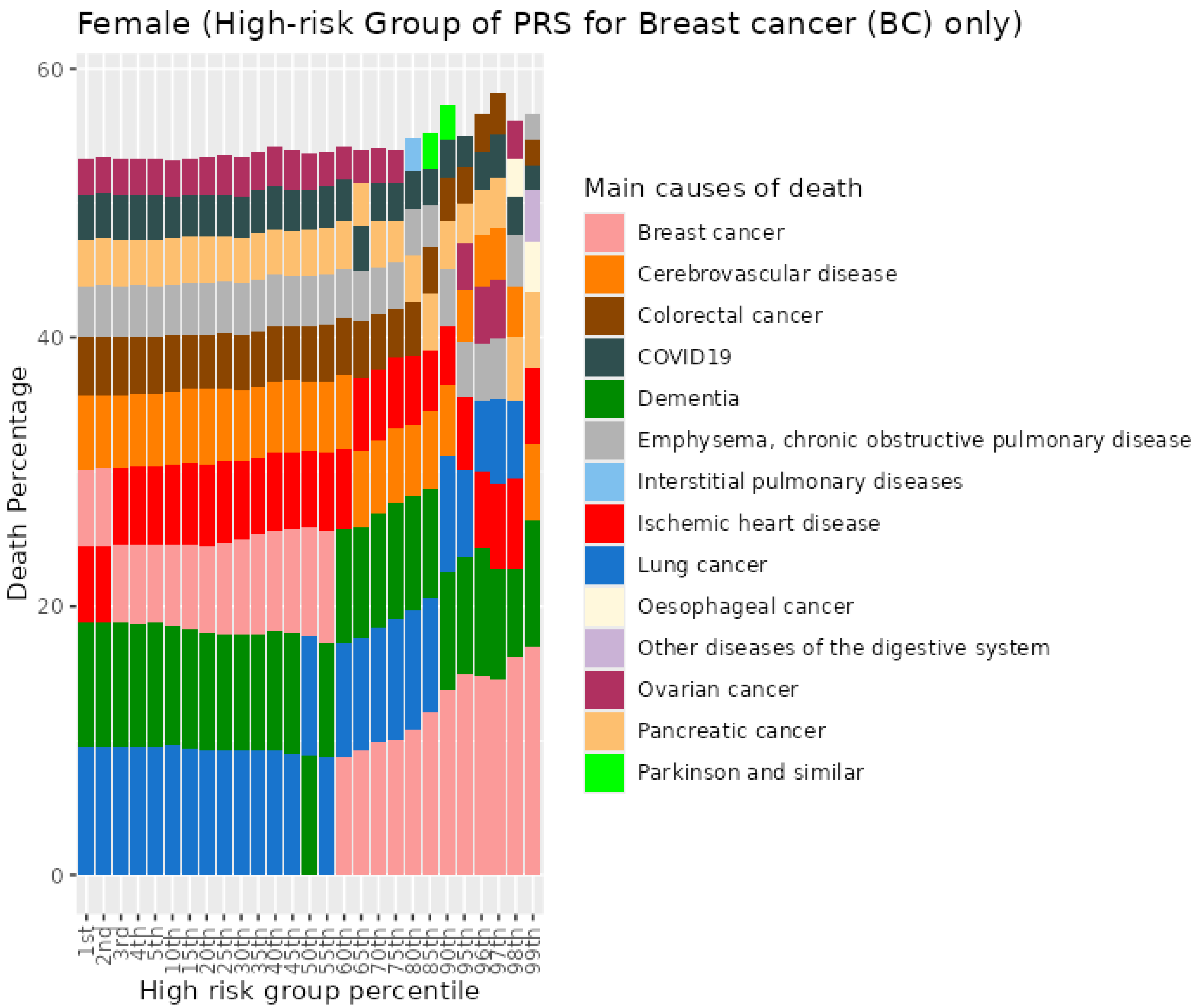


Figure S51. Percentiles from 1 to 99 for breast cancer PRS among female previous smokers who died at age 70+

##### 6.7 Female current smokers who died at age <=59

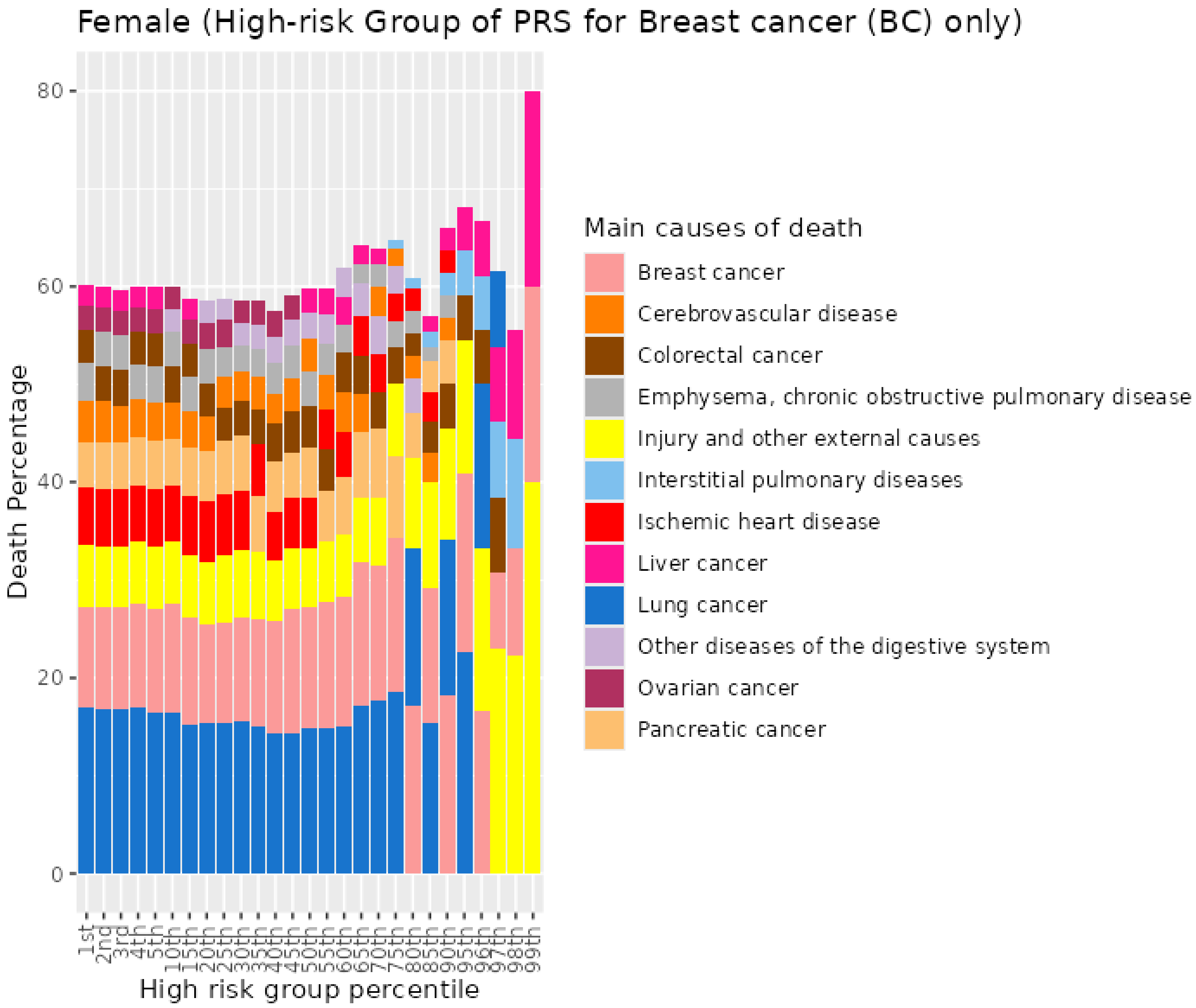


Figure S52. Percentiles from 1 to 99 for breast cancer PRS among female current smokers who died at age <=59

##### 6.8 Female current smokers who died at age 60-69

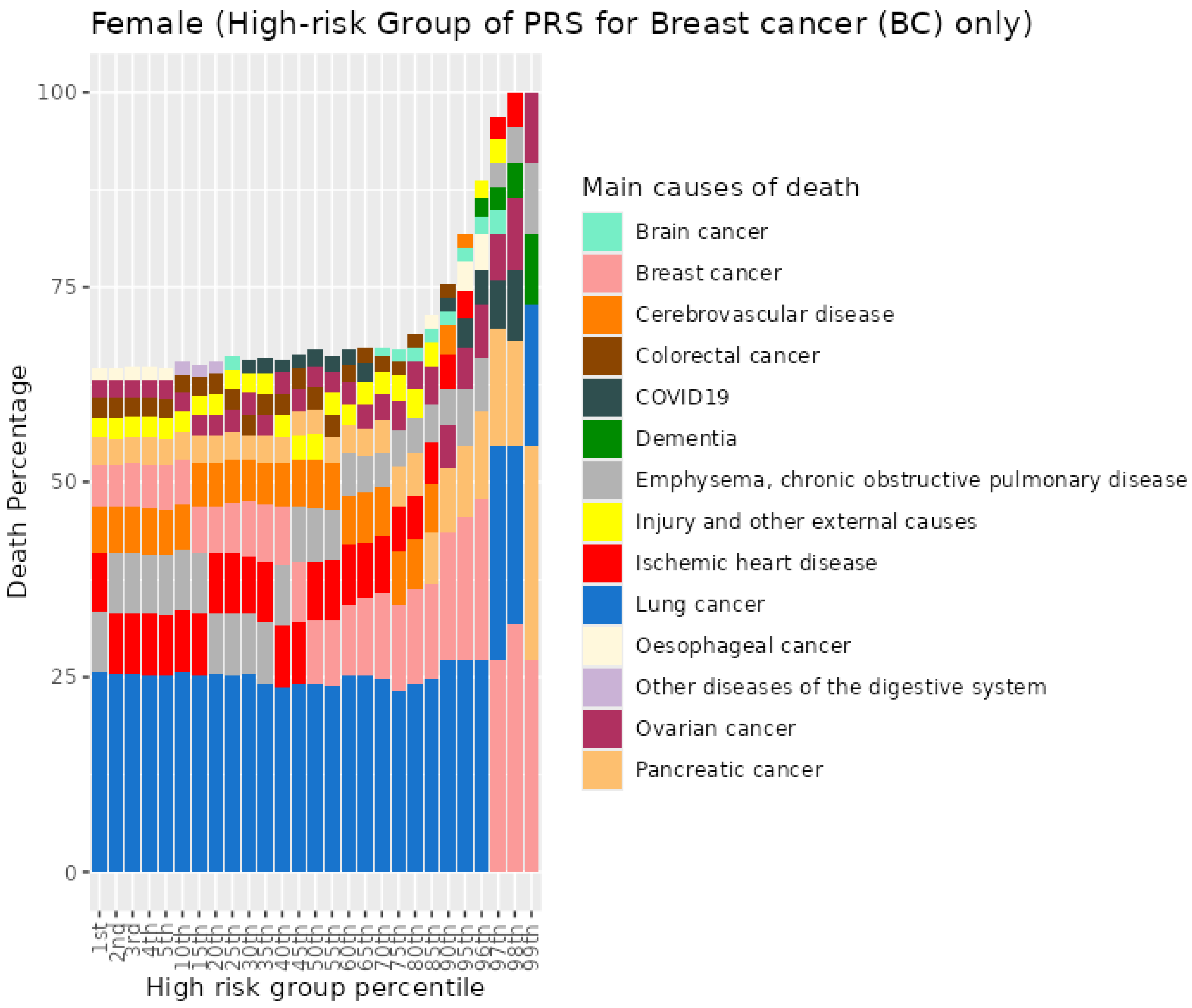


Figure S53. Percentiles from 1 to 99 for breast cancer PRS among female current smokers who died at age 60-69

#### 6.9 Female current smokers who died at age 70+

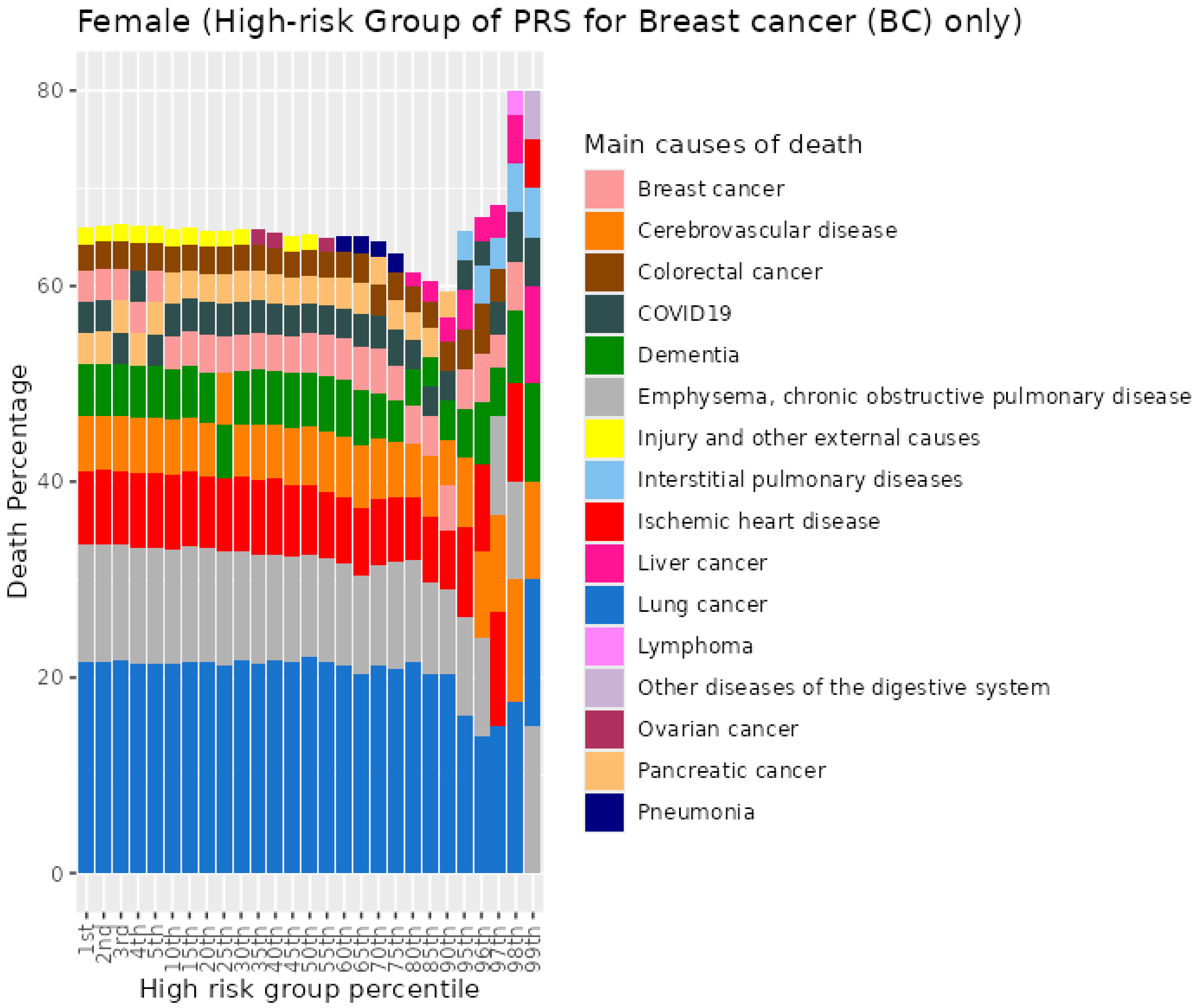


Figure S54. Percentiles from 1 to 99 for breast cancer PRS among female current smokers who died at age 70+

### 7 Ovarian cancer (EOC) PRS for UKB females

#### 7.1 Female never smokers who died at age <=59

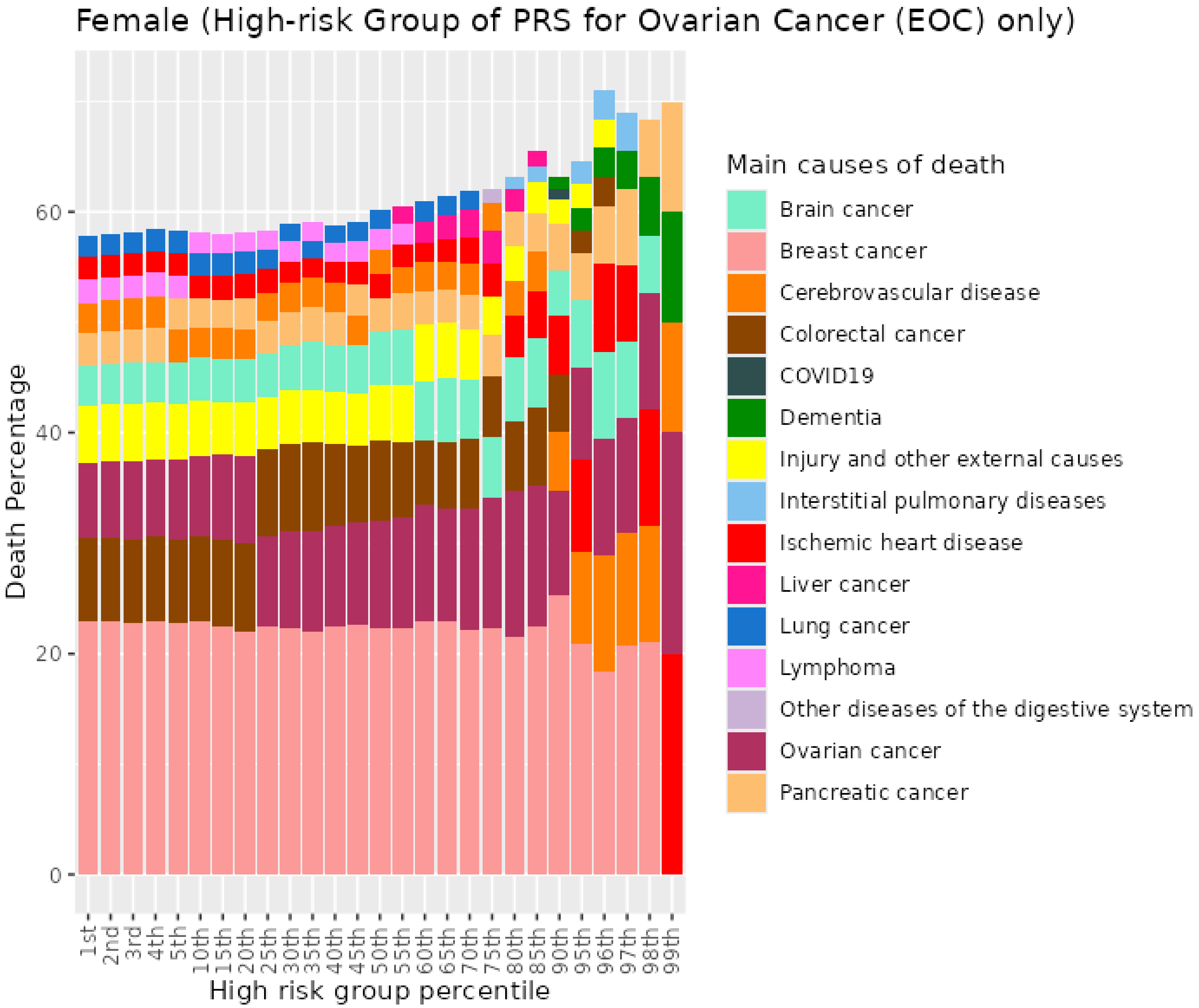


Figure S55. Percentiles from 1 to 99 for epithelial ovarian cancer PRS among female never smokers who died at age <=59

#### 7.2 Female never smokers who died at age 60-69

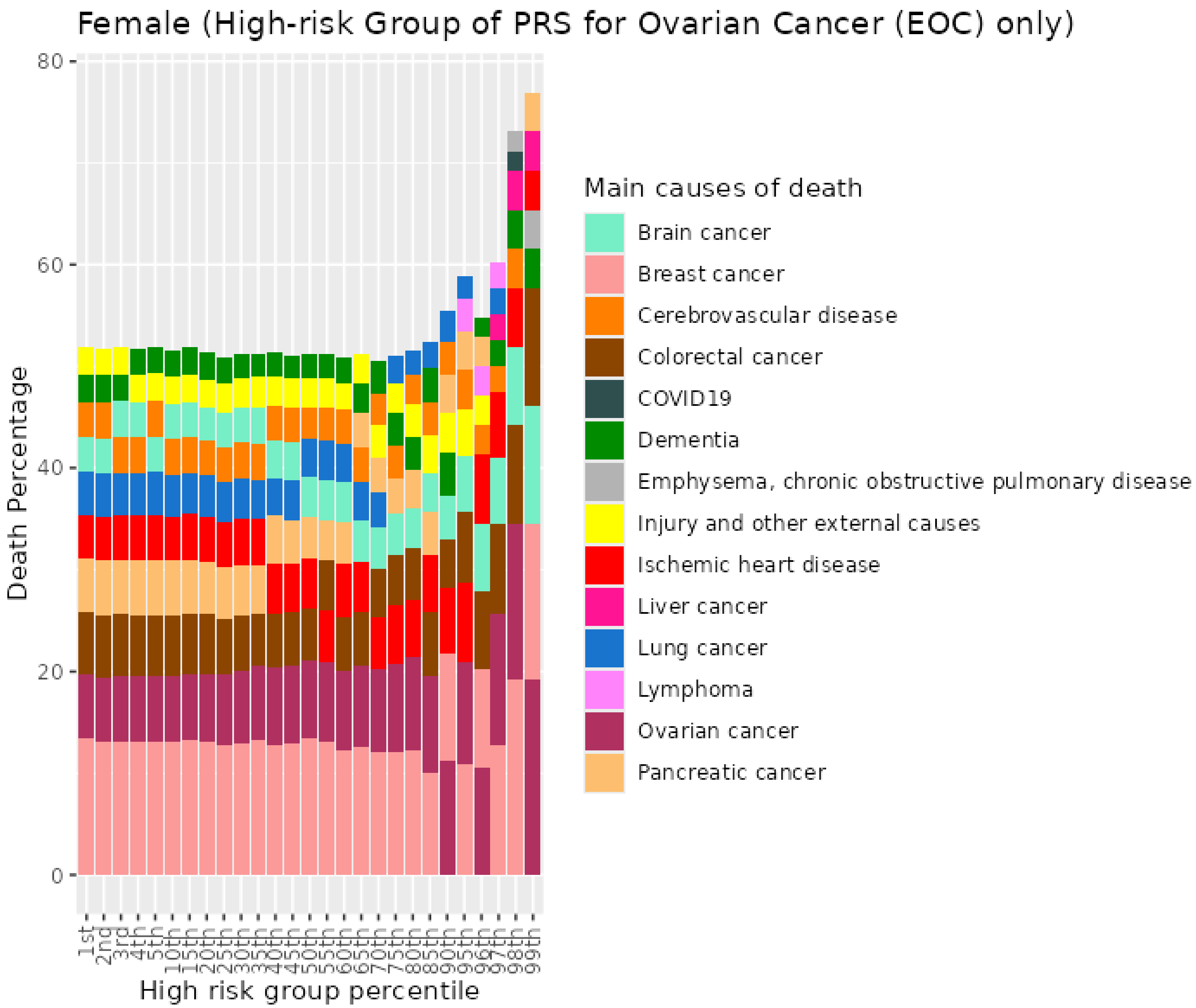


Figure S56. Percentiles from 1 to 99 for epithelial ovarian cancer PRS among female never smokers who died at age 60-69

##### 7.3 Female never smokers who died at age 70+

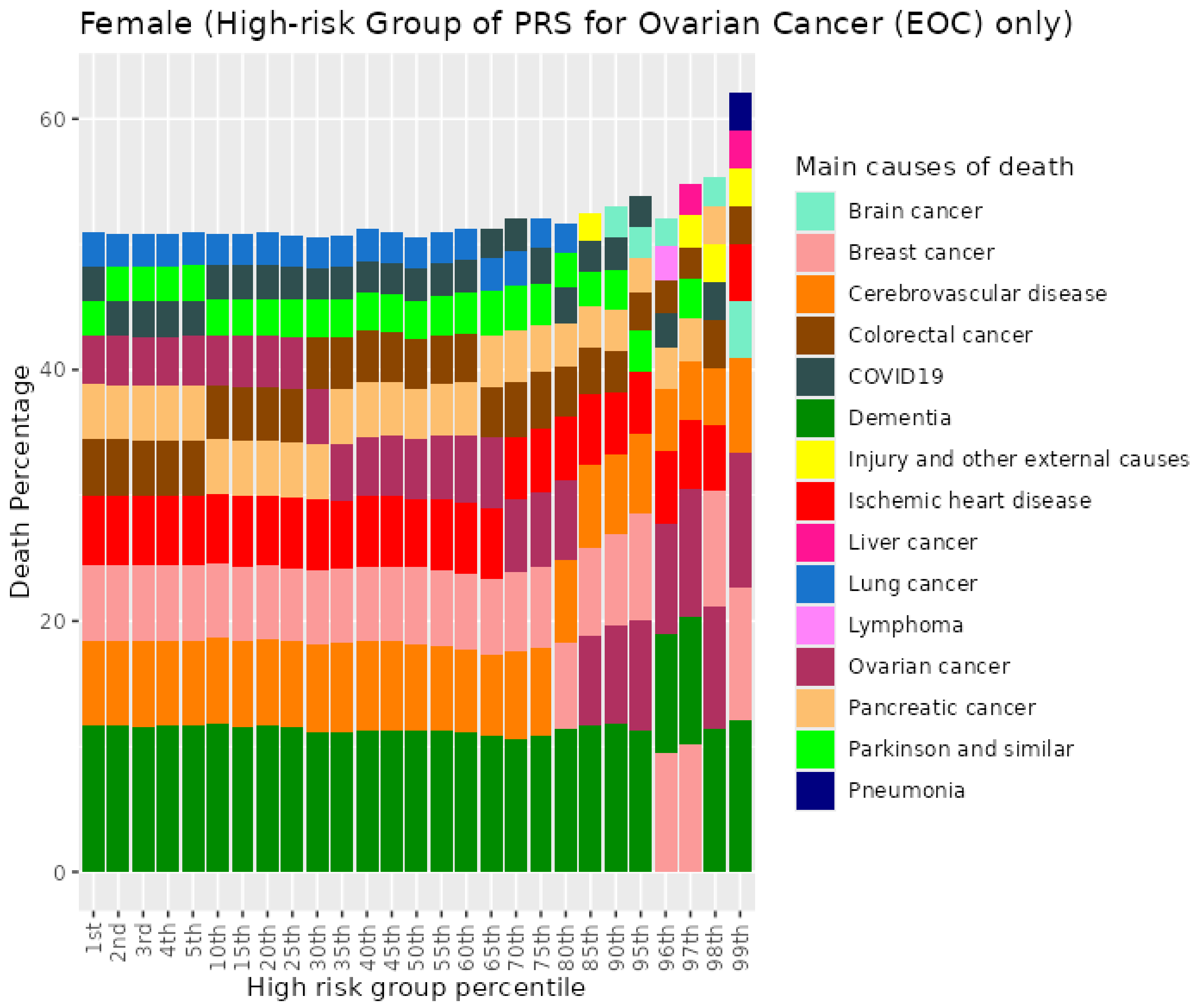


Figure S57. Percentiles from 1 to 99 for epithelial ovarian cancer PRS among female never smokers who died at age 70+

#### 7.4 Female previous smokers who died at age <=59

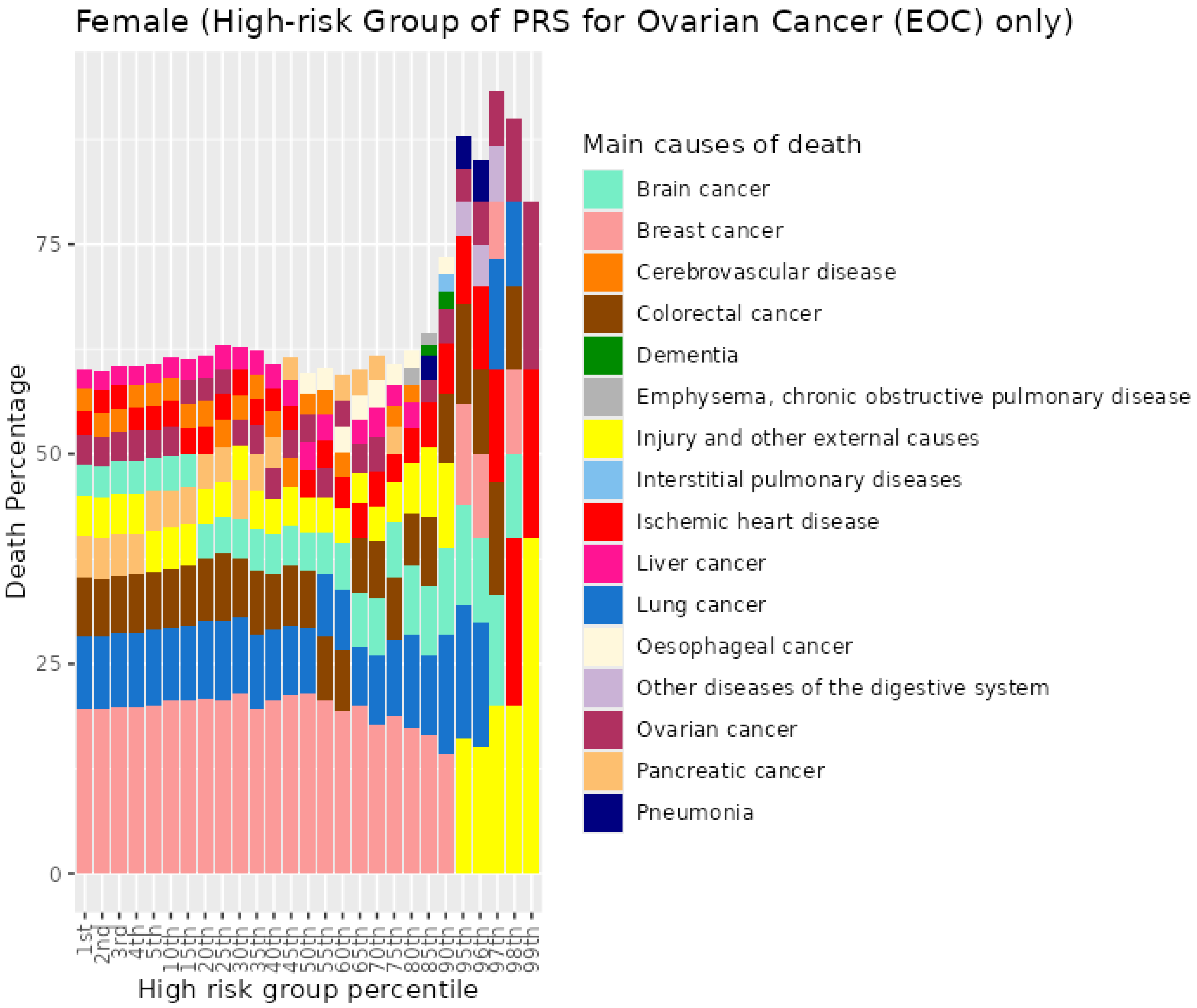


Figure S58. Percentiles from 1 to 99 for epithelial ovarian cancer among female previous smokers who died at age <=59

##### 7.5 Female previous smokers who died at age 60-69

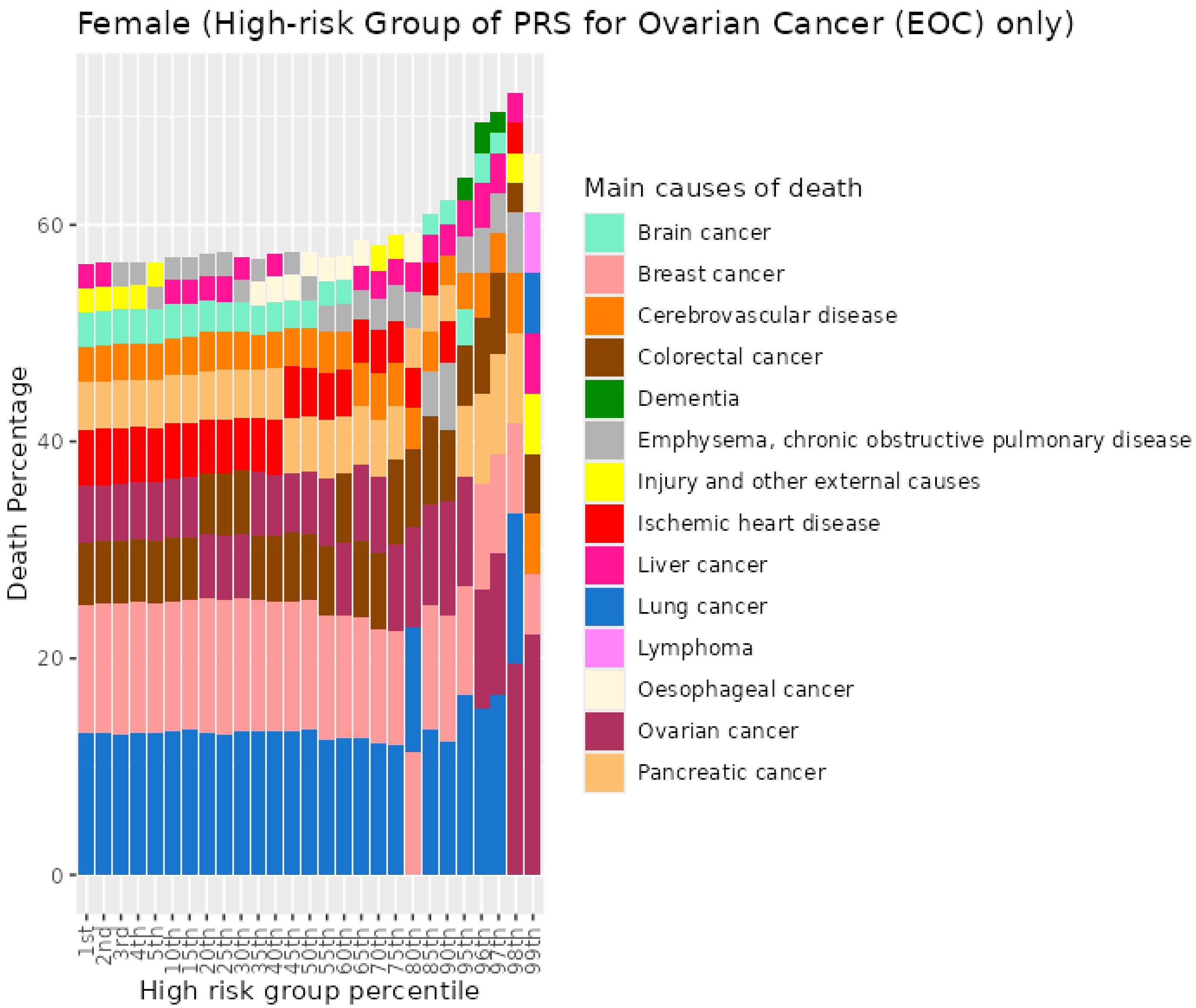


Figure S59. Percentiles from 1 to 99 for epithelial ovarian cancer PRS among female previous smokers who died at age 60-69

##### 7.6 Female previous smokers who died at age 70+

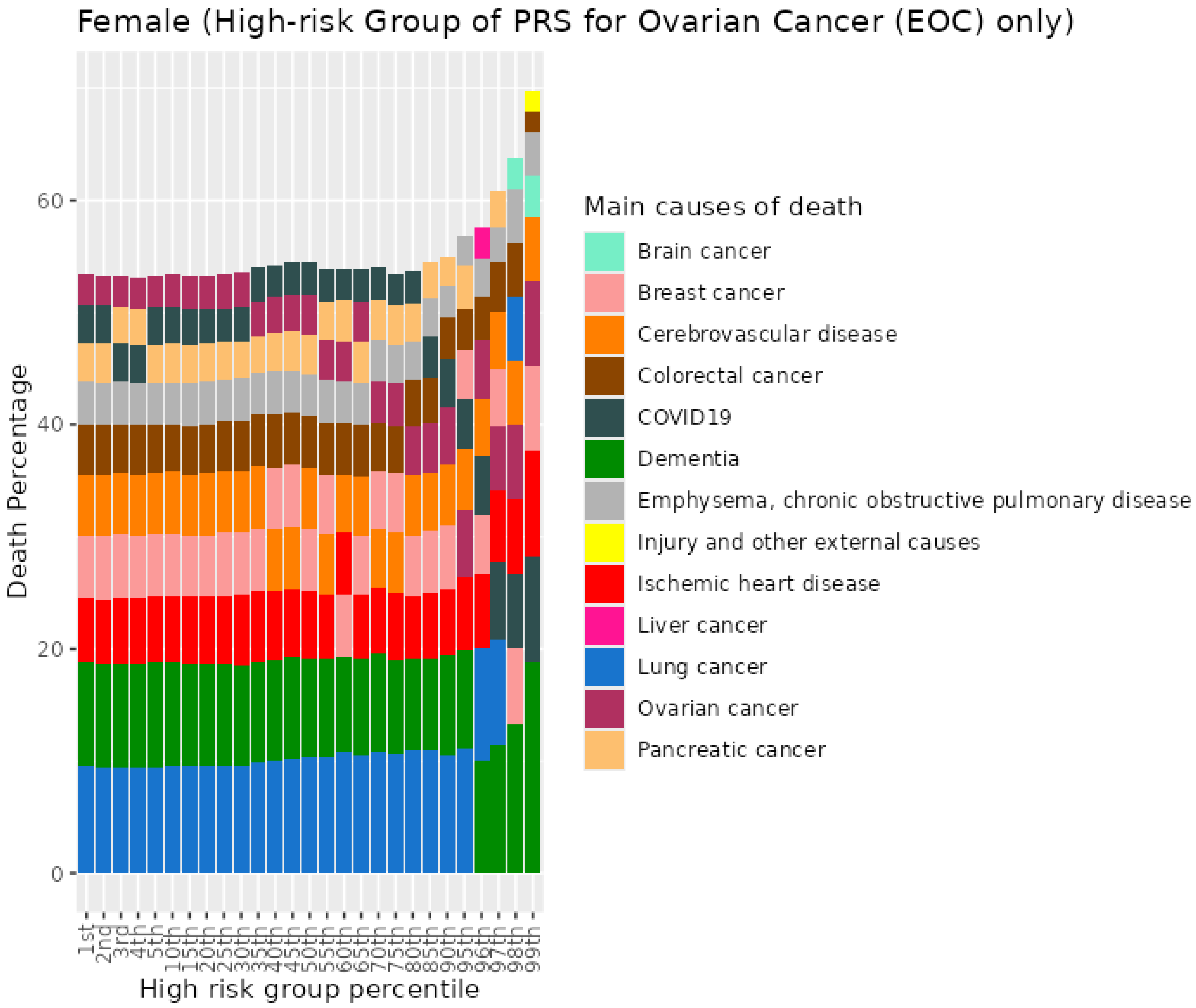


Figure S60. Percentiles from 1 to 99 for epithelial ovarian cancer PRS among female previous smokers who died at age 70+

#### 7.7 Female current smokers who died at age <=59

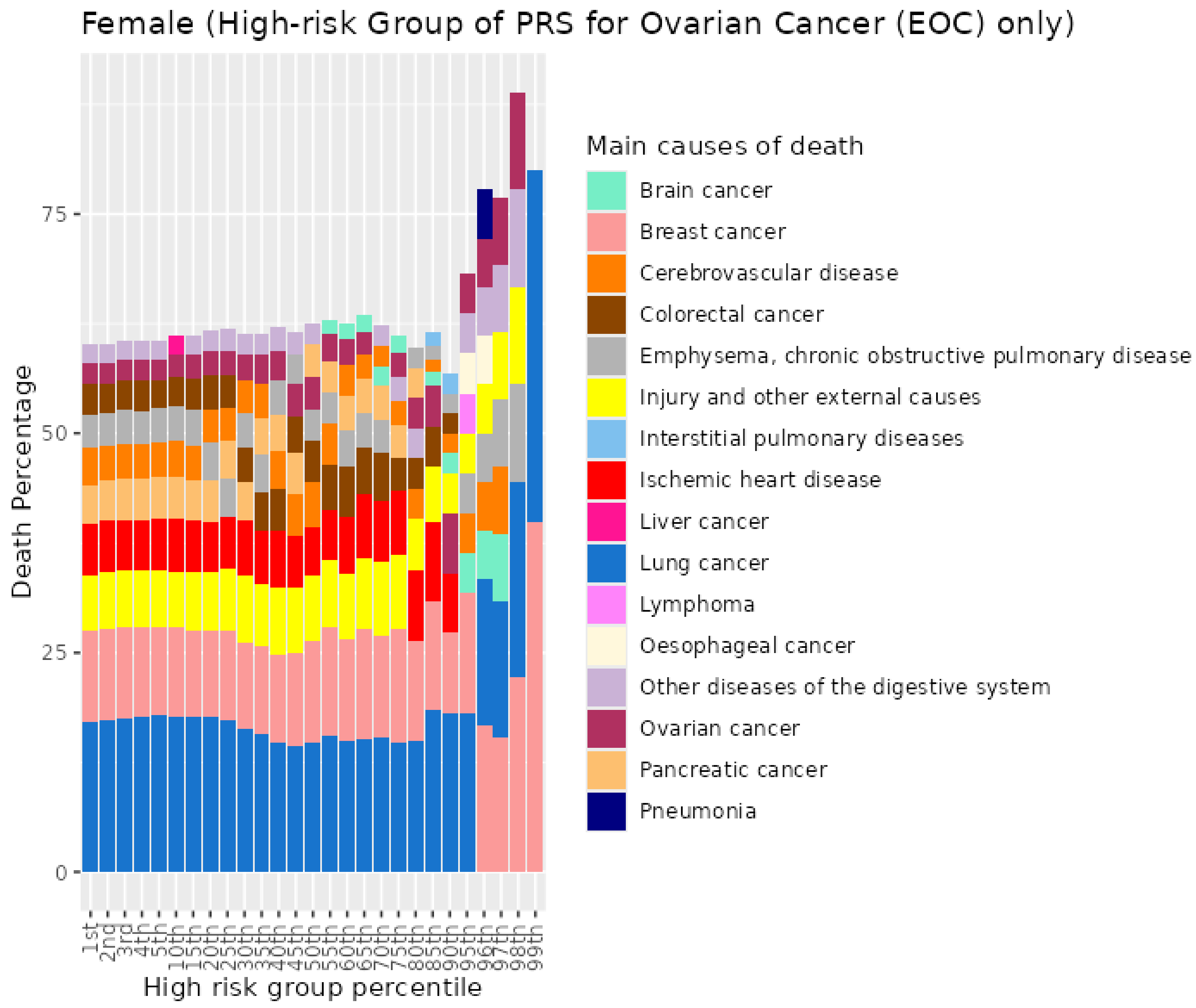


Figure S61. Percentiles from 1 to 99 for epithelial ovarian cancer PRS among female current smokers who died at age <=59

#### 7.8 Female current smokers who died at age 60-69

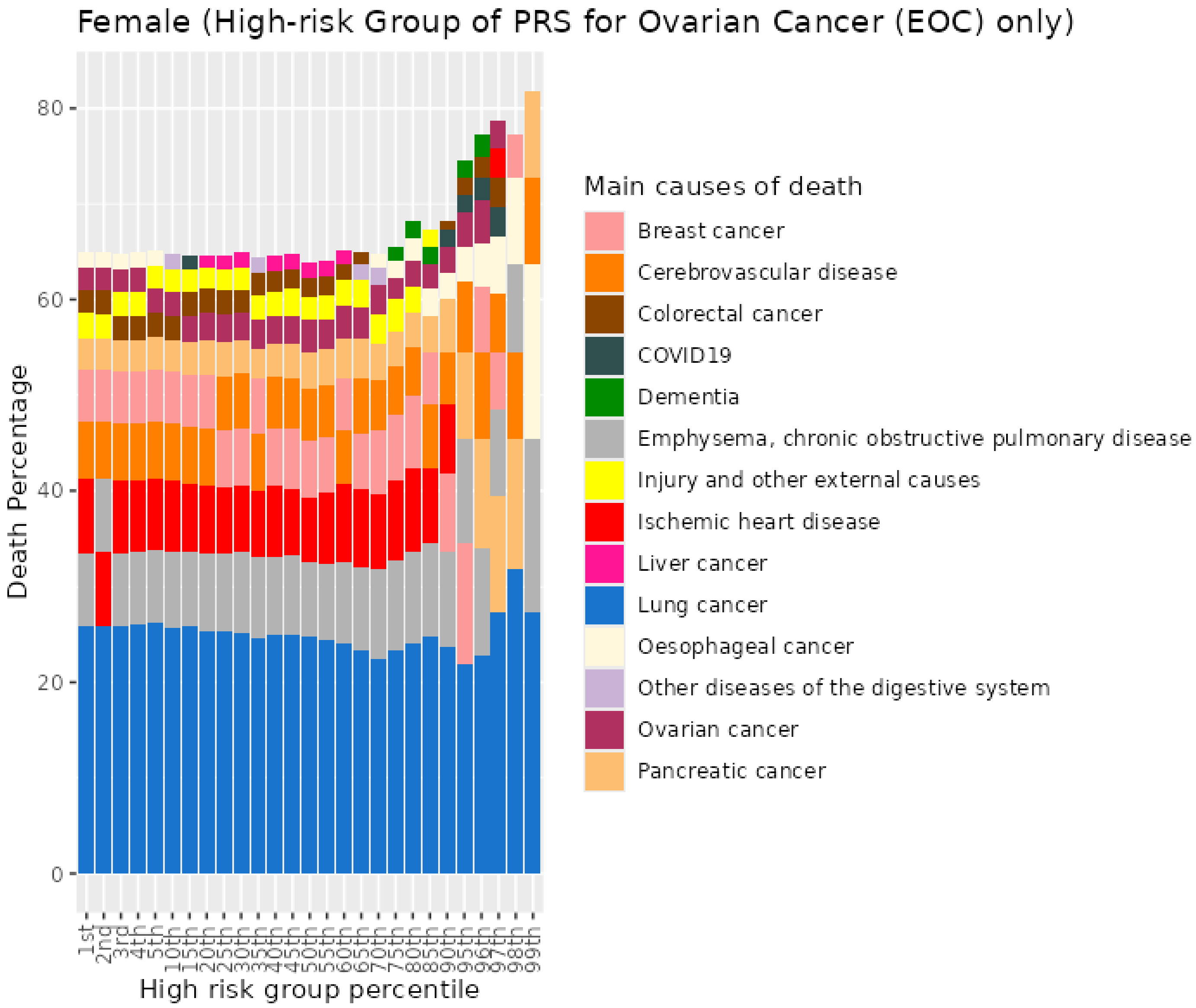


Figure S62. Percentiles from 1 to 99 for epithelial ovarian cancer PRS among female current smokers who died at age 60-69

##### 7.9 Female current smokers who died at age 70+

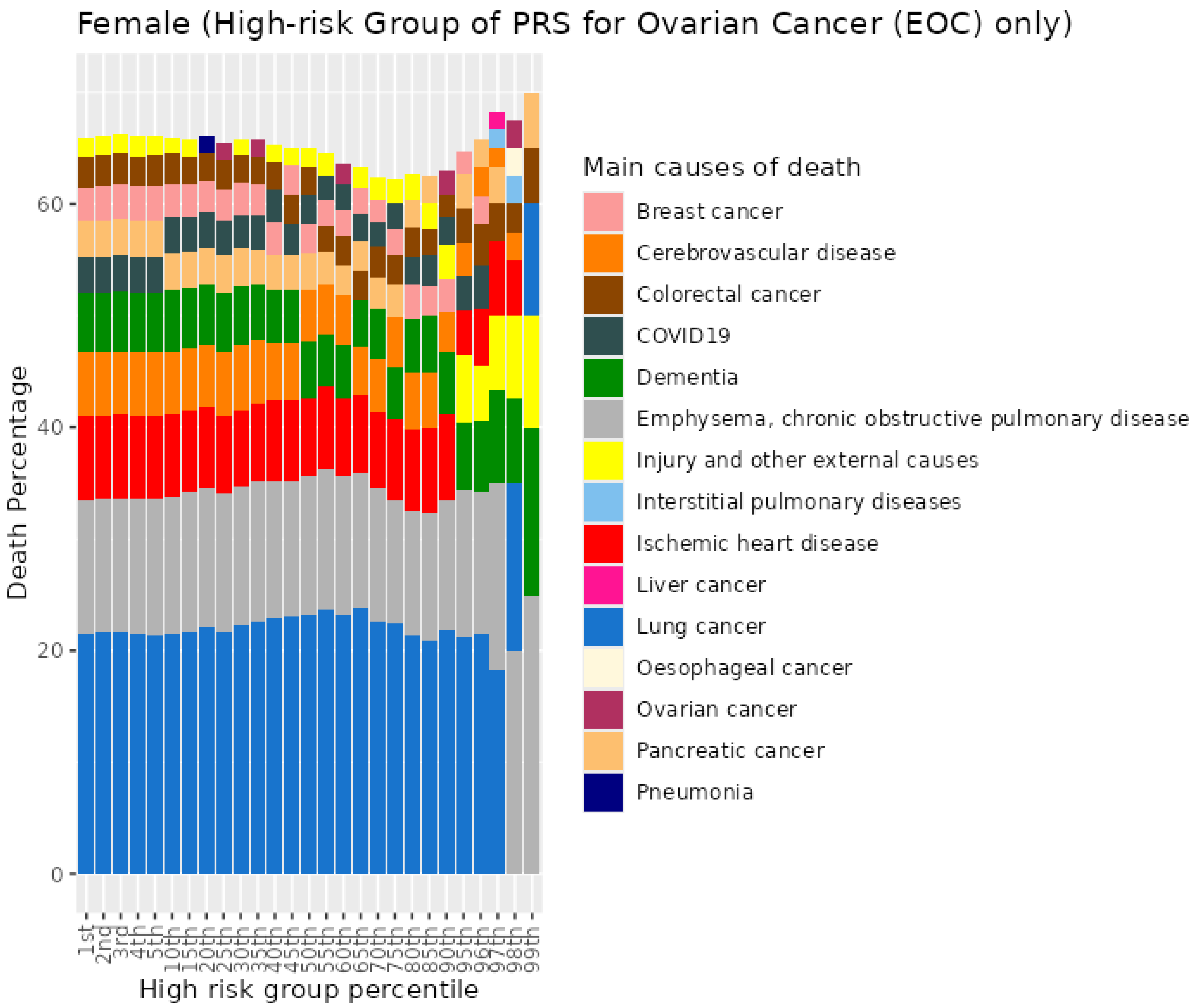


Figure S63. Percentiles from 1 to 99 for epithelial ovarian cancer PRS among female current smokers who died at age 70+

### 8 Alzheimer's disease (AD) PRS for UKB males

#### 8.1 Male never smokers who died at age <=59

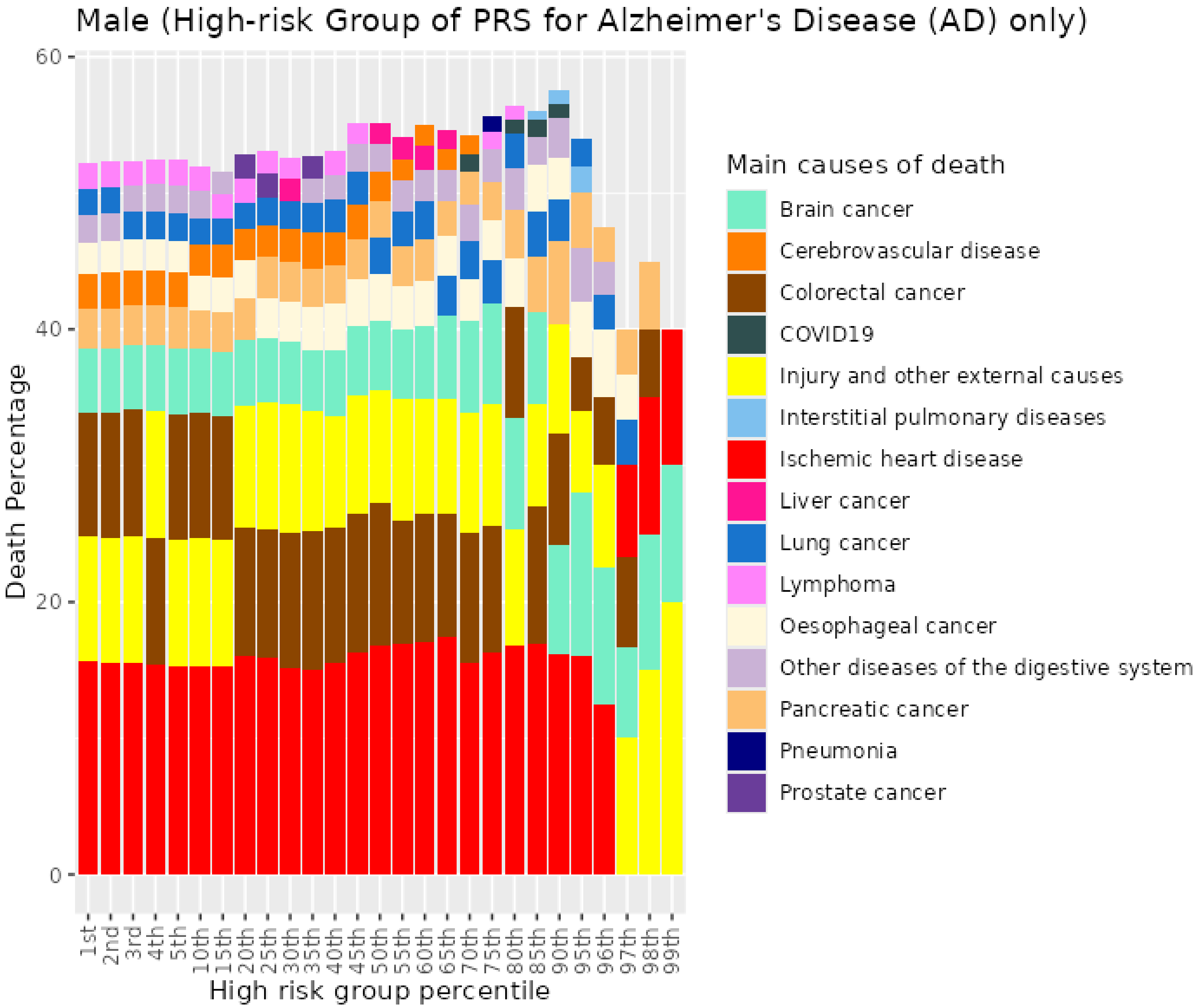


*Figure S64. Percentiles from 1 to 99 for Alzheimer's disease PRS among male never smokers who died at age <=59*

##### 8.2 Male never smokers who died at age 60-69

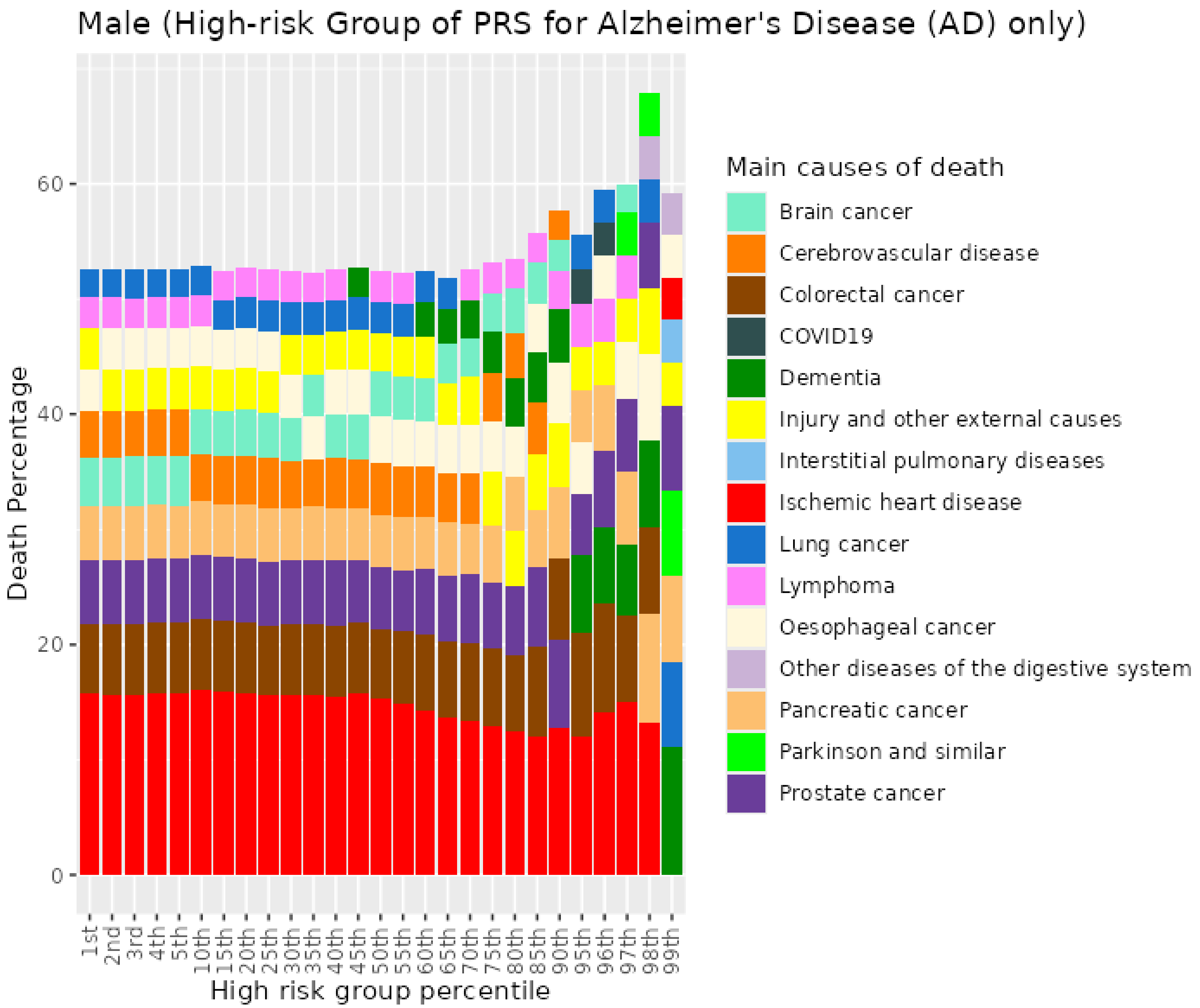


*Figure S65. Percentiles from 1 to 99 for Alzheimer's disease PRS among male never smokers who died at age 60-69*

##### 8.3 Male never smokers who died at age 70+

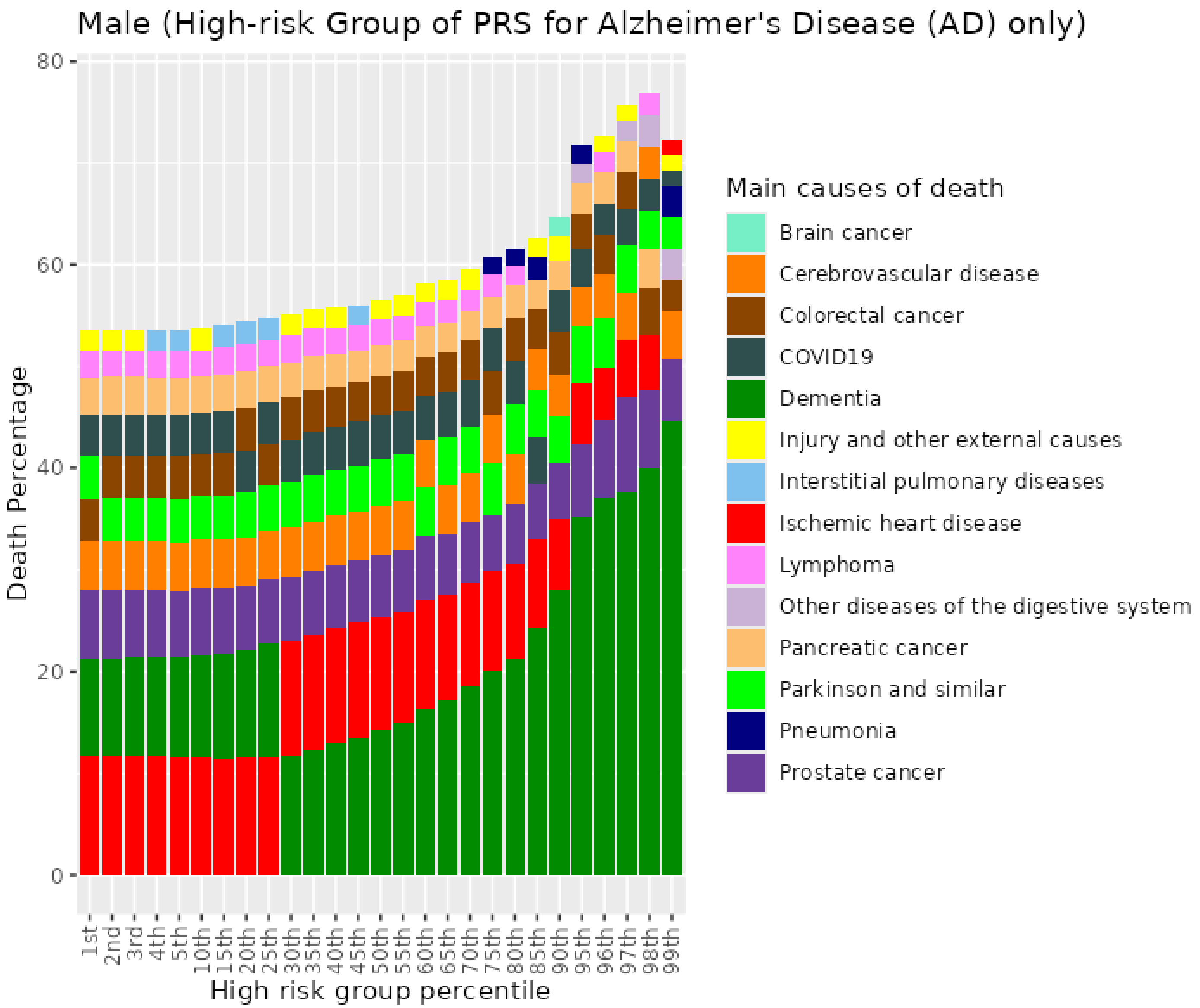


*Figure S66. Percentiles from 1 to 99 for Alzheimer's disease PRS among male never smokers who died at age 70+*

##### 8.4 Male previous smokers who died at <=59

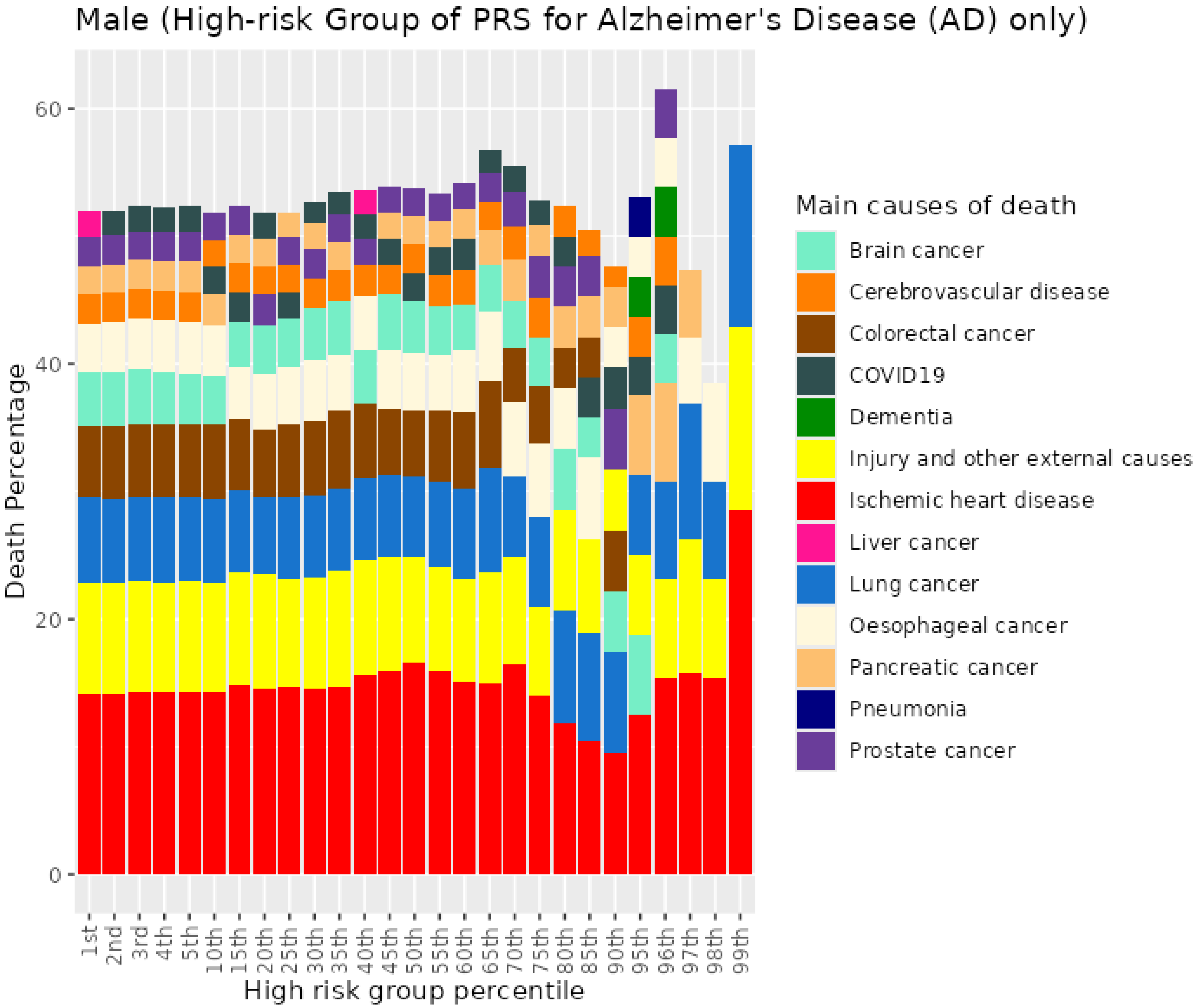


*Figure S67. Percentiles from 1 to 99 for Alzheimer's disease PRS among male previous smokers who died at age <=59*

##### 8.5 Male previous smokers who died at age 60-69

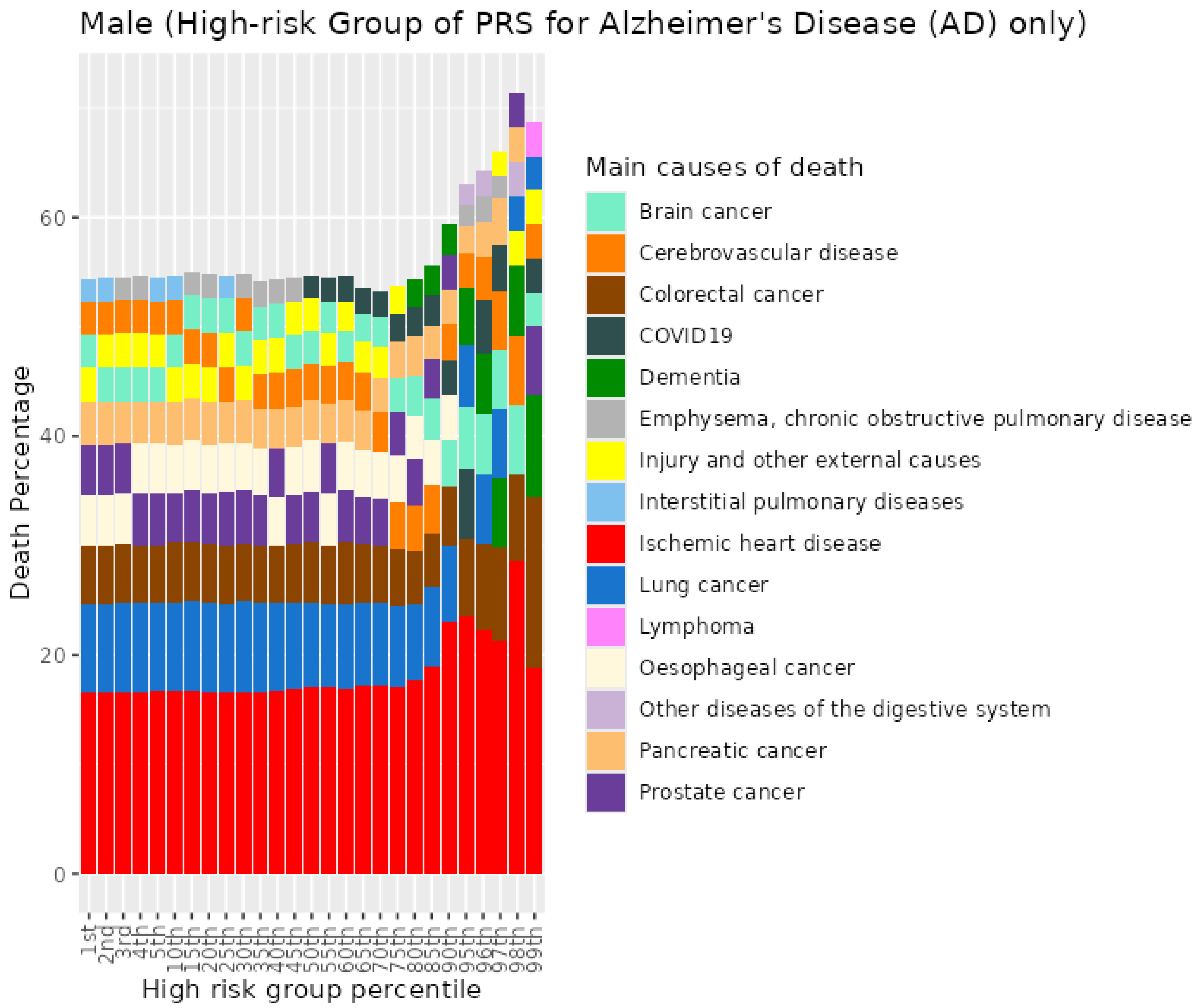


*Figure S68. Percentiles from 1 to 99 for Alzheimer's disease PRS among male previous smokers who died at age 60-69*

#### 8.6 Male previous smokers who died at age 70+

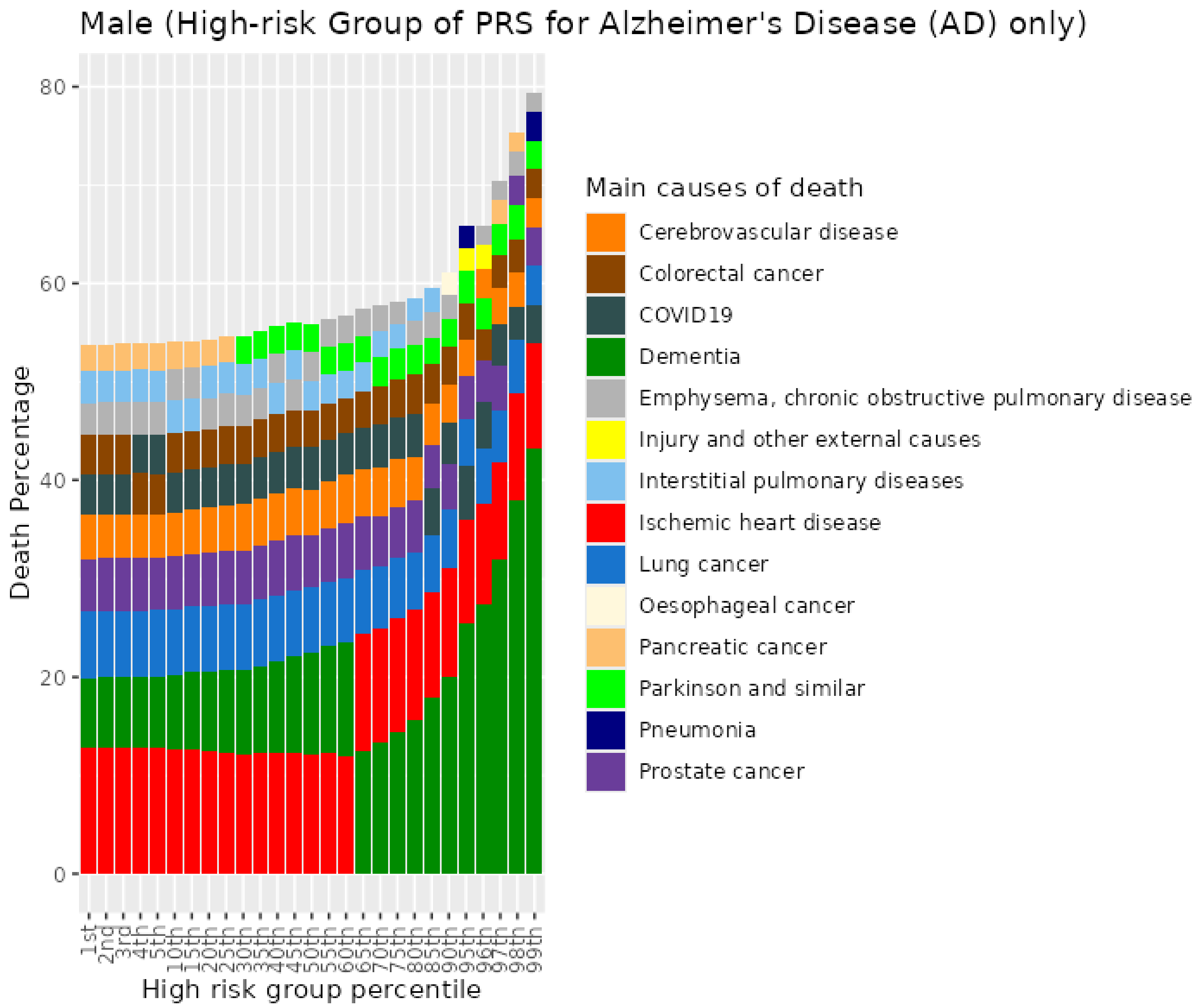


*Figure S69. Percentiles from 1 to 99 for Alzheimer's disease PRS among male previous smokers who died at age 70+*

#### 8.7 Male current smokers who died at age <=59

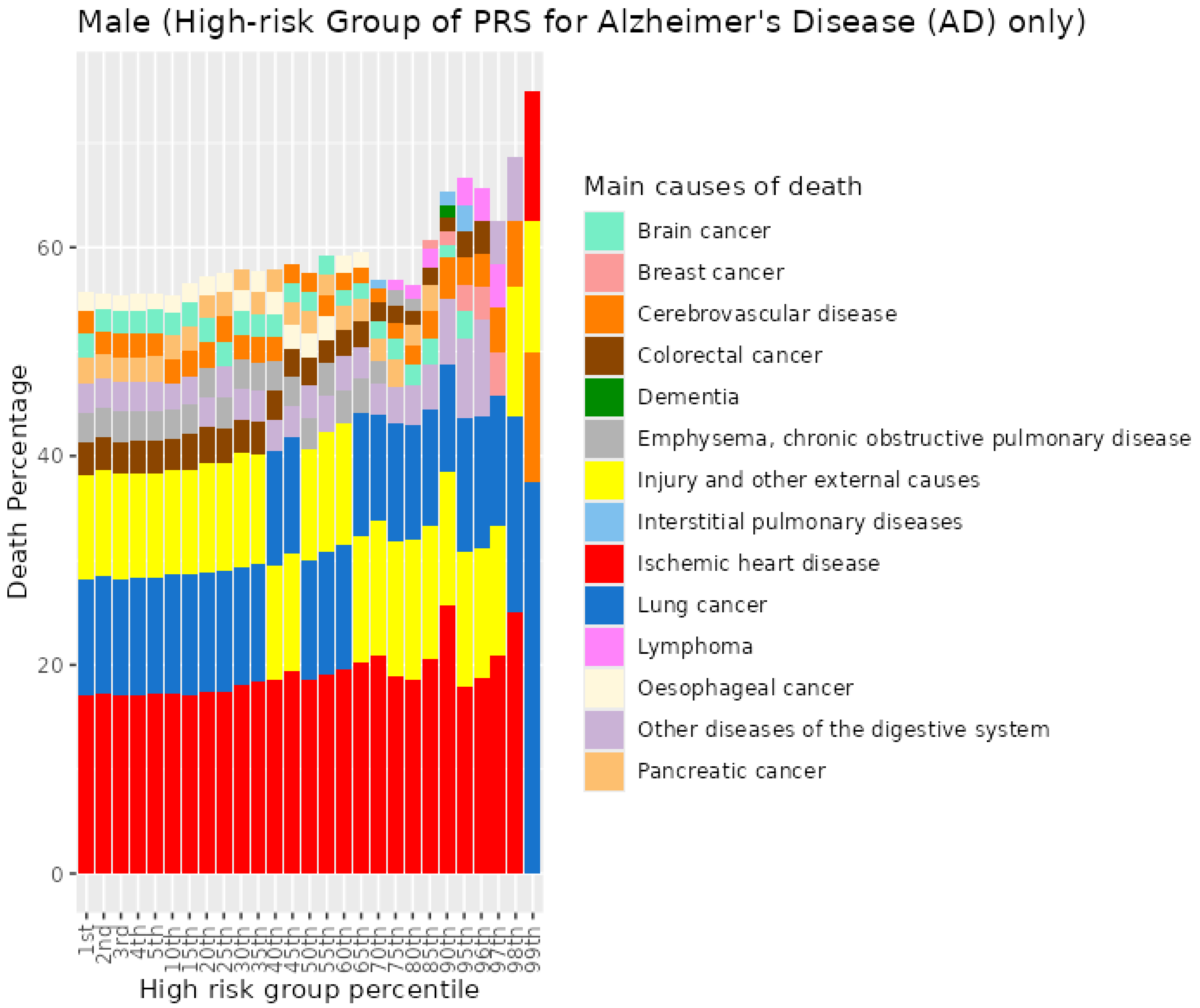


*Figure S70. Percentiles from 1 to 99 for Alzheimer's disease PRS among male current smokers who died at age <=59*

#### 8.8 Male current smokers who died at age 60-69

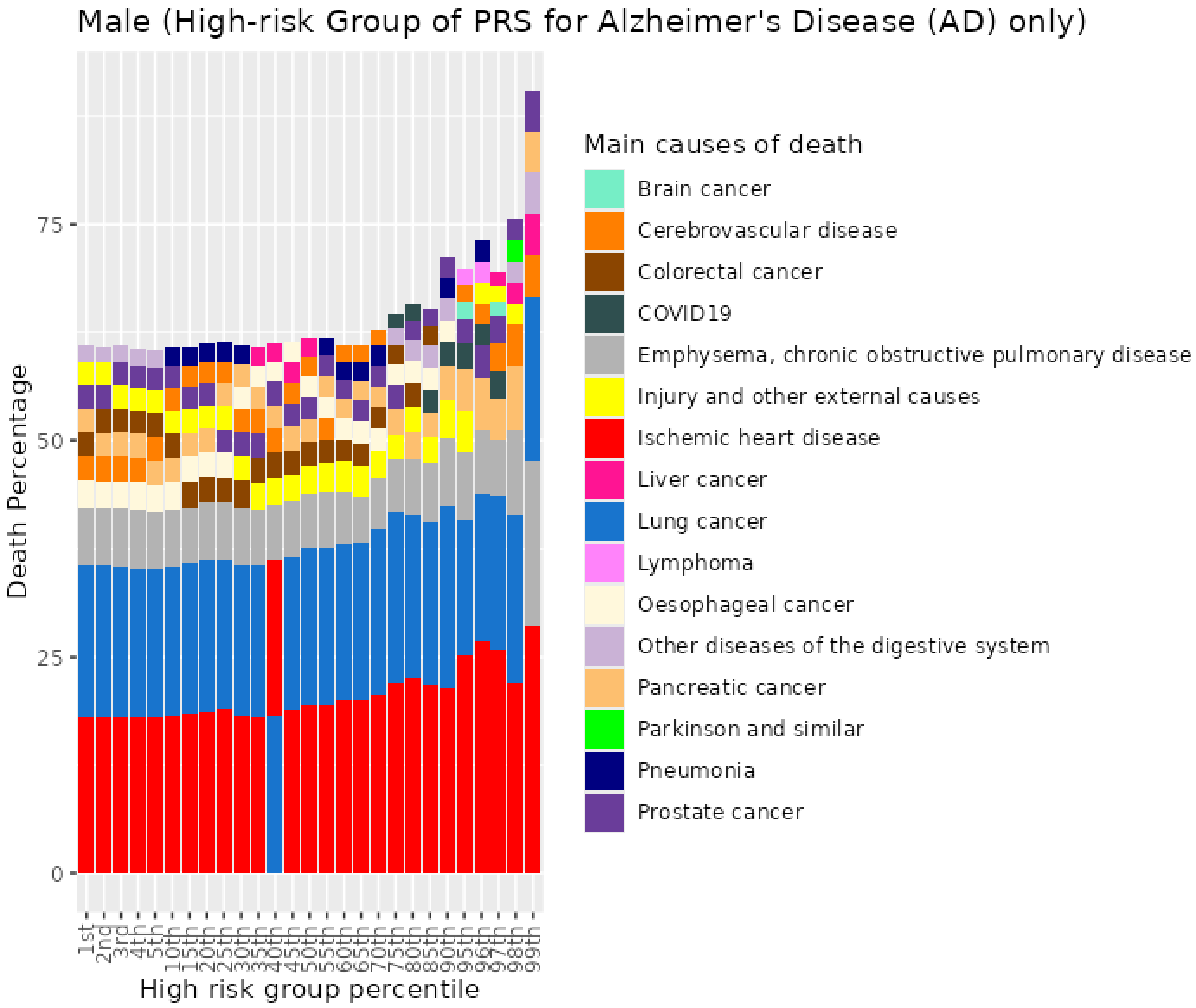


*Figure S71. Percentiles from 1 to 99 for Alzheimer's disease PRS among male current smokers who died at age 60-69*

##### 8.9 Male current smokers who died at age 70+

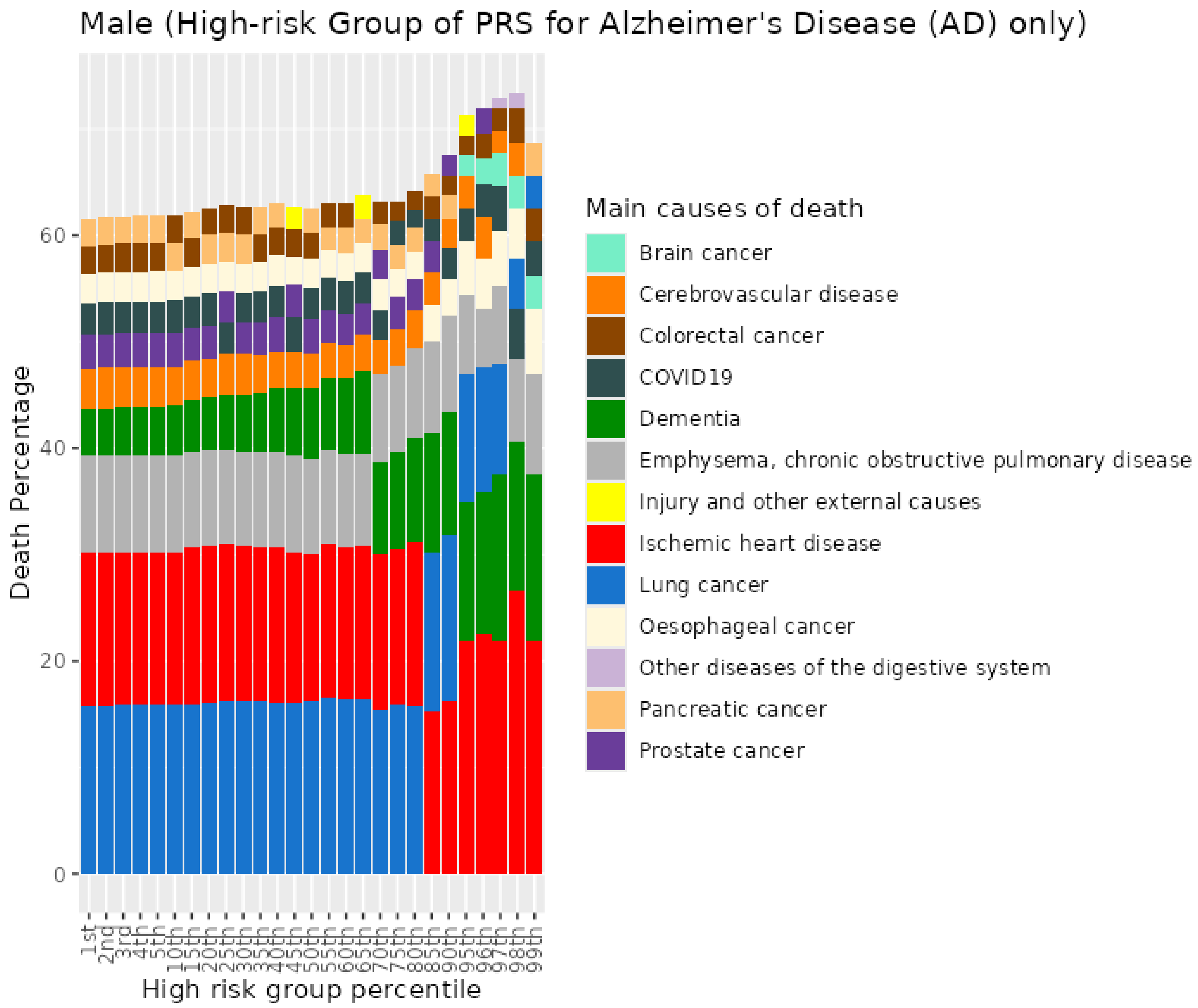


*Figure S72. Percentiles from 1 to 99 for Alzheimer's disease PRS among male current smokers who died at age 70+*

### 9 Bowel Cancer (CRC) PRS for UKB males

#### 9.1 Male never smokers who died at age <=59

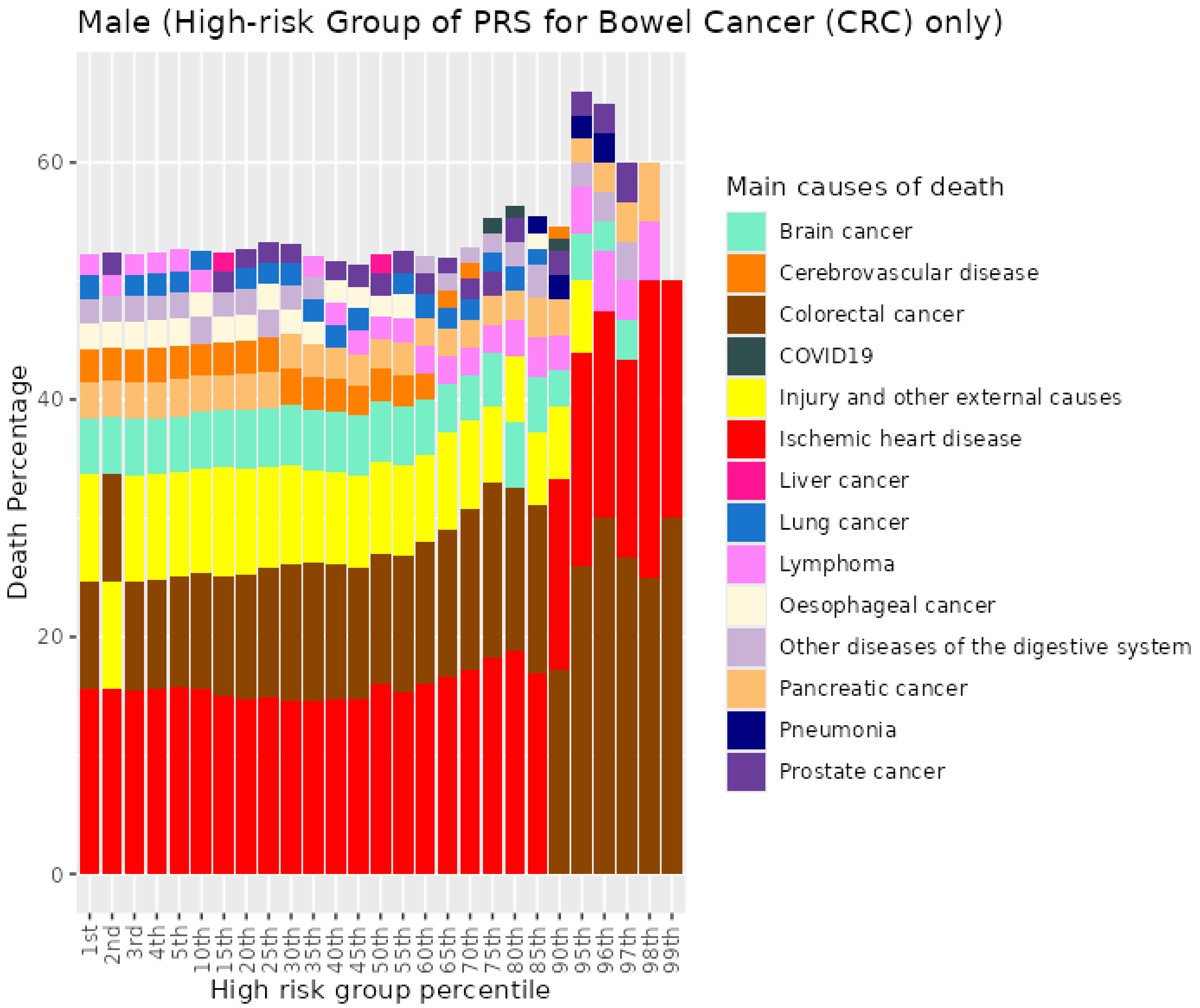


*Figure S73. Percentiles from 1 to 99 for bowel cancer PRS among male never smokers who died at age <=59*

#### 9.2 Male never smokers who died at age 60-69

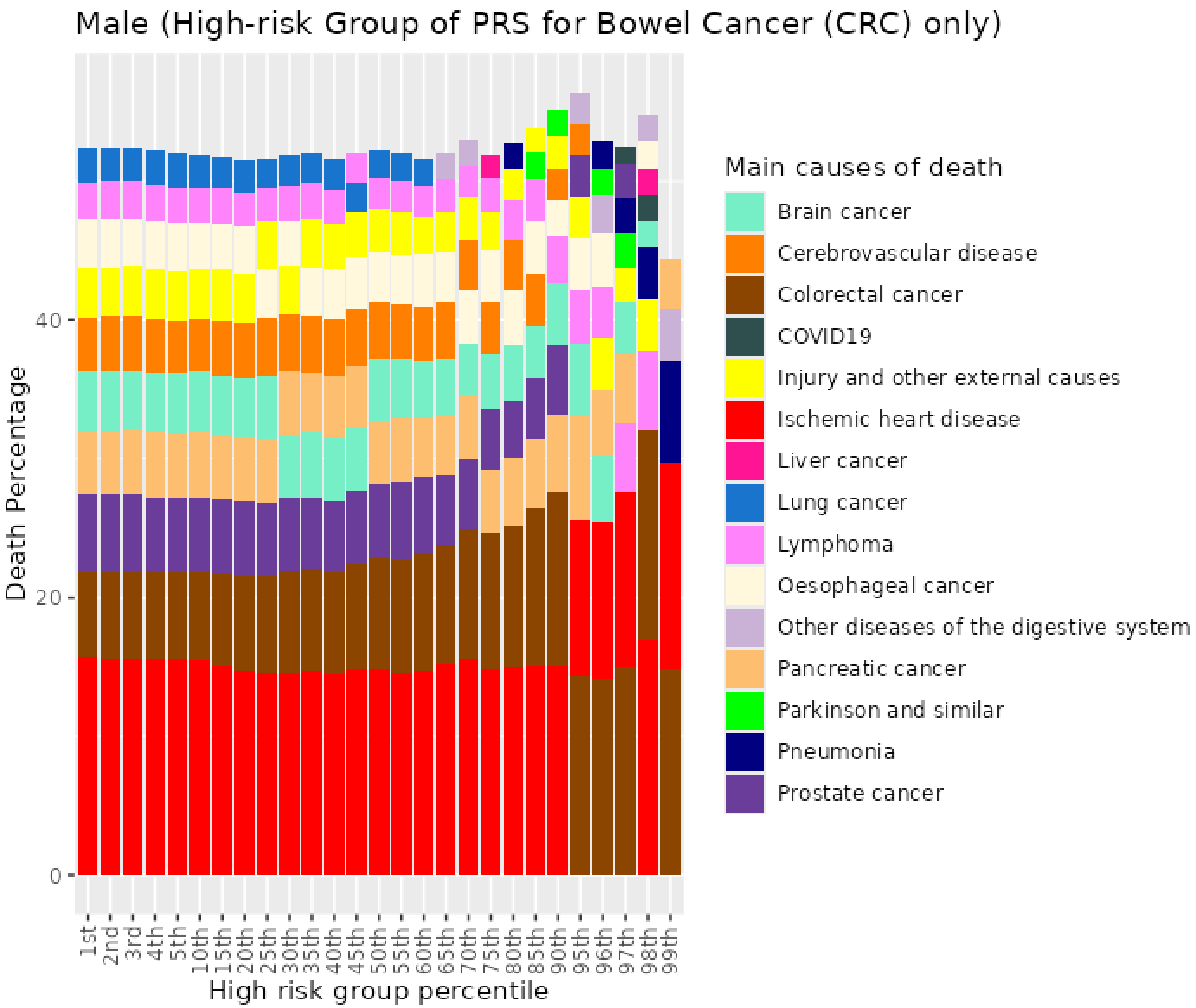


*Figure S74. Percentiles from 1 to 99 for bowel cancer PRS among male never smokers who died at age 60-69*

##### 9.3 Male never smokers who died at age 70+

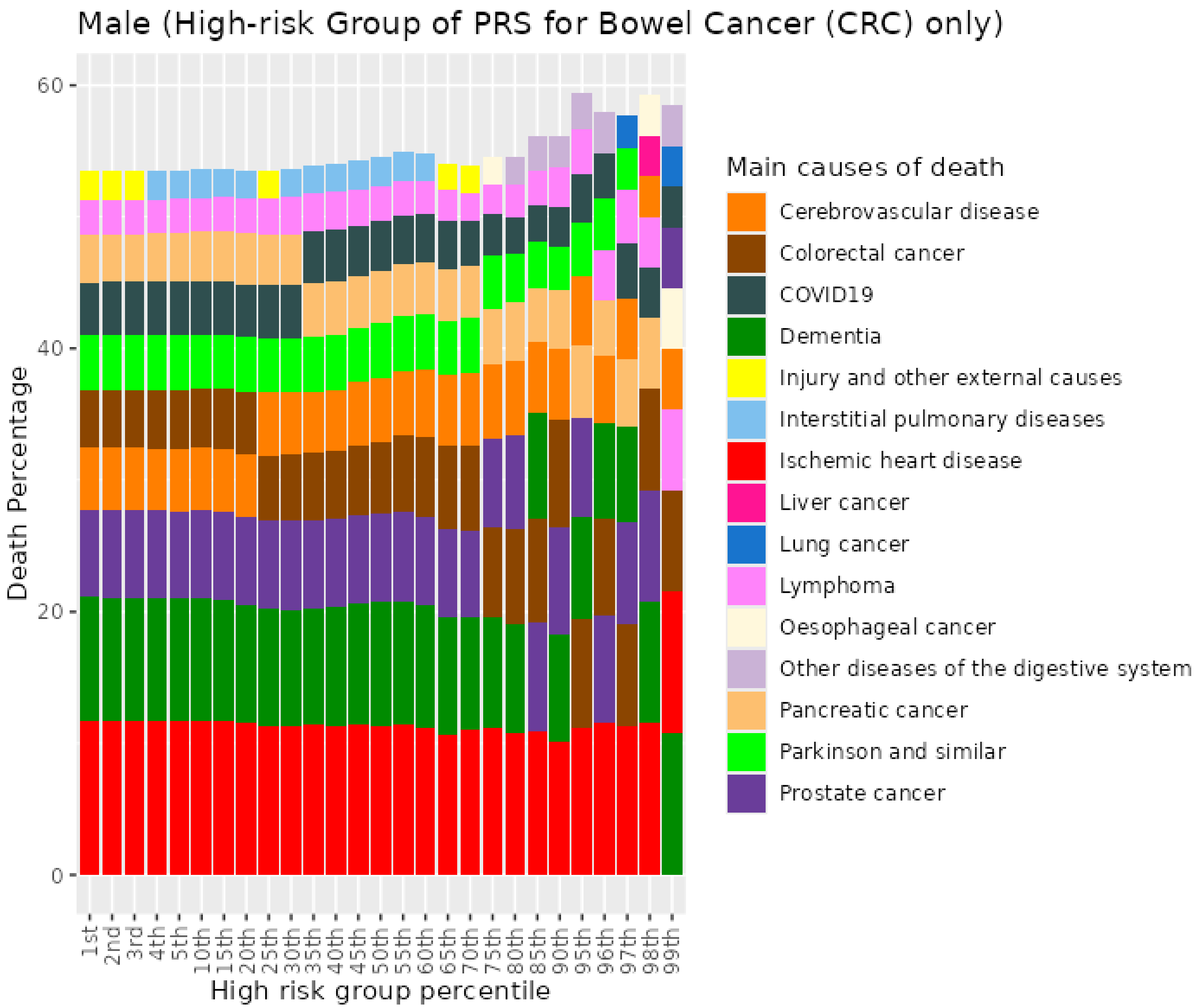


*Figure S75. Percentiles from 1 to 99 for bowel cancer PRS among male never smokers who died at age 70+*

##### 9.4 Male previous smokers who died at age <=59

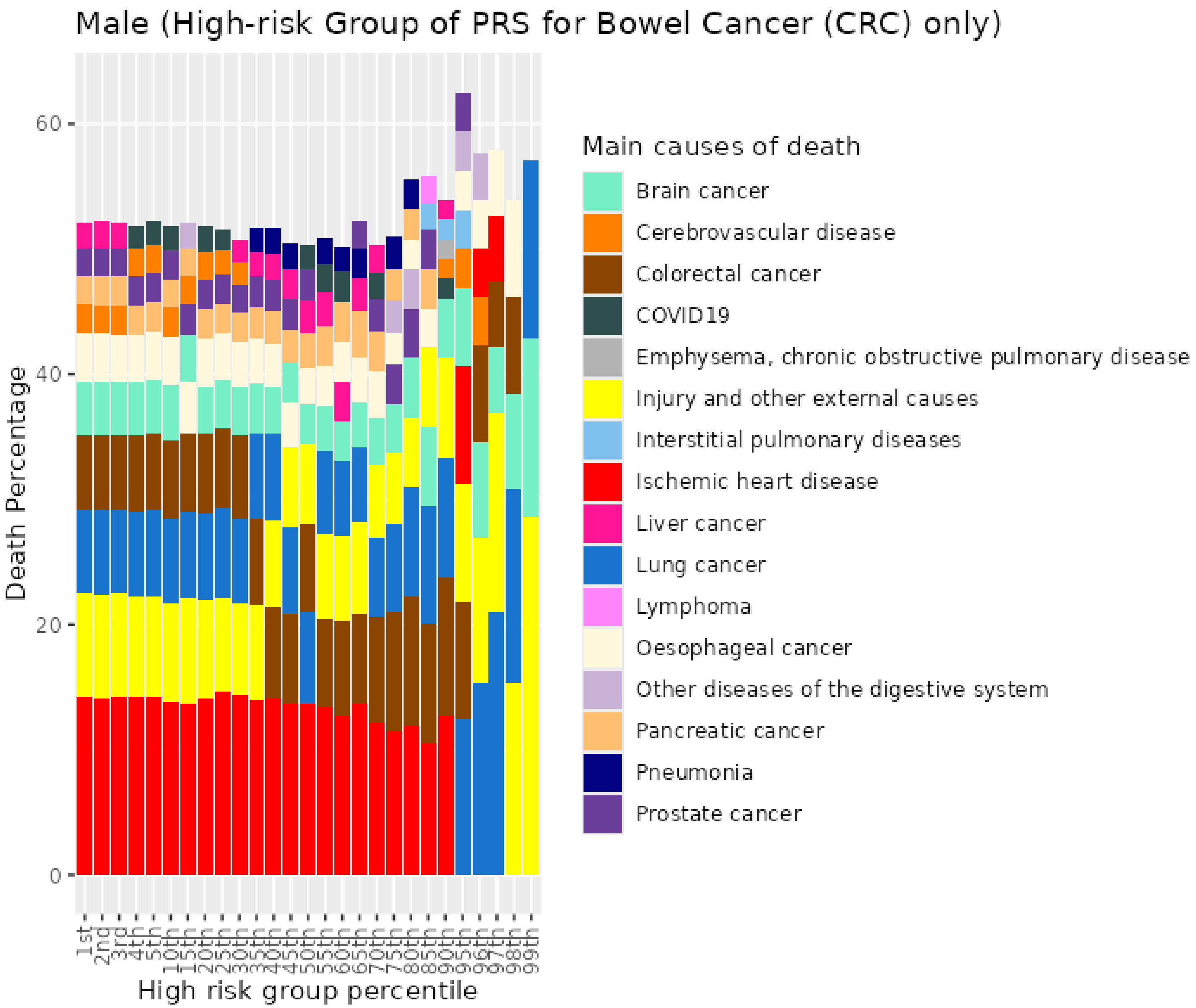


*Figure S76. Percentiles from 1 to 99 for bowel cancer PRS among male previous smokers who died at age <=59*

#### 9.5 Male previous smokers who died at age 60-69

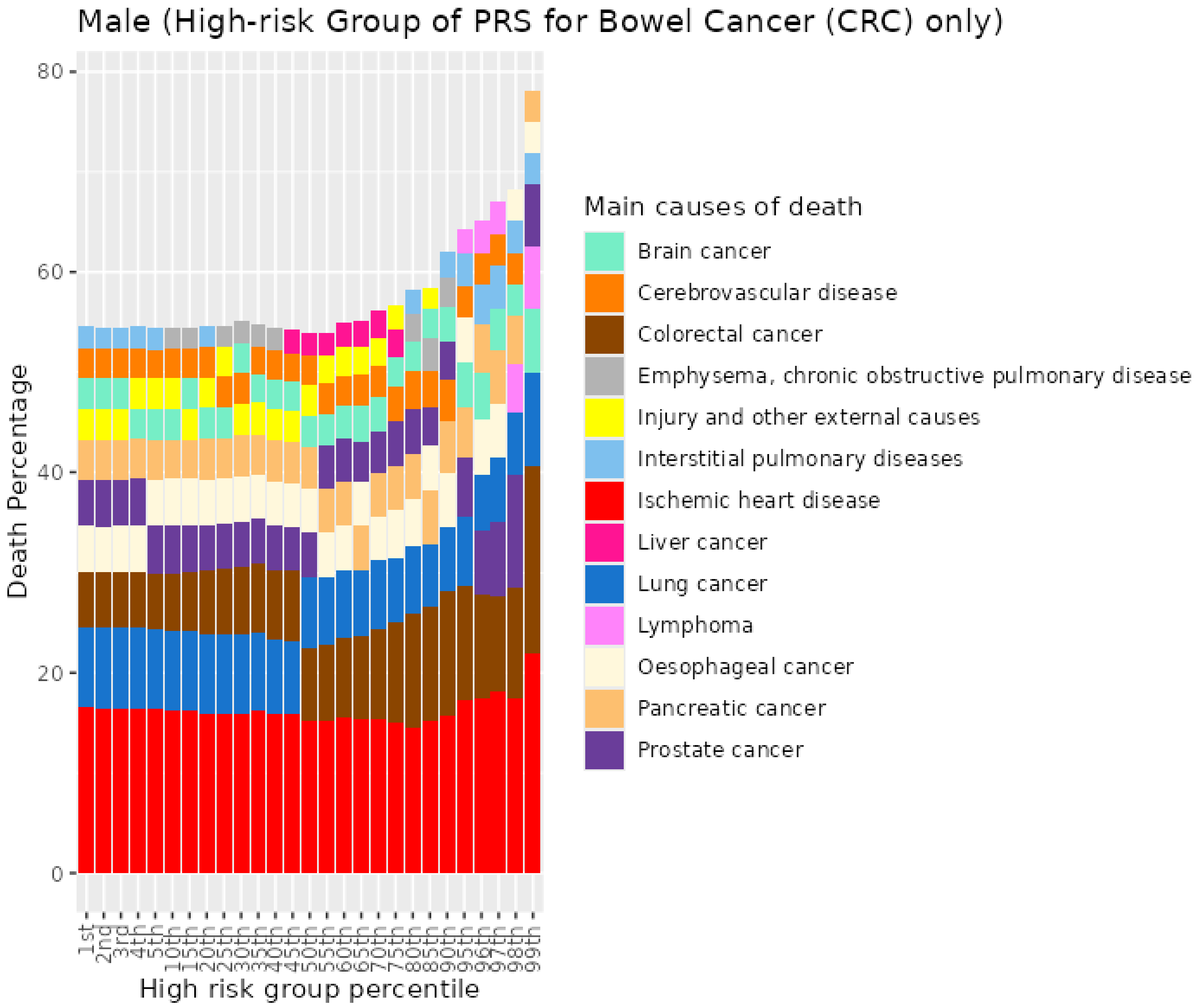


*Figure S77. Percentiles from 1 to 99 for bowel cancer PRS among male previous smokers who died at age 60-69*

##### 9.6 Male previous smokers who died at age 70+

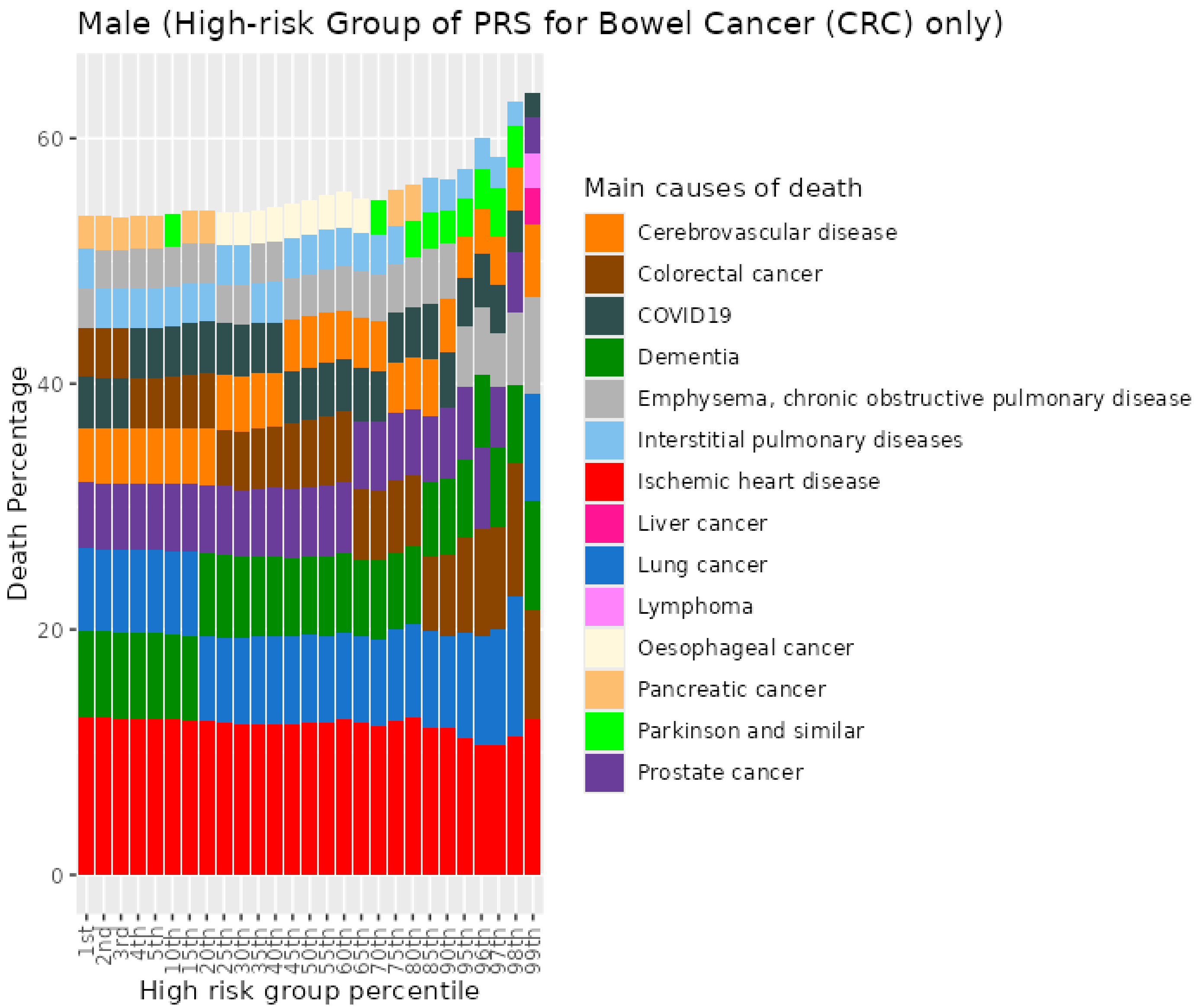


*Figure S78. Percentiles from 1 to 99 for bowel cancer PRS among male previous smokers who died at age 70+*

#### 9.7 Male current smokers who died at age <=59

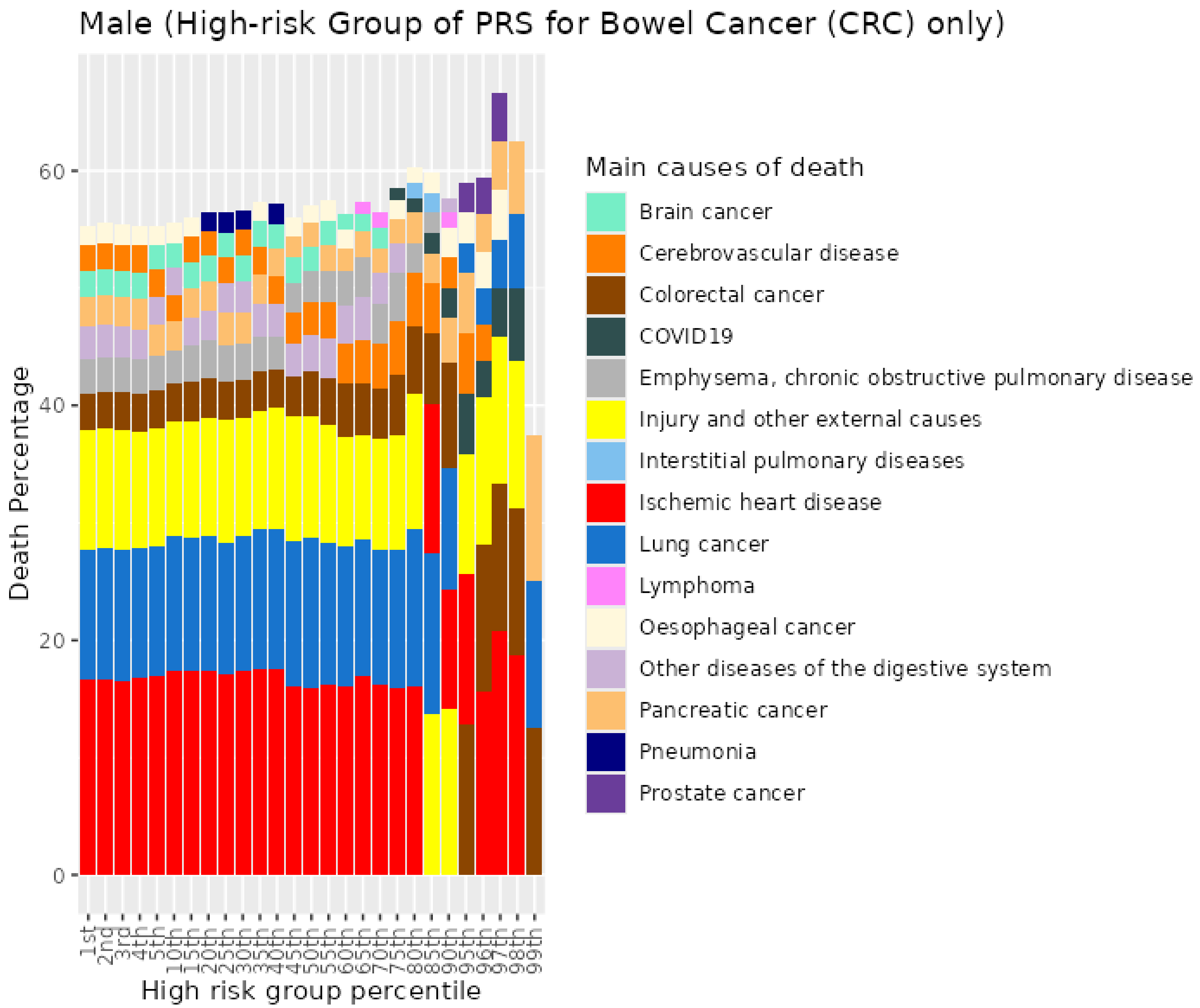


*Figure S79. Percentiles from 1 to 99 for bowel cancer PRS among male current smokers who died at age <=59*

##### 9.8 Male current smokers who died at age 60-69

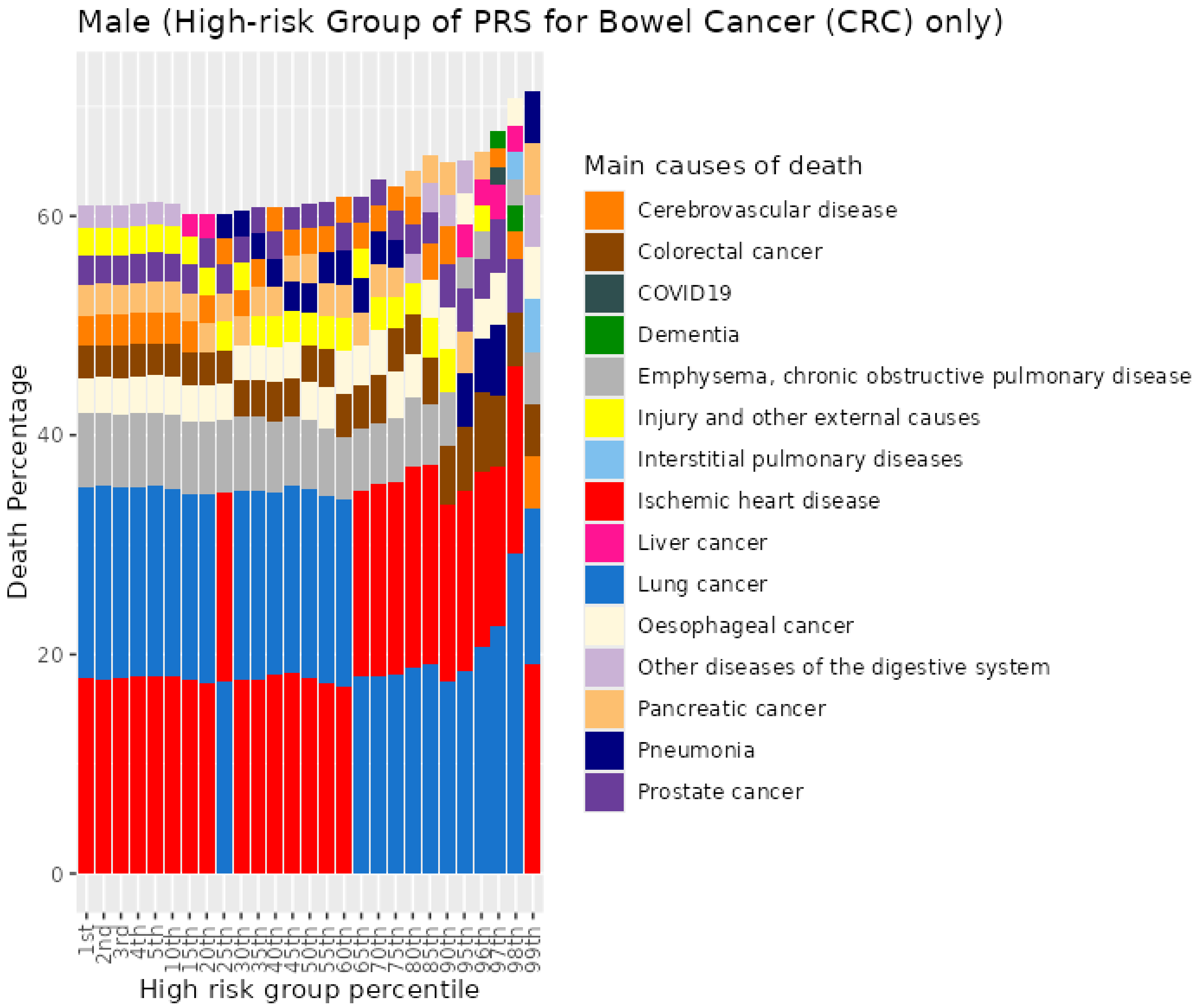


*Figure S80. Percentiles from 1 to 99 for bowel cancer PRS among male current smokers who died at age 60-69*

##### 9.9 Male current smokers who died at age 70+

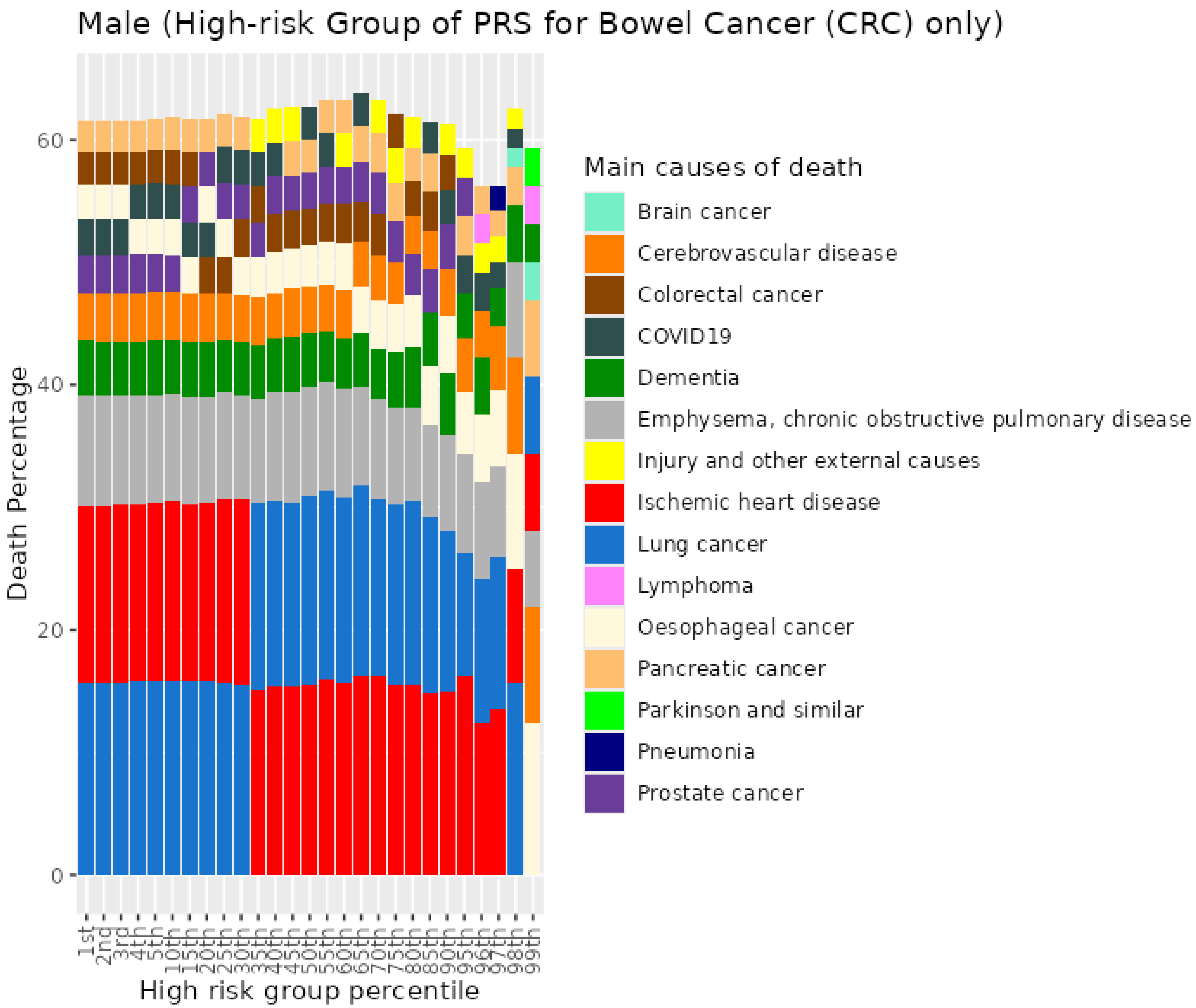


*Figure S81. Percentiles from 1 to 99 for bowel cancer PRS among male current smokers who died at age 70+*

### 10 Cardiovascular disease (CVD) PRS for UKB males

#### 10.1 Male never smokers who died at age <=59

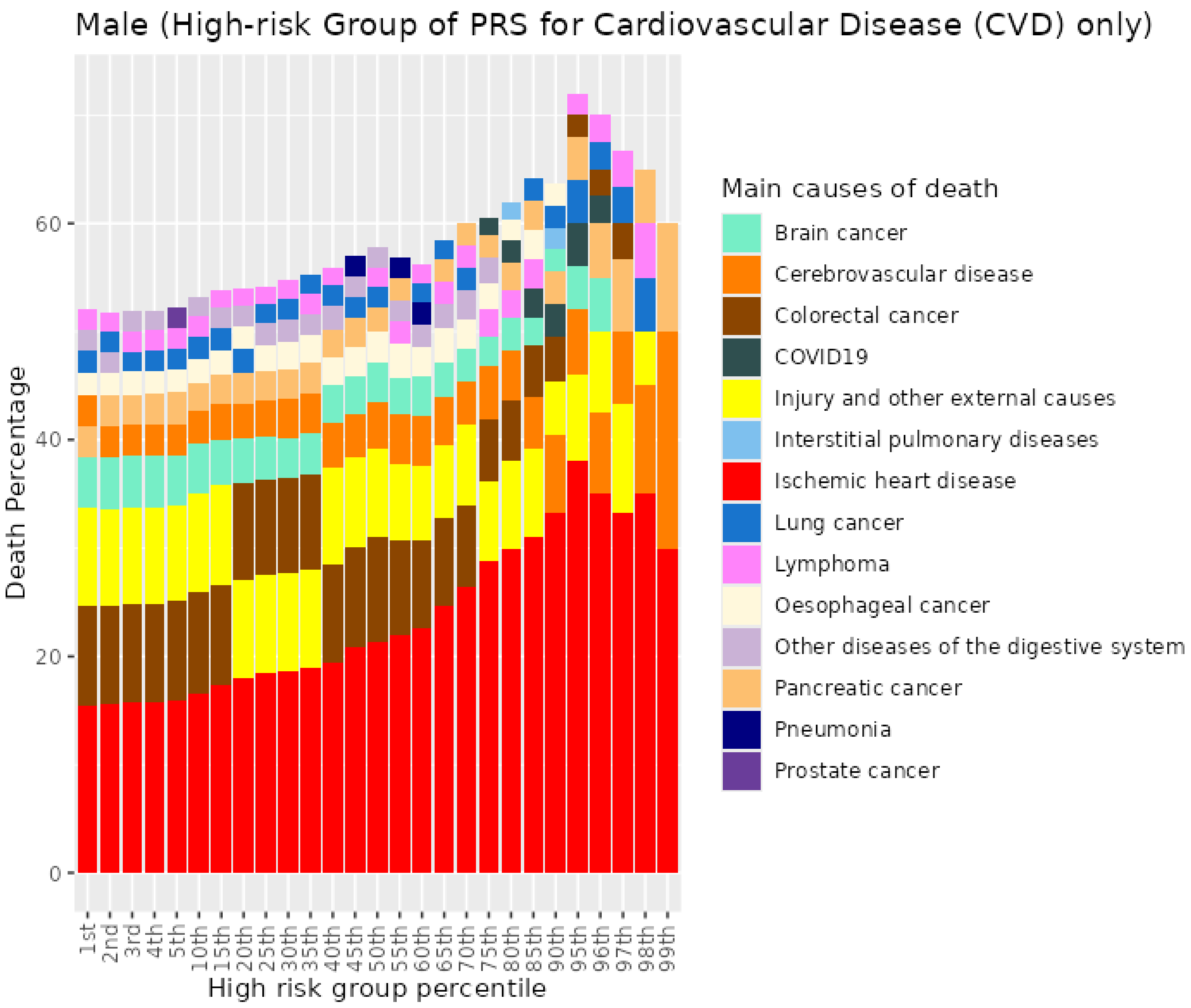


*Figure S82. Percentiles from 1 to 99 for cardiovascular disease PRS among male never smokers who died at age <=59*

##### 10.2 Male never smokers who died at age 60-69

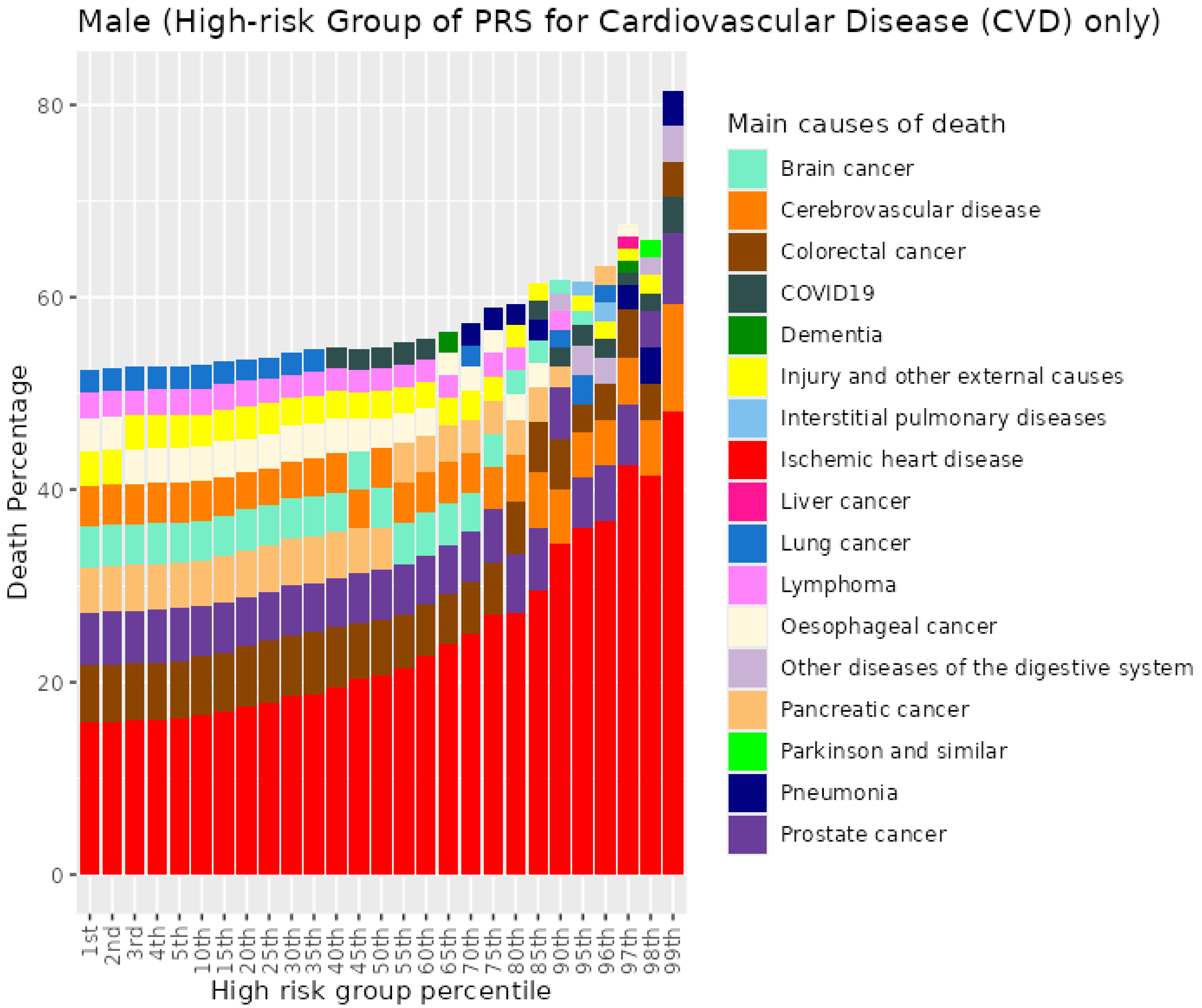


*Figure S83. Percentiles from 1 to 99 for cardiovascular disease PRS among male never smokers who died at age 60-69*

##### 10.3 Male never smokers who died at age 70+

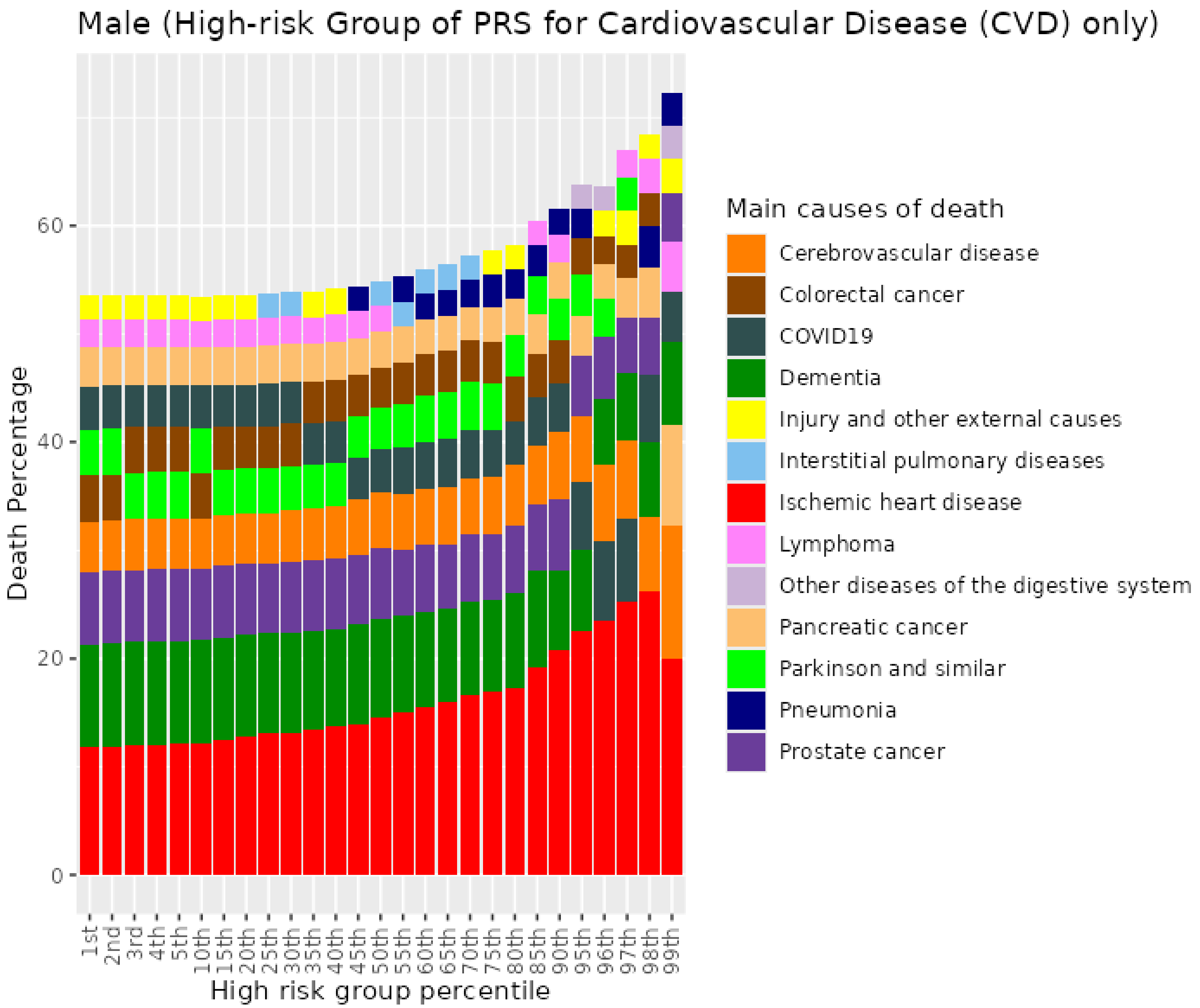


*Figure S84. Percentiles from 1 to 99 for cardiovascular disease PRS among male never smokers who died at age 70+*

##### 10.4 Male previous smokers who died at age <=59

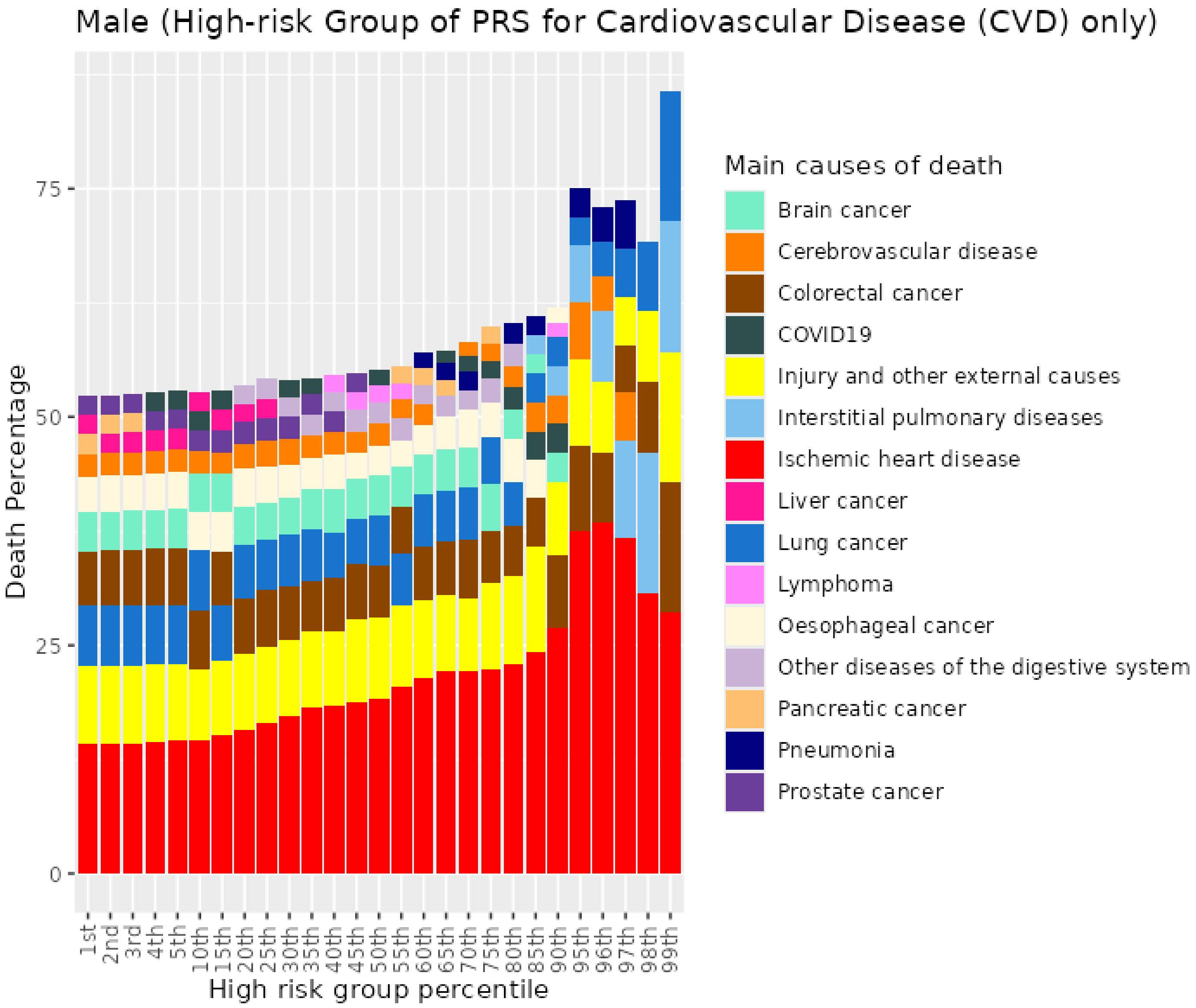


*Figure S85. Percentiles from 1 to 99 for cardiovascular disease PRS among male previous smokers who died at age <=59*

#### 10.5 Male previous smokers who died at age 60-69

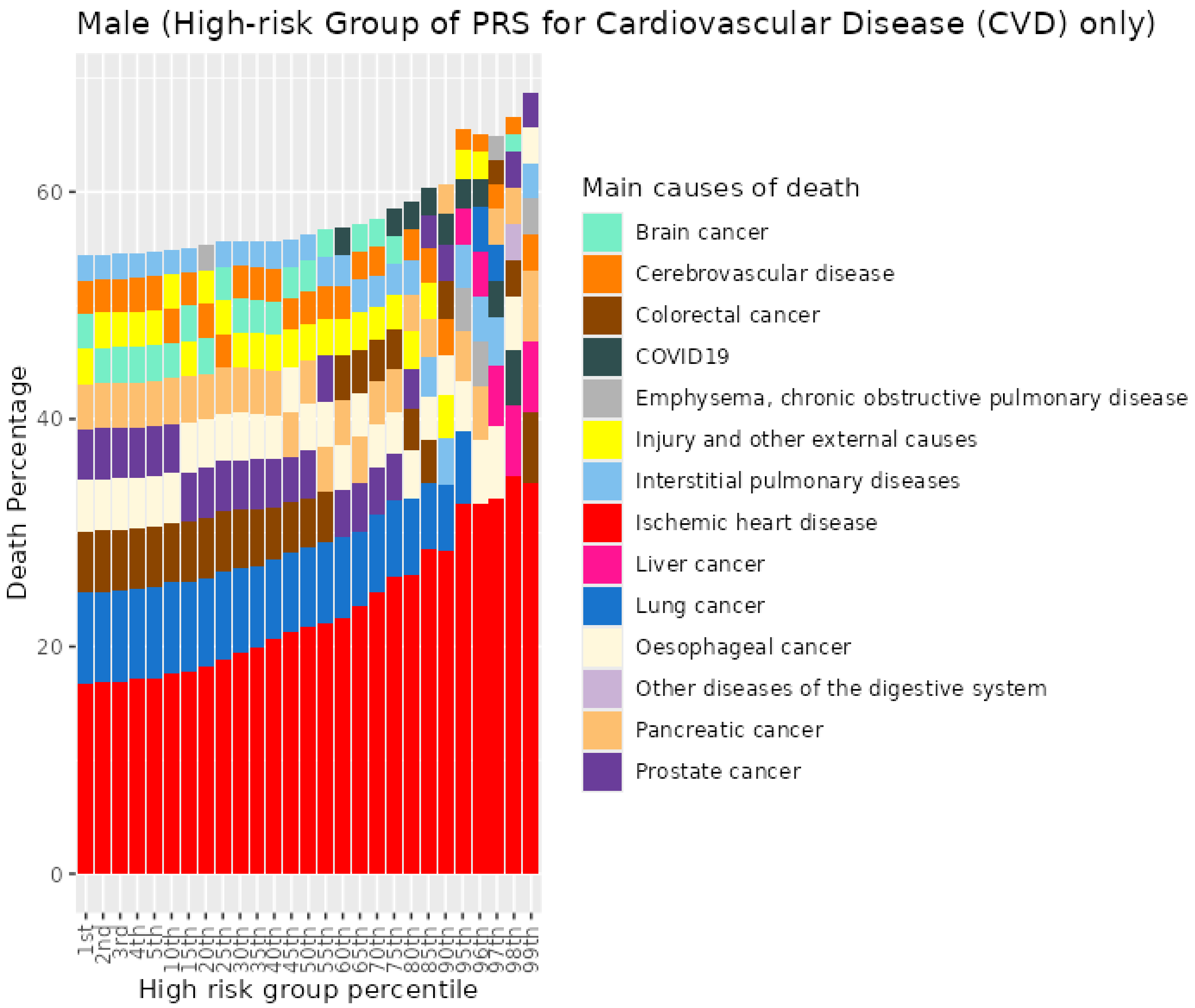


*Figure S86. Percentiles from 1 to 99 for cardiovascular disease PRS among male previous smokers who died at age 60-69*

##### 10.6 Male previous smokers who died at age 70+

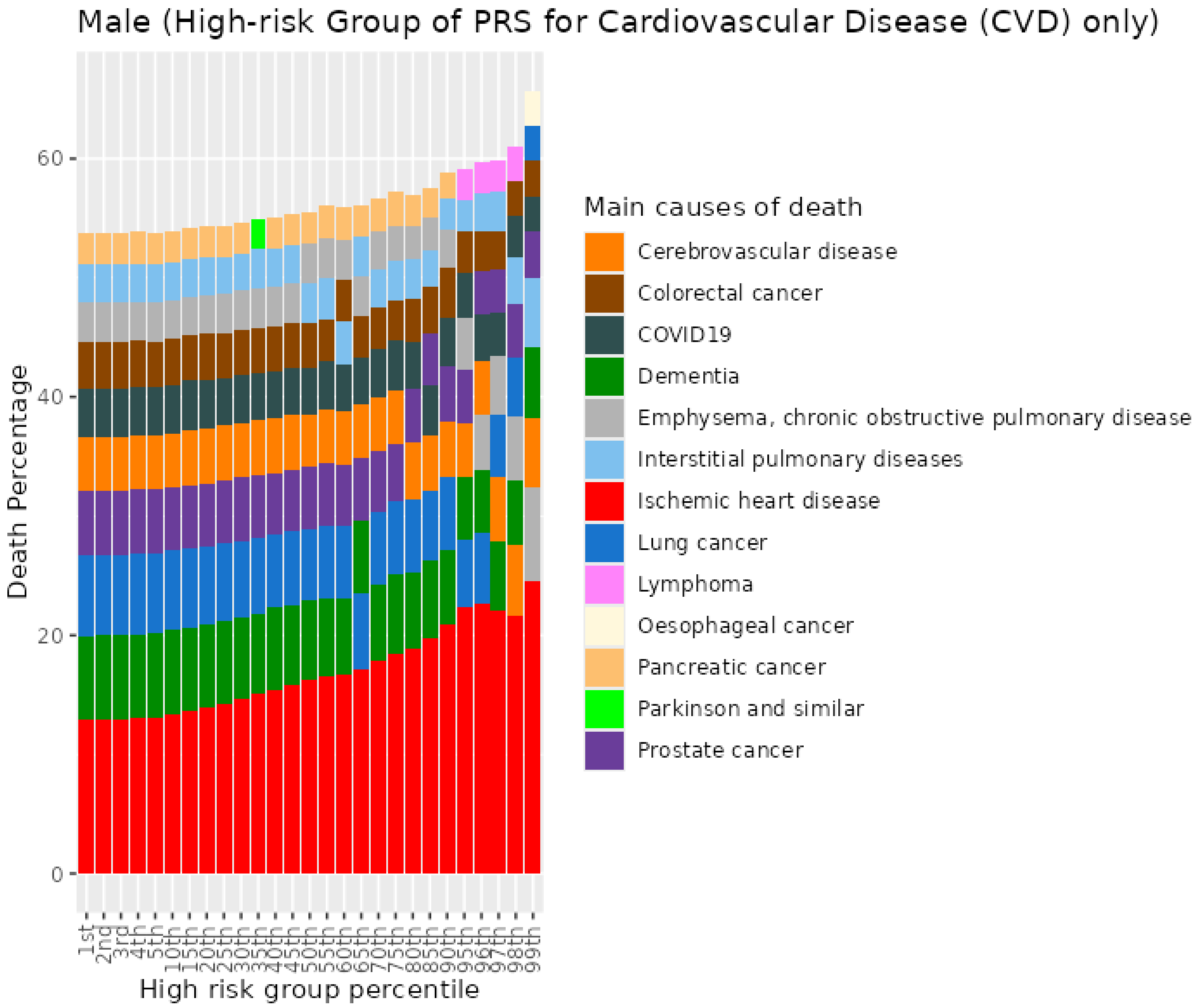


*Figure S87. Percentiles from 1 to 99 for cardiovascular disease PRS among male previous smokers who died at age 70+*

##### 10.7 Male current smokers who died at age <=59

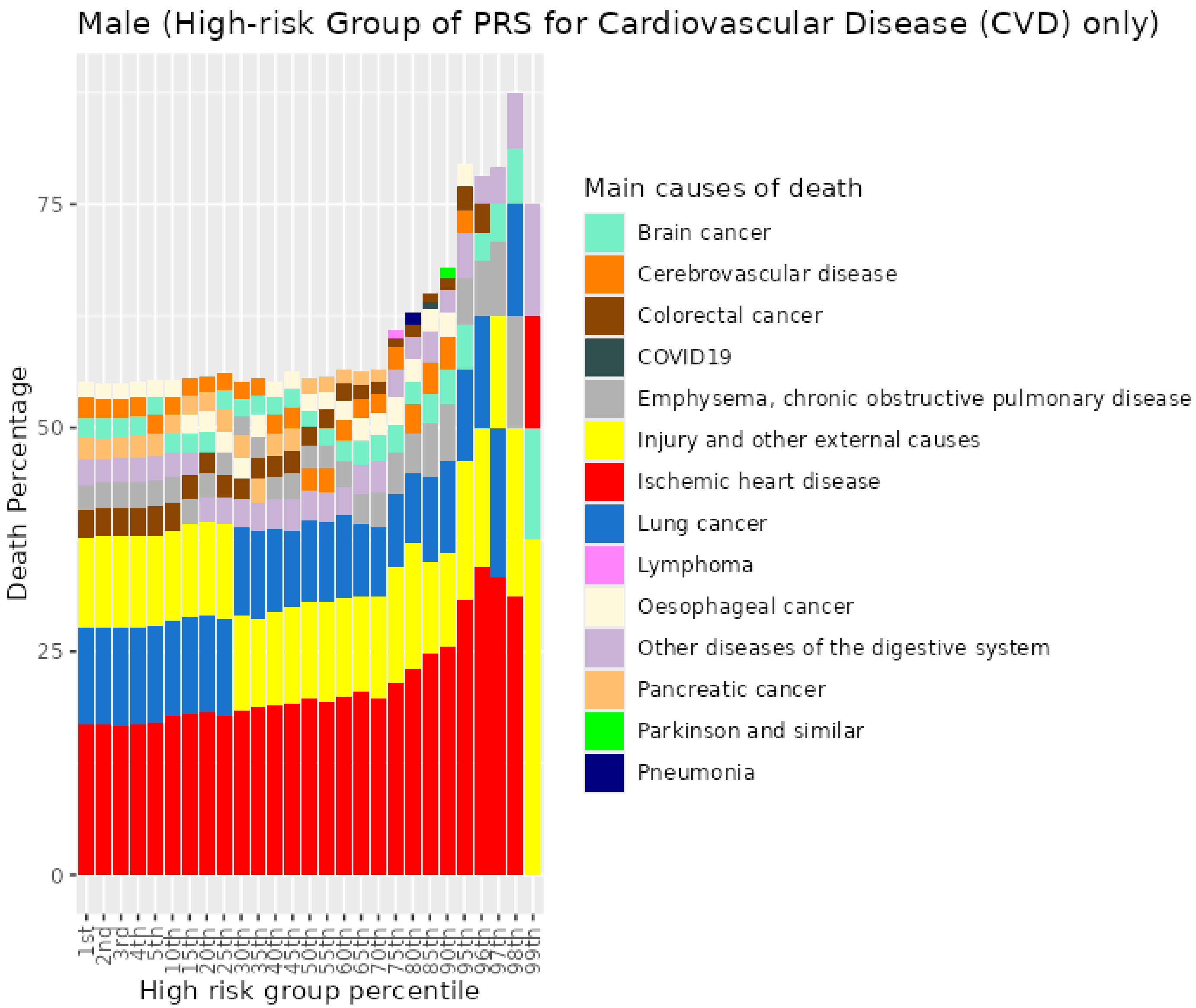


*Figure S88. Percentiles from 1 to 99 for cardiovascular disease PRS among male current smokers who died at age <=59*

##### 10.8 Male current smokers who died at age 60-69

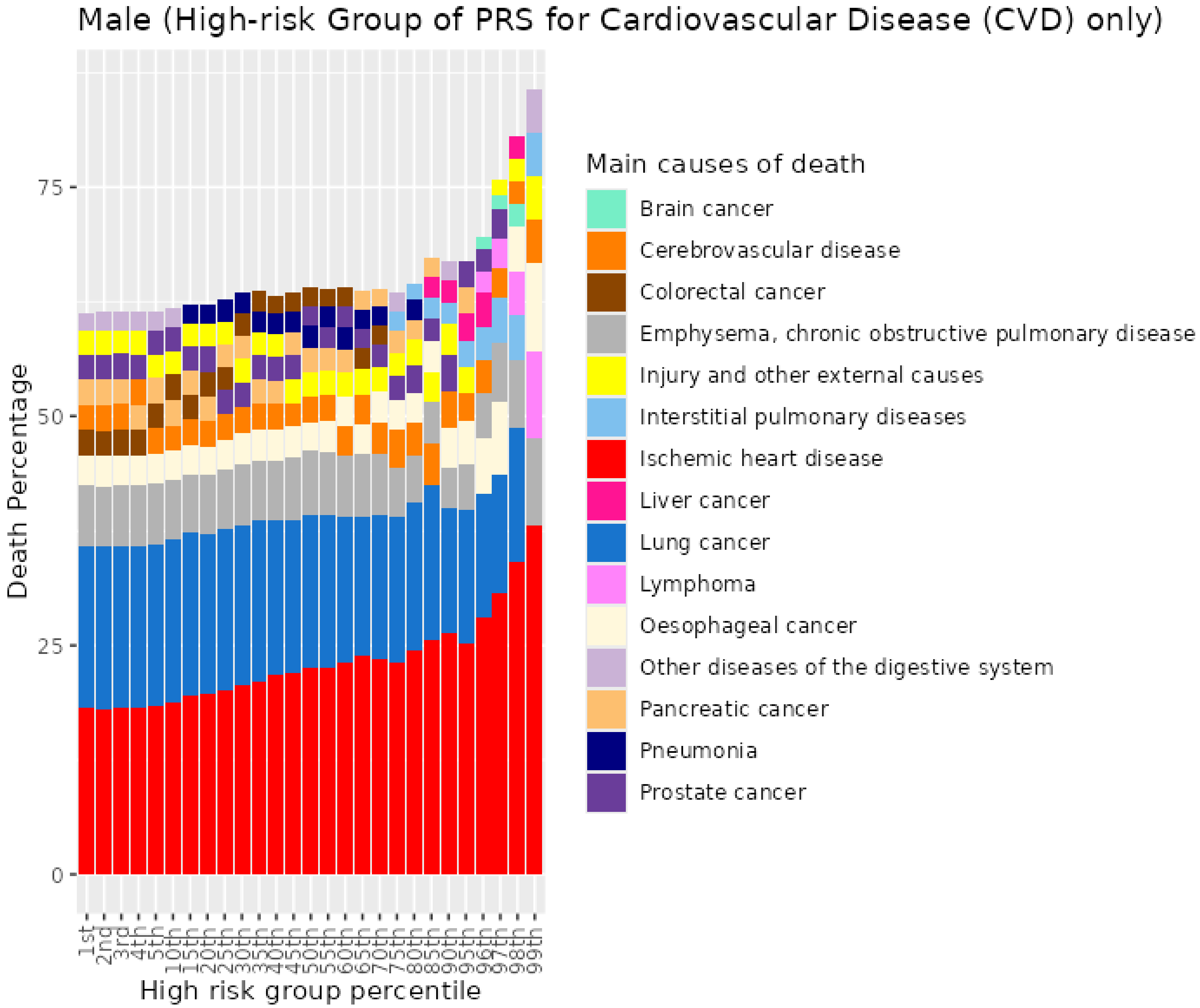


*Figure S89. Percentiles from 1 to 99 for cardiovascular disease PRS among male current smokers who died at age 60-69*

#### 10.9 Male current smokers who died at age 70+

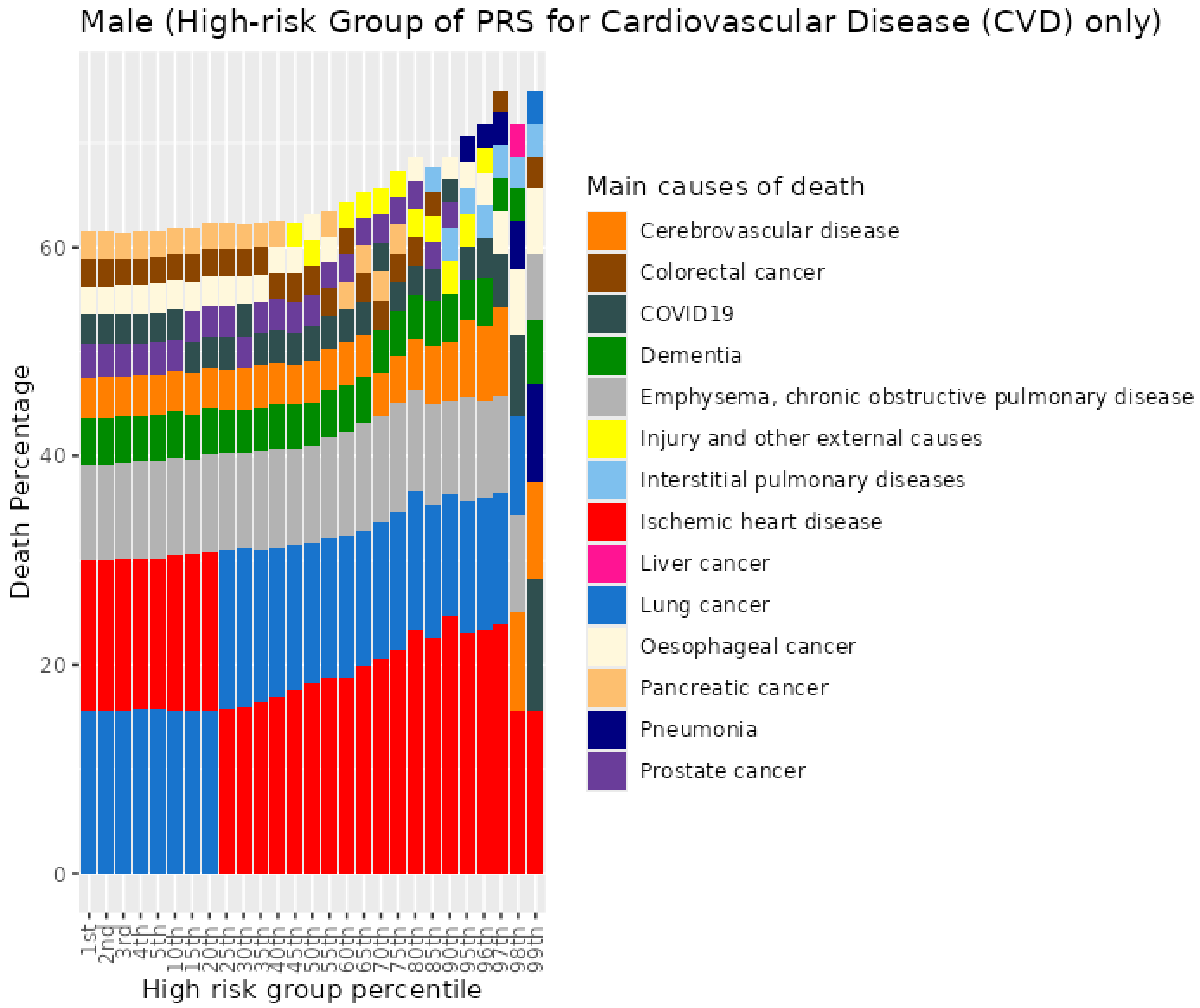


*Figure S90. Percentiles from 1 to 99 for cardiovascular disease PRS among male current smokers who died at age 70+*

### 11 Coronary artery disease (CAD) PRS for UKB males

##### 11.1 Male never smokers who died at age <=59

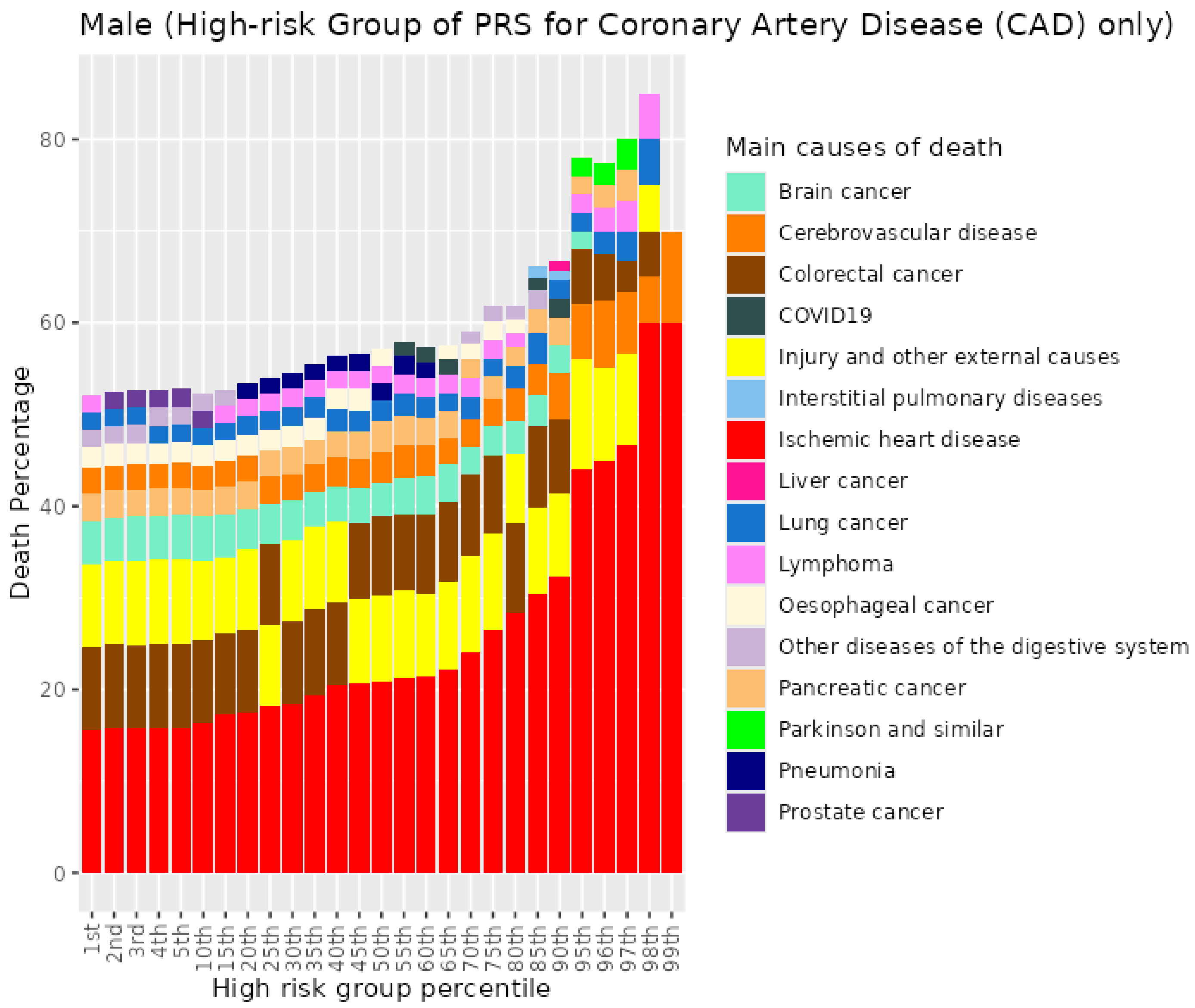


*Figure S91. Percentiles from 1 to 99 for coronary artery disease PRS among male never smokers who died at age <=59*

##### 11.2 Male never smokers who died at age 60-69

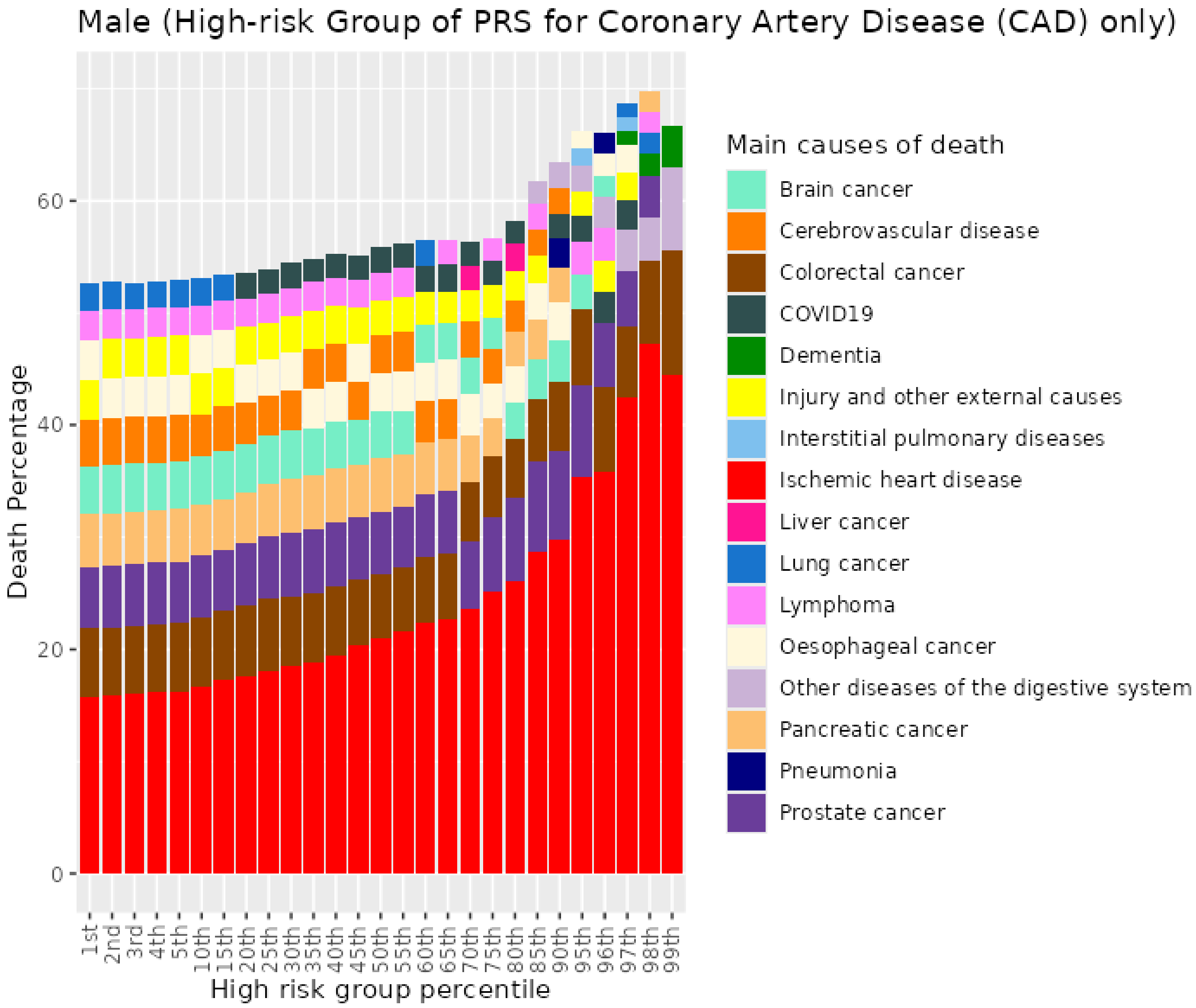


*Figure S92. Percentiles from 1 to 99 for coronary artery disease among male never smokers who died at age 60-69*

##### 11.3 Male never smokers who died at age 70+

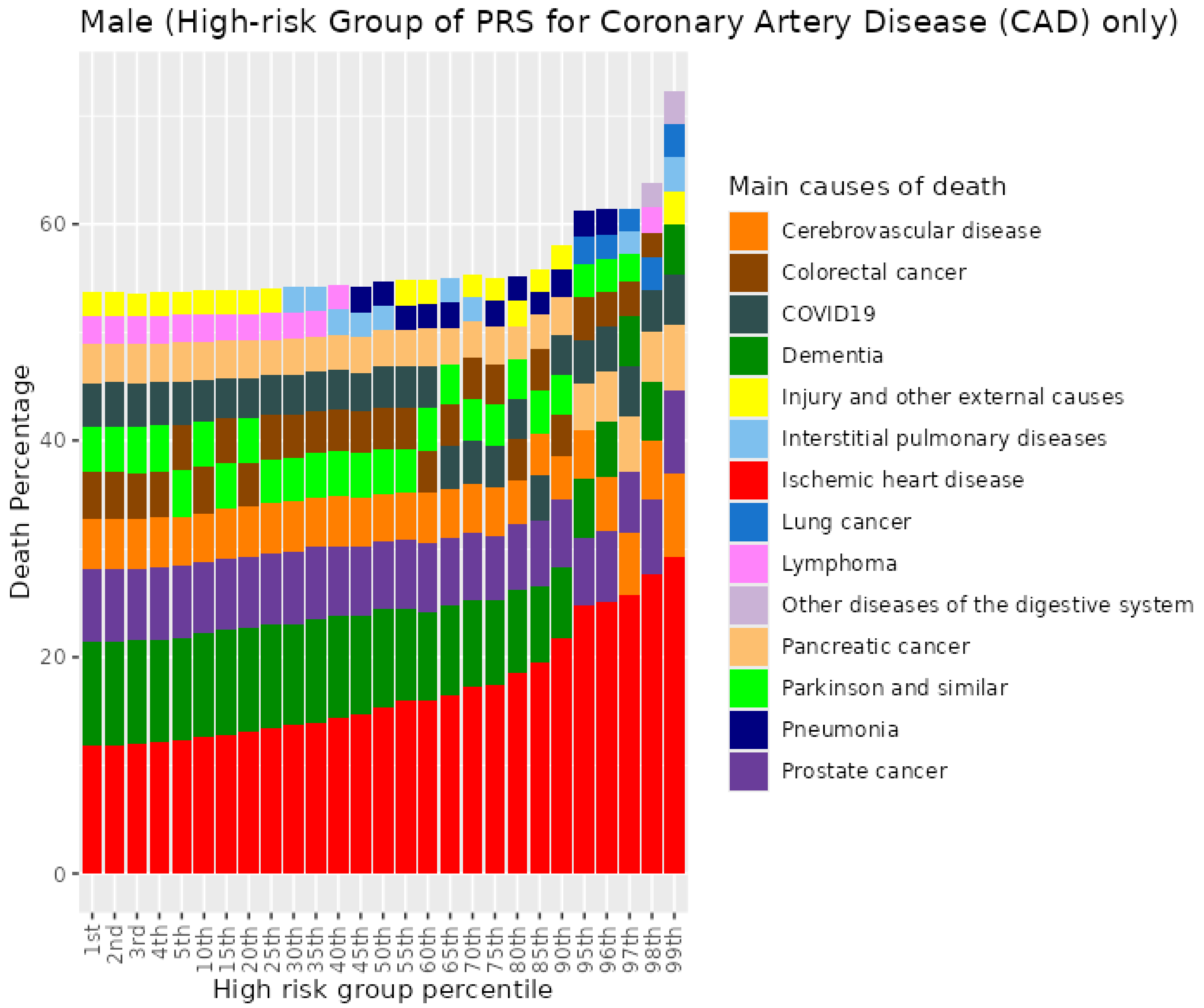


*Figure S93. Percentiles from 1 to 99 for coronary artery disease PRS among male never smokers who died at age 70+*

##### 11.4 Male previous smokers who died at age <=59

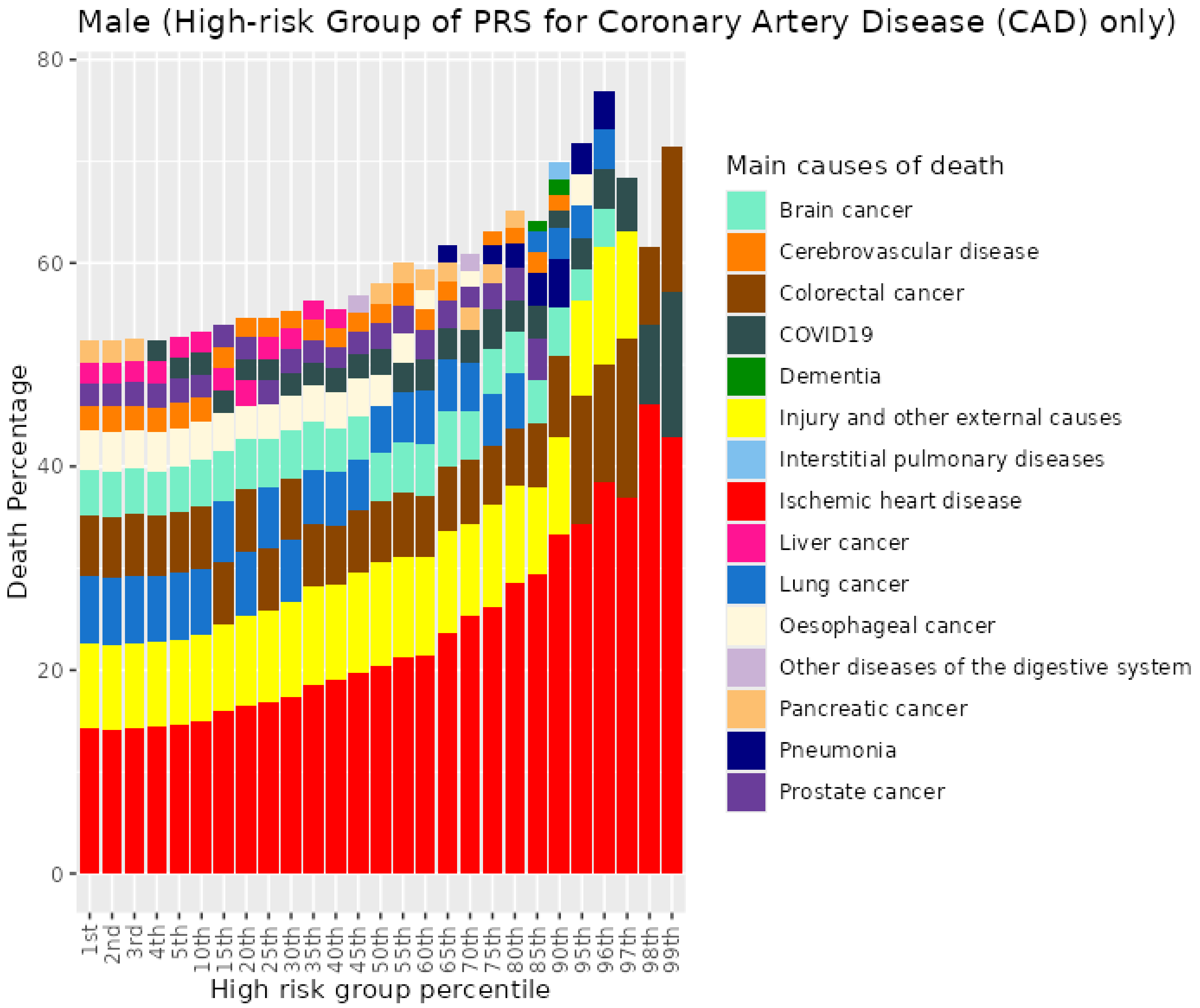


*Figure S94. Percentiles from 1 to 99 for coronary artery disease PRS among male previous smokers who died at age <=59*

##### 11.5 Male previous smokers who died at age 60-69

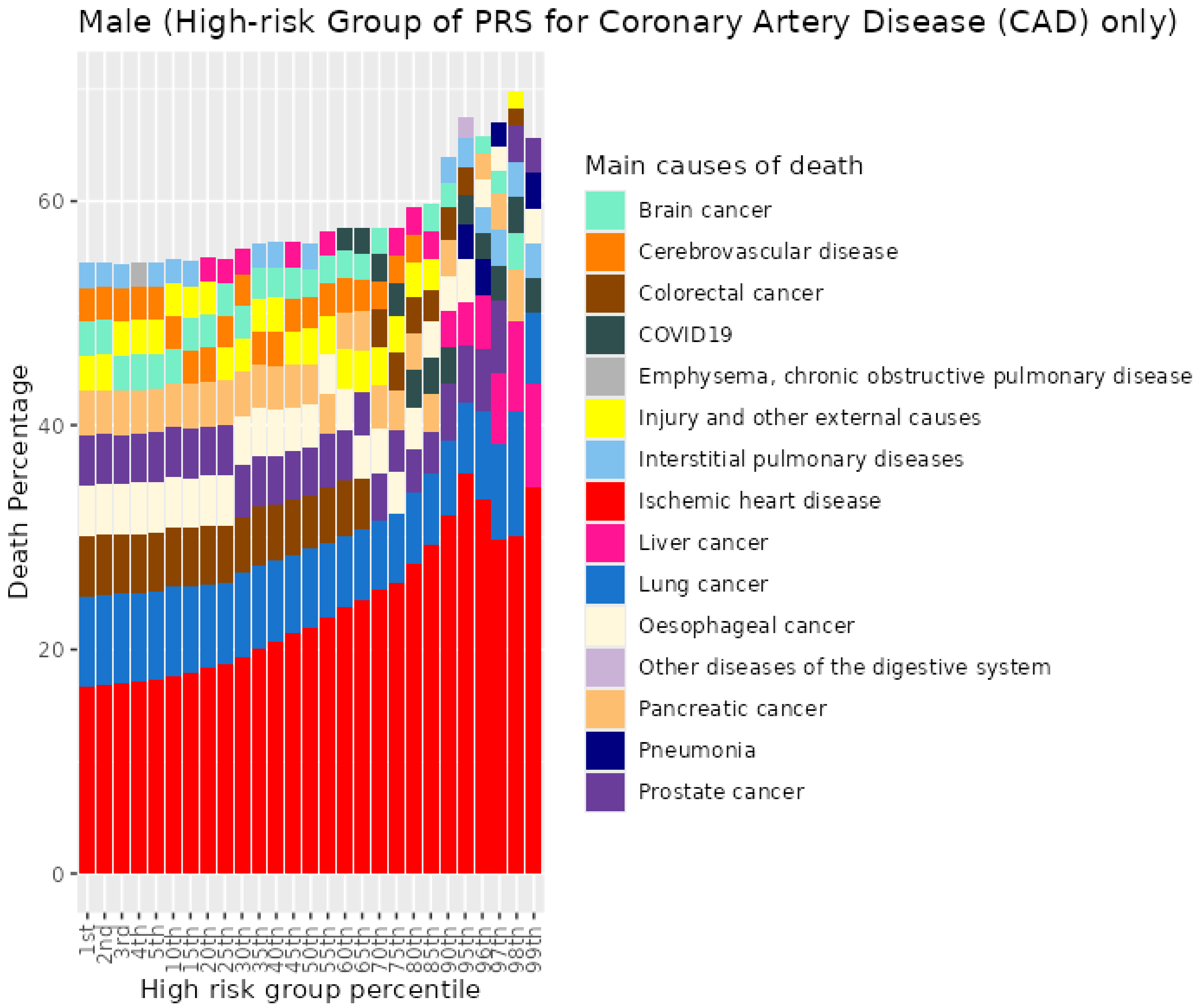


*Figure S95. Percentiles from 1 to 99 for coronary artery disease PRS among male previous smokers who died at age 60-69*

##### 11.6 Male previous smokers who died at age 70+

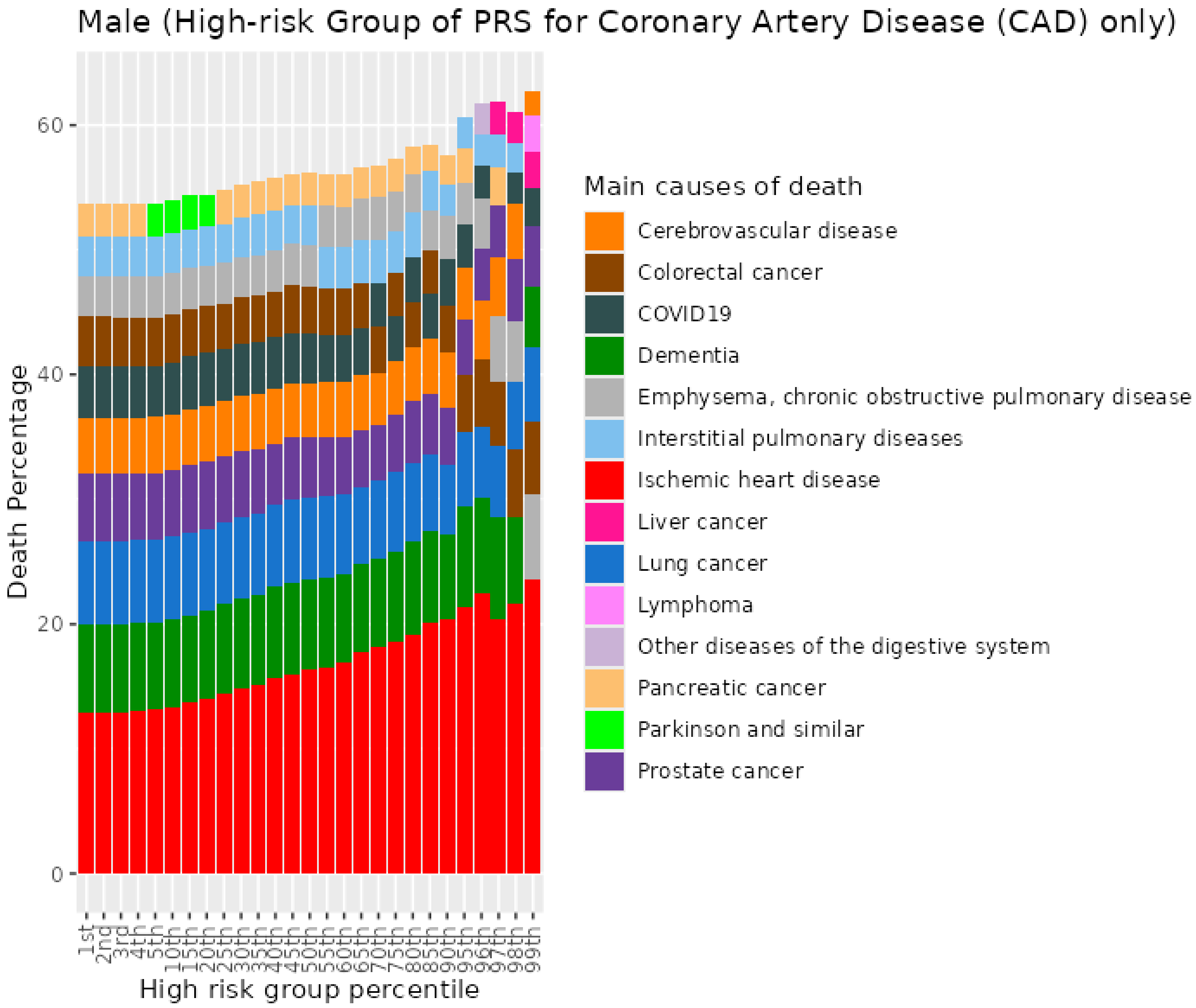


*Figure S96. Percentiles from 1 to 99 for coronary artery disease PRS among male previous smokers who died at age 70+*

#### 11.7 Male current smokers who died at age <=59

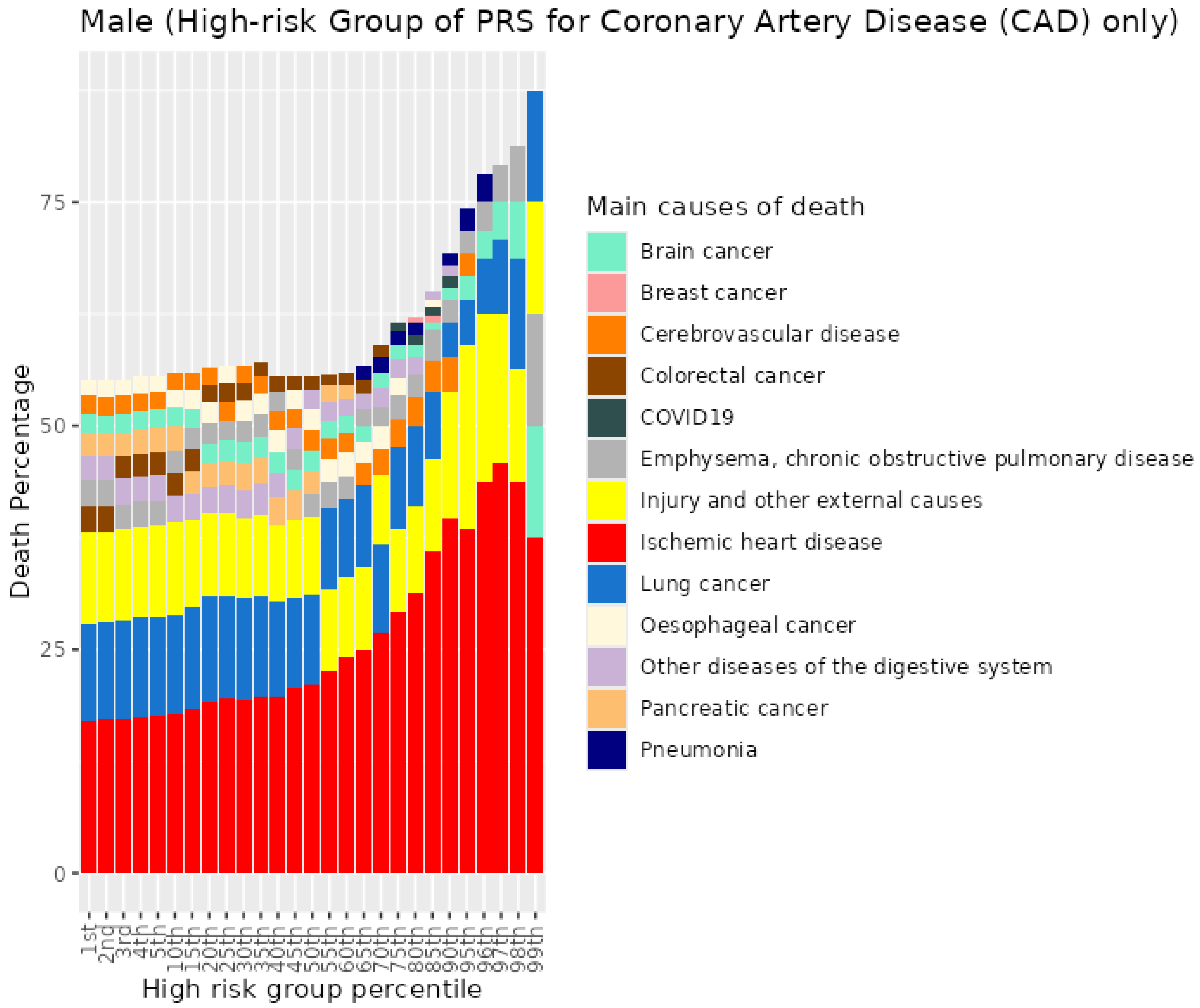


*Figure S97. Percentiles from 1 to 99 for coronary artery disease PRS among male current smokers who died at age <=59*

##### 11.8 Male current smokers who died at age 60-69

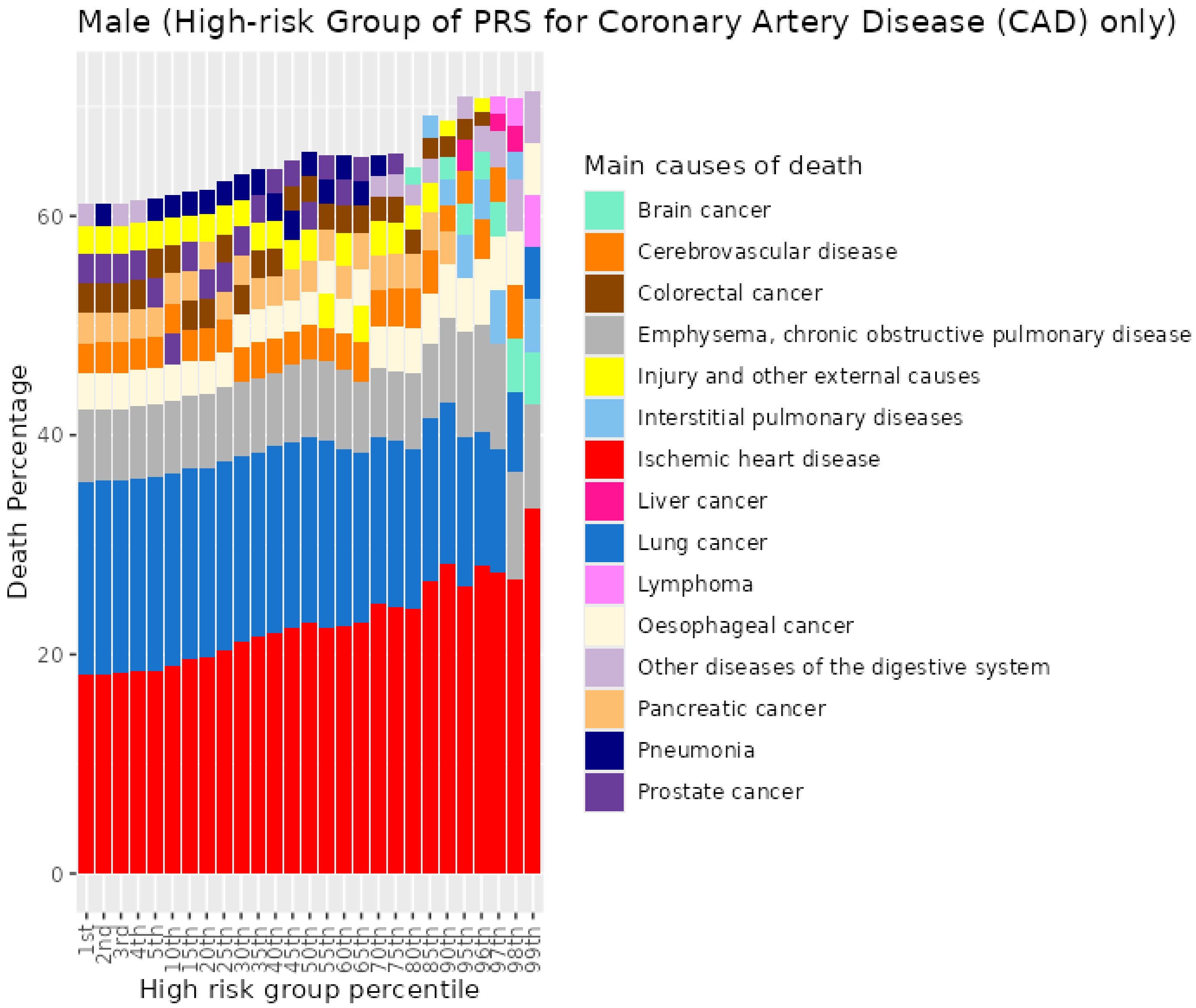


*Figure S98. Percentiles from 1 to 99 for coronary artery disease PRS among male current smokers who died at age 60-69*

##### 11.9 Male current smokers who died at age 70+

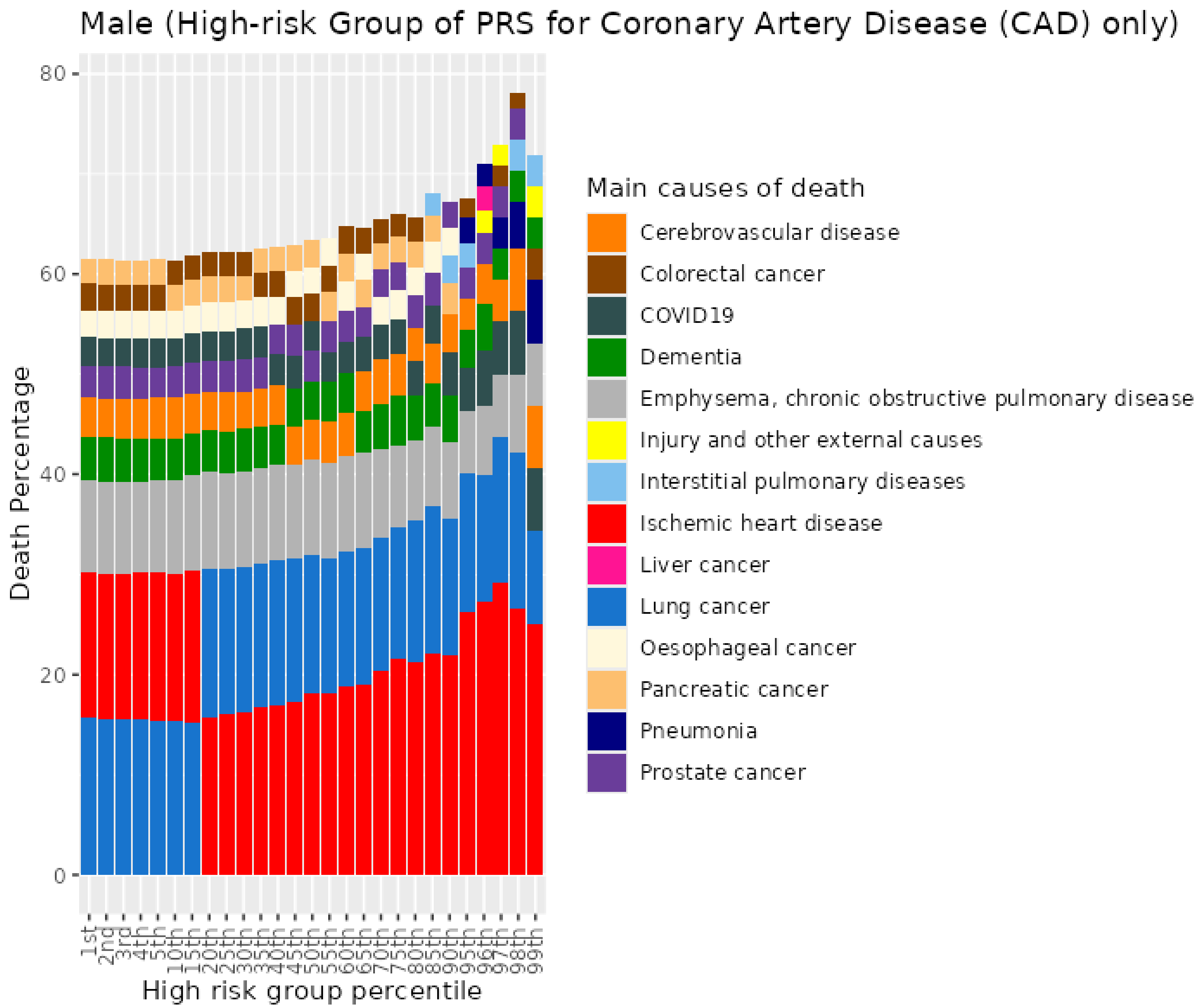


*Figure S99. Percentiles from 1 to 99 for coronary artery disease PRS among male current smokers who died at age 70+*

### 12 Ischaemic stroke (ISS) PRS for UKB males

#### 12.1 Male never smokers who died at age <=59

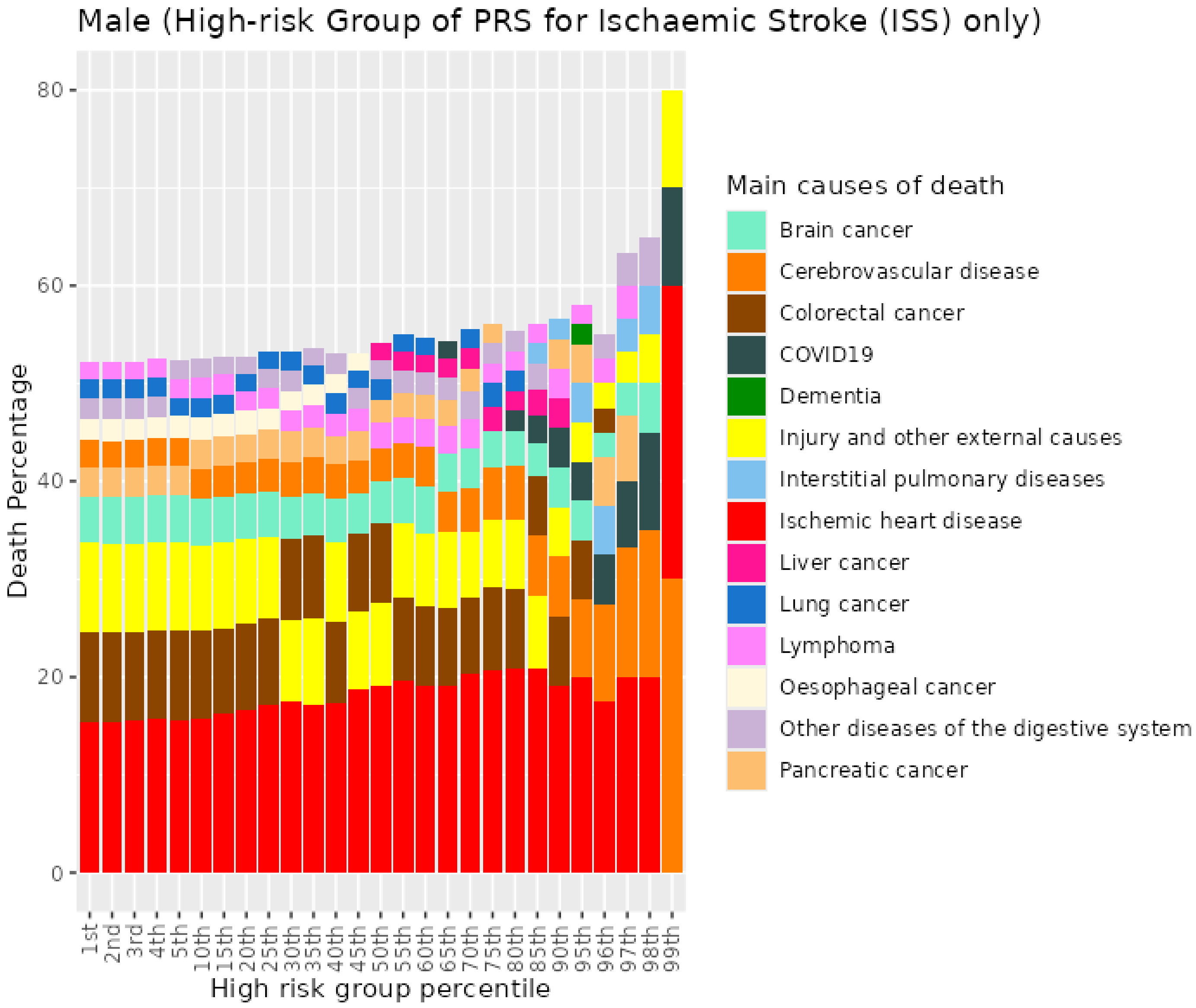


*Figure S100. Percentiles from 1 to 99 for ischaemic stroke PRS among male never smokers who died at age <=59*

##### 12.2 Male never smokers who died at age 60-69

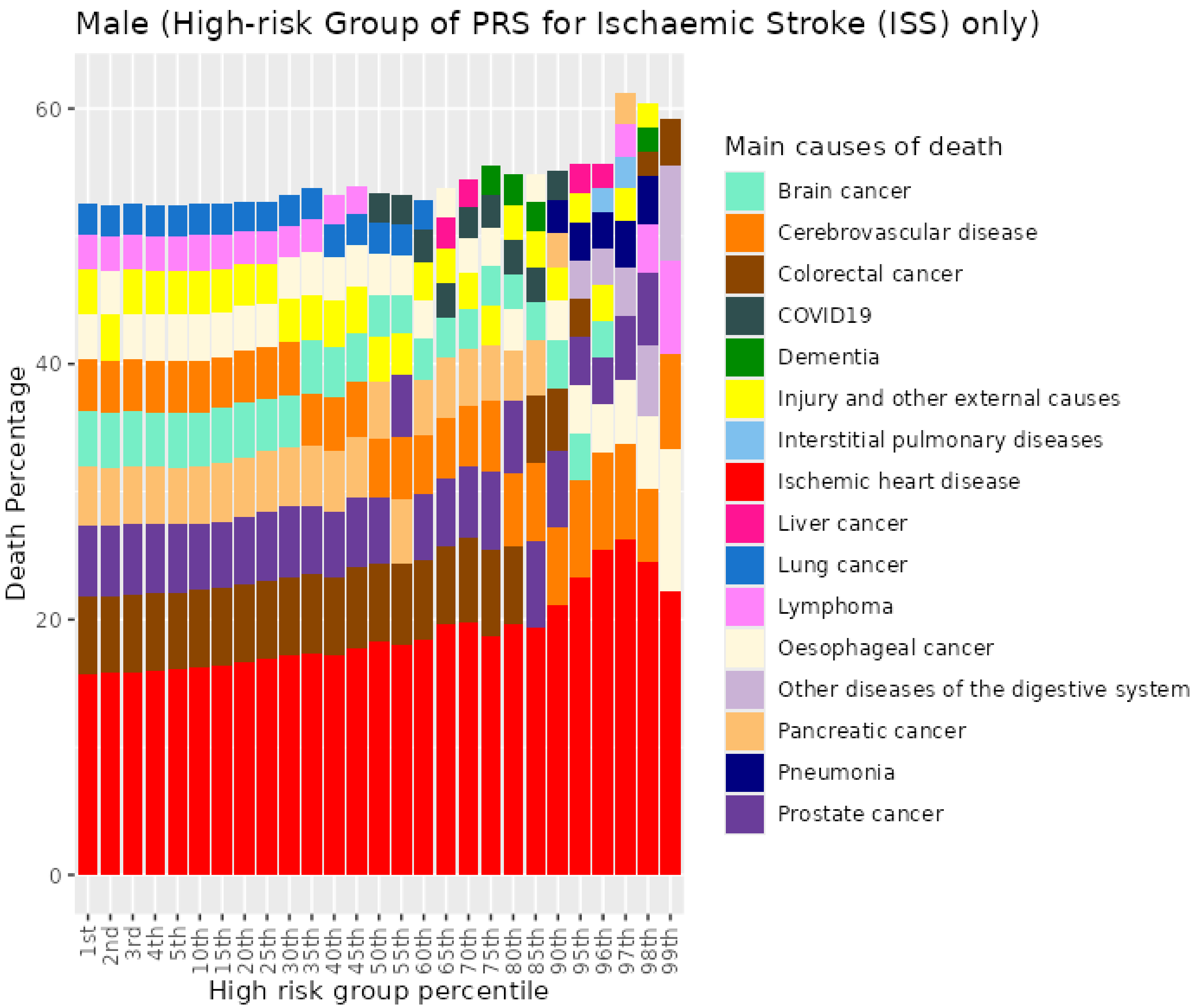


*Figure S101. Percentiles from 1 to 99 for ischaemic stroke PRS among male never smokers who died at age 60-69*

##### 12.3 Male never smokers who died at age 70+

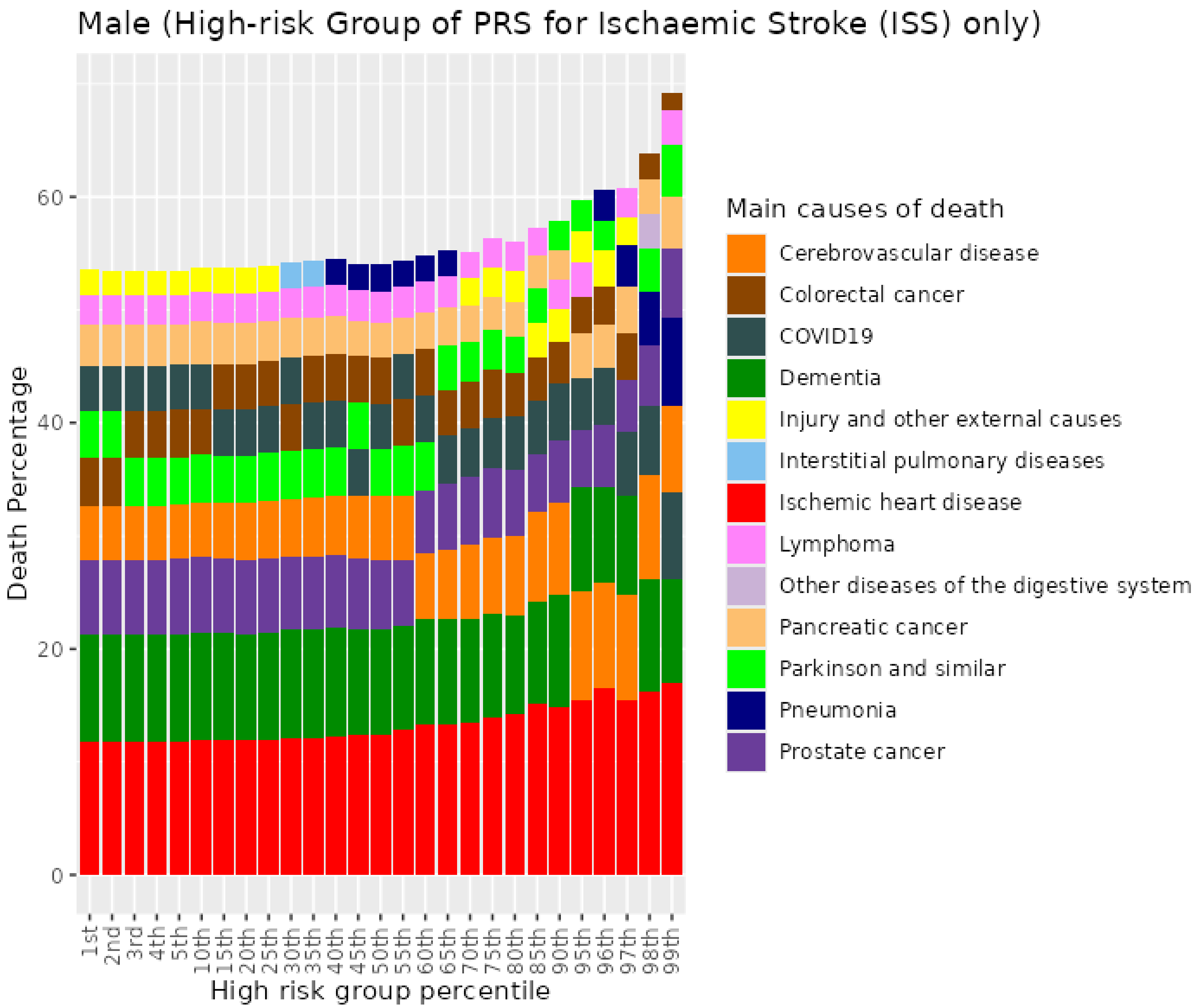


Figure S102. Percentiles from 1 to 99 for ischaemic stroke PRS among male never smokers who died at age 70+

##### 12.4 Male previous smokers who died at age <=59

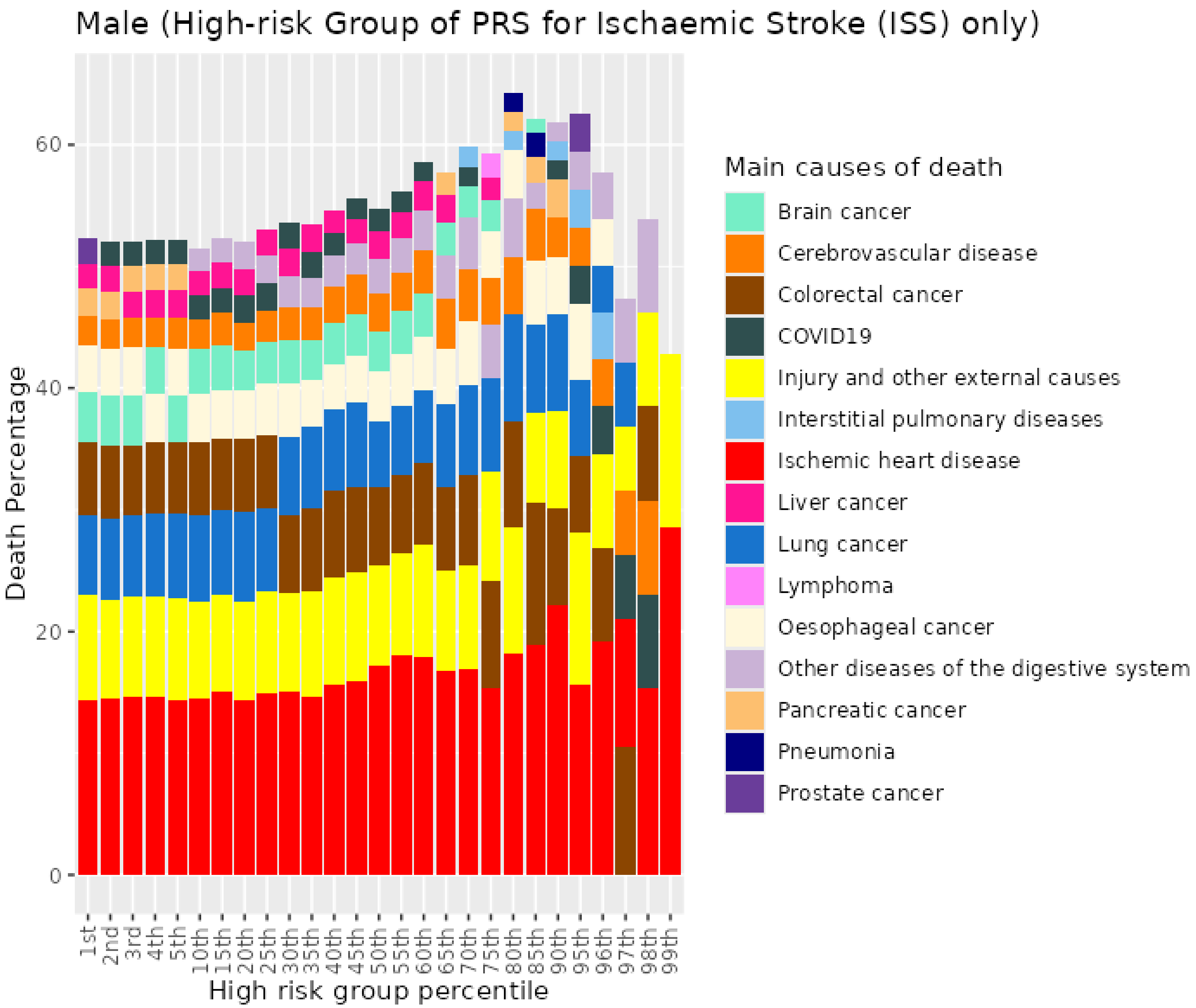


*Figure S103. Percentiles from 1 to 99 for ischaemic stroke PRS among male previous smokers who died at age <=59*

##### 12.5 Male previous smokers who died at age 60-69

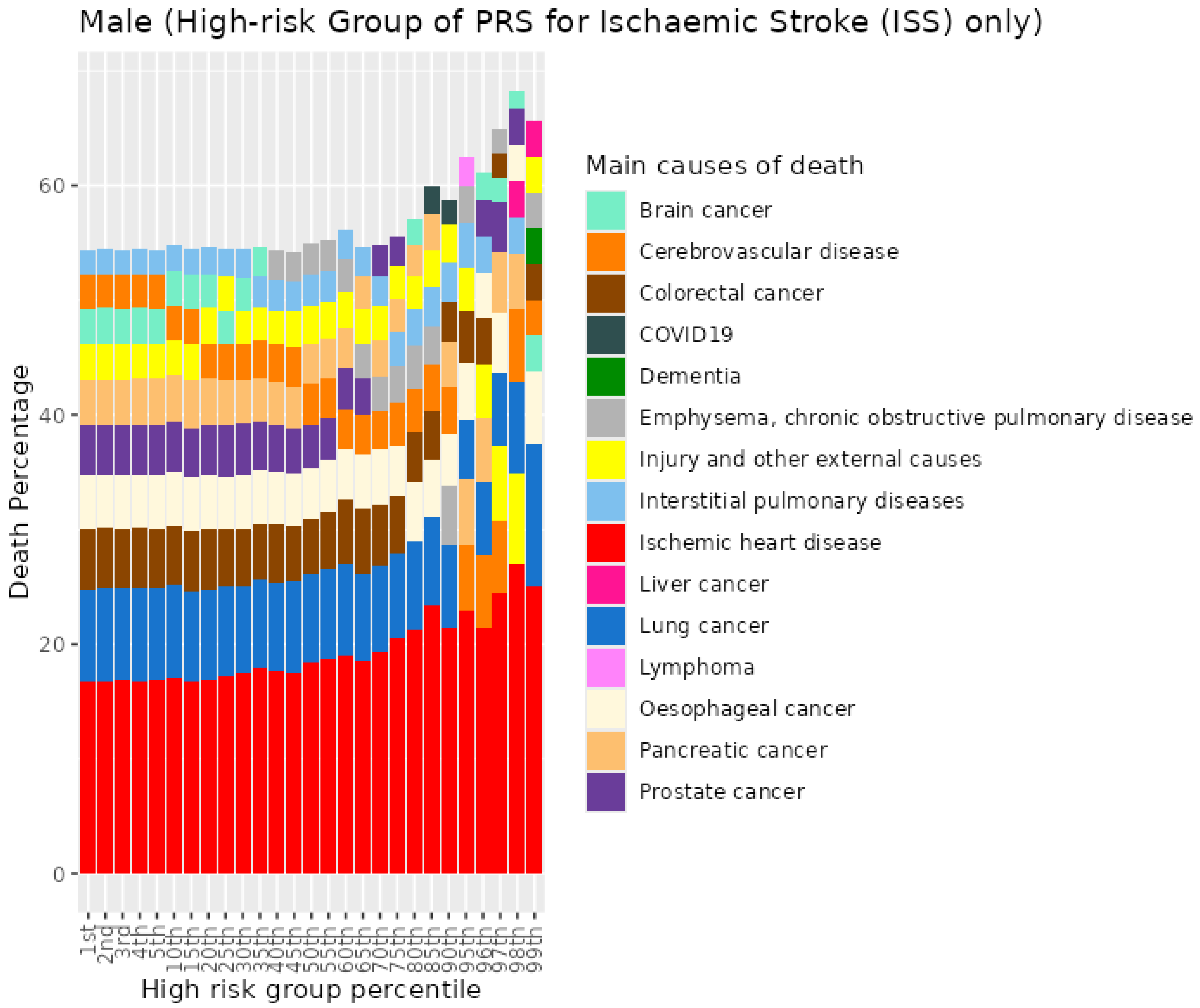


*Figure S104. Percentiles from 1 to 99 for ischaemic stroke PRS among male previous smokers who died at age 60-69*

##### 12.6 Male previous smokers who died at age 70+

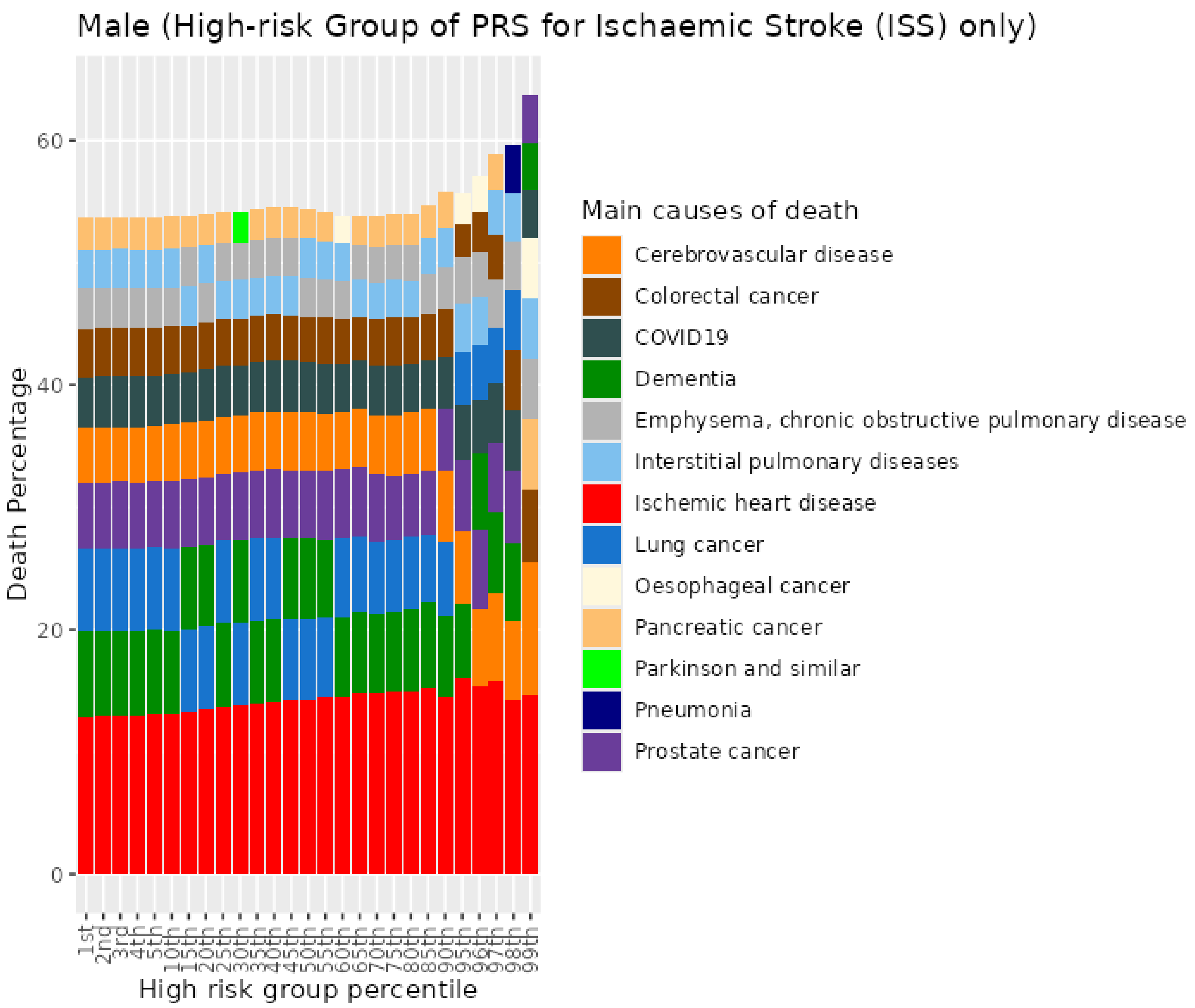


Figure S105. Percentiles from 1 to 99 for ischaemic stroke PRS among male previous smokers who died at age 70+

#### 12.7 Male current smokers who died at age <=59

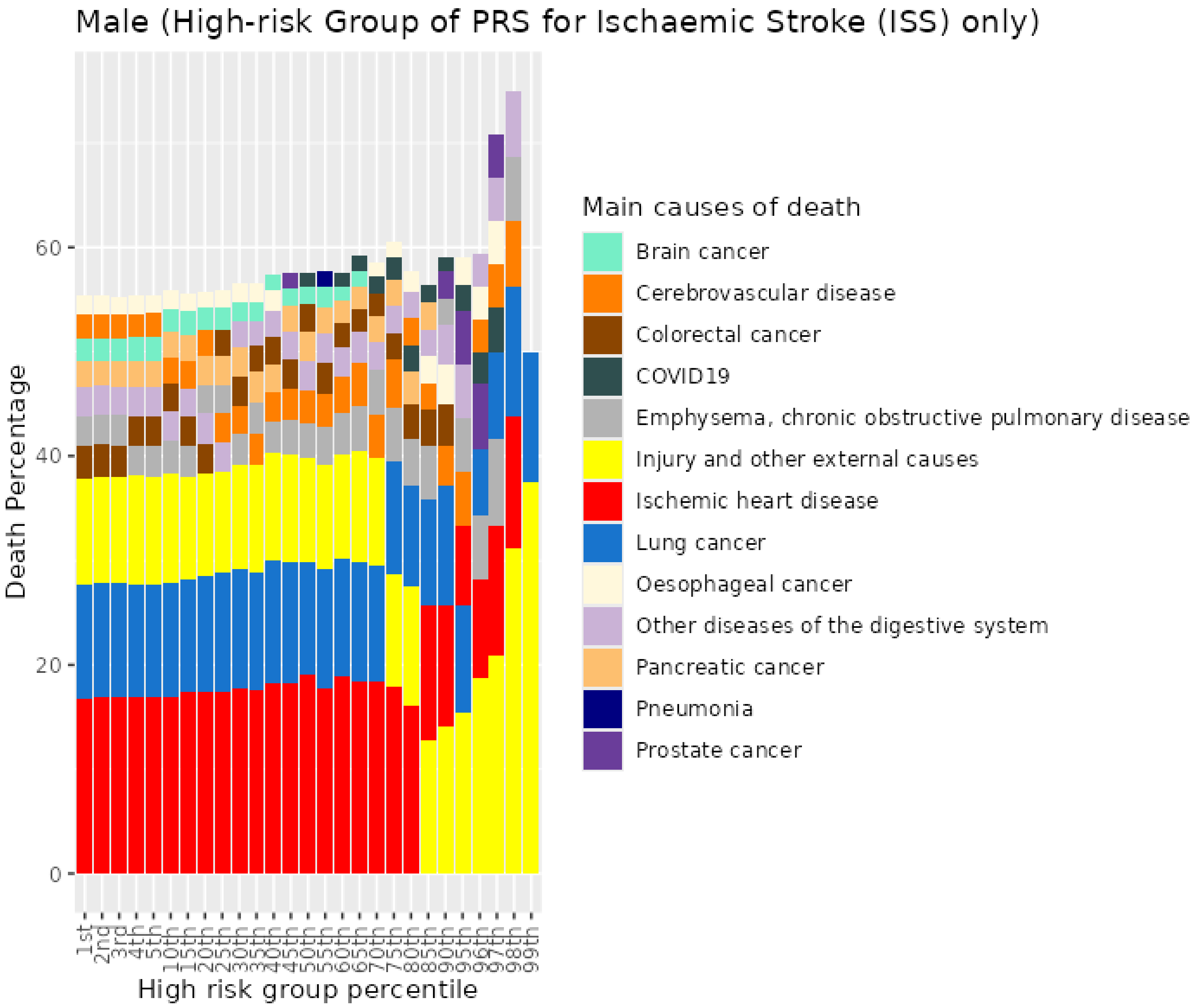


*Figure S106. Percentiles from 1 to 99 for ischaemic stroke PRS among male current smokers who died at age <=59*

#### 12.8 Male current smokers who died at age 60-69

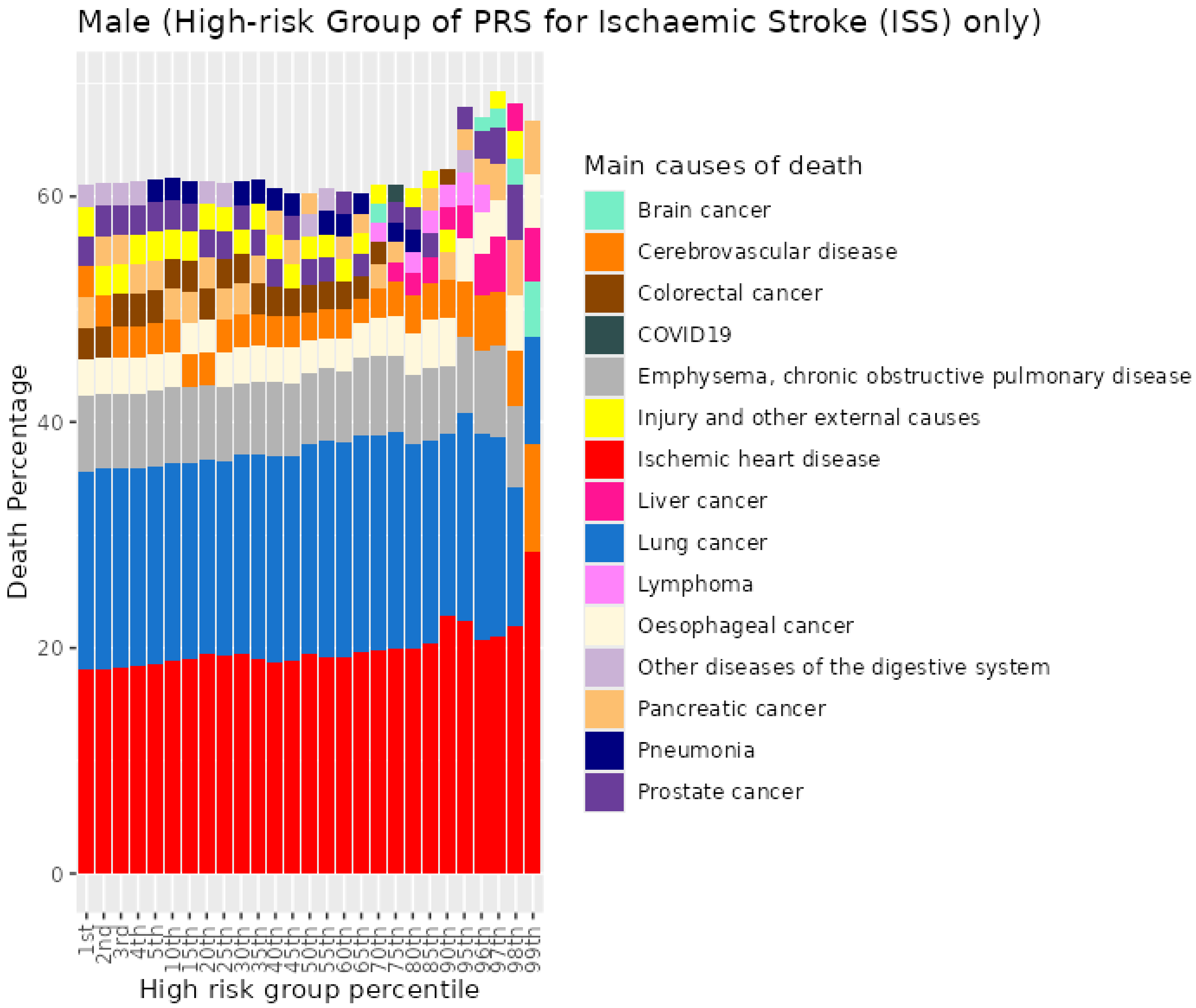


*Figure S107. Percentiles from 1 to 99 for ischaemic stroke PRS among male current smokers who died at age 60-69*

#### 12.9 Male current smokers who died at age 70+

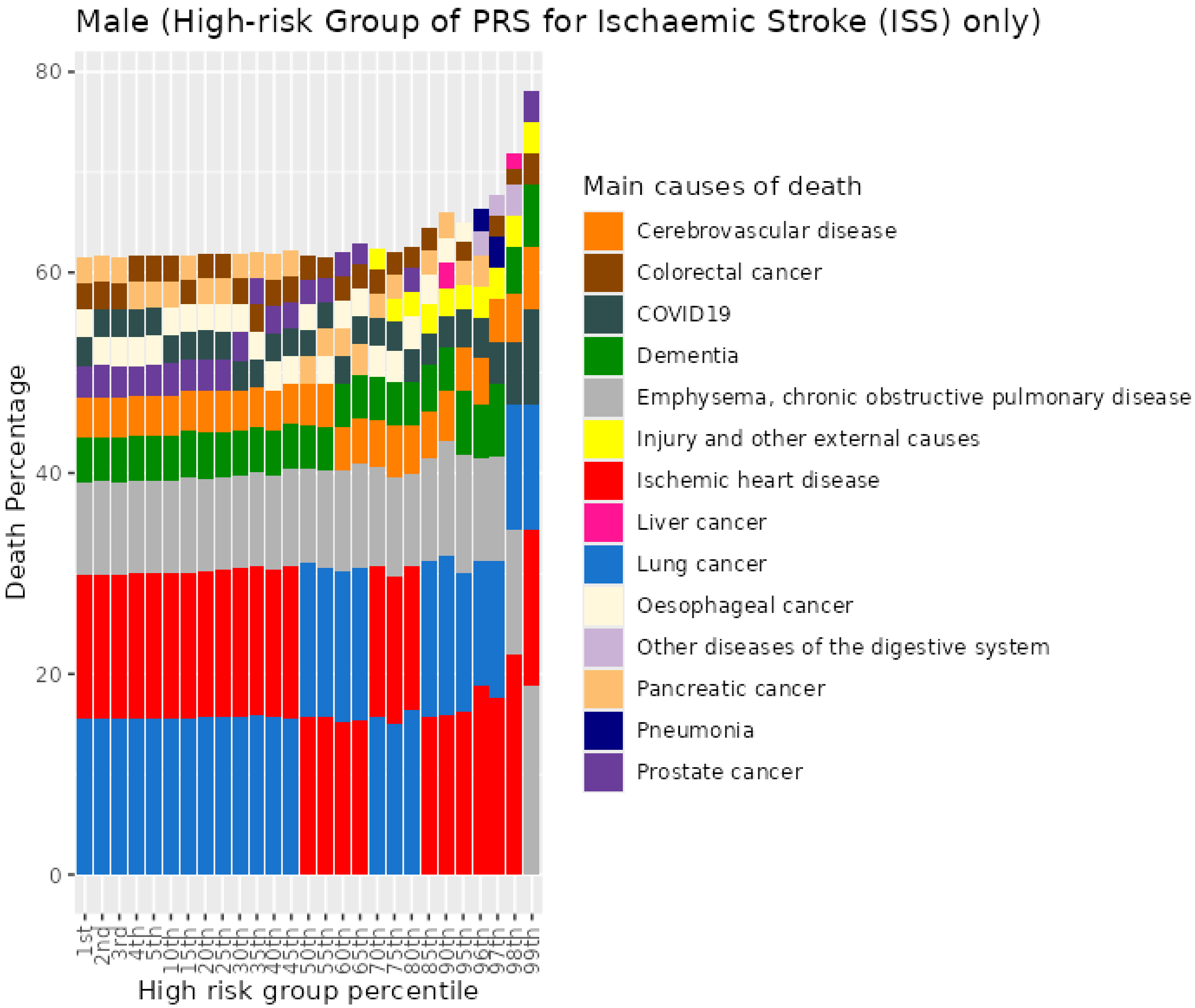


Figure S108. Percentiles from 1 to 99 for ischaemic stroke PRS among male current smokers who died at age 70+

#### 13 Peostate cancer (PC) PRS for UKB males

##### 13.1 Male never smokers who died at age <=59

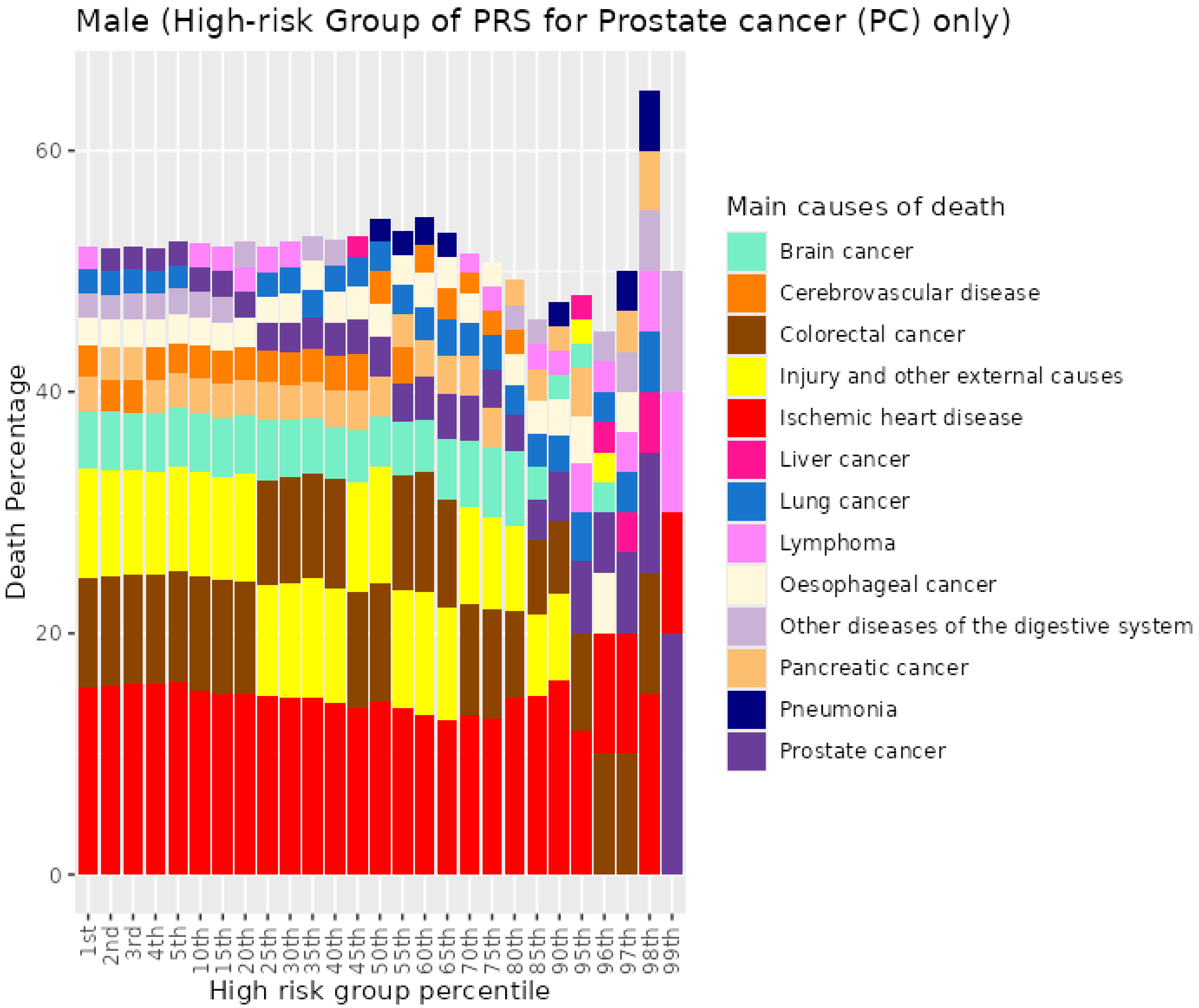


Figure S109. Percentiles from 1 to 99 for prostate cancer PRS among male never smokers who died at age <=59

##### 13.2 Male never smokers who died at age 60-69

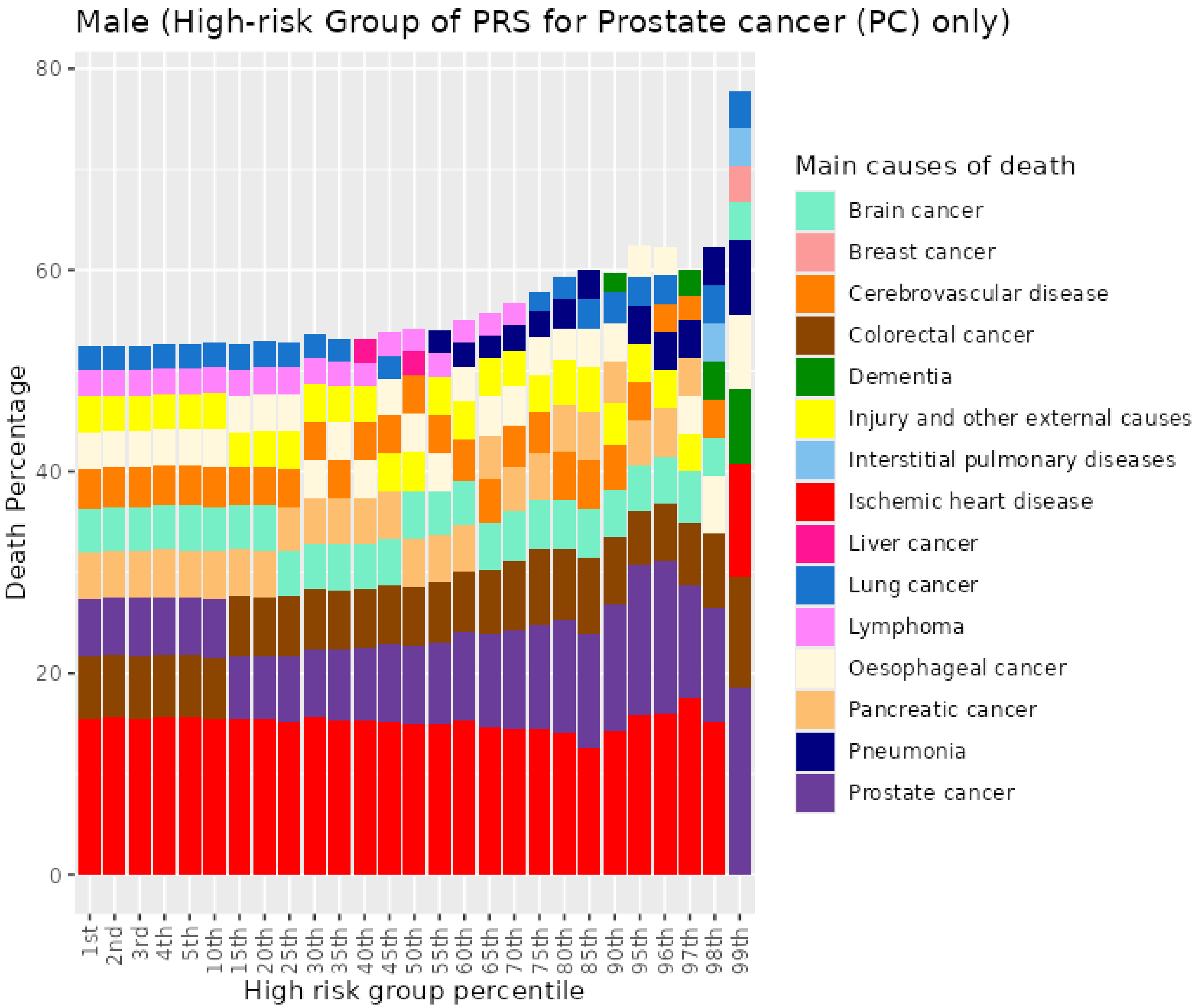


Figure S110. Percentiles from 1 to 99 for prostate cancer PRS among male never smokers who died at age 60-69

##### 13.3 Male never smokers who died at age 70+

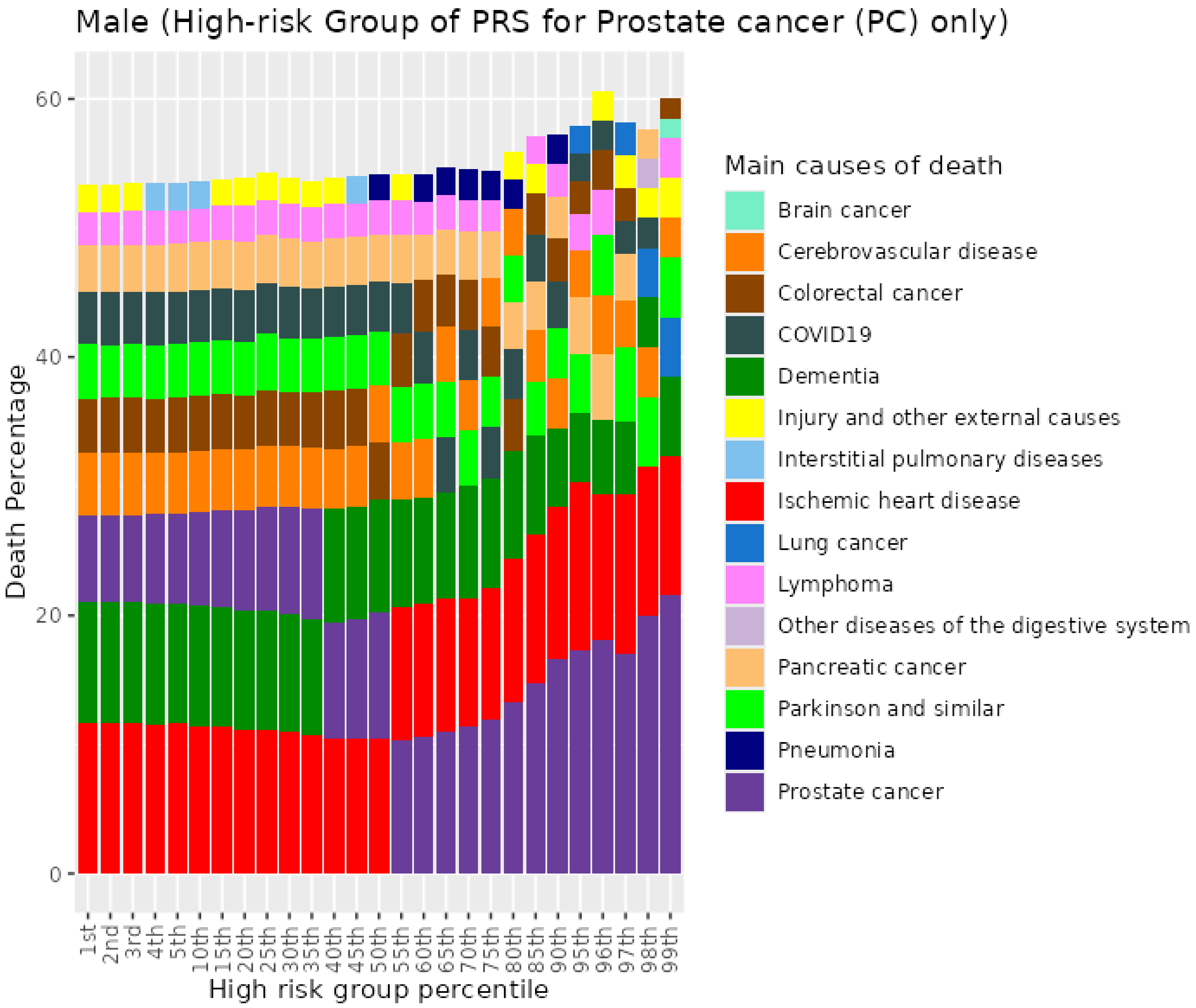


Figure S111. Percentiles from 1 to 99 for prostate cancer PRS among male never smokers who died at age 70+

#### 13.4 Male previous smokers who died at age <=59

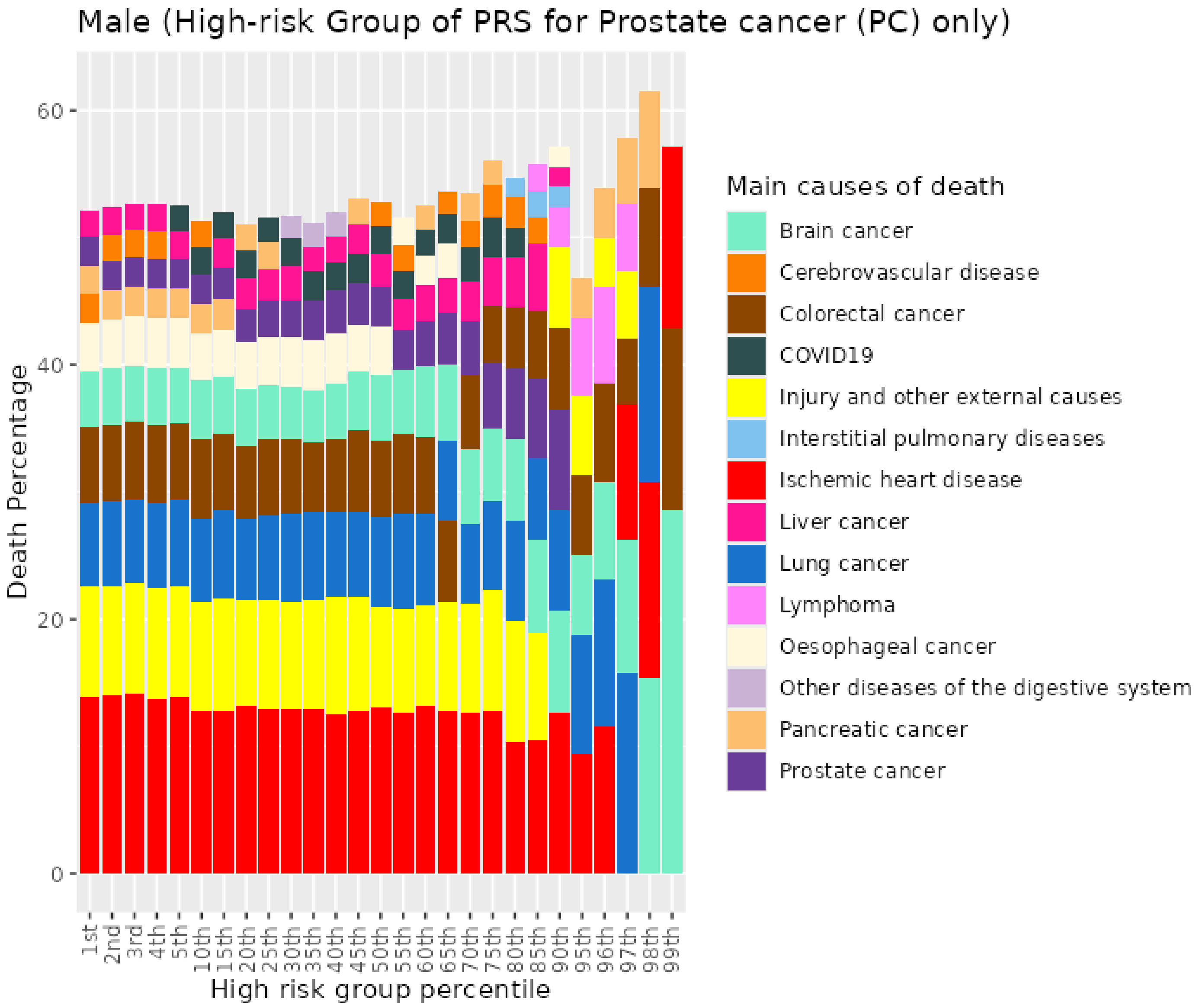


Figure S112. Percentiles from 1 to 99 for prostate cancer PRS among male previous smokers who died at age <=59

#### 13.5 Male previous smokers who died at age 60-69

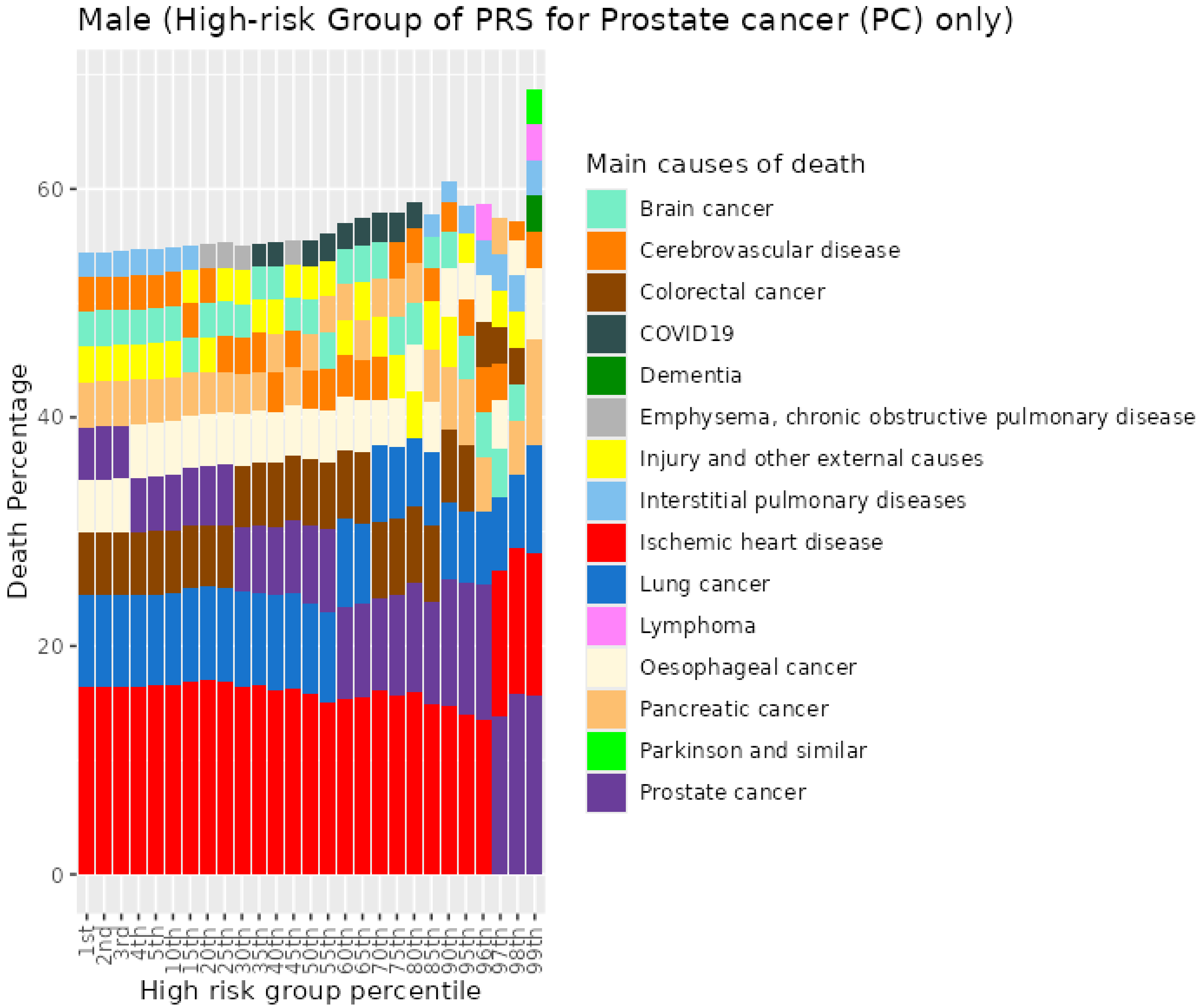


Figure S113. Percentiles from 1 to 99 for prostate cancer PRS among male previous smokers who died at age 60-69

#### 13.6 Male previous smokers who died at age 70+

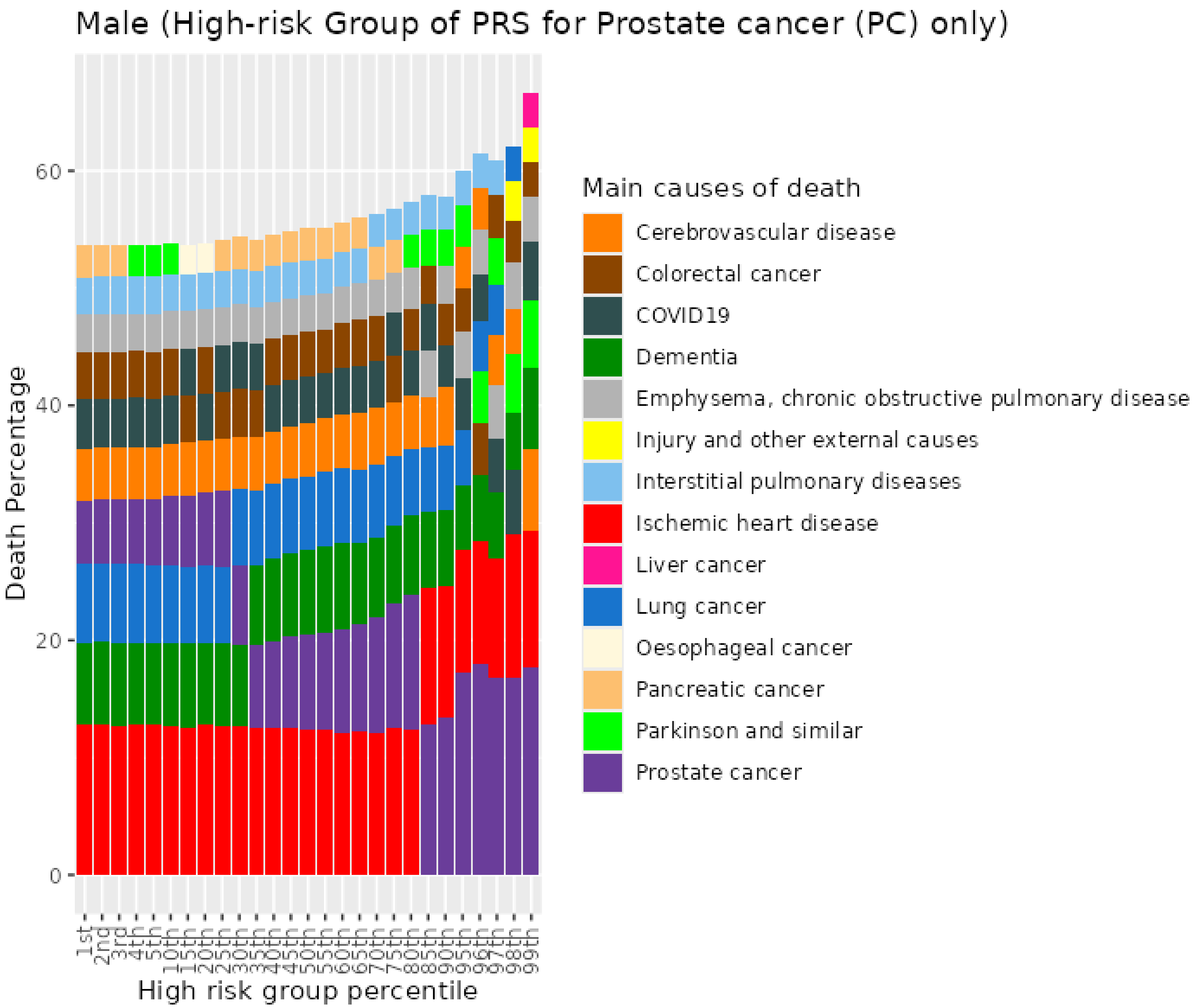


Figure S114. Percentiles from 1 to 99 for prostate cancer PRS among male previous smokers who died at age 70+

##### 13.7 Male current smokers who died at age <=59

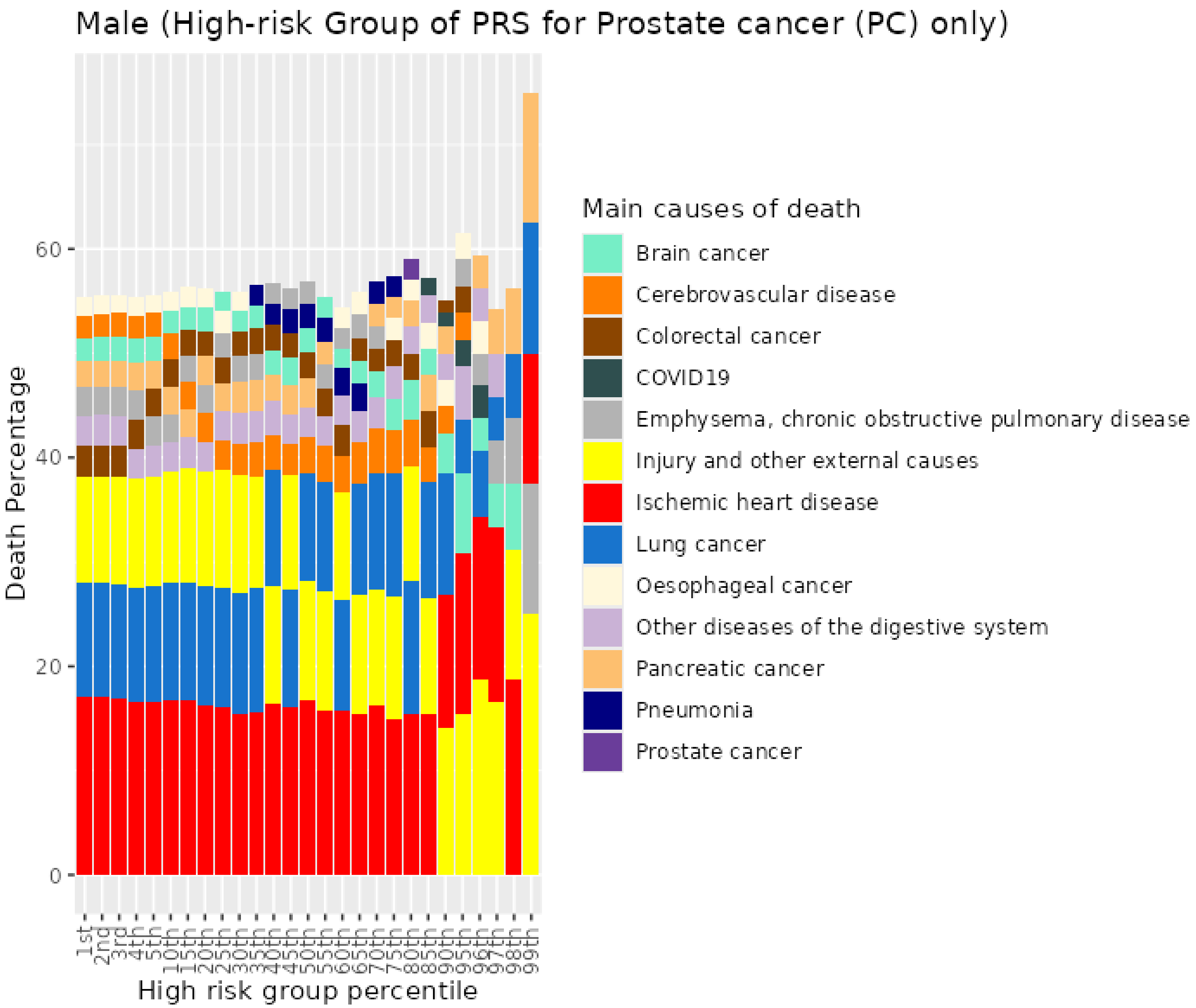


Figure S115. Percentiles from 1 to 99 for prostate cancer PRS among male current smokers who died at age <=59

##### 13.8 Male current smokers who died at age 60-69

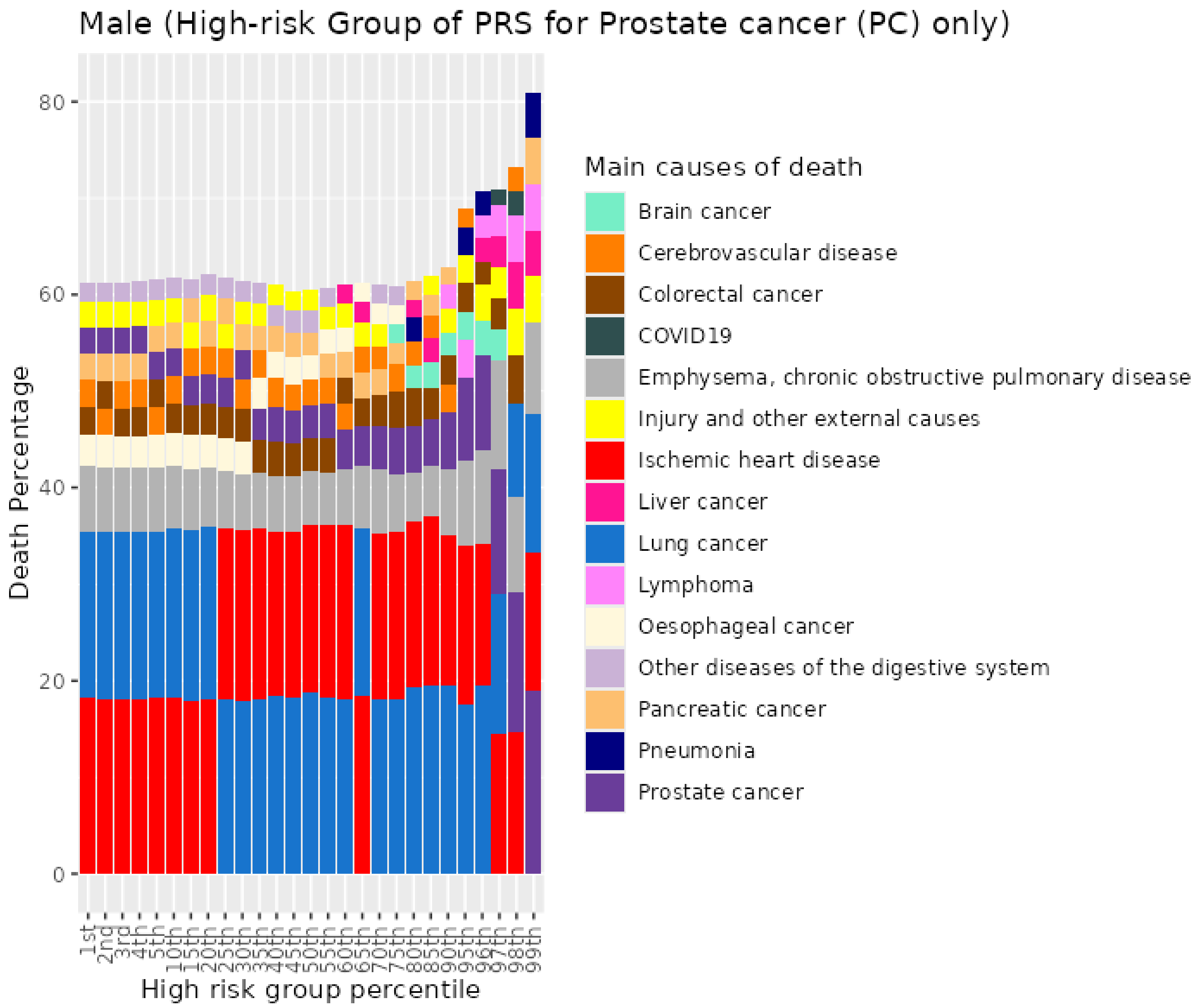


Figure S116. Percentiles from 1 to 99 for prostate cancer PRS among male current smokers who died at age 60-69

#### 13.9 Male current smokers who died at age 70+

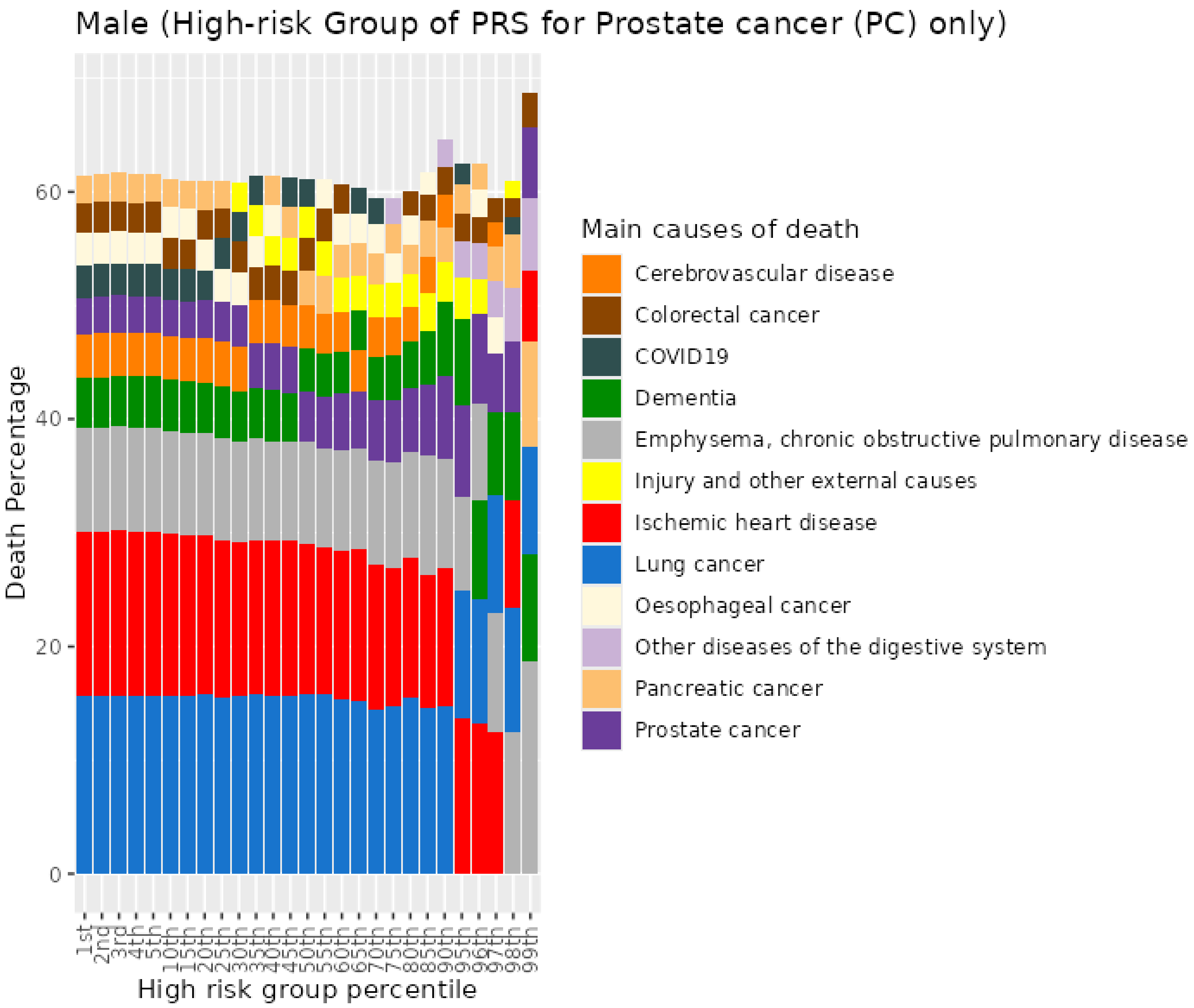


Figure S117. Percentiles from 1 to 99 for prostate cancer PRS among male current smokers who died at age 70+
